# Early Life Experiences and Adult Community Participation in Secular and Religious Contexts in 22 Countries

**DOI:** 10.1101/2025.03.14.25323977

**Authors:** Ying Chen, Brendan W. Case, Katelyn N. G. Long, Robert Woodberry, Eric S. Kim, R. Noah Padgett, Byron R. Johnson, Tyler J. VanderWeele

## Abstract

Participation in community groups (including both secular and religious groups) is associated with improved health, well-being, and societal cohesion. However, less is known about conditions that increase community participation across the life-course, especially childhood factors. Using data from 202,898 adults across 22 countries, this study evaluated childhood candidate antecedents of community participation in both secular and religious contexts during adulthood. The associations were examined in each country separately, and also cross-nationally by meta-analytically pooling country-specific estimates. The results suggest that the childhood experiences of living comfortably financially, child abuse, feeling like an outsider in the family, having excellent self-rated health, frequent religious service attendance, a later birth year, and being male were each associated with a greater likelihood of weekly^+^ secular community participation in adulthood. In comparison, the childhood experiences of having good relationship with father, having married parents, frequent religious service attendance, and an earlier year of birth were each associated with a higher likelihood of weekly^+^ religious service attendance in adulthood. The direction and strength of these associations differed by country, indicating diverse societal influences. The findings provide valuable insights into early-life experiences that may shape different forms of community participation later in life across societies.

## INTRODUCTION

As local communities weaken across much of the world, participation in community groups remains an essential means of social connection^1^. Community participation empowers people to engage meaningfully with society, which may lead to individual and communal flourishing^2–5^. Community participation also provides a meaningful avenue for enhancing social cohesion, and addressing significant societal challenges such as the burgeoning pandemic of loneliness^4,6^. Certain forms of community participation, such as civic engagement, are even regarded as the bedrock of democratic societies^7,8^. Nevertheless, some traditional forms of community participation have been declining in many countries^9,10^. It is, therefore, important to understand effective approaches to enhancing community participation across diverse populations.

Community participation (i.e., formal social participation) is often defined as “active interactions with established organizations such as volunteer, cultural, sports, leisure, and religious communities”^11,12^. Community participation is associated with improved health and well-being outcomes including lower risks of mortality^13,14^ and mental illness^15,16^, as well as better physical functioning^17^ and psychosocial well-being^14,16,18^. There is also evidence that participation in faith-based communities (e.g., religious service attendance) may have stronger connections with favorable health outcomes than participation in secular groups^13,19^, but such dynamics may vary by national contexts (e.g., country-level religious regulation)^20^. In addition, community participation may enhance a range of significant societal and economic outcomes, such as building social trust^21,22^, fostering economic mobility^23^, and strengthening democratic institutions^24^.

Early life is likely a sensitive period for fostering a propensity for social engagement^25,26^. While empirical evidence on childhood antecedents of adult community participation is limited, the studies that do exist suggest a complex interplay between individual, familial, and societal experiences in childhood that shapes one’s community participation later in life. First, childhood familial environment ranging from positive parent-child relationships, stressful events (e.g., parental divorce), to adverse experiences (e.g., abuse and neglect) may all affect the child’s development of social skills, trust, and empathy, which shape adult community participation^27,28^. For instance, prior studies found that positive parent-child relationship in adolescence is associated with greater civic engagement, political participation, volunteering, and religious service attendance during young adulthood^28–30^, whereas child maltreatment may undermine adult social functioning and increase the risk of social isolation^31,32^. Second, childhood socioeconomic status (SES) may influence the opportunities and resources available for community participation. Prior studies found that people who grow up in families with lower versus (vs.) higher SES often reported a lower rate of adult participation in secular communities (e.g., civic, political, voluntary groups)^27,33,34^. This may be partly because families with higher SES are more likely to possess resources, have broad social networks, and stay updated of current affairs, creating a cycle of opportunities, resources, and values that altogether shape one’s community engagement later in life^9,35^. In comparison, how childhood SES shapes adult religious community participation is less clear. Existing evidence generally suggests that the dynamics between SES and religious service attendance may vary depending on the indicator of SES, religious denomination, and different national context^36,37^. Third, other social identity factors such as immigration status may also influence access to community participation. For instance, prior evidence suggests that immigrant youth may encounter more challenges in integrating into the civic life of the host society than the native-born, but the patterns may vary depending on the host county’s integration policies^38,39^. Fourth, childhood health status may also influence one’s capacity to participate in community groups. For instance, there is evidence suggesting that adolescents with mental distress and functional limitations are less likely to engage in civic, political, and volunteering activities when they grow up into adults, compared to healthy adolescents^40,41^. Fifth, a religious upbringing may help young people develop broader social networks, foster stronger civic character, and nurture social trust and shared responsibility^22^. For instance, prior evidence from the United States suggests that adolescents with a religious upbringing on average reported more frequent participation in volunteering, civic, and religious communities as adults^37^, although it is plausible that this pattern may differ by contextual factors (e.g., country-level religious composition and religious regulation)^20^.

Prior studies have advanced our understanding of childhood antecedents of community participation, but some knowledge gaps remain. First, the rising rates of secularization in many societies may affect early life experiences of participation in faith-based communities for those who hold religious beliefs, especially among more recent cohorts. This may have significant implications for intervention programs that aim to enhance community participation across diverse populations. However, prior studies have seldom directly compared childhood antecedents of secular and religious community participation. Such knowledge may be essential for developing targeted, culturally relevant, and socially inclusive programs for enhancing people’s participation in different forms of communities starting from early life. Second, most prior studies are based on data from a single country, which limits a comprehensive understanding of how childhood experiences may affect individuals’ engagement in different forms of community groups under different cultural and societal contexts. Third, prior studies on childhood antecedents of social participation are mostly based on data from Western, Educated, Industrialized, Rich, and Democratic (WEIRD) countries and/or used non-representative samples from those countries^42,43^, which limits the generalizability of the study findings even within the countries where these studies were conducted.

To begin addressing these knowledge gaps, this study used data from a large sample of adults across 22 diverse countries, with the samples weighted to be nationally representative within each country, to investigate several childhood personal attributes, and familial or social circumstances as potential childhood antecedents of adult community participation, with participation in secular and religious communities examined separately. We hypothesized that 1) the childhood factors that we examined would have varied associations with adult participation in secular and religious communities, and 2) the direction and strength of these associations would differ by country, reflecting diverse societal influences.

## METHODS

This study used Wave 1 data from the Global Flourishing Study (GFS). The description of the methods below has been adapted from VanderWeele et al.^44^ (the paper by VanderWeele et al. is scheduled to be published before the present study, and the reference will be updated when it becomes available). Further methodological detail regarding the GFS is available elsewhere^45–50^. The present study is a secondary analysis of the existing GFS data.

### Study Population

The Global Flourishing Study is a longitudinal study that enrolled 202,898 adults (age range: 18 to 99 years) from 22 culturally and geographically diverse countries, with the samples weighted to be nationally representative within each country. Survey items queried multiple aspects of well-being including happiness, health status, sense of meaning and purpose, character, social relationships, and financial stability, along with a range of demographic, socioeconomic, political, religious, personality, childhood, community, and behavior factors. Data collection was carried out by Gallup via a combination of modes (e.g., in-person, phone, web) that varied across countries^50^. Data for Wave 1 were collected principally during 2023, while some countries began data collection in 2022^45^. The following countries were included in Wave 1 data collection: Argentina, Australia, Brazil, Egypt, Germany, Hong Kong (Special Administrative Region of China with mainland China included from 2024 onwards), India, Indonesia, Israel, Japan, Kenya, Mexico, Nigeria, the Philippines, Poland, South Africa, Spain, Sweden, Tanzania, Turkey, United Kingdom, and the United States. These countries were chosen to 1) maximize coverage of the world’s population, 2) ensure geographic, cultural, and religious diversity, and 3) prioritize feasibility and existing data collection infrastructure. The precise sampling design to ensure nationally representative samples varied by country^45^. During the survey translation process, Gallup adhered to TRAPD model (translation, review, adjudication, pretesting, and documentation) for cross-cultural survey research (ccsg.isr.umich.edu/chapters/translation/overview)^49^. The data that supports this research are publicly available via the Center for Open Science (COS, https://www.cos.io/gfs). Details about the GFS study methodology and survey development were reported elsewhere^45^ (see also https://osf.io/y3t6m).

The present study used data from all participants in Wave 1 of GFS (N=202,898). Poststratification and nonresponse adjustments were performed to ensure the sample was representative of the adult population in each country^45,50^. Ethical approval was granted by the institutional review boards at Baylor University and Gallup, and all participants provided informed consent. All methods were carried out in accordance with relevant ethical guidelines and regulations.

### Assessment of Community Participation

#### Secular community participation

The participants reported their participation in secular community groups in response to this question: “How often do you participate in groups that are not religious, such as book clubs, sports, or political organizations?”. Response options include “never”, “a few times a year”, “one to three times a month”, “once a week”, and “more than once a week”. Because prior studies suggest that at least once/week of community participation is often associated with improved health and well-being outcomes^18,37,51^, we dichotomized the responses as at least once/week vs. less than once/week or never.

#### Religious service attendance

Frequency of religious service attendance was assessed with one questionnaire item: “How often do you attend religious services?”. Response options include “never”, “a few times a year”, “one to three times a month”, “once a week”, and “more than once a week”. Similar to secular community participation (see above), we dichotomized the frequency of religious service attendance as at least once/week vs. less than once/week or never.

### Assessment of Childhood Factors

We investigated a range of individual-, interpersonal-, and familial-level childhood factors, as described below.

#### Relationship with parents

Relationship with one’s parents in childhood was assessed with the question: “Please think about your relationship with your mother/father when you were growing up. In general, would you say that relationship was very good, somewhat good, somewhat bad, or very bad?”. Relationships with mother and father were queried separately. To reduce collinearity, in regression models the variables of relationship with mother and father were both dichotomized as “very/somewhat good” and “very /somewhat bad”. Responding “Does not apply” to either parental relationship indicator was treated as a dichotomous control variable for respondents who did not have a mother or father due to death or absence.

#### Parents’ marital status

Parents’ marital status was assessed with the question: “Were your parents married to each other when you were around 12 years old?”. Response options included “married”, “divorced”, “never married”, and “one or both of them had died”.

#### Subjective financial status

Participants reported the financial status of their family as it was when they were growing up, in response to the question: “which of these phrases comes closest to your own feelings about your family’s household income when you were growing up, such as when you were around 12 years old?”. Response options included “lived comfortably”, “got by”, “found it difficult”, and “found it very difficult”.

#### Childhood abuse

Childhood abuse was assessed with the question: “Were you ever physically or sexually abused when you were growing up?”. Responses included “yes” and “no”. This information was not obtained in Israel due to restrictions on asking such questions.

#### Felt like an outsider in family

Participants’ sense of family belonging during childhood was assessed with the question: “When you were growing up, did you feel like an outsider in your family”. Response options included “yes” and “no”.

#### Self-rated health

Participants reported their health status during childhood in response to the question: “In general, how was your health when you were growing up? Was it excellent, very good, good, fair, or poor?”

#### Immigration status

Immigration status was assessed with the question “Were you born in this country?” The responses include “yes” and “no”.

#### Religious service attendance in childhood

Frequency of religious service attendance during childhood was assessed with a question: “How often did you attend religious services or worship at a temple, mosque, shrine, church or other religious building when you were around 12 years old”. The response options range from 1 (at least once a week) to 4 (never).

#### Birth Year

Participants reported their current age (in years), and their birth years were calculated as the year of data collection minus their current age.

#### Gender

Participants reported their gender, and the response options include “male”, “female”, and “other gender identities”.

#### Religious affiliation

Religious affiliation in childhood was measured with the question: “What was your religion when you were 12 years old”. Response options include 15 major religions (e.g., Christianity, Islam, Hinduism, Buddhism, Judaism etc.), “some other religion”, and “No religion/Atheist/Agnostic”. The response categories varied as appropriate for each country, and this variable was included in the country-specific analyses only but not in the meta-analysis across countries^52^. To reduce data sparsity, in regression models the response categories of religious affiliation with a prevalence <3% were collapsed. “No religion/Atheist/Agnostic” was used as the reference group when at least 3% of the observed sample within the country endorsed this category; otherwise, the most prominent religious group was used as the reference category.

#### Race and ethnicity

Race and ethnicity were assessed in most but not all countries (Germany, Japan, Spain, and Sweden had restriction on collecting such data). Response options varied as appropriate for each country, so this variable was included in the country-specific analyses only but not in the meta-analysis across countries. To reduce data sparsity, in regression models race and ethnicity was dichotomized as racial and ethnic plurality (the category with the largest proportion) and minority (collapsing other categories) in each country.

### Statistical Analyses

The descriptive analyses examined the distribution of childhood factors in individual countries and in the total combined sample. The results were weighted to be nationally representative within each country.

In country-specific analyses, a weighted modified Poisson regression model with complex survey adjusted standard errors was used to regress community participation (at least once/week vs. less than once/week or never) on *all childhood factors simultaneously* to evaluate their associations with community participation independently from each other. Secular and religious community participation were examined separately as the outcome variable. A Wald-type test was conducted to obtain a global (joint) test of the effect of all categories within a childhood factor, resulting in a single global p-value for each childhood factor.

Random effects meta-analysis^53,54^ was then conducted to pool the country-specific regression coefficients, along with confidence intervals, the proportion of calibrated effects by threshold (above a risk ratio [RR] of 1.10 or below a RR of 0.90), heterogeneity estimate (τ), and I^2^ for evidence concerning variation within a given childhood predictor category across countries^55^. Forest plots of estimates are presented in the supplement. Religious affiliation and race and ethnicity (when available) were used as control variables within country (their coefficients are reported in the supplement), but these coefficients themselves were not included in the meta-analyses because their response categories varied by country. A pooled global p-value^56^ was reported across countries to evaluate if the association between each childhood factor and adult community participation varied within at least one country. Bonferroni corrected p-value thresholds are provided based on the number of childhood factors that we examined^57,58^.

All analyses were conducted in R^59^, and the random effects meta-analysis used the metafor package^60^. For each childhood predictor, in both the meta-analyses and country-specific analyses we calculated E-values to evaluate sensitivity of the observed associations to potential unmeasured confounding. An E-value is the minimum strength of the association an unmeasured confounder must have with both the outcome variable and the predictor, above and beyond all measured covariates, for an unmeasured confounder to explain away the observed predictor-outcome association^61^. As a supplementary analysis, we performed a population weighted meta-analysis to pool the country-specific estimates. The population weighted meta-analysis effectively treats each person equally, rather than treating each country equally as the random effects meta-analysis does. All analyses were pre-registered with COS prior to data access, with only slight subsequent modification in covariate adjustments due to multicollinearity (https://doi.org/10.17605/OSF.IO/VM7W9and https://doi.org/10.17605/OSF.IO/6W392); all code to reproduce analyses are openly available in an online repository^47^.

### Missing Data

In the total sample combined across countries, 0.31% of participants had missing data on secular community participation and 0.35% had missing data on religious service attendance.

Missing data on childhood factors ranged from 0.01% to 9.78% (except race/ethnicity, which was not measured in some countries).We conducted multivariate imputation by chained equations (5 imputed datasets created) to impute missing data on all variables in each country separately^62–64^. We included sampling weights in the imputation models to account for specific-variable missingness that may have been related to probability of inclusion in the study^50^.

### Accounting for Complex Sampling Design

The GFS used different sampling schemes across countries based on availability of existing panels and recruitment needs^65^. All analyses in the present study accounted for the complex survey design components by including weights, primary sampling units, and strata. Additional methodological details are reported elsewhere^50,65^.

## RESULTS

### Descriptive Analyses

Table 1 presents the distribution of childhood factors in the weighted cross-national sample. Around 17.7% of the participants engaged in secular groups at least once/week (a measure referred to hereafter as “weekly^+^”) and 32% attended religious services weekly^+^ in adulthood. In the weighted sample, most participants reported their childhood experiences as follows: very/somewhat good relationships with mother (89%) and father (80%), parents married (75%), lived comfortably financially or got by (76%), no experience of child abuse (82%), did not feel like an outsider in the family (84%), had excellent or very good health (64%), and were native-born (94%). A large proportion (41%) attended religious services weekly^+^ at age 12. There were fewer participants from the earliest birth cohorts (born in or before year 1953) than those from more recent birth cohorts. The weighted sample included a similar proportion of women (51%) and men (49%), and a small percentage (0.3%) who identified as other gender. Participant characteristics in each individual country are reported in Supplementary Table S1a to S22a.

**Table 1.** Distribution of Childhood Factors in the Overall Sample Combined Across 22 Countries (N=202,898, with Nationally Representative Sample within Each Country)

| **Childhood factors** | **N (%)** |
| --- | --- |
| **Relationship with mother** |  |
| Very good | 127,836 (63%) |
| Somewhat good | 52,439 (26%) |
| Somewhat bad | 11,060 (5.5%) |
| Very bad | 4,642 (2.3%) |
| Does not apply | 5,965 (2.9%) |
| (Missing) | 956 (0.5%) |
| **Relationship with father** |  |
| Very good | 107,742 (53%) |
| Somewhat good | 55,714 (27%) |
| Somewhat bad | 15,807 (7.8%) |
| Very bad | 8,278 (4.1%) |
| Does not apply | 13,985 (6.9%) |
| (Missing) | 1,372 (0.7%) |
| **Parent marital status** |  |
| Parents married | 152,001 (75%) |
| Divorced | 17,726 (8.7%) |
| Parents were never married | 15,534 (7.7%) |
| One or both parents had died | 7,794 (3.8%) |
| (Missing) | 9,843 (4.9%) |
| **Subjective financial status of family when growing up** |  |
| Lived comfortably | 70,861 (35%) |
| Got by | 82,905 (41%) |
| Found it difficult | 35,852 (18%) |
| Found it very difficult | 12,606 (6.2%) |
| (Missing) | 674 (0.3%) |
| **Experienced Abuse** |  |
| Yes | 29,139 (14%) |
| No | 167,279 (82%) |
| (Missing) | 6,479 (3.2%) |
| **Felt like an outsider in the family** |  |
| Yes | 28,732 (14%) |
| No | 170,577 (84%) |
| (Missing) | 3,589 (1.8%) |
| **Self-rated health when growing up** |  |
| Excellent | 67,121 (33%) |
| Very good | 63,086 (31%) |
| Good | 47,378 (23%) |
| Fair | 19,877 (9.8%) |
| Poor | 4,906 (2.4%) |
| (Missing) | 530 (0.3%) |
| **Immigration status** |  |
| Born in this country | 190,998 (94%) |
| Born in another country | 9,791 (4.8%) |
| (Missing) | 2,110 (1.0%) |
| **Frequency of religious service attendance (age 12)** |  |
| At least 1/week | 83,237 (41%) |
| 1-3/month | 33,308 (16%) |
| <1/month | 36,928 (18%) |
| Never | 47,445 (23%) |
| (Missing) | 1,980 (1.0%) |
| **Year of birth** |  |
| 1998-2005; age 18-24 | 27,007 (13%) |
| 1993-1998; age 25-29 | 20,700 (10%) |
| 1983-1993; age 30-39 | 40,256 (20%) |
| 1973-1983; age 40-49 | 34,464 (17%) |
| 1963-1973; age 50-59 | 31,793 (16%) |
| 1953-1963; age 60-69 | 27,763 (14%) |
| 1943-1953; age 70-79 | 16,776 (8.3%) |
| 1943 or earlier; age 80+ | 4,119 (2.0%) |
| (Missing) | 20 (<0.1%) |
| **Gender** |  |
| Male | 98,411 (49%) |
| Female | 103,488 (51%) |
| Other | 602 (0.3%) |
| (Missing) | 397 (0.2%) |
| **Country** |  |
| Argentina | 6,724 (3.3%) |
| Australia | 3,844 (1.9%) |
| Brazil | 13,204 (6.5%) |
| Egypt | 4,729 (2.3%) |
| Germany | 9,506 (4.7%) |
| Hong Kong (S.A.R of China) | 3,012 (1.5%) |
| India | 12,765 (6.3%) |
| Indonesia | 6,992 (3.4%) |
| Israel | 3,669 (1.8%) |
| Japan | 20,543 (10%) |
| Kenya | 11,389 (5.6%) |
| Mexico | 5,776 (2.8%) |
| Nigeria | 6,827 (3.4%) |
| Philippines | 5,292 (2.6%) |
| Poland | 10,389 (5.1%) |
| South Africa | 2,651 (1.3%) |
| Spain | 6,290 (3.1%) |
| Sweden | 15,068 (7.4%) |
| Tanzania | 9,075 (4.5%) |
| Turkey | 1,473 (0.7%) |
| United Kingdom | 5,368 (2.6%) |
| United States | 38,312 (19%) |

Note. S.A.R. = Special Administrative Region.

### Childhood Factors and Adult Secular Community Participation: Pooled Estimates Across Countries

On average across countries, most childhood factors that we examined were associated with adult secular community participation (Table 2). The global p-values for most childhood factors (except for relationship with mother and father, subjective financial status of family, and immigration status) were below the Bonferroni corrected threshold for statistical significance (*p*<0.0045), suggesting that each of these childhood factors was associated with adult secular community participation in at least one of the 22 countries. First, compared to participants with good self-rated health in childhood, those with excellent or very good self-rated childhood health had a higher likelihood of adult weekly^+^ secular community participation by 1.12-fold (risk ratio [RR] =1.12, 95% confidence interval [CI]: 1.03, 1.22) and 1.07-fold (RR=1.07, 95% CI: 1.01, 1.15), respectively. It is worth noting, however, that having poor vs. good self-rated childhood health was also positively associated with adult weekly^+^ secular community participation to a similar extent (RR=1.12, 95% CI: 1.00, 1.25). Second, religious service attendance in childhood was positively associated with adult weekly^+^ secular community participation in a monotonic fashion. For instance, those attending religious services weekly^+^ in childhood were 1.54 times more likely to participate in secular groups weekly^+^ when they grew up into adults, as compared to those never attending services (RR =1.54, 95% CI: 1.29, 1.84). Third, an earlier birth year/older current age was inversely associated with the likelihood of weekly^+^ secular community participation in a nearly monotonic pattern (e.g., RR _birth cohort 1953-1963 vs. 1998-2005_=0.70, 95% CI: 0.62, 0.78). Fourth, women and those identified as other gender were less likely to engage in secular groups weekly^+^ than men (e.g., RR _female vs. male_=0.78, 95% CI: 0.72, 0.85). Fifth, somewhat unexpectedly, the experience of child abuse (RR=1.12, 95% CI: 1.05, 1.21) and feeling like an outsider in the family (RR=1.09, 95% CI: 1.01, 1.17) were each associated with a higher likelihood of adult weekly^+^ secular community participation on average across countries. To a lesser extent, several other childhood factors were also associated with adult secular community participation. Specifically, those who lived comfortably financially when growing up were more likely to participate in secular communities in adulthood by 1.09-fold (95% CI: 1.05, 1.12) than those who reported “got by”, although the global *p*-value did not reach <.05 after Bonferroni correction. Next, while the association between very good/somewhat good (vs. very bad/somewhat bad) relationship with father in childhood and adult secular community participation did not include the null value (RR=1.08, 95% CI: 1.03, 1.13), the global p-value did not reach p<.05. Likewise, there was little evidence suggesting that relationship with mother was associated with adult secular community participation. It is worth noting though there are only a small number of participants who reported “very/somewhat bad relationships” with their parents in this sample (<5% and <10% reported very/somewhat bad relationship with mother and father in most countries). Finally, although the association for immigration status included the null value, the global *p*-value reached p<.05 before Bonferroni correction, indicating that immigration status might be associated with secular community participation in at least one country, though possibly in different directions, and not a clear average association across countries.

**Table 2.**
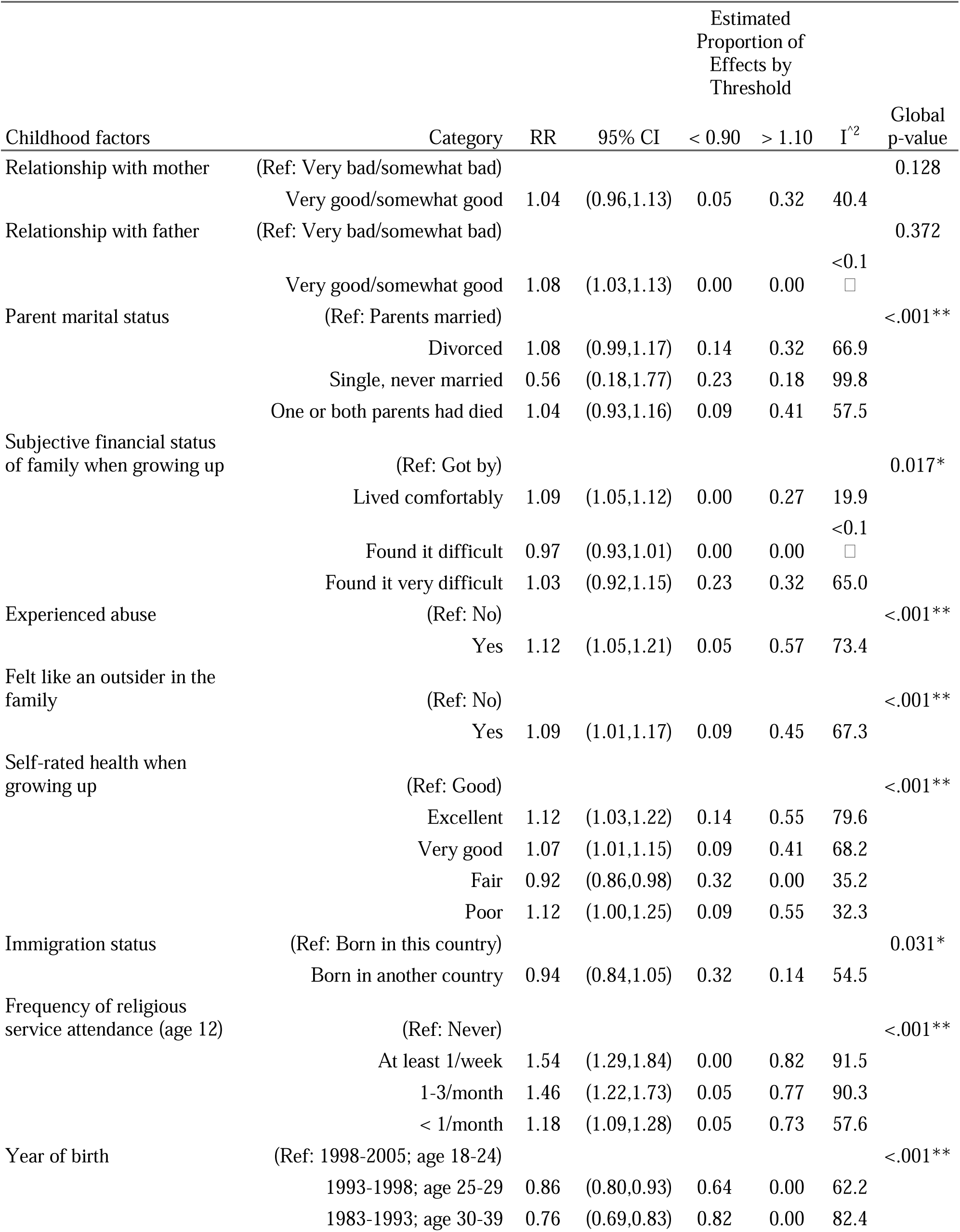
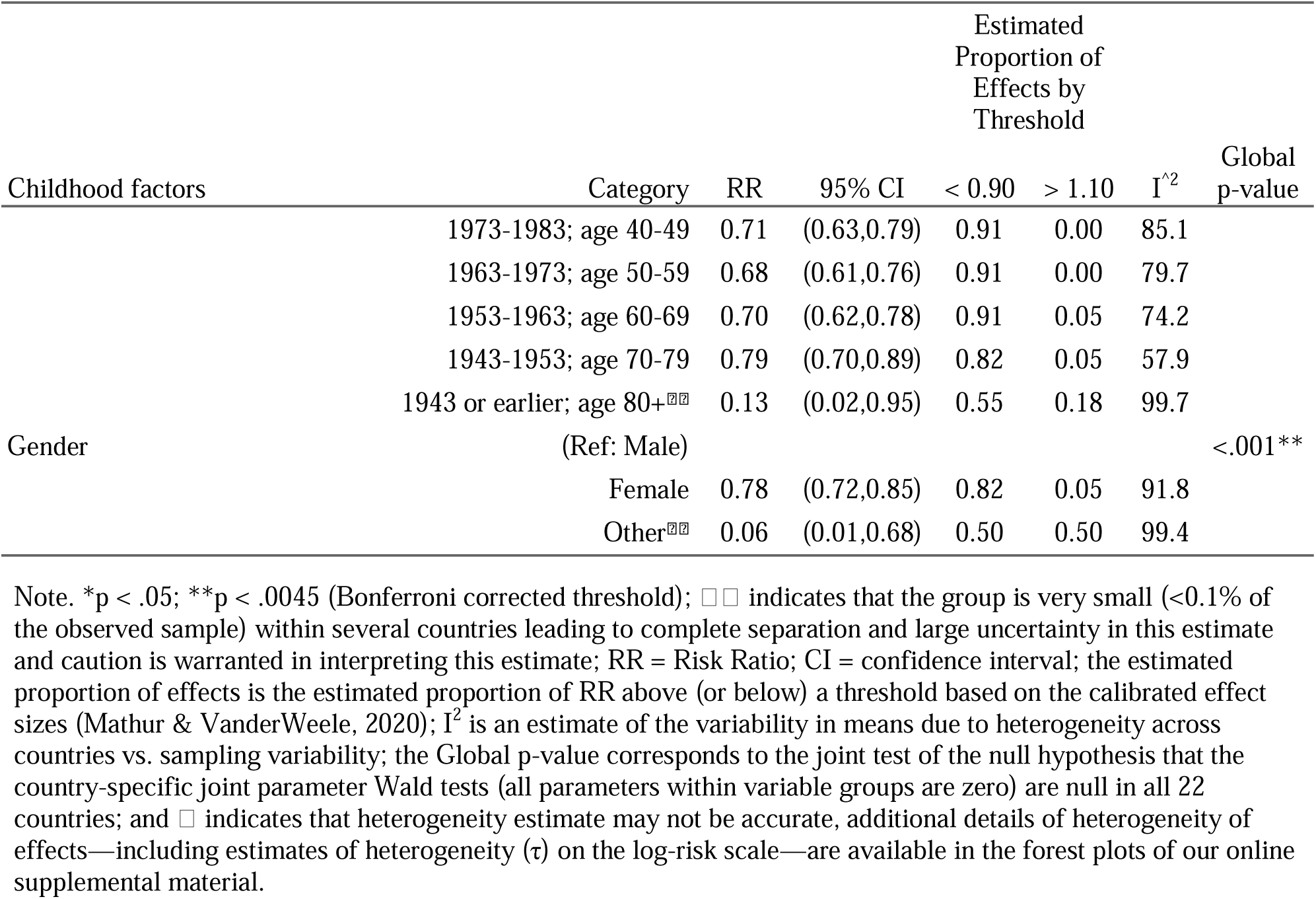
Random Effects Meta-Analysis of Association between Childhood Factors and Secular Community Participation in Adulthood (N=202,898).

There is notable evidence for heterogeneity across countries, and the I^2^ estimates for certain categories of parent marital status, religious service attendance at age 12, year of birth, and gender were larger (above 90) than other childhood factor categories. This suggests that variability in the estimates for these childhood categories is more likely due to heterogeneity across countries than to sampling variability. We comment further on cross-country variation below.

### Childhood Factors and Adult Secular Community Participation: Country-Specific Analyses

Some associations with particular childhood predictors of secular community participation were fairly consistent across countries. In most of the 22 countries, religious service attendance at age 12, birth year, and gender were each associated with adult secular community participation with a global *p*-value <.05 (Supplementary Tables S1b to S22b and Supplementary Figures S1 to S27). First, in 17 countries frequent religious service attendance at age 12 was associated with a substantially higher likelihood of adult weekly^+^ secular community participation, with the largest effect sizes observed in some of the most secular countries/territories in the world (Germany, Hong Kong, Japan, Sweden, United Kingdom, and Spain)^66^. For instance, in Hong Kong participants who attended services weekly^+^ (vs. never) at age 12 had a higher likelihood of adult weekly^+^ secular group participation by 5.57-fold (95% CI: 3.50, 8.86). Second, birth year was also associated with adult community participation across 18 economically and culturally diverse countries. For instance, almost every earlier (vs. the most recent) birth cohort reported a lower likelihood of adult weekly^+^ secular community participation in Argentina, Egypt, Indonesia, India, and Kenya. Third, gender was associated with adult secular community participation in almost all countries except Brazil and Spain. Consistently across these countries (except Japan), male (vs. female) participants were more likely to engage in secular communities weekly^+^.

Other patterns were present in many but not all countries. In around half of the counties, subjective financial status of family when growing up and self-rated childhood health were each associated with adult secular community participation. Specifically, living comfortably (vs. got by) financially in childhood was positively associated with weekly^+^ secular community participation in Mexico, the United Kingdom, the United States, Indonesia, and Sweden, with the RRs ranging from 1.11 in Sweden to 1.23 in Mexico. It is, however, worth noting that those who reported difficult or poor (vs. got by) financial status when growing up were also more likely to participate in secular communities weekly^+^ in Philippines (RR=1.38, 95% CI: 1.04, 1.83), Sweden (RR=1.55, 95% CI: 1.03, 2.35), and Poland (RR=1.69, 95% CI: 1.01, 2.83). Next, self-rated childhood health was associated with adult secular community participation in several predominantly high-income or upper-middle income countries/territories. Specifically, excellent/very good (vs. good) childhood health was positively associated with adult secular community participation in Argentina, Japan, Indonesia, Australia, Hong Kong, Mexico, Sweden, and Germany, whereas fair (vs. good) childhood health was associated with a lower likelihood of adult weekly^+^ secular community participation in South Africa, Japan, Sweden, Philippine, and Brazil. It is worth noting that poor (vs. good) childhood health was also positively associated with adult secular community participation in Indonesia (RR=1.82, 95% CI: 1.20, 2.76), and maybe also in Tanzania, Poland, and Spain (although the associations were only around borderline statistical significance in these 3 latter countries).

In a handful of countries, the childhood factors of abuse, feeling like an outsider in the family, relationship with parents, parent marital status, and immigration status were also associated with adult secular community participation. For instance, somewhat unexpectedly, experiencing abuse was consistently associated with an increased likelihood of weekly^+^ secular community participation by 1.13 to 1.53-fold in several African and Asian countries (including South Africa, India, Japan, Kenya, Hong Kong, Nigeria). Likewise, participants who felt like an outsider in the family were more likely to report weekly^+^ secular community participation by up to a 1.28-fold across several Asian, European, and American countries (including Hong Kong, Japan, Spain, United Kingdom, and Argentina). It is worth noting that neither child abuse nor outsider growing up was strongly associated with a lower likelihood of adult weekly^+^ secular community participation in any of the 22 countries. Being an immigrant was positively associated with adult secular community participation in Poland, but the association was negative in Spain and Hong Kong, and the association was null in other countries. The country-specific estimates for religious affiliation and race/ethnicity are also provided in Supplementary Tables S1b to S22b.

### Childhood Factors Associated with Religious Service Attendance in Adulthood: Pooled Estimates Across Countries

The childhood factors of religious service attendance at age 12, and certain categories of birth cohorts, self-rated health, parent marital status, and relationship with father were strongly associated with adult weekly^+^ religious service attendance on average across countries (Table 3). The global p-values for most childhood factors (except for relationship with parents, parent marital status, and subjective financial status of family) were below the Bonferroni corrected threshold for statistical significance (*p*<0.0045), indicating that each of these childhood factors was associated with adult service attendance in at least one of the 22 countries. Specifically, the strongest evidence was with religious service attendance in childhood, whereby frequent service attendance at age 12 was associated with a substantially higher likelihood of adult weekly^+^ service attendance in a monotonic pattern. For instance, compared to those never attending services in childhood, those attending services weekly^+^ were more likely to continue doing so in adulthood by 3.19-fold (RR=3.19, 95% CI: 2.03, 5.01). Some earlier birth cohorts (with current age≥50 years) were also more likely to attend services weekly^+^ in adulthood (e.g., RR _birth cohort 1943 or earlier vs. 1998-2005_=1.22, 95% CI: 1.01, 1.47), compared to the most recent birth cohort. In addition, having very good (vs. good) self-rated health when growing up was associated with a higher likelihood of adult weekly^+^ service attendance by 1.03-fold (95% CI: 1.00, 1.08).

**Table 3.** Random Effects Meta-Analysis of Association between Childhood Factors and Religious Service Attendance in Adulthood (N=202,898).

| Childhood factors | Category | RR | 95% CI | Estimated Proportion of Effects by Threshold |  | I<sup>2</sup> | Global p-value |
| --- | --- | --- | --- | --- | --- | --- | --- |
|  |  |  |  | < 0.90 | > 1.10 |  |  |
| Relationship with mother | (Ref: Very bad/somewhat bad) |  |  |  |  |  | 0.282 |
|  | Very good/somewhat good | 1.04 | (0.99,1.09) | 0.00 | 0.27 | 28.1 |  |
| Relationship with father | (Ref: Very bad/somewhat bad) |  |  |  |  |  | 0.012* |
|  | Very good/somewhat good | 1.11 | (1.06,1.16) | 0.00 | 0.59 | 48.6 |  |
| Parent marital status | (Ref: Parents married) |  |  |  |  |  | 0.007* |
|  | Divorced | 0.88 | (0.81,0.96) | 0.50 | 0.05 | 80.1 |  |
|  | Single, never married | 0.95 | (0.90,1.00) | 0.18 | 0.05 | 49.2 |  |
|  | One or both parents had died | 0.96 | (0.91,1.00) | 0.14 | 0.00 | 39.9 |  |
| Subjective financial status of family when growing up | (Ref: Got by) |  |  |  |  |  | 0.210 |
|  | Lived comfortably | 1.01 | (0.98,1.03) | 0.00 | 0.00 | 52.0 |  |
|  | Found it difficult | 1.02 | (0.99,1.05) | 0.00 | 0.00 | 38.6 |  |
|  | Found it very difficult | 1.02 | (1.00,1.05) | 0.00 | 0.00 | 4.9 |  |
| Experienced abuse | (Ref: No) |  |  |  |  |  | <.001** |
|  | Yes | 0.99 | (0.94,1.05) | 0.14 | 0.19 | 83.4 |  |
| Felt like an outsider in the family | (Ref: No) |  |  |  |  |  | <.001** |
|  | Yes | 1.05 | (0.95,1.15) | 0.09 | 0.23 | 94.1 |  |
| Self-rated health when growing up | (Ref: Good) |  |  |  |  |  | <.001** |
|  | Excellent | 1.03 | (0.97,1.08) | 0.05 | 0.23 | 84.3 |  |
|  | Very good | 1.03 | (1.00,1.08) | 0.00 | 0.27 | 69.5 |  |
|  | Fair | 0.97 | (0.92,1.02) | 0.18 | 0.00 | 68.1 |  |
|  | Poor | 1.04 | (0.93,1.16) | 0.18 | 0.23 | 78.0 |  |
| Immigration status | (Ref: Born in this country) |  |  |  |  |  | <.001** |
|  | Born in another country | 1.05 | (0.96,1.14) | 0.09 | 0.32 | 58.2 |  |
| Frequency of religious service attendance (age 12) | (Ref: Never) |  |  |  |  |  | <.001** |
|  | At least 1/week | 3.19 | (2.03,5.01) | 0.00 | 0.86 | 99.4 |  |
|  | 1-3/month | 2.24 | (1.51,3.33) | 0.05 | 0.77 | 99.0 |  |
|  | < 1/month | 1.19 | (1.00,1.42) | 0.18 | 0.50 | 92.4 |  |
| Year of birth | (Ref: 1998-2005; age 18-24) |  |  |  |  |  | <.001** |
|  | 1993-1998; age 25-29 | 1.02 | (0.96,1.08) | 0.05 | 0.32 | 81.8 |  |
|  | 1983-1993; age 30-39 | 1.03 | (0.96,1.10) | 0.18 | 0.32 | 91.1 |  |
|  | 1973-1983; age 40-49 | 1.04 | (0.97,1.11) | 0.18 | 0.36 | 89.0 |  |
|  | 1963-1973; age 50-59 | 1.11 | (1.01,1.21) | 0.09 | 0.41 | 92.2 |  |

|  |  | RR | 95% CI | Estimated Proportion of Effects by Threshold |  | I<sup>2</sup> | Global p-value |
| --- | --- | --- | --- | --- | --- | --- | --- |
|  |  |  |  | < 0.90 | > 1.10 |  |  |
| Childhood factors | Category |  |  |  |  |  |  |
|  | 1953-1963; age 60-69 | 1.11 | (0.99,1.25) | 0.18 | 0.55 | 92.1 |  |
|  | 1943-1953; age 70-79 | 1.10 | (0.96,1.25) | 0.18 | 0.59 | 86.8 |  |
|  | 1943 or earlier; age 80+ | 1.22 | (1.01,1.47) | 0.14 | 0.64 | 83.4 |  |
| Gender | (Ref: Male) |  |  |  |  |  | <.001** |
|  | Female | 0.98 | (0.89,1.07) | 0.32 | 0.32 | 97.9 |  |
|  | Other | 0.17 | (0.02,1.12) | 0.50 | 0.28 | 99.9 |  |

Note. \*p < .05; \*\*p < .0045 (Bonferroni corrected threshold); RR = Risk Ratio; CI = confidence interval; the estimated proportion of effects is the estimated proportion of RR above (or below) a threshold based on the calibrated effect sizes (Mathur & VanderWeele, 2020); I<sup>2</sup> is an estimate of the variability in means due to heterogeneity across countries vs. sampling variability; and the Global p-value corresponds to the joint test of the null hypothesis that the country-specific joint parameter Wald tests (all parameters within variable groups are zero) are null in all 22 countries.

Compared to participants with married parents, those whose parents were unmarried during childhood were less likely to attend services weekly^+^ when they grew up into adults, although the global *p*-value reached <.05 only before Bonferroni correction. For instance, those with divorced parents (vs. with married parents) were less likely to attend services weekly^+^ in adulthood (RR=0.88, 95% CI: 0.81, 0.96). Likewise, participants who recalled having very/somewhat bad (vs. very/somewhat good) relationship with father were less likely to attend services weekly^+^ in adulthood (RR=0.88, 95% CI: 0.81, 0.96), although the global *p*-value was again below <.05 only before Bonferroni correction. In addition, the associations for abuse, feeling like an outsider in the family, immigration status, and gender all included the null value but the global *p*-values were <.001, indicating that each of these childhood factors were associated with adult service attendance in at least one country but not clearly on average across countries. There was, however, little evidence that the childhood factors of relationship with mother and subjective financial status were associated with adult religious service attendance.

There was evidence of heterogeneity in effect sizes across countries; also the I^2^ estimates for certain categories of several childhood factors including feeling like an outsider in the family, religious service attendance at age 12, year of birth, and gender were relatively large (above 90) compared to other childhood categories. This indicates that variability in the estimates for these categories is more likely due to heterogeneity across countries than to sampling variability, which we comment on below.

### Childhood Factors Associated with Religious Service Attendance in Adulthood: Country-Specific Analyses

Consistently across all 22 countries, frequent religious service attendance in childhood was associated with a substantially higher likelihood of weekly^+^ service attendance in adulthood, with the strongest association observed in some of the most secular countries in the world (e.g., Sweden, Germany, Japan, United Kingdom, and Spain; Supplementary Tables S1b to S22b and Supplementary Figures S1 to S27). For instance, in Sweden participants who attended services weekly^+^ in childhood were more likely to continue doing so in adulthood by 34.34-fold (95% CI: 22.94, 51.40). Birth year was also associated with adult service attendance across 15 diverse countries, but the specific patterning varied widely by country. For instance, compared to those from the most recent birth cohort (1998-2005), every earlier birth cohort reported a higher likelihood of adult weekly^+^ service attendance nearly monotonically in Brazil and India, whereas only some older birth cohorts had a higher likelihood of weekly^+^ attendance in Indonesia, Kenya, Mexico, Poland, the United Kingdom, and the United States. Conversely, almost all earlier birth cohorts in Sweden and some of the earlier birth cohorts in Israel and Egypt reported a lower likelihood of adult weekly^+^ service attendance. In addition, gender was associated with adult service attendance in 19 countries, but in different directions. Specifically, female (vs. male) participants were more likely to attend services weekly^+^ in predominantly Christian countries (Argentina, Kenya, Poland, Philippine, Tanzania, United States, and Brazil), whereas they were less likely to attend services weekly^+^ in countries that are considered more secular (Germany, Hong Kong, Japan, Spain, and Sweden) or in countries dominated by Muslims or Hindus (Egypt, India, and Indonesia).

In 9 of 22 countries, child abuse was associated with adult service attendance, but not in a consistent direction. Experiencing abuse was associated with a higher likelihood of weekly^+^ attendance in Sweden, Hong Kong, Egypt, and India (RR ranged from 1.14 in India to 1.38 in Sweden), but a lower likelihood of weekly^+^ attendance in Poland, Turkey, South Africa, Nigeria, and Kenya (RR ranged from 0.95 in Kenya to 0.70 in Poland). Likewise, participants who felt like an outsider in the family when growing up reported a higher likelihood of adult weekly^+^ attendance in Hong Kong, Japan, Spain, and the United Kingdom (RR ranged from 1.32 in Spain to 2.22 in Japan), but the pattern was reversed in Kenya and the United States (RR ranged from 0.95 in Kenya to 0.68 in United States). Parental marital status was also associated with adult service attendance in 8 countries, but the specific patterning varied across countries.

The childhood factors of subjective financial status, self-rated health, relationship with parents, and immigration status were each associated with adult service attendance in a handful of countries only. For example, as compared to those reporting “got by” financially in childhood, participants reporting difficult/very difficult financial status were more likely to attend religious services weekly^+^ during adulthood in the United States and Mexico, and those who lived comfortably financially in childhood were less likely to attend services weekly^+^ in Poland, but more likely to in Spain and the Philippines. As another example, excellent/very good (vs. good) self-rated childhood health was related to a higher likelihood of weekly^+^ service attendance in adulthood in Hong Kong, Indonesia, and Israel, whereas fair/poor (vs. good) self-rated childhood health was also positively associated with adult service attendance in Israel. The country-specific estimates for childhood religious affiliation and race/ethnicity were also reported in Supplementary Tables S1b to S22b.

### Additional Analyses

Table 4 (for adult secular community participation) and Table 5 (for adult service attendance) present the “E-values” for assessing robustness of the meta-analyzed associations between childhood experiences and adult community participation to potential unmeasured confounding. For example, to explain away the association between weekly^+^ (vs. never) religious service attendance at age 12 and adult secular community participation (RR=1.54), an unmeasured confounder associated with increased likelihood of weekly^+^ service attendance at age 12 and weekly^+^ secular community participation in adulthood by risk ratios of 2.45 each, above and beyond all measured covariates, could suffice but weaker joint confounder associations could not (Table 4). Similarly, to shift the confidence interval to include the null value, an unmeasured confounder associated with increased likelihood of weekly^+^ service attendance at age 12 and weekly^+^ secular community participation in adulthood by risk ratios of 1.90-fold each at minimum could suffice. The E-values for country-specific estimates are provided in Supplementary Tables S1c to S22c.

**Table 4.** Sensitivity of Meta-Analyzed Associations between Childhood Factors and Secular Community Participation in Adulthood to Potential Unmeasured Confounding.

| Variable | Category | E-value for Estimate | E-value for 95% CI |
| --- | --- | --- | --- |
| Relationship with mother | (Ref: Very bad/somewhat bad) |  |  |
|  | Very good/somewhat good | 1.25 | 1.00 |
| Relationship with father | (Ref: Very bad/somewhat bad) |  |  |
|  | Very good/somewhat good | 1.38 | 1.22 |
| Parent marital status | (Ref: Parents married) |  |  |
|  | Divorced | 1.36 | 1.00 |
|  | Single, never married | 2.94 | 1.00 |
|  | One or both parents had died | 1.24 | 1.00 |
| Subjective financial status of family when growing up | (Ref: Got by) |  |  |
|  | Lived comfortably | 1.39 | 1.28 |
|  | Found it difficult | 1.21 | 1.00 |
|  | Found it very difficult | 1.19 | 1.00 |
| Experienced abuse | (Ref: No) |  |  |
|  | Yes | 1.50 | 1.26 |
| Felt like an outsider in the family | (Ref: No) |  |  |
|  | Yes | 1.40 | 1.14 |
| Self-rated health when growing up | (Ref: Good) |  |  |
|  | Excellent | 1.48 | 1.20 |
|  | Very good | 1.36 | 1.08 |
|  | Fair | 1.40 | 1.15 |
|  | Poor | 1.47 | 1.00 |
| Immigration status | (Ref: Born in this country) |  |  |
|  | Born in another country | 1.33 | 1.00 |
| Frequency of religious service attendance (age 12) | (Ref: Never) |  |  |
|  | At least 1/week | 2.45 | 1.90 |
|  | 1-3/month | 2.27 | 1.74 |
|  | < 1/month | 1.64 | 1.39 |
| Year of birth | (Ref: 1998-2005; age 18-24) |  |  |
|  | 1993-1998; age 25-29 | 1.59 | 1.36 |
|  | 1983-1993; age 30-39 | 1.96 | 1.69 |
|  | 1973-1983; age 40-49 | 2.18 | 1.84 |
|  | 1963-1973; age 50-59 | 2.28 | 1.94 |
|  | 1953-1963; age 60-69 | 2.23 | 1.87 |
|  | 1943-1953; age 70-79 | 1.85 | 1.48 |
|  | 1943 or earlier; age 80+<sup>□</sup> | 15.17 | 1.30 |
| Gender | (Ref: Male) |  |  |
|  | Female | 1.88 | 1.63 |
|  | Other<sup>□</sup> | 33.45 | 2.32 |

Note. N = 202,898; the E-value is the minimum strength of the association an unmeasured confounder must have with both the outcome and the predictor, above and beyond all measured covariates, for an unmeasured confounder to explain away an association (VanderWeele & Ding, 2017, p. 269-270); and <sup>□</sup> indicates that the group is very small (<0.1% of the observed sample) within several countries potentially leading to complete separation and large uncertainty in this estimate and caution is warranted in interpreting this estimate.

**Table 5.** Sensitivity of Meta-Analyzed Associations between Childhood Factors and Religious Service Attendance in Adulthood to Potential Unmeasured Confounding.

| Variable | Category | E-value for Estimate | E-value for 95% CI |
| --- | --- | --- | --- |
| Relationship with mother | (Ref: Very bad/somewhat bad) |  |  |
|  | Very good/somewhat good | 1.25 | 1.00 |
| Relationship with father | (Ref: Very bad/somewhat bad) |  |  |
|  | Very good/somewhat good | 1.46 | 1.31 |
| Parent marital status | (Ref: Parents married) |  |  |
|  | Divorced | 1.52 | 1.25 |
|  | Single, never married | 1.30 | 1.07 |
|  | One or both parents had died | 1.27 | 1.00 |
| Subjective financial status of family when growing up | (Ref: Got by) |  |  |
|  | Lived comfortably | 1.08 | 1.00 |
|  | Found it difficult | 1.16 | 1.00 |
|  | Found it very difficult | 1.18 | 1.00 |
| Experienced abuse | (Ref: No) |  |  |
|  | Yes | 1.10 | 1.00 |
| Felt like an outsider in the family | (Ref: No) |  |  |
|  | Yes | 1.27 | 1.00 |
| Self-rated health when growing up | (Ref: Good) |  |  |
|  | Excellent | 1.19 | 1.00 |
|  | Very good | 1.22 | 1.00 |
|  | Fair | 1.21 | 1.00 |
|  | Poor | 1.23 | 1.00 |
| Immigration status | (Ref: Born in this country) |  |  |
|  | Born in another country | 1.27 | 1.00 |
| Frequency of religious service attendance (age 12) | (Ref: Never) |  |  |
|  | At least 1/week | 5.84 | 3.49 |
|  | 1-3/month | 3.91 | 2.39 |
|  | < 1/month | 1.67 | 1.07 |
| Year of birth | (Ref: 1998-2005; age 18-24) |  |  |
|  | 1993-1998; age 25-29 | 1.16 | 1.00 |
|  | 1983-1993; age 30-39 | 1.20 | 1.00 |
|  | 1973-1983; age 40-49 | 1.23 | 1.00 |
|  | 1963-1973; age 50-59 | 1.45 | 1.10 |
|  | 1953-1963; age 60-69 | 1.47 | 1.00 |
|  | 1943-1953; age 70-79 | 1.42 | 1.00 |
|  | 1943 or earlier; age 80+□ | 1.74 | 1.13 |
| Gender | (Ref: Male) |  |  |
|  | Female | 1.17 | 1.00 |
|  | Other ☐ | 11.46 | 1.00 |

Note. N = 202,898; the E-value is the minimum strength of the association an unmeasured confounder must have with both the outcome and the predictor, above and beyond all measured covariates, for an unmeasured confounder to explain away an association (VanderWeele & Ding, 2017, p. 269-270); and ☐ indicates that the group is very small (<0.1% of the observed sample) within several countries potentially leading to complete separation and large uncertainty in this estimate and caution is warranted in interpreting this estimate.

Results from the population weighted meta-analysis (Supplementary Tables S23 and S24) were generally similar to the random effects meta-analysis, but there were a few exceptions. For adult secular community participation, the associations for some categories of relationship with father, feeling like an outsider in the family, and self-related childhood health met p<.05 in the random effects meta-analysis only, whereas the association for parent marital status (never married/one of both parents died vs. married) reached p<.05 in the population weighted meta-analysis only. For adult service attendance, the association for some categories of parental marital status (one or both parents had died vs. parents married), self-rated childhood health (excellent vs. good), and year of birth (1943-1963 and 1973-1998) reached p<.05 in the population weighted meta-analysis only. These differences may be attributed to the large weighting given to India in the population weighted meta-analysis.

## DISCUSSION

Participation in community groups, whether within secular or religious contexts, may cultivate a sense of agency, belonging, and purpose that fosters individual and community growth^2–4^. As the world becomes increasingly fragmented and the pandemic of loneliness spreads further, the needs to strengthen social connection and community life become ever more urgent^22^. However, the antecedents of community participation, especially childhood factors that shape adult community participation, remain unclear. Such knowledge is essential for developing effective approaches to enhancing community participation. This study expands the literature by examining various personal, familial, and societal-level childhood experiences as potential antecedents of adult participation in secular and religious communities across different national contexts, using a large sample of adults from 22 diverse countries, with the sample weighted to be nationally representative within each country.

In this sample, frequent religious service attendance in childhood is the strongest childhood antecedent of adult participation in both secular and religious communities in a majority of the 22 countries. On average across the 22 countries, people who attended religious services weekly^+^ in childhood were 1.54 times more likely to participate in secular communities weekly^+^ in adulthood. In some countries, this association was particularly strong. For example, in Hong Kong, participants who attended religious services weekly^+^ (vs. never) in childhood were 5.57 times more likely to participate in secular communities weekly^+^ as adults. Similarly, on average across countries those who attended services weekly^+^ in childhood were 3.19 times more likely to continue doing so during adulthood; in Sweden, this likelihood was as high as 34.34 times. These findings are congruent with prior studies^37,67^ (mainly based on data from WEIRD countries) but now extend such evidence to a wider range of countries. These findings may be understood with Putnam’s Theory on Social Capital^1^, highlighting the potential profound impact of religious community participation. For young people, participating in religious communities serves as a gateway to bridging social networks (e.g., meeting diverse individuals through intergenerational activities, forming meaningful connections with peers and mentors), nurturing civic skills (e.g., organizing community events, volunteering in leadership roles within the congregation, participating in interfaith initiatives/dialogues), cultivating altruistic values (e.g., learning about compassion and service through religious teachings and experiential activities such as fundraising and directly helping various groups in need), and engaging in broader community life that leads to collective good (e.g., joining voluntary associations such as food banks and the Red Cross); through these experiences, young people develop a deeper sense of social trust and mutual responsibility, creating social bonds that last a lifetime^1,68^. These connections, often rooted in trust and reciprocity, in turn may lay the groundwork for continued community participation as young people grow into adults. As Putnam argued in his book *Bowling Alone*, faith communities – where people come together to practice faith, celebrate rituals, and strengths their shared beliefs - are perhaps the most important reservoir of social capital, at least in the United States^22^. Interestingly, in this sample, the associations between religious service attendance in early life and continued attendance in adulthood are strongest in several of the most secular societies in the world (e.g., Sweden, Germany, Japan, Spain). We speculate that families that practice religious upbringing in secular countries may be those with a particularly strong faith and faith-related networks, thus their children may be more likely to sustain religious community involvement into adulthood.

This study also found that living comfortably financially (vs. got by) in childhood was associated with greater adult participation in secular communities on average across countries. This finding aligns with prior literature suggesting that people with higher SES tend to have greater civic, political, and voluntary participation^27,33,34^. Prior researchers hypothesized that people from higher SES backgrounds have more resources (e.g., financial resources, memberships, time), greater access to opportunities (e.g., are more aware of current affairs and have broader social connections to access community activities), and are more likely to have high educational attainment (education teaches civic knowledge and skills, and helps foster civic character), all contributing to greater capacities to engage in civic life^35,69^. Beyond civic and voluntary associations, many other secular communities (e.g., sports teams, leisure clubs) require memberships and access fees, thus may be likewise more affordable to those from higher SES backgrounds. Conversely, on average, childhood financial status was not associated with adult religious service attendance across countries. This may be because members of religious communities are brought together not by their social status, but by their shared faith and a common desire to connect with something greater than themselves. Many religious teachings value love and respect for all people in their communities, regardless of their backgrounds^72^.

People with optimal childhood health (excellent vs. good) are also more likely to engage in secular communities weekly^+^ in adulthood on average across countries, possibly because they are less likely to experience health barriers to engaging in communities. It is worth noting, however, that the association for childhood health appeared curvilinear, whereby those who rated their childhood health as “poor” (vs. “good”) were also more likely to participate in secular communities in adulthood on average across countries. We speculate that people who experience health challenges and other adversities early in life may develop heightened empathy and strong motivations to give back to the community when they grow into adults^70^, through serving in communities that aim to promote social justice and equity. They may also seek out community involvement as an approach to building supportive social networks and overcome social disconnection that they may have experienced in childhood due to health limitations^71^.

Somewhat unexpectedly, on average across countries, people who experienced child abuse or felt like an outsider in family when growing up are both more likely to participate in secular communities in adulthood. This is contrary to some prior evidence suggesting that child abuse and neglect may result in low self-esteem, anxiety, and mistrust of others that in turn triggers “a fight-or-flight response”, leading to difficulties in community life that often requires trust and collaboration^26,31,32^. Conversely, in our sample neither child abuse nor feeling like an outsider in family showed strong associations with a lower likelihood of adult weekly^+^ secular community participation in any of the 22 individual countries. This may be partly because our broad measure of secular community participation can cover various recovery programs, support groups, and social service organizations, where victims of childhood trauma may turn to for healing. Supportive communities may also provide their members with a sense of identity, connection and belonging^1^, which could be particularly meaningful for people who have not had a chance to develop nurturing interpersonal relationships in earlier life. In addition, there is also prior evidence suggesting that childhood adversities may sometimes lead to heightened empathy towards the well-being of others, which one might refer to as “altruism-born-of-suffering”^73^. To this end, victims of childhood adversities may engage in civic and voluntary activities as a way to give back to society and offer support for others in need, while attaining a sense of worth, meaning, and growth in the process. It is also worth noting that in our sample the countries with the strongest associations between child abuse and adult secular community participation are mostly those that are ranked middle or low in national commitment and infrastructure for addressing violence towards children (e.g., Kenya, India, Japan, Nigeria)^74^, highlighting the needs to enhance societal engagement and address structural barriers in the prevention of, and recovery from, child maltreatment. In comparison, in most of the countries that we surveyed, neither child abuse nor feeling like an outsider in the family was strongly associated with adult religious service attendance. On one hand, victims of childhood trauma may turn to religion for spiritual guidance in finding meaning and attaining growth from their suffering^75^. On the other hand, children who are mistreated, particularly within religious institutions, may question the existence of a benevolent higher power, rebel against the authoritarianism in religious teachings, and turn away from faith communities to seek healing elsewhere^76^. It is possible that these opposing dynamics balance each other out, leading to a null association overall.

This study has some limitations. First, we were not able to investigate specific types of secular communities (e.g., political groups, civic associations, sports clubs, leisure groups) or religious groups (prayer or meditation groups, bible study groups, spiritual healing groups, groups specific to certain religious denominations) due to lack of data. It is possible that childhood antecedents may differ across participation in these specific types of communities. In addition, as the use of digital media increases sharply especially among younger cohorts, the antecedents of participation in emerging forms of communities such as online communities will need to be understood as well. Second, because the participants recall their childhood experiences and reported their current community participation simultaneously, there may be recall bias and we cannot infer causality from the data. Third, there may be collinearity between some of the childhood factors (e.g., relationship with mother, relationship with father, child abuse, felt like an outsider in the family) in the regression models. To reduce concerns about collinearity, we collapsed some response categories and removed some variables from the regression models (see details in our pre-registration), but we cannot entirely rule out the possibility of collinearity. Fourth, there are other important childhood factors at the school (e.g., school types, peer relationships), neighborhood (e.g., neighborhood composition), and societal level (e.g., welfare systems, country-level religiosity) that may shape adults’ community participation within different national contexts^79–82^, which need to be better understood in future research.

Participating in group-based communities, whether secular or faith-based, contributes to both individual and communal flourishing, through enhancing personal well-being, while also strengthening social capital and the proper functioning of civic institutions^3,4,22^. Findings from this study align with the expanding body of research indicating that an orientation towards community engagement and collective good may be cultivated starting from early life^26^, highlighting a range of personal, familial, and societal factors that may matter within different national contexts. The findings also indicate that interventions that consider addressing societal and structural conditions may more effectively enhance social participation and foster equitable access to community life. Further research on antecedents of community participation across societies will deepen our understanding of the approaches that empower people to create inclusive, supportive, and resilient communities where individuals flourish and contribute to the greater good.

## Acknowledgements

The Global Flourishing Study was supported by funding from the John Templeton Foundation (grant #61665), Templeton Religion Trust (#1308), Templeton World Charity Foundation (#0605), Well-Being for Planet Earth Foundation, Fetzer Institute (#4354), Well Being Trust, Paul L. Foster Family Foundation, and the David and Carol Myers Foundation. The funding source had no impact on the study design; on the collection, analysis and interpretation of data; on the writing of the report; or on the decision to submit the article for publication.

## Author Contributions

B.R. J., and T.J.V. developed the study concept. Y.C., B.W.C, K.N.G.L., R.W., E.S.K., R.N.P., B.R. J., and T.J.V. contributed to the study design. Y.C. and R.N.P. had full access to the data, conducted data analyses, and take responsibility for the integrity of the data and accuracy of the data analysis. Y.C. drafted the manuscript. B.W.C, K.N.G.L., R.W., E.S.K., R.N.P., B.R. J., and T.J.V. provided critical revisions, and approved the final submitted version of the manuscript.

## Ethics approval and consent to participate

Ethical approval was granted by the institutional review boards at Baylor University (IRB Reference #: 6431841317) and Gallup (IRB Reference #: 2021-11-02), and all participants provided informed consent.

## Data Availability

Data for Wave 1 of the Global Flourishing Study is available through the Center for Open Science upon submission of a pre-registration (https://doi.org/10.17605/OSF.IO/3JTZ8), and will be openly available without pre-registration beginning February 2025. Please see https://www.cos.io/gfs-access-data for more information about data access.

## Code Availability

This study was pre-registered with Center for Open Science prior to data access (https://doi.org/10.17605/OSF.IO/VM7W9 and https://doi.org/10.17605/OSF.IO/6W392). All code to reproduce analyses are openly available in the online OSF repository (https://doi.org/10.17605/osf.io/vbype).

## Completing Interests

Tyler VanderWeele reports consulting fees from Gloo Inc., along with shared revenue received by Harvard University in its license agreement with Gloo according to the University IP policy. Other authors have no conflicts of interest to declare.

