## Supplementary file for "Early Life Experiences and Adult Community Participation in Secular and Religious Contexts in 22 Countries"

- Table S1a – S22c: Supplementary tables for country-specific analyses.
- Table S23: Supplementary table for population weighted meta-analysis.
- Table S24: Supplementary table for population weighted meta-analysis of E-values.
- Figure S1– S27: Supplementary figures for estimates of childhood predictor categories.

Notes: This online supplement has several important caveats to interpretation. First, this study focused on childhood predictors using a retrospective (recall) approach to obtain information about the respondents’ lives at age 12. Several childhood characteristics such as relationship quality with parents, subjective financial status growing up, etc. are necessarily highly correlated. This led to multicollinearity issues, so some effects may be unstable. Secondly, the low degree of variability of the outcome variable in some countries for some subgroups leads to the issues of complete separation in the modified Poisson regression model. The complete separation results in uninterpretable estimates for those effects, as there were no observed differences between the exposure and the outcome in this sample. We have tried to note when this occurs in the meta-analyses. Third, in rare instances the confidence interval of the effect estimate can contradict the reported global p-value (e.g., for the single-category effects of relationship with mother). In such cases, the reported confidence interval is more robust with corrected degrees of freedom from the pooling across multiple imputations, whereas the global p-value is based on a Wald-type test and is less robust to uncertainty attributable to multiple imputation. Lastly, comparing results across countries should be done with caution due to possible measurement non-invariance and differences in translation.

Table S1a. Nationally representative descriptive statistics for Argentina

| **Characteristic** | **N = 6,724**^1^ |
| --- | --- |
| **Relationship with mother** |  |
| Very good | 4,463 (66%) |
| Somewhat good | 1,436 (21%) |
| Somewhat bad | 299 (4.4%) |
| Very bad | 216 (3.2%) |
| Does not apply | 273 (4.1%) |
| (Missing) | 36 (0.5%) |
| **Relationship with father** |  |
| Very good | 3,612 (54%) |
| Somewhat good | 1,537 (23%) |
| Somewhat bad | 440 (6.5%) |
| Very bad | 401 (6.0%) |
| Does not apply | 694 (10%) |
| (Missing) | 39 (0.6%) |
| **Parent marital status** |  |
| Parents married | 4,110 (61%) |
| Divorced | 637 (9.5%) |
| Parents were never married | 1,368 (20%) |
| One or both parents had died | 199 (3.0%) |
| (Missing) | 410 (6.1%) |
| **Subjective financial status of family growing up** |  |
| Lived comfortably | 2,042 (30%) |
| Got by | 2,305 (34%) |
| Found it difficult | 1,789 (27%) |
| Found it very difficult | 569 (8.5%) |
| (Missing) | 19 (0.3%) |
| **Experienced Abuse** |  |
| Yes | 1,302 (19%) |
| No | 5,271 (78%) |
| (Missing) | 151 (2.2%) |
| **Felt like an outsider in the family** |  |
| Yes | 1,165 (17%) |
| No | 5,458 (81%) |
| (Missing) | 101 (1.5%) |
| **Self-rated health growing up** |  |
| Excellent | 2,402 (36%) |
| Very good | 1,819 (27%) |
| Good | 1,830 (27%) |
| Fair | 505 (7.5%) |
| Poor | 156 (2.3%) |
| (Missing) | 12 (0.2%) |
| **Immigration status** |  |
| Born in this country | 6,346 (94%) |
| Born in another country | 348 (5.2%) |
| (Missing) | 29 (0.4%) |
| **Religious service attendance (age 12)** |  |
| At least 1/week | 2,601 (39%) |
| 1-3/month | 1,204 (18%) |
| <1/month | 1,059 (16%) |
| Never | 1,808 (27%) |
| (Missing) | 53 (0.8%) |
| **Year of birth** |  |
| 1998-2005; age 18-24 | 1,108 (16%) |
| 1993-1998; age 25-29 | 719 (11%) |
| 1983-1993; age 30-39 | 1,432 (21%) |
| 1973-1983; age 40-49 | 1,254 (19%) |
| 1963-1973; age 50-59 | 1,014 (15%) |
| 1953-1963; age 60-69 | 730 (11%) |
| 1943-1953; age 70-79 | 356 (5.3%) |
| 1943 or earlier; age 80+ | 112 (1.7%) |
| (Missing) | 0 (0%) |
| **Gender** |  |
| Male | 3,143 (47%) |
| Female | 3,542 (53%) |
| Other | 21 (0.3%) |
| (Missing) | 18 (0.3%) |
| **Religious affiliation** |  |
| Christianity | 5,805 (86%) |
| Islam | 11 (0.2%) |
| Hinduism | 2 (<0.1%) |
| Buddhism | 3 (<0.1%) |
| Judaism | 51 (0.8%) |
| Sikhism | 5 (<0.1%) |
| Baha'i | 0 (0%) |
| Jainism | 0 (0%) |
| Shinto | 0 (0%) |
| Taoism | 1 (<0.1%) |
| Confucianism | 0 (0%) |
| Primal, Animist, or Folk religion | 17 (0.2%) |
| Spiritism | 0 (0%) |
| Umbanda, Candomble, and other African-derived religions | 0 (0%) |
| Chinese folk/traditional religion | 0 (0%) |
| Some other religion | 10 (0.2%) |
| No religion/Atheist/Agnostic | 697 (10%) |
| (Missing) | 122 (1.8%) |
| **Race/Ethnicity** |  |
| Asian | 43 (0.6%) |
| Black | 95 (1.4%) |
| Indigenous | 129 (1.9%) |
| Mestizo(a) | 1,801 (27%) |
| Mullato(a) | 75 (1.1%) |
| Other | 104 (1.5%) |
| White | 3,406 (51%) |
| (Missing) | 1,070 (16%) |
| ^1^n (%) | |

Table S1b. Regression of Community Participation on Childhood Predictors for Argentina

|  | | Secular Community Participation | | | |  | Religious Service Attendance | | | |
| --- | --- | --- | --- | --- | --- | --- | --- | --- | --- | --- |
| Variable | Category | Risk-Ratio | RR 95% CI | log(RR) SE | Global  p-value |  | Risk-Ratio | RR 95% CI | log(RR) SE | Global  p-value |
| Relationship with mother | (Ref: Very bad/somewhat bad) |  |  |  | 0.034 |  |  |  |  | 0.485 |
|  | Very good/somewhat good | 1.34 | (1.01,1.78) | 0.14 |  |  | 0.91 | (0.69,1.21) | 0.14 |  |
| Relationship with father | (Ref: Very bad/somewhat bad) |  |  |  | 0.177 |  |  |  |  | 0.130 |
|  | Very good/somewhat good | 1.16 | (0.93,1.44) | 0.11 |  |  | 1.20 | (0.95,1.52) | 0.12 |  |
| Parent marital status | (Ref: Parents married) |  |  |  | 0.068 |  |  |  |  | 0.793 |
|  | Divorced | 1.11 | (0.88,1.40) | 0.12 |  |  | 0.96 | (0.73,1.26) | 0.14 |  |
|  | Parents were never married | 0.95 | (0.79,1.16) | 0.10 |  |  | 1.00 | (0.82,1.23) | 0.11 |  |
|  | One or both parents had died | 1.44 | (1.01,2.07) | 0.18 |  |  | 0.81 | (0.48,1.36) | 0.27 |  |
| Subjective financial status of family growing up | (Ref: Got by) |  |  |  | 0.041 |  |  |  |  | 0.983 |
|  | Lived comfortably | 1.12 | (0.96,1.31) | 0.08 |  |  | 1.02 | (0.86,1.22) | 0.09 |  |
|  | Found it difficult | 0.86 | (0.71,1.03) | 0.09 |  |  | 1.02 | (0.85,1.24) | 0.10 |  |
|  | Found it very difficult | 0.98 | (0.73,1.32) | 0.15 |  |  | 1.05 | (0.80,1.38) | 0.14 |  |
| Experienced Abuse | (Ref: No) |  |  |  | 0.731 |  |  |  |  | 0.733 |
|  | Yes | 1.02 | (0.85,1.23) | 0.09 |  |  | 1.03 | (0.86,1.23) | 0.09 |  |
| Felt like an outsider in the family | (Ref: No) |  |  |  | 0.017 |  |  |  |  | 0.395 |
|  | Yes | 1.25 | (1.04,1.50) | 0.09 |  |  | 1.10 | (0.88,1.36) | 0.11 |  |
| Self-rated health growing up | (Ref: Good) |  |  |  | 0.009 |  |  |  |  | 0.687 |
|  | Excellent | 1.41 | (1.17,1.71) | 0.10 |  |  | 0.94 | (0.78,1.13) | 0.09 |  |
|  | Very good | 1.35 | (1.10,1.64) | 0.10 |  |  | 0.89 | (0.74,1.08) | 0.10 |  |
|  | Fair | 1.21 | (0.91,1.60) | 0.14 |  |  | 1.02 | (0.77,1.36) | 0.14 |  |
|  | Poor | 1.26 | (0.72,2.22) | 0.29 |  |  | 1.13 | (0.72,1.79) | 0.23 |  |
| Immigration status | (Ref: Born in this country) |  |  |  | 0.275 |  |  |  |  | 0.883 |
|  | Born in another country | 0.83 | (0.59,1.16) | 0.17 |  |  | 0.99 | (0.72,1.35) | 0.16 |  |
| Religious service attendance (age 12) | (Ref: Never) |  |  |  | <.001 |  |  |  |  | <.001 |
|  | At least 1/week | 1.89 | (1.56,2.28) | 0.10 |  |  | 2.36 | (1.89,2.96) | 0.11 |  |
|  | 1-3/month | 1.62 | (1.29,2.03) | 0.12 |  |  | 1.44 | (1.10,1.89) | 0.14 |  |
|  | < 1/month | 1.13 | (0.88,1.45) | 0.13 |  |  | 0.97 | (0.71,1.32) | 0.16 |  |
| Year of birth | (Ref: 1998-2005; current age: 18-24) |  |  |  | 0.002 |  |  |  |  | 0.257 |
|  | 1993-1998; age 25-29 | 0.91 | (0.71,1.16) | 0.12 |  |  | 1.38 | (1.04,1.82) | 0.14 |  |
|  | 1983-1993; age 30-39 | 0.80 | (0.65,0.98) | 0.10 |  |  | 1.15 | (0.88,1.48) | 0.13 |  |
|  | 1973-1983; age 40-49 | 0.78 | (0.63,0.96) | 0.11 |  |  | 1.10 | (0.85,1.43) | 0.13 |  |
|  | 1963-1973; age 50-59 | 0.84 | (0.67,1.06) | 0.12 |  |  | 1.30 | (0.98,1.72) | 0.14 |  |
|  | 1953-1963; age 60-69 | 0.71 | (0.53,0.94) | 0.15 |  |  | 1.02 | (0.75,1.40) | 0.16 |  |
|  | 1943-1953; age 70-79 | 0.46 | (0.30,0.69) | 0.21 |  |  | 1.30 | (0.89,1.91) | 0.20 |  |
|  | 1943 or earlier; age 80+ | 0.37 | (0.15,0.91) | 0.46 |  |  | 1.14 | (0.65,2.01) | 0.29 |  |
| Gender | (Ref: Male) |  |  |  | <.001 |  |  |  |  | 0.028 |
|  | Female | 0.77 | (0.67,0.88) | 0.07 |  |  | 1.16 | (1.00,1.35) | 0.08 |  |
|  | Other | 1.57 | (0.60,4.08) | 0.49 |  |  | 0.19 | (0.03,1.33) | 1.00 |  |
| Religious affiliation | (Ref: No religion/Atheist/Agnostic) |  |  |  | 0.064 |  |  |  |  | 0.023 |
|  | Christianity | 0.79 | (0.63,1.00) | 0.12 |  |  | 1.60 | (1.11,2.32) | 0.19 |  |
|  | Collapsed affiliations with prevalence<3% | 1.09 | (0.66,1.80) | 0.26 |  |  | 1.93 | (1.03,3.60) | 0.32 |  |
| Race/ethnicity | (Ref: Plurality group) |  |  |  | 0.337 |  |  |  |  | 0.220 |
|  | Non-plurality groups | 1.06 | (0.92,1.23) | 0.07 |  |  | 1.09 | (0.93,1.27) | 0.08 |  |

Table S1c. Sensitivity of the regression estimates to unmeasured confounding for Argentina

|  | | Secular Community Participation | |  | Religious Service Attendance | |
| --- | --- | --- | --- | --- | --- | --- |
| Variable | Category | E-value for Estimate | E-value for 95% CI |  | E-value for Estimate | E-value for 95% CI |
| Relationship with mother | (Ref: Very bad/somewhat bad) |  |  |  |  |  |
|  | Very good/somewhat good | 2.02 | 1.13 |  | 1.42 | 1.00 |
| Relationship with father | (Ref: Very bad/somewhat bad) |  |  |  |  |  |
|  | Very good/somewhat good | 1.59 | 1.00 |  | 1.69 | 1.00 |
| Parent marital status | (Ref: Parents married) |  |  |  |  |  |
|  | Divorced | 1.47 | 1.00 |  | 1.26 | 1.00 |
|  | Parents were never married | 1.27 | 1.00 |  | 1.05 | 1.00 |
|  | One or both parents had died | 2.25 | 1.09 |  | 1.78 | 1.00 |
| Subjective financial status of family growing up | (Ref: Got by) |  |  |  |  |  |
|  | Lived comfortably | 1.49 | 1.00 |  | 1.18 | 1.00 |
|  | Found it difficult | 1.60 | 1.00 |  | 1.18 | 1.00 |
|  | Found it very difficult | 1.17 | 1.00 |  | 1.28 | 1.00 |
| Experienced Abuse | (Ref: No) |  |  |  |  |  |
|  | Yes | 1.18 | 1.00 |  | 1.19 | 1.00 |
| Felt like an outsider in the family | (Ref: No) |  |  |  |  |  |
|  | Yes | 1.80 | 1.24 |  | 1.42 | 1.00 |
| Self-rated health growing up | (Ref: Good) |  |  |  |  |  |
|  | Excellent | 2.18 | 1.62 |  | 1.33 | 1.00 |
|  | Very good | 2.03 | 1.44 |  | 1.49 | 1.00 |
|  | Fair | 1.72 | 1.00 |  | 1.17 | 1.00 |
|  | Poor | 1.84 | 1.00 |  | 1.52 | 1.00 |
| Immigration status | (Ref: Born in this country) |  |  |  |  |  |
|  | Born in another country | 1.71 | 1.00 |  | 1.13 | 1.00 |
| Religious service attendance (age 12) | (Ref: Never) |  |  |  |  |  |
|  | At least 1/week | 3.19 | 2.50 |  | 4.15 | 3.18 |
|  | 1-3/month | 2.62 | 1.89 |  | 2.24 | 1.43 |
|  | < 1/month | 1.50 | 1.00 |  | 1.20 | 1.00 |
| Year of birth | (Ref: 1998-2005; current age: 18-24) |  |  |  |  |  |
|  | 1993-1998; age 25-29 | 1.43 | 1.00 |  | 2.10 | 1.24 |
|  | 1983-1993; age 30-39 | 1.81 | 1.16 |  | 1.55 | 1.00 |
|  | 1973-1983; age 40-49 | 1.90 | 1.24 |  | 1.43 | 1.00 |
|  | 1963-1973; age 50-59 | 1.65 | 1.00 |  | 1.92 | 1.00 |
|  | 1953-1963; age 60-69 | 2.19 | 1.33 |  | 1.17 | 1.00 |
|  | 1943-1953; age 70-79 | 3.80 | 2.28 |  | 1.93 | 1.00 |
|  | 1943 or earlier; age 80+ | 4.88 | 1.44 |  | 1.55 | 1.00 |
| Gender | (Ref: Male) |  |  |  |  |  |
|  | Female | 1.92 | 1.53 |  | 1.60 | 1.05 |
|  | Other | 2.51 | 1.00 |  | 10.17 | 1.00 |
| Religious affiliation | (Ref: No religion/Atheist/Agnostic) |  |  |  |  |  |
|  | Christianity | 1.83 | 1.07 |  | 2.58 | 1.45 |
|  | Collapsed affiliations with prevalence<3% | 1.39 | 1.00 |  | 3.27 | 1.22 |
| Race/ethnicity | (Ref: Plurality group) |  |  |  |  |  |
|  | Non-plurality groups | 1.33 | 1.00 |  | 1.40 | 1.00 |

Table S2a. Nationally representative descriptive statistics for Australia

| **Characteristic** | **N = 3,844**^1^ |
| --- | --- |
| **Relationship with mother** |  |
| Very good | 2,554 (66%) |
| Somewhat good | 925 (24%) |
| Somewhat bad | 218 (5.7%) |
| Very bad | 107 (2.8%) |
| Does not apply | 32 (0.8%) |
| (Missing) | 7 (0.2%) |
| **Relationship with father** |  |
| Very good | 2,032 (53%) |
| Somewhat good | 1,144 (30%) |
| Somewhat bad | 315 (8.2%) |
| Very bad | 196 (5.1%) |
| Does not apply | 148 (3.9%) |
| (Missing) | 9 (0.2%) |
| **Parent marital status** |  |
| Parents married | 3,048 (79%) |
| Divorced | 462 (12%) |
| Parents were never married | 187 (4.9%) |
| One or both parents had died | 96 (2.5%) |
| (Missing) | 52 (1.4%) |
| **Subjective financial status of family growing up** |  |
| Lived comfortably | 1,756 (46%) |
| Got by | 1,496 (39%) |
| Found it difficult | 422 (11%) |
| Found it very difficult | 154 (4.0%) |
| (Missing) | 16 (0.4%) |
| **Experienced Abuse** |  |
| Yes | 995 (26%) |
| No | 2,790 (73%) |
| (Missing) | 59 (1.5%) |
| **Felt like an outsider in the family** |  |
| Yes | 756 (20%) |
| No | 3,062 (80%) |
| (Missing) | 26 (0.7%) |
| **Self-rated health growing up** |  |
| Excellent | 1,736 (45%) |
| Very good | 1,087 (28%) |
| Good | 603 (16%) |
| Fair | 308 (8.0%) |
| Poor | 106 (2.8%) |
| (Missing) | 4 (<0.1%) |
| **Immigration status** |  |
| Born in this country | 2,953 (77%) |
| Born in another country | 885 (23%) |
| (Missing) | 6 (0.2%) |
| **Religious service attendance (age 12)** |  |
| At least 1/week | 1,362 (35%) |
| 1-3/month | 486 (13%) |
| <1/month | 600 (16%) |
| Never | 1,307 (34%) |
| (Missing) | 90 (2.3%) |
| **Year of birth** |  |
| 1998-2005; age 18-24 | 345 (9.0%) |
| 1993-1998; age 25-29 | 282 (7.3%) |
| 1983-1993; age 30-39 | 641 (17%) |
| 1973-1983; age 40-49 | 618 (16%) |
| 1963-1973; age 50-59 | 691 (18%) |
| 1953-1963; age 60-69 | 589 (15%) |
| 1943-1953; age 70-79 | 498 (13%) |
| 1943 or earlier; age 80+ | 178 (4.6%) |
| (Missing) | 2 (<0.1%) |
| **Gender** |  |
| Male | 1,861 (48%) |
| Female | 1,941 (50%) |
| Other | 36 (0.9%) |
| (Missing) | 6 (0.2%) |
| **Religious affiliation** |  |
| Christianity | 2,678 (70%) |
| Islam | 48 (1.2%) |
| Hinduism | 39 (1.0%) |
| Buddhism | 16 (0.4%) |
| Judaism | 29 (0.8%) |
| Sikhism | 6 (0.2%) |
| Baha'i | 5 (0.1%) |
| Jainism | 0 (0%) |
| Shinto | 0 (0%) |
| Taoism | 1 (<0.1%) |
| Confucianism | 0 (0%) |
| Primal, Animist, or Folk religion | 4 (<0.1%) |
| Spiritism | 0 (0%) |
| Umbanda, Candomble, and other African-derived religions | 0 (0%) |
| Chinese folk/traditional religion | 0 (0%) |
| Some other religion | 8 (0.2%) |
| No religion/Atheist/Agnostic | 990 (26%) |
| (Missing) | 21 (0.5%) |
| **Race/Ethnicity** |  |
| Aboriginal | 53 (1.4%) |
| Australian | 1,946 (51%) |
| Australian British/European | 1,047 (27%) |
| Chinese | 75 (1.9%) |
| Indian | 58 (1.5%) |
| Japanese | 1 (<0.1%) |
| Malay | 11 (0.3%) |
| New Zealander | 91 (2.4%) |
| Other | 163 (4.2%) |
| Other European | 357 (9.3%) |
| Russian | 7 (0.2%) |
| Samoan | 4 (0.1%) |
| Sinhalese | 1 (<0.1%) |
| Spanish | 2 (<0.1%) |
| Sri Lankan Moor | 1 (<0.1%) |
| Sri Lankan Tamil | 7 (0.2%) |
| Vietnamese | 7 (0.2%) |
| (Missing) | 14 (0.4%) |
| ^1^n (%) | |

Table S2b. Regression of Community Participation on Childhood Predictors for Australia

|  | | Secular Community Participation | | | |  | Religious Service Attendance | | | |
| --- | --- | --- | --- | --- | --- | --- | --- | --- | --- | --- |
| Variable | Category | Risk-Ratio | RR 95% CI | log(RR) SE | Global  p-value |  | Risk-Ratio | RR 95% CI | log(RR) SE | Global  p-value |
| Relationship with mother | (Ref: Very bad/somewhat bad) |  |  |  | 0.100 |  |  |  |  | 0.215 |
|  | Very good/somewhat good | 1.28 | (0.95,1.72) | 0.15 |  |  | 1.36 | (0.83,2.24) | 0.25 |  |
| Relationship with father | (Ref: Very bad/somewhat bad) |  |  |  | 0.636 |  |  |  |  | 0.921 |
|  | Very good/somewhat good | 0.95 | (0.77,1.18) | 0.11 |  |  | 1.02 | (0.72,1.44) | 0.18 |  |
| Parent marital status | (Ref: Parents married) |  |  |  | 0.940 |  |  |  |  | 0.006 |
|  | Divorced | 1.06 | (0.84,1.34) | 0.12 |  |  | 0.36 | (0.19,0.66) | 0.31 |  |
|  | Parents were never married | 0.98 | (0.63,1.51) | 0.22 |  |  | 0.74 | (0.33,1.63) | 0.40 |  |
|  | One or both parents had died | 0.95 | (0.60,1.48) | 0.23 |  |  | 0.75 | (0.42,1.34) | 0.30 |  |
| Subjective financial status of family growing up | (Ref: Got by) |  |  |  | 0.297 |  |  |  |  | 0.373 |
|  | Lived comfortably | 0.88 | (0.77,1.01) | 0.07 |  |  | 0.85 | (0.69,1.04) | 0.11 |  |
|  | Found it difficult | 0.98 | (0.78,1.22) | 0.11 |  |  | 0.80 | (0.55,1.15) | 0.19 |  |
|  | Found it very difficult | 1.01 | (0.68,1.50) | 0.20 |  |  | 0.93 | (0.50,1.75) | 0.32 |  |
| Experienced Abuse | (Ref: No) |  |  |  | 0.075 |  |  |  |  | 0.491 |
|  | Yes | 0.86 | (0.73,1.02) | 0.08 |  |  | 1.08 | (0.85,1.37) | 0.12 |  |
| Felt like an outsider in the family | (Ref: No) |  |  |  | 0.107 |  |  |  |  | 0.431 |
|  | Yes | 0.85 | (0.69,1.04) | 0.11 |  |  | 0.88 | (0.64,1.21) | 0.16 |  |
| Self-rated health growing up | (Ref: Good) |  |  |  | 0.012 |  |  |  |  | 0.631 |
|  | Excellent | 1.37 | (1.11,1.68) | 0.11 |  |  | 1.02 | (0.76,1.37) | 0.15 |  |
|  | Very good | 1.28 | (1.03,1.59) | 0.11 |  |  | 1.07 | (0.78,1.46) | 0.16 |  |
|  | Fair | 0.92 | (0.64,1.33) | 0.19 |  |  | 1.40 | (0.88,2.22) | 0.24 |  |
|  | Poor | 1.42 | (0.90,2.23) | 0.23 |  |  | 1.03 | (0.48,2.18) | 0.38 |  |
| Immigration status | (Ref: Born in this country) |  |  |  | 0.450 |  |  |  |  | 0.371 |
|  | Born in another country | 0.94 | (0.79,1.11) | 0.09 |  |  | 1.12 | (0.87,1.45) | 0.13 |  |
| Religious service attendance (age 12) | (Ref: Never) |  |  |  | 0.002 |  |  |  |  | <.001 |
|  | At least 1/week | 1.33 | (1.11,1.59) | 0.09 |  |  | 4.44 | (2.98,6.61) | 0.20 |  |
|  | 1-3/month | 0.98 | (0.77,1.25) | 0.12 |  |  | 1.91 | (1.15,3.18) | 0.26 |  |
|  | < 1/month | 1.10 | (0.88,1.36) | 0.11 |  |  | 1.47 | (0.87,2.49) | 0.27 |  |
| Year of birth | (Ref: 1998-2005; current age: 18-24) |  |  |  | <.001 |  |  |  |  | <.001 |
|  | 1993-1998; age 25-29 | 0.78 | (0.54,1.14) | 0.19 |  |  | 1.51 | (0.77,2.98) | 0.35 |  |
|  | 1983-1993; age 30-39 | 0.72 | (0.54,0.97) | 0.15 |  |  | 1.07 | (0.60,1.92) | 0.30 |  |
|  | 1973-1983; age 40-49 | 0.69 | (0.52,0.92) | 0.15 |  |  | 1.26 | (0.72,2.21) | 0.29 |  |
|  | 1963-1973; age 50-59 | 0.55 | (0.41,0.73) | 0.15 |  |  | 0.92 | (0.52,1.62) | 0.29 |  |
|  | 1953-1963; age 60-69 | 0.64 | (0.48,0.84) | 0.14 |  |  | 1.05 | (0.60,1.84) | 0.29 |  |
|  | 1943-1953; age 70-79 | 1.03 | (0.78,1.36) | 0.14 |  |  | 1.20 | (0.68,2.11) | 0.29 |  |
|  | 1943 or earlier; age 80+ | 1.01 | (0.73,1.39) | 0.16 |  |  | 2.27 | (1.28,4.01) | 0.29 |  |
| Gender | (Ref: Male) |  |  |  | 0.364 |  |  |  |  | 0.122 |
|  | Female | 1.09 | (0.96,1.23) | 0.06 |  |  | 1.23 | (1.01,1.50) | 0.10 |  |
|  | Other | 0.75 | (0.26,2.16) | 0.54 |  |  | 0.99 | (0.13,7.58) | 1.04 |  |
| Religious affiliation | (Ref: No religion/Atheist/Agnostic) |  |  |  | 0.034 |  |  |  |  | <.001 |
|  | Christianity | 1.02 | (0.84,1.25) | 0.10 |  |  | 2.71 | (1.63,4.48) | 0.26 |  |
|  | Collapsed affiliations with prevalence<3% | 0.56 | (0.35,0.90) | 0.24 |  |  | 3.40 | (1.79,6.45) | 0.33 |  |
| Race/ethnicity | (Ref: Plurality group) |  |  |  | 0.860 |  |  |  |  | 0.896 |
|  | Non-plurality groups | 1.01 | (0.88,1.16) | 0.07 |  |  | 1.00 | (0.80,1.25) | 0.12 |  |

Table S2c. Sensitivity of the regression estimates to unmeasured confounding for Australia

|  | | Secular Community Participation | |  | Religious Service Attendance | |
| --- | --- | --- | --- | --- | --- | --- |
| Variable | Category | E-value for Estimate | E-value for 95% CI |  | E-value for Estimate | E-value for 95% CI |
| Relationship with mother | (Ref: Very bad/somewhat bad) |  |  |  |  |  |
|  | Very good/somewhat good | 1.87 | 1.00 |  | 2.07 | 1.00 |
| Relationship with father | (Ref: Very bad/somewhat bad) |  |  |  |  |  |
|  | Very good/somewhat good | 1.28 | 1.00 |  | 1.15 | 1.00 |
| Parent marital status | (Ref: Parents married) |  |  |  |  |  |
|  | Divorced | 1.31 | 1.00 |  | 5.02 | 2.38 |
|  | Parents were never married | 1.17 | 1.00 |  | 2.04 | 1.00 |
|  | One or both parents had died | 1.31 | 1.00 |  | 2.01 | 1.00 |
| Subjective financial status of family growing up | (Ref: Got by) |  |  |  |  |  |
|  | Lived comfortably | 1.53 | 1.00 |  | 1.65 | 1.00 |
|  | Found it difficult | 1.18 | 1.00 |  | 1.82 | 1.00 |
|  | Found it very difficult | 1.10 | 1.00 |  | 1.36 | 1.00 |
| Experienced Abuse | (Ref: No) |  |  |  |  |  |
|  | Yes | 1.59 | 1.00 |  | 1.38 | 1.00 |
| Felt like an outsider in the family | (Ref: No) |  |  |  |  |  |
|  | Yes | 1.65 | 1.00 |  | 1.53 | 1.00 |
| Self-rated health growing up | (Ref: Good) |  |  |  |  |  |
|  | Excellent | 2.07 | 1.46 |  | 1.15 | 1.00 |
|  | Very good | 1.88 | 1.20 |  | 1.34 | 1.00 |
|  | Fair | 1.39 | 1.00 |  | 2.15 | 1.00 |
|  | Poor | 2.19 | 1.00 |  | 1.19 | 1.00 |
| Immigration status | (Ref: Born in this country) |  |  |  |  |  |
|  | Born in another country | 1.33 | 1.00 |  | 1.49 | 1.00 |
| Religious service attendance (age 12) | (Ref: Never) |  |  |  |  |  |
|  | At least 1/week | 1.99 | 1.46 |  | 8.34 | 5.40 |
|  | 1-3/month | 1.15 | 1.00 |  | 3.24 | 1.56 |
|  | < 1/month | 1.42 | 1.00 |  | 2.30 | 1.00 |
| Year of birth | (Ref: 1998-2005; current age: 18-24) |  |  |  |  |  |
|  | 1993-1998; age 25-29 | 1.87 | 1.00 |  | 2.39 | 1.00 |
|  | 1983-1993; age 30-39 | 2.12 | 1.21 |  | 1.34 | 1.00 |
|  | 1973-1983; age 40-49 | 2.25 | 1.38 |  | 1.83 | 1.00 |
|  | 1963-1973; age 50-59 | 3.07 | 2.07 |  | 1.40 | 1.00 |
|  | 1953-1963; age 60-69 | 2.52 | 1.66 |  | 1.28 | 1.00 |
|  | 1943-1953; age 70-79 | 1.21 | 1.00 |  | 1.69 | 1.00 |
|  | 1943 or earlier; age 80+ | 1.10 | 1.00 |  | 3.96 | 1.88 |
| Gender | (Ref: Male) |  |  |  |  |  |
|  | Female | 1.39 | 1.00 |  | 1.77 | 1.10 |
|  | Other | 1.99 | 1.00 |  | 1.08 | 1.00 |
| Religious affiliation | (Ref: No religion/Atheist/Agnostic) |  |  |  |  |  |
|  | Christianity | 1.18 | 1.00 |  | 4.85 | 2.65 |
|  | Collapsed affiliations with prevalence<3% | 2.96 | 1.47 |  | 6.26 | 2.99 |
| Race/ethnicity | (Ref: Plurality group) |  |  |  |  |  |
|  | Non-plurality groups | 1.09 | 1.00 |  | 1.01 | 1.00 |

Table S3a. Nationally representative descriptive statistics for Brazil

| **Characteristic** | **N = 13,204**^1^ |
| --- | --- |
| **Relationship with mother** |  |
| Very good | 8,369 (63%) |
| Somewhat good | 3,559 (27%) |
| Somewhat bad | 483 (3.7%) |
| Very bad | 214 (1.6%) |
| Does not apply | 507 (3.8%) |
| (Missing) | 73 (0.6%) |
| **Relationship with father** |  |
| Very good | 6,364 (48%) |
| Somewhat good | 3,654 (28%) |
| Somewhat bad | 1,035 (7.8%) |
| Very bad | 756 (5.7%) |
| Does not apply | 1,303 (9.9%) |
| (Missing) | 93 (0.7%) |
| **Parent marital status** |  |
| Parents married | 8,546 (65%) |
| Divorced | 1,384 (10%) |
| Parents were never married | 1,985 (15%) |
| One or both parents had died | 508 (3.8%) |
| (Missing) | 781 (5.9%) |
| **Subjective financial status of family growing up** |  |
| Lived comfortably | 4,998 (38%) |
| Got by | 4,616 (35%) |
| Found it difficult | 2,484 (19%) |
| Found it very difficult | 1,027 (7.8%) |
| (Missing) | 79 (0.6%) |
| **Experienced Abuse** |  |
| Yes | 2,606 (20%) |
| No | 10,147 (77%) |
| (Missing) | 451 (3.4%) |
| **Felt like an outsider in the family** |  |
| Yes | 1,659 (13%) |
| No | 11,234 (85%) |
| (Missing) | 311 (2.4%) |
| **Self-rated health growing up** |  |
| Excellent | 5,312 (40%) |
| Very good | 3,392 (26%) |
| Good | 2,873 (22%) |
| Fair | 1,368 (10%) |
| Poor | 228 (1.7%) |
| (Missing) | 30 (0.2%) |
| **Immigration status** |  |
| Born in this country | 12,688 (96%) |
| Born in another country | 153 (1.2%) |
| (Missing) | 363 (2.7%) |
| **Religious service attendance (age 12)** |  |
| At least 1/week | 6,306 (48%) |
| 1-3/month | 2,491 (19%) |
| <1/month | 2,629 (20%) |
| Never | 1,707 (13%) |
| (Missing) | 71 (0.5%) |
| **Year of birth** |  |
| 1998-2005; age 18-24 | 1,986 (15%) |
| 1993-1998; age 25-29 | 1,468 (11%) |
| 1983-1993; age 30-39 | 2,908 (22%) |
| 1973-1983; age 40-49 | 2,638 (20%) |
| 1963-1973; age 50-59 | 2,131 (16%) |
| 1953-1963; age 60-69 | 1,435 (11%) |
| 1943-1953; age 70-79 | 510 (3.9%) |
| 1943 or earlier; age 80+ | 126 (1.0%) |
| (Missing) | 0 (0%) |
| **Gender** |  |
| Male | 6,320 (48%) |
| Female | 6,820 (52%) |
| Other | 35 (0.3%) |
| (Missing) | 30 (0.2%) |
| **Religious affiliation** |  |
| Christianity | 11,403 (86%) |
| Islam | 15 (0.1%) |
| Hinduism | 1 (<0.1%) |
| Buddhism | 27 (0.2%) |
| Judaism | 40 (0.3%) |
| Sikhism | 0 (0%) |
| Baha'i | 1 (<0.1%) |
| Jainism | 4 (<0.1%) |
| Shinto | 4 (<0.1%) |
| Taoism | 1 (<0.1%) |
| Confucianism | 7 (<0.1%) |
| Primal, Animist, or Folk religion | 17 (0.1%) |
| Spiritism | 336 (2.5%) |
| Umbanda, Candomble, and other African-derived religions | 262 (2.0%) |
| Chinese folk/traditional religion | 0 (0%) |
| Some other religion | 87 (0.7%) |
| No religion/Atheist/Agnostic | 908 (6.9%) |
| (Missing) | 94 (0.7%) |
| **Race/Ethnicity** |  |
| Amarela | 238 (1.8%) |
| Branca | 5,169 (39%) |
| Indígena | 131 (1.0%) |
| Other | 61 (0.5%) |
| Parda | 5,125 (39%) |
| Preta | 1,615 (12%) |
| (Missing) | 865 (6.6%) |
| ^1^n (%) | |

Table S3b. Regression of Community Participation on Childhood Predictors for Brazil

|  | | Secular Community Participation | | | |  | Religious Service Attendance | | | |
| --- | --- | --- | --- | --- | --- | --- | --- | --- | --- | --- |
| Variable | Category | Risk-Ratio | RR 95% CI | log(RR) SE | Global  p-value |  | Risk-Ratio | RR 95% CI | log(RR) SE | Global  p-value |
| Relationship with mother | (Ref: Very bad/somewhat bad) |  |  |  | 0.480 |  |  |  |  | 0.312 |
|  | Very good/somewhat good | 1.07 | (0.87,1.32) | 0.11 |  |  | 1.07 | (0.93,1.23) | 0.07 |  |
| Relationship with father | (Ref: Very bad/somewhat bad) |  |  |  | 0.037 |  |  |  |  | 0.023 |
|  | Very good/somewhat good | 1.15 | (1.01,1.32) | 0.07 |  |  | 1.11 | (1.01,1.21) | 0.05 |  |
| Parent marital status | (Ref: Parents married) |  |  |  | 0.973 |  |  |  |  | 0.112 |
|  | Divorced | 0.99 | (0.85,1.14) | 0.07 |  |  | 0.91 | (0.82,1.01) | 0.05 |  |
|  | Parents were never married | 1.00 | (0.86,1.16) | 0.08 |  |  | 0.91 | (0.82,1.00) | 0.05 |  |
|  | One or both parents had died | 1.02 | (0.79,1.32) | 0.13 |  |  |  | (0.78,1.11) | 0.09 |  |
| Subjective financial status of family growing up | (Ref: Got by) |  |  |  | 0.228 |  |  |  |  | 0.283 |
|  | Lived comfortably | 1.07 | (0.97,1.19) | 0.05 |  |  | 1.01 | (0.94,1.08) | 0.04 |  |
|  | Found it difficult | 0.94 | (0.81,1.08) | 0.07 |  |  | 1.06 | (0.97,1.16) | 0.04 |  |
|  | Found it very difficult | 1.01 | (0.81,1.25) | 0.11 |  |  | 1.10 | (0.97,1.23) | 0.06 |  |
| Experienced Abuse | (Ref: No) |  |  |  | 0.144 |  |  |  |  | 0.843 |
|  | Yes | 0.92 | (0.81,1.03) | 0.06 |  |  | 1.00 | (0.92,1.08) | 0.04 |  |
| Felt like an outsider in the family | (Ref: No) |  |  |  | 0.054 |  |  |  |  | 0.582 |
|  | Yes | 1.13 | (1.00,1.29) | 0.07 |  |  | 0.98 | (0.89,1.07) | 0.05 |  |
| Self-rated health growing up | (Ref: Good) |  |  |  | 0.056 |  |  |  |  | 0.538 |
|  | Excellent | 1.05 | (0.92,1.19) | 0.06 |  |  | 0.95 | (0.88,1.04) | 0.04 |  |
|  | Very good | 0.95 | (0.83,1.09) | 0.07 |  |  | 0.94 | (0.87,1.03) | 0.04 |  |
|  | Fair | 0.82 | (0.68,1.00) | 0.10 |  |  | 0.98 | (0.87,1.10) | 0.06 |  |
|  | Poor | 1.12 | (0.78,1.61) | 0.18 |  |  | 1.08 | (0.87,1.34) | 0.11 |  |
| Immigration status | (Ref: Born in this country) |  |  |  | 0.840 |  |  |  |  | 0.686 |
|  | Born in another country | 0.96 | (0.64,1.45) | 0.21 |  |  | 0.94 | (0.66,1.32) | 0.17 |  |
| Religious service attendance (age 12) | (Ref: Never) |  |  |  | <.001 |  |  |  |  | <.001 |
|  | At least 1/week | 2.15 | (1.74,2.66) | 0.11 |  |  | 1.84 | (1.63,2.09) | 0.06 |  |
|  | 1-3/month | 2.01 | (1.60,2.51) | 0.11 |  |  | 1.27 | (1.10,1.46) | 0.07 |  |
|  | < 1/month | 1.27 | (1.01,1.61) | 0.12 |  |  | 0.78 | (0.67,0.91) | 0.08 |  |
| Year of birth | (Ref: 1998-2005; current age: 18-24) |  |  |  | <.001 |  |  |  |  | <.001 |
|  | 1993-1998; age 25-29 | 0.83 | (0.72,0.96) | 0.07 |  |  | 1.12 | (1.00,1.26) | 0.06 |  |
|  | 1983-1993; age 30-39 | 0.73 | (0.64,0.82) | 0.06 |  |  | 1.11 | (1.01,1.23) | 0.05 |  |
|  | 1973-1983; age 40-49 | 0.71 | (0.62,0.81) | 0.07 |  |  | 1.19 | (1.07,1.32) | 0.05 |  |
|  | 1963-1973; age 50-59 | 0.64 | (0.54,0.75) | 0.08 |  |  | 1.32 | (1.18,1.47) | 0.06 |  |
|  | 1953-1963; age 60-69 | 0.56 | (0.44,0.70) | 0.12 |  |  | 1.39 | (1.22,1.59) | 0.07 |  |
|  | 1943-1953; age 70-79 | 0.71 | (0.50,1.01) | 0.18 |  |  | 1.40 | (1.15,1.71) | 0.10 |  |
|  | 1943 or earlier; age 80+ | 0.77 | (0.41,1.45) | 0.32 |  |  | 1.44 | (1.03,2.02) | 0.17 |  |
| Gender | (Ref: Male) |  |  |  | <.001 |  |  |  |  | 0.027 |
|  | Female | 0.60 | (0.55,0.66) | 0.05 |  |  | 1.08 | (1.01,1.14) | 0.03 |  |
|  | Other | 0.82 | (0.31,2.15) | 0.49 |  |  | 0.56 | (0.23,1.37) | 0.46 |  |
| Religious affiliation | (Ref: No religion/Atheist/Agnostic) |  |  |  | 0.008 |  |  |  |  | 0.002 |
|  | Christianity | 0.77 | (0.65,0.92) | 0.09 |  |  | 0.95 | (0.83,1.10) | 0.07 |  |
|  | Collapsed affiliations with prevalence<3% | 0.87 | (0.68,1.10) | 0.12 |  |  | 1.17 | (0.98,1.40) | 0.09 |  |
| Race/ethnicity | (Ref: Plurality group) |  |  |  | 0.661 |  |  |  |  | <.001 |
|  | Non-plurality groups | 1.01 | (0.91,1.12) | 0.05 |  |  | 1.16 | (1.08,1.24) | 0.03 |  |

Table S3c. Sensitivity of the regression estimates to unmeasured confounding for Brazil

|  | | Secular Community Participation | |  | Religious Service Attendance | |
| --- | --- | --- | --- | --- | --- | --- |
| Variable | Category | E-value for Estimate | E-value for 95% CI |  | E-value for Estimate | E-value for 95% CI |
| Relationship with mother | (Ref: Very bad/somewhat bad) |  |  |  |  |  |
|  | Very good/somewhat good | 1.35 | 1.00 |  | 1.35 | 1.00 |
| Relationship with father | (Ref: Very bad/somewhat bad) |  |  |  |  |  |
|  | Very good/somewhat good | 1.57 | 1.09 |  | 1.45 | 1.11 |
| Parent marital status | (Ref: Parents married) |  |  |  |  |  |
|  | Divorced | 1.13 | 1.00 |  | 1.42 | 1.00 |
|  | Parents were never married | 1.05 | 1.00 |  | 1.44 | 1.00 |
|  | One or both parents had died | 1.15 | 1.00 |  | 1.36 | 1.00 |
| Subjective financial status of family growing up | (Ref: Got by) |  |  |  |  |  |
|  | Lived comfortably | 1.35 | 1.00 |  | 1.09 | 1.00 |
|  | Found it difficult | 1.34 | 1.00 |  | 1.32 | 1.00 |
|  | Found it very difficult | 1.08 | 1.00 |  | 1.42 | 1.00 |
| Experienced Abuse | (Ref: No) |  |  |  |  |  |
|  | Yes | 1.41 | 1.00 |  | 1.06 | 1.00 |
| Felt like an outsider in the family | (Ref: No) |  |  |  |  |  |
|  | Yes | 1.53 | 1.00 |  | 1.19 | 1.00 |
| Self-rated health growing up | (Ref: Good) |  |  |  |  |  |
|  | Excellent | 1.27 | 1.00 |  | 1.27 | 1.00 |
|  | Very good | 1.30 | 1.00 |  | 1.31 | 1.00 |
|  | Fair | 1.73 | 1.00 |  | 1.16 | 1.00 |
|  | Poor | 1.49 | 1.00 |  | 1.37 | 1.00 |
| Immigration status | (Ref: Born in this country) |  |  |  |  |  |
|  | Born in another country | 1.24 | 1.00 |  | 1.34 | 1.00 |
| Religious service attendance (age 12) | (Ref: Never) |  |  |  |  |  |
|  | At least 1/week | 3.73 | 2.87 |  | 3.09 | 2.64 |
|  | 1-3/month | 3.43 | 2.59 |  | 1.85 | 1.44 |
|  | < 1/month | 1.87 | 1.11 |  | 1.88 | 1.42 |
| Year of birth | (Ref: 1998-2005; current age: 18-24) |  |  |  |  |  |
|  | 1993-1998; age 25-29 | 1.70 | 1.25 |  | 1.49 | 1.00 |
|  | 1983-1993; age 30-39 | 2.10 | 1.73 |  | 1.47 | 1.08 |
|  | 1973-1983; age 40-49 | 2.18 | 1.77 |  | 1.66 | 1.35 |
|  | 1963-1973; age 50-59 | 2.52 | 2.00 |  | 1.96 | 1.64 |
|  | 1953-1963; age 60-69 | 2.99 | 2.21 |  | 2.14 | 1.74 |
|  | 1943-1953; age 70-79 | 2.15 | 1.00 |  | 2.15 | 1.55 |
|  | 1943 or earlier; age 80+ | 1.92 | 1.00 |  | 2.24 | 1.20 |
| Gender | (Ref: Male) |  |  |  |  |  |
|  | Female | 2.71 | 2.39 |  | 1.36 | 1.12 |
|  | Other | 1.74 | 1.00 |  | 2.98 | 1.00 |
| Religious affiliation | (Ref: No religion/Atheist/Agnostic) |  |  |  |  |  |
|  | Christianity | 1.92 | 1.41 |  | 1.28 | 1.00 |
|  | Collapsed affiliations with prevalence<3% | 1.58 | 1.00 |  | 1.62 | 1.00 |
| Race/ethnicity | (Ref: Plurality group) |  |  |  |  |  |
|  | Non-plurality groups | 1.11 | 1.00 |  | 1.59 | 1.38 |

Table S4a. Nationally representative descriptive statistics for Egypt

| **Characteristic** | **N = 4,729**^1^ |
| --- | --- |
| **Relationship with mother** |  |
| Very good | 4,110 (87%) |
| Somewhat good | 505 (11%) |
| Somewhat bad | 21 (0.4%) |
| Very bad | 10 (0.2%) |
| Does not apply | 83 (1.8%) |
| (Missing) | 0 (0%) |
| **Relationship with father** |  |
| Very good | 3,713 (79%) |
| Somewhat good | 683 (14%) |
| Somewhat bad | 56 (1.2%) |
| Very bad | 30 (0.6%) |
| Does not apply | 233 (4.9%) |
| (Missing) | 14 (0.3%) |
| **Parent marital status** |  |
| Parents married | 4,049 (86%) |
| Divorced | 131 (2.8%) |
| Parents were never married | 9 (0.2%) |
| One or both parents had died | 485 (10%) |
| (Missing) | 55 (1.2%) |
| **Subjective financial status of family growing up** |  |
| Lived comfortably | 1,251 (26%) |
| Got by | 2,352 (50%) |
| Found it difficult | 857 (18%) |
| Found it very difficult | 268 (5.7%) |
| (Missing) | 1 (<0.1%) |
| **Experienced Abuse** |  |
| Yes | 405 (8.6%) |
| No | 4,293 (91%) |
| (Missing) | 30 (0.6%) |
| **Felt like an outsider in the family** |  |
| Yes | 260 (5.5%) |
| No | 4,456 (94%) |
| (Missing) | 13 (0.3%) |
| **Self-rated health growing up** |  |
| Excellent | 2,687 (57%) |
| Very good | 1,174 (25%) |
| Good | 497 (11%) |
| Fair | 265 (5.6%) |
| Poor | 106 (2.2%) |
| (Missing) | 1 (<0.1%) |
| **Immigration status** |  |
| Born in this country | 4,713 (100%) |
| Born in another country | 16 (0.3%) |
| (Missing) | 1 (<0.1%) |
| **Religious service attendance (age 12)** |  |
| At least 1/week | 2,307 (49%) |
| 1-3/month | 570 (12%) |
| <1/month | 629 (13%) |
| Never | 1,165 (25%) |
| (Missing) | 57 (1.2%) |
| **Year of birth** |  |
| 1998-2005; age 18-24 | 960 (20%) |
| 1993-1998; age 25-29 | 607 (13%) |
| 1983-1993; age 30-39 | 1,204 (25%) |
| 1973-1983; age 40-49 | 897 (19%) |
| 1963-1973; age 50-59 | 613 (13%) |
| 1953-1963; age 60-69 | 387 (8.2%) |
| 1943-1953; age 70-79 | 54 (1.1%) |
| 1943 or earlier; age 80+ | 7 (0.2%) |
| (Missing) | 0 (0%) |
| **Gender** |  |
| Male | 2,394 (51%) |
| Female | 2,334 (49%) |
| Other | 0 (0%) |
| (Missing) | 0 (<0.1%) |
| **Religious affiliation** |  |
| Christianity | 123 (2.6%) |
| Islam | 4,602 (97%) |
| Hinduism | 0 (0%) |
| Buddhism | 0 (0%) |
| Judaism | 0 (0%) |
| Sikhism | 0 (0%) |
| Baha'i | 0 (0%) |
| Jainism | 1 (<0.1%) |
| Shinto | 0 (0%) |
| Taoism | 0 (<0.1%) |
| Confucianism | 0 (0%) |
| Primal, Animist, or Folk religion | 0 (0%) |
| Spiritism | 0 (0%) |
| Umbanda, Candomble, and other African-derived religions | 0 (0%) |
| Chinese folk/traditional religion | 0 (0%) |
| Some other religion | 0 (0%) |
| No religion/Atheist/Agnostic | 0 (0%) |
| (Missing) | 3 (<0.1%) |
| **Race/Ethnicity** |  |
| Arab | 4,585 (97%) |
| Bedouin Arab | 4 (<0.1%) |
| Greek | 1 (<0.1%) |
| Nubian | 27 (0.6%) |
| Turkish | 9 (0.2%) |
| (Missing) | 102 (2.2%) |
| ^1^n (%) | |

Table S4b. Regression of Community Participation on Childhood Predictors for Egypt

|  | | Secular Community Participation | | | |  | Religious Service Attendance | | | |
| --- | --- | --- | --- | --- | --- | --- | --- | --- | --- | --- |
| Variable | Category | Risk-Ratio | RR 95% CI | log(RR) SE | Global  p-value |  | Risk-Ratio | RR 95% CI | log(RR) SE | Global  p-value |
| Relationship with mother | (Ref: Very bad/somewhat bad) |  |  |  | 0.457 |  |  |  |  | 0.404 |
|  | Very good/somewhat good | 0.62 | (0.18,2.18) | 0.64 |  |  | 0.85 | (0.56,1.27) | 0.21 |  |
| Relationship with father | (Ref: Very bad/somewhat bad) |  |  |  | 0.210 |  |  |  |  | 0.749 |
|  | Very good/somewhat good | 0.65 | (0.34,1.27) | 0.34 |  |  | 1.05 | (0.76,1.44) | 0.16 |  |
| Parent marital status | (Ref: Parents married) |  |  |  | <.001 |  |  |  |  | 0.013 |
|  | Divorced | 0.89 | (0.41,1.94) | 0.39 |  |  | 0.69 | (0.49,0.96) | 0.17 |  |
|  | Parents were never married | 0.00 | (0.00,0.00) | 0.44 |  |  | 0.81 | (0.41,1.61) | 0.35 |  |
|  | One or both parents had died | 0.91 | (0.55,1.50) | 0.26 |  |  | 1.19 | (1.03,1.37) | 0.07 |  |
| Subjective financial status of family growing up | (Ref: Got by) |  |  |  | 0.541 |  |  |  |  | 0.113 |
|  | Lived comfortably | 1.23 | (0.90,1.68) | 0.16 |  |  | 0.94 | (0.85,1.04) | 0.05 |  |
|  | Found it difficult | 1.22 | (0.84,1.77) | 0.19 |  |  | 1.12 | (0.99,1.26) | 0.06 |  |
|  | Found it very difficult | 1.32 | (0.71,2.47) | 0.32 |  |  | 1.10 | (0.90,1.33) | 0.10 |  |
| Experienced Abuse | (Ref: No) |  |  |  | 0.066 |  |  |  |  | 0.020 |
|  | Yes | 1.44 | (0.98,2.10) | 0.19 |  |  | 1.17 | (1.03,1.33) | 0.07 |  |
| Felt like an outsider in the family | (Ref: No) |  |  |  | 0.172 |  |  |  |  | 0.226 |
|  | Yes | 1.41 | (0.86,2.30) | 0.25 |  |  | 0.89 | (0.73,1.08) | 0.10 |  |
| Self-rated health growing up | (Ref: Good) |  |  |  | 0.282 |  |  |  |  | 0.081 |
|  | Excellent | 1.26 | (0.79,2.01) | 0.24 |  |  | 1.10 | (0.94,1.27) | 0.08 |  |
|  | Very good | 0.88 | (0.55,1.42) | 0.24 |  |  | 1.06 | (0.89,1.27) | 0.09 |  |
|  | Fair | 0.94 | (0.49,1.79) | 0.33 |  |  | 0.85 | (0.68,1.07) | 0.12 |  |
|  | Poor | 1.17 | (0.51,2.69) | 0.43 |  |  | 1.05 | (0.79,1.40) | 0.15 |  |
| Immigration status | (Ref: Born in this country) |  |  |  | 0.303 |  |  |  |  | 0.646 |
|  | Born in another country | 2.30 | (0.48,11.09) | 0.80 |  |  | 1.16 | (0.61,2.20) | 0.33 |  |
| Religious service attendance (age 12) | (Ref: Never) |  |  |  | 0.077 |  |  |  |  | 0.004 |
|  | At least 1/week | 1.55 | (1.03,2.33) | 0.21 |  |  | 1.29 | (1.11,1.49) | 0.07 |  |
|  | 1-3/month | 1.16 | (0.71,1.91) | 0.25 |  |  | 1.22 | (1.04,1.43) | 0.08 |  |
|  | < 1/month | 1.37 | (0.83,2.27) | 0.26 |  |  | 1.04 | (0.87,1.25) | 0.09 |  |
| Year of birth | (Ref: 1998-2005; current age: 18-24) |  |  |  | <.001 |  |  |  |  | 0.109 |
|  | 1993-1998; age 25-29 | 0.46 | (0.31,0.69) | 0.21 |  |  | 0.93 | (0.79,1.09) | 0.08 |  |
|  | 1983-1993; age 30-39 | 0.37 | (0.27,0.49) | 0.15 |  |  | 0.85 | (0.74,0.97) | 0.07 |  |
|  | 1973-1983; age 40-49 | 0.28 | (0.19,0.41) | 0.19 |  |  | 0.86 | (0.75,0.98) | 0.07 |  |
|  | 1963-1973; age 50-59 | 0.28 | (0.17,0.46) | 0.25 |  |  | 0.89 | (0.79,1.02) | 0.07 |  |
|  | 1953-1963; age 60-69 | 0.11 | (0.04,0.32) | 0.57 |  |  | 0.97 | (0.81,1.16) | 0.09 |  |
|  | 1943-1953; age 70-79 | 0.15 | (0.03,0.68) | 0.78 |  |  | 0.57 | (0.35,0.94) | 0.25 |  |
|  | 1943 or earlier; age 80+ | 0.00 | (0.00,0.00) | 0.69 |  |  | 0.72 | (0.18,2.97) | 0.72 |  |
| Gender | (Ref: Male) |  |  |  | <.001 |  |  |  |  | <.001 |
|  | Female | 0.46 | (0.34,0.61) | 0.14 |  |  | 0.57 | (0.51,0.63) | 0.06 |  |
| Religious affiliation | (Ref: Islam) |  |  |  | 0.268 |  |  |  |  | <.001 |
|  | Collapsed affiliations with prevalence<3% | 1.41 | (0.77,2.56) | 0.31 |  |  | 1.70 | (1.44,2.01) | 0.09 |  |
| Race/ethnicity | (Ref: Plurality group) |  |  |  | 0.461 |  |  |  |  | 0.469 |
|  | Non-plurality groups | 1.34 | (0.61,2.94) | 0.40 |  |  | 0.77 | (0.39,1.54) | 0.35 |  |

Table S4c. Sensitivity of the regression estimates to unmeasured confounding for Egypt

|  | | Secular Community Participation | |  | Religious Service Attendance | |
| --- | --- | --- | --- | --- | --- | --- |
| Variable | Category | E-value for Estimate | E-value for 95% CI |  | E-value for Estimate | E-value for 95% CI |
| Relationship with mother | (Ref: Very bad/somewhat bad) |  |  |  |  |  |
|  | Very good/somewhat good | 2.61 | 1.00 |  | 1.64 | 1.00 |
| Relationship with father | (Ref: Very bad/somewhat bad) |  |  |  |  |  |
|  | Very good/somewhat good | 2.43 | 1.00 |  | 1.27 | 1.00 |
| Parent marital status | (Ref: Parents married) |  |  |  |  |  |
|  | Divorced | 1.48 | 1.00 |  | 2.27 | 1.25 |
|  | Parents were never married | 756576.62 | 320368.63 |  | 1.78 | 1.00 |
|  | One or both parents had died | 1.43 | 1.00 |  | 1.66 | 1.19 |
| Subjective financial status of family growing up | (Ref: Got by) |  |  |  |  |  |
|  | Lived comfortably | 1.76 | 1.00 |  | 1.33 | 1.00 |
|  | Found it difficult | 1.74 | 1.00 |  | 1.48 | 1.00 |
|  | Found it very difficult | 1.98 | 1.00 |  | 1.43 | 1.00 |
| Experienced Abuse | (Ref: No) |  |  |  |  |  |
|  | Yes | 2.23 | 1.00 |  | 1.61 | 1.19 |
| Felt like an outsider in the family | (Ref: No) |  |  |  |  |  |
|  | Yes | 2.17 | 1.00 |  | 1.50 | 1.00 |
| Self-rated health growing up | (Ref: Good) |  |  |  |  |  |
|  | Excellent | 1.83 | 1.00 |  | 1.42 | 1.00 |
|  | Very good | 1.52 | 1.00 |  | 1.32 | 1.00 |
|  | Fair | 1.33 | 1.00 |  | 1.63 | 1.00 |
|  | Poor | 1.61 | 1.00 |  | 1.28 | 1.00 |
| Immigration status | (Ref: Born in this country) |  |  |  |  |  |
|  | Born in another country | 4.03 | 1.00 |  | 1.60 | 1.00 |
| Religious service attendance (age 12) | (Ref: Never) |  |  |  |  |  |
|  | At least 1/week | 2.47 | 1.21 |  | 1.90 | 1.47 |
|  | 1-3/month | 1.60 | 1.00 |  | 1.74 | 1.25 |
|  | < 1/month | 2.09 | 1.00 |  | 1.25 | 1.00 |
| Year of birth | (Ref: 1998-2005; current age: 18-24) |  |  |  |  |  |
|  | 1993-1998; age 25-29 | 3.74 | 2.25 |  | 1.38 | 1.00 |
|  | 1983-1993; age 30-39 | 4.89 | 3.48 |  | 1.65 | 1.22 |
|  | 1973-1983; age 40-49 | 6.59 | 4.37 |  | 1.61 | 1.16 |
|  | 1963-1973; age 50-59 | 6.56 | 3.73 |  | 1.48 | 1.00 |
|  | 1953-1963; age 60-69 | 18.22 | 5.63 |  | 1.22 | 1.00 |
|  | 1943-1953; age 70-79 | 13.06 | 2.30 |  | 2.90 | 1.32 |
|  | 1943 or earlier; age 80+ | 1531464.84 | 392737.04 |  | 2.11 | 1.00 |
| Gender | (Ref: Male) |  |  |  |  |  |
|  | Female | 3.80 | 2.68 |  | 2.93 | 2.53 |
| Religious affiliation | (Ref: Islam) |  |  |  |  |  |
|  | Collapsed affiliations with prevalence<3% | 2.16 | 1.00 |  | 2.80 | 2.24 |
| Race/ethnicity | (Ref: Plurality group) |  |  |  |  |  |
|  | Non-plurality groups | 2.02 | 1.00 |  | 1.91 | 1.00 |

Table S5a. Nationally representative descriptive statistics for Germany

| **Characteristic** | **N = 9,506**^1^ |
| --- | --- |
| **Relationship with mother** |  |
| Very good | 5,497 (58%) |
| Somewhat good | 3,031 (32%) |
| Somewhat bad | 496 (5.2%) |
| Very bad | 187 (2.0%) |
| Does not apply | 241 (2.5%) |
| (Missing) | 54 (0.6%) |
| **Relationship with father** |  |
| Very good | 4,652 (49%) |
| Somewhat good | 3,012 (32%) |
| Somewhat bad | 846 (8.9%) |
| Very bad | 385 (4.0%) |
| Does not apply | 538 (5.7%) |
| (Missing) | 73 (0.8%) |
| **Parent marital status** |  |
| Parents married | 7,620 (80%) |
| Divorced | 927 (9.8%) |
| Parents were never married | 578 (6.1%) |
| One or both parents had died | 245 (2.6%) |
| (Missing) | 136 (1.4%) |
| **Subjective financial status of family growing up** |  |
| Lived comfortably | 3,177 (33%) |
| Got by | 4,508 (47%) |
| Found it difficult | 1,481 (16%) |
| Found it very difficult | 314 (3.3%) |
| (Missing) | 26 (0.3%) |
| **Experienced Abuse** |  |
| Yes | 1,086 (11%) |
| No | 8,321 (88%) |
| (Missing) | 99 (1.0%) |
| **Felt like an outsider in the family** |  |
| Yes | 1,105 (12%) |
| No | 8,262 (87%) |
| (Missing) | 139 (1.5%) |
| **Self-rated health growing up** |  |
| Excellent | 2,633 (28%) |
| Very good | 3,518 (37%) |
| Good | 2,582 (27%) |
| Fair | 612 (6.4%) |
| Poor | 134 (1.4%) |
| (Missing) | 26 (0.3%) |
| **Immigration status** |  |
| Born in this country | 8,722 (92%) |
| Born in another country | 744 (7.8%) |
| (Missing) | 40 (0.4%) |
| **Religious service attendance (age 12)** |  |
| At least 1/week | 1,943 (20%) |
| 1-3/month | 1,899 (20%) |
| <1/month | 2,887 (30%) |
| Never | 2,749 (29%) |
| (Missing) | 27 (0.3%) |
| **Year of birth** |  |
| 1998-2005; age 18-24 | 829 (8.7%) |
| 1993-1998; age 25-29 | 774 (8.1%) |
| 1983-1993; age 30-39 | 1,438 (15%) |
| 1973-1983; age 40-49 | 1,494 (16%) |
| 1963-1973; age 50-59 | 1,729 (18%) |
| 1953-1963; age 60-69 | 1,915 (20%) |
| 1943-1953; age 70-79 | 1,137 (12%) |
| 1943 or earlier; age 80+ | 190 (2.0%) |
| (Missing) | 0 (0%) |
| **Gender** |  |
| Male | 4,641 (49%) |
| Female | 4,843 (51%) |
| Other | 11 (0.1%) |
| (Missing) | 11 (0.1%) |
| **Religious affiliation** |  |
| Christianity | 5,751 (61%) |
| Islam | 350 (3.7%) |
| Hinduism | 15 (0.2%) |
| Buddhism | 25 (0.3%) |
| Judaism | 18 (0.2%) |
| Sikhism | 5 (<0.1%) |
| Baha'i | 2 (<0.1%) |
| Jainism | 1 (<0.1%) |
| Shinto | 0 (0%) |
| Taoism | 0 (0%) |
| Confucianism | 4 (<0.1%) |
| Primal, Animist, or Folk religion | 19 (0.2%) |
| Spiritism | 0 (0%) |
| Umbanda, Candomble, and other African-derived religions | 0 (0%) |
| Chinese folk/traditional religion | 0 (0%) |
| Some other religion | 67 (0.7%) |
| No religion/Atheist/Agnostic | 3,163 (33%) |
| (Missing) | 85 (0.9%) |
| ^1^n (%) | |

Table S5b. Regression of Community Participation on Childhood Predictors for Germany

|  | | Secular Community Participation | | | |  | Religious Service Attendance | | | |
| --- | --- | --- | --- | --- | --- | --- | --- | --- | --- | --- |
| Variable | Category | Risk-Ratio | RR 95% CI | log(RR) SE | Global  p-value |  | Risk-Ratio | RR 95% CI | log(RR) SE | Global  p-value |
| Relationship with mother | (Ref: Very bad/somewhat bad) |  |  |  | 0.025 |  |  |  |  | 0.027 |
|  | Very good/somewhat good | 1.25 | (1.03,1.51) | 0.10 |  |  | 1.52 | (1.02,2.26) | 0.20 |  |
| Relationship with father | (Ref: Very bad/somewhat bad) |  |  |  | 0.805 |  |  |  |  | 0.015 |
|  | Very good/somewhat good | 0.99 | (0.84,1.15) | 0.08 |  |  | 1.47 | (1.07,2.01) | 0.16 |  |
| Parent marital status | (Ref: Parents married) |  |  |  | 0.898 |  |  |  |  | 0.014 |
|  | Divorced | 1.04 | (0.89,1.22) | 0.08 |  |  | 0.78 | (0.54,1.12) | 0.18 |  |
|  | Parents were never married | 0.97 | (0.78,1.20) | 0.11 |  |  | 1.50 | (1.07,2.11) | 0.17 |  |
|  | One or both parents had died | 1.06 | (0.79,1.42) | 0.15 |  |  | 0.64 | (0.30,1.36) | 0.39 |  |
| Subjective financial status of family growing up | (Ref: Got by) |  |  |  | 0.351 |  |  |  |  | 0.221 |
|  | Lived comfortably | 1.09 | (0.97,1.23) | 0.06 |  |  | 1.22 | (0.98,1.53) | 0.11 |  |
|  | Found it difficult | 1.04 | (0.89,1.21) | 0.08 |  |  | 1.06 | (0.79,1.44) | 0.15 |  |
|  | Found it very difficult | 0.89 | (0.66,1.19) | 0.15 |  |  | 0.70 | (0.34,1.48) | 0.38 |  |
| Experienced Abuse | (Ref: No) |  |  |  | 0.860 |  |  |  |  | 0.607 |
|  | Yes | 1.01 | (0.87,1.18) | 0.08 |  |  | 1.08 | (0.81,1.43) | 0.15 |  |
| Felt like an outsider in the family | (Ref: No) |  |  |  | 0.879 |  |  |  |  | 0.246 |
|  | Yes | 0.99 | (0.86,1.15) | 0.08 |  |  | 1.15 | (0.91,1.45) | 0.12 |  |
| Self-rated health growing up | (Ref: Good) |  |  |  | 0.295 |  |  |  |  | 0.841 |
|  | Excellent | 1.16 | (1.00,1.34) | 0.07 |  |  | 0.85 | (0.66,1.11) | 0.14 |  |
|  | Very good | 1.09 | (0.96,1.24) | 0.07 |  |  | 0.93 | (0.73,1.18) | 0.12 |  |
|  | Fair | 1.06 | (0.85,1.32) | 0.11 |  |  | 0.92 | (0.60,1.41) | 0.22 |  |
|  | Poor | 1.29 | (0.86,1.94) | 0.21 |  |  | 0.98 | (0.44,2.21) | 0.41 |  |
| Immigration status | (Ref: Born in this country) |  |  |  | 0.772 |  |  |  |  | 0.006 |
|  | Born in another country | 1.03 | (0.85,1.25) | 0.10 |  |  | 1.55 | (1.13,2.13) | 0.16 |  |
| Religious service attendance (age 12) | (Ref: Never) |  |  |  | <.001 |  |  |  |  | <.001 |
|  | At least 1/week | 1.38 | (1.19,1.60) | 0.08 |  |  | 21.73 | (13.01,36.29) | 0.26 |  |
|  | 1-3/month | 1.27 | (1.10,1.48) | 0.08 |  |  | 13.53 | (8.00,22.89) | 0.27 |  |
|  | < 1/month | 1.20 | (1.05,1.38) | 0.07 |  |  | 2.21 | (1.20,4.05) | 0.31 |  |
| Year of birth | (Ref: 1998-2005; current age: 18-24) |  |  |  | <.001 |  |  |  |  | 0.455 |
|  | 1993-1998; age 25-29 | 1.07 | (0.88,1.30) | 0.10 |  |  | 0.79 | (0.52,1.21) | 0.22 |  |
|  | 1983-1993; age 30-39 | 0.82 | (0.68,0.98) | 0.09 |  |  | 0.99 | (0.69,1.41) | 0.18 |  |
|  | 1973-1983; age 40-49 | 0.78 | (0.64,0.94) | 0.10 |  |  | 0.86 | (0.60,1.25) | 0.19 |  |
|  | 1963-1973; age 50-59 | 0.66 | (0.54,0.80) | 0.10 |  |  | 0.89 | (0.62,1.29) | 0.19 |  |
|  | 1953-1963; age 60-69 | 0.77 | (0.63,0.93) | 0.10 |  |  | 0.73 | (0.49,1.08) | 0.20 |  |
|  | 1943-1953; age 70-79 | 0.80 | (0.65,0.99) | 0.11 |  |  | 0.79 | (0.52,1.21) | 0.21 |  |
|  | 1943 or earlier; age 80+ | 0.58 | (0.39,0.86) | 0.20 |  |  | 0.57 | (0.29,1.15) | 0.35 |  |
| Gender | (Ref: Male) |  |  |  | 0.001 |  |  |  |  | <.001 |
|  | Female | 0.92 | (0.83,1.01) | 0.05 |  |  | 0.82 | (0.68,0.99) | 0.09 |  |
|  | Other | 2.46 | (1.36,4.47) | 0.30 |  |  | 0.00 | (0.00,0.00) | 0.60 |  |
| Religious affiliation | (Ref: No religion/Atheist/Agnostic) |  |  |  | 0.614 |  |  |  |  | <.001 |
|  | Islam | 0.90 | (0.68,1.19) | 0.14 |  |  | 1.27 | (0.88,1.82) | 0.19 |  |
|  | Christianity | 0.96 | (0.86,1.08) | 0.06 |  |  | 0.68 | (0.54,0.85) | 0.11 |  |
|  | Collapsed affiliations with prevalence<3% | 1.17 | (0.80,1.69) | 0.19 |  |  | 1.58 | (0.89,2.81) | 0.29 |  |

Table S5c. Sensitivity of the regression estimates to unmeasured confounding for Germany

|  | | Secular Community Participation | |  | Religious Service Attendance | |
| --- | --- | --- | --- | --- | --- | --- |
| Variable | Category | E-value for Estimate | E-value for 95% CI |  | E-value for Estimate | E-value for 95% CI |
| Relationship with mother | (Ref: Very bad/somewhat bad) |  |  |  |  |  |
|  | Very good/somewhat good | 1.80 | 1.19 |  | 2.41 | 1.16 |
| Relationship with father | (Ref: Very bad/somewhat bad) |  |  |  |  |  |
|  | Very good/somewhat good | 1.14 | 1.00 |  | 2.30 | 1.36 |
| Parent marital status | (Ref: Parents married) |  |  |  |  |  |
|  | Divorced | 1.26 | 1.00 |  | 1.90 | 1.00 |
|  | Parents were never married | 1.21 | 1.00 |  | 2.38 | 1.35 |
|  | One or both parents had died | 1.31 | 1.00 |  | 2.50 | 1.00 |
| Subjective financial status of family growing up | (Ref: Got by) |  |  |  |  |  |
|  | Lived comfortably | 1.41 | 1.00 |  | 1.74 | 1.00 |
|  | Found it difficult | 1.24 | 1.00 |  | 1.32 | 1.00 |
|  | Found it very difficult | 1.50 | 1.00 |  | 2.19 | 1.00 |
| Experienced Abuse | (Ref: No) |  |  |  |  |  |
|  | Yes | 1.11 | 1.00 |  | 1.36 | 1.00 |
| Felt like an outsider in the family | (Ref: No) |  |  |  |  |  |
|  | Yes | 1.10 | 1.00 |  | 1.56 | 1.00 |
| Self-rated health growing up | (Ref: Good) |  |  |  |  |  |
|  | Excellent | 1.58 | 1.04 |  | 1.62 | 1.00 |
|  | Very good | 1.40 | 1.00 |  | 1.37 | 1.00 |
|  | Fair | 1.32 | 1.00 |  | 1.41 | 1.00 |
|  | Poor | 1.91 | 1.00 |  | 1.16 | 1.00 |
| Immigration status | (Ref: Born in this country) |  |  |  |  |  |
|  | Born in another country | 1.20 | 1.00 |  | 2.48 | 1.52 |
| Religious service attendance (age 12) | (Ref: Never) |  |  |  |  |  |
|  | At least 1/week | 2.10 | 1.66 |  | 42.95 | 25.51 |
|  | 1-3/month | 1.86 | 1.42 |  | 26.56 | 15.49 |
|  | < 1/month | 1.69 | 1.27 |  | 3.84 | 1.70 |
| Year of birth | (Ref: 1998-2005; current age: 18-24) |  |  |  |  |  |
|  | 1993-1998; age 25-29 | 1.34 | 1.00 |  | 1.85 | 1.00 |
|  | 1983-1993; age 30-39 | 1.75 | 1.16 |  | 1.14 | 1.00 |
|  | 1973-1983; age 40-49 | 1.89 | 1.32 |  | 1.58 | 1.00 |
|  | 1963-1973; age 50-59 | 2.40 | 1.80 |  | 1.48 | 1.00 |
|  | 1953-1963; age 60-69 | 1.92 | 1.34 |  | 2.09 | 1.00 |
|  | 1943-1953; age 70-79 | 1.80 | 1.08 |  | 1.83 | 1.00 |
|  | 1943 or earlier; age 80+ | 2.82 | 1.58 |  | 2.89 | 1.00 |
| Gender | (Ref: Male) |  |  |  |  |  |
|  | Female | 1.41 | 1.00 |  | 1.73 | 1.13 |
|  | Other | 4.36 | 2.05 |  | 670829.80 | 207017.84 |
| Religious affiliation | (Ref: No religion/Atheist/Agnostic) |  |  |  |  |  |
|  | Islam | 1.47 | 1.00 |  | 1.84 | 1.00 |
|  | Christianity | 1.24 | 1.00 |  | 2.31 | 1.63 |
|  | Collapsed affiliations with prevalence<3% | 1.61 | 1.00 |  | 2.55 | 1.00 |

Table S6a. Nationally representative descriptive statistics for Hong Kong

| **Characteristic** | **N = 3,012**^1^ |
| --- | --- |
| **Relationship with mother** |  |
| Very good | 1,077 (36%) |
| Somewhat good | 1,164 (39%) |
| Somewhat bad | 293 (9.7%) |
| Very bad | 49 (1.6%) |
| Does not apply | 426 (14%) |
| (Missing) | 3 (<0.1%) |
| **Relationship with father** |  |
| Very good | 868 (29%) |
| Somewhat good | 1,089 (36%) |
| Somewhat bad | 393 (13%) |
| Very bad | 102 (3.4%) |
| Does not apply | 557 (19%) |
| (Missing) | 3 (0.1%) |
| **Parent marital status** |  |
| Parents married | 2,752 (91%) |
| Divorced | 114 (3.8%) |
| Parents were never married | 40 (1.3%) |
| One or both parents had died | 50 (1.7%) |
| (Missing) | 56 (1.8%) |
| **Subjective financial status of family growing up** |  |
| Lived comfortably | 906 (30%) |
| Got by | 1,527 (51%) |
| Found it difficult | 473 (16%) |
| Found it very difficult | 84 (2.8%) |
| (Missing) | 22 (0.7%) |
| **Experienced Abuse** |  |
| Yes | 318 (11%) |
| No | 2,688 (89%) |
| (Missing) | 5 (0.2%) |
| **Felt like an outsider in the family** |  |
| Yes | 664 (22%) |
| No | 2,224 (74%) |
| (Missing) | 124 (4.1%) |
| **Self-rated health growing up** |  |
| Excellent | 545 (18%) |
| Very good | 1,073 (36%) |
| Good | 863 (29%) |
| Fair | 426 (14%) |
| Poor | 91 (3.0%) |
| (Missing) | 13 (0.4%) |
| **Immigration status** |  |
| Born in this country | 2,637 (88%) |
| Born in another country | 321 (11%) |
| (Missing) | 53 (1.8%) |
| **Religious service attendance (age 12)** |  |
| At least 1/week | 432 (14%) |
| 1-3/month | 528 (18%) |
| <1/month | 753 (25%) |
| Never | 1,295 (43%) |
| (Missing) | 4 (0.1%) |
| **Year of birth** |  |
| 1998-2005; age 18-24 | 217 (7.2%) |
| 1993-1998; age 25-29 | 198 (6.6%) |
| 1983-1993; age 30-39 | 507 (17%) |
| 1973-1983; age 40-49 | 580 (19%) |
| 1963-1973; age 50-59 | 711 (24%) |
| 1953-1963; age 60-69 | 620 (21%) |
| 1943-1953; age 70-79 | 164 (5.5%) |
| 1943 or earlier; age 80+ | 15 (0.5%) |
| (Missing) | 0 (0%) |
| **Gender** |  |
| Male | 1,390 (46%) |
| Female | 1,620 (54%) |
| Other | 2 (<0.1%) |
| (Missing) | 0 (0%) |
| **Religious affiliation** |  |
| Christianity | 715 (24%) |
| Islam | 86 (2.9%) |
| Hinduism | 27 (0.9%) |
| Buddhism | 323 (11%) |
| Judaism | 16 (0.5%) |
| Sikhism | 4 (0.1%) |
| Baha'i | 0 (0%) |
| Jainism | 1 (<0.1%) |
| Shinto | 18 (0.6%) |
| Taoism | 81 (2.7%) |
| Confucianism | 10 (0.3%) |
| Primal, Animist, or Folk religion | 15 (0.5%) |
| Spiritism | 0 (0%) |
| Umbanda, Candomble, and other African-derived religions | 0 (0%) |
| Chinese folk/traditional religion | 108 (3.6%) |
| Some other religion | 5 (0.2%) |
| No religion/Atheist/Agnostic | 1,601 (53%) |
| (Missing) | 1 (<0.1%) |
| **Race/Ethnicity** |  |
| Chinese (Cantonese) | 1,930 (64%) |
| Chinese (Chaoshan) | 201 (6.7%) |
| Chinese (Fujianese) | 117 (3.9%) |
| Chinese (Hakka) | 121 (4.0%) |
| Chinese (Other ethnicity) | 264 (8.8%) |
| Chinese (Shanghainese) | 89 (2.9%) |
| East Asian (Korean, Japanese) | 10 (0.3%) |
| Other | 4 (0.1%) |
| South Asian (Indian, Nepalese, Pakistani) | 17 (0.6%) |
| Southeast Asian (Filipino, Indonesian, Thailand) | 46 (1.5%) |
| Taiwanese | 14 (0.4%) |
| White | 15 (0.5%) |
| (Missing) | 184 (6.1%) |
| ^1^n (%) | |

Table S6b. Regression of Community Participation on Childhood Predictors for Hong Kong

|  | | Secular Community Participation | | | |  | Religious Service Attendance | | | |
| --- | --- | --- | --- | --- | --- | --- | --- | --- | --- | --- |
| Variable | Category | Risk-Ratio | RR 95% CI | log(RR) SE | Global  p-value |  | Risk-Ratio | RR 95% CI | log(RR) SE | Global  p-value |
| Relationship with mother | (Ref: Very bad/somewhat bad) |  |  |  | 0.011 |  |  |  |  | 0.370 |
|  | Very good/somewhat good | 0.73 | (0.57,0.93) | 0.13 |  |  | 0.91 | (0.74,1.12) | 0.10 |  |
| Relationship with father | (Ref: Very bad/somewhat bad) |  |  |  | 0.640 |  |  |  |  | 0.326 |
|  | Very good/somewhat good | 0.94 | (0.71,1.25) | 0.14 |  |  | 1.15 | (0.87,1.51) | 0.14 |  |
| Parent marital status | (Ref: Parents married) |  |  |  | 0.254 |  |  |  |  | 0.366 |
|  | Divorced | 0.58 | (0.30,1.12) | 0.33 |  |  | 0.60 | (0.32,1.13) | 0.32 |  |
|  | Parents were never married | 0.73 | (0.41,1.31) | 0.30 |  |  | 0.76 | (0.36,1.60) | 0.38 |  |
|  | One or both parents had died | 1.49 | (0.35,6.33) | 0.74 |  |  | 1.27 | (0.39,4.14) | 0.60 |  |
| Subjective financial status of family growing up | (Ref: Got by) |  |  |  | 0.001 |  |  |  |  | 0.995 |
|  | Lived comfortably | 1.00 | (0.86,1.17) | 0.08 |  |  | 1.00 | (0.86,1.16) | 0.07 |  |
|  | Found it difficult | 0.51 | (0.29,0.89) | 0.29 |  |  | 0.97 | (0.70,1.33) | 0.16 |  |
|  | Found it very difficult | 0.10 | (0.02,0.41) | 0.73 |  |  | 0.96 | (0.47,1.98) | 0.37 |  |
| Experienced Abuse | (Ref: No) |  |  |  | 0.022 |  |  |  |  | 0.023 |
|  | Yes | 1.28 | (1.03,1.58) | 0.11 |  |  | 1.23 | (1.03,1.47) | 0.09 |  |
| Felt like an outsider in the family | (Ref: No) |  |  |  | <.001 |  |  |  |  | <.001 |
|  | Yes | 1.53 | (1.32,1.78) | 0.08 |  |  | 1.38 | (1.21,1.57) | 0.07 |  |
| Self-rated health growing up | (Ref: Good) |  |  |  | 0.013 |  |  |  |  | 0.002 |
|  | Excellent | 1.34 | (1.00,1.79) | 0.15 |  |  | 1.18 | (0.93,1.52) | 0.13 |  |
|  | Very good | 1.32 | (1.01,1.71) | 0.13 |  |  | 1.24 | (1.01,1.53) | 0.11 |  |
|  | Fair | 0.66 | (0.37,1.16) | 0.29 |  |  | 0.54 | (0.36,0.81) | 0.20 |  |
|  | Poor | 0.54 | (0.19,1.57) | 0.54 |  |  | 1.34 | (0.59,3.08) | 0.42 |  |
| Immigration status | (Ref: Born in this country) |  |  |  | 0.018 |  |  |  |  | 0.877 |
|  | Born in another country | 0.64 | (0.44,0.93) | 0.19 |  |  | 1.00 | (0.70,1.43) | 0.18 |  |
| Religious service attendance (age 12) | (Ref: Never) |  |  |  | <.001 |  |  |  |  | <.001 |
|  | At least 1/week | 5.57 | (3.50,8.86) | 0.24 |  |  | 3.60 | (2.34,5.56) | 0.22 |  |
|  | 1-3/month | 4.27 | (2.80,6.52) | 0.22 |  |  | 2.76 | (1.83,4.16) | 0.21 |  |
|  | < 1/month | 1.22 | (0.77,1.94) | 0.24 |  |  | 1.05 | (0.70,1.60) | 0.21 |  |
| Year of birth | (Ref: 1998-2005; current age: 18-24) |  |  |  | <.001 |  |  |  |  | 0.019 |
|  | 1993-1998; age 25-29 | 0.90 | (0.67,1.21) | 0.15 |  |  | 1.00 | (0.72,1.39) | 0.17 |  |
|  | 1983-1993; age 30-39 | 0.89 | (0.68,1.18) | 0.14 |  |  | 1.27 | (0.97,1.66) | 0.14 |  |
|  | 1973-1983; age 40-49 | 0.87 | (0.65,1.15) | 0.14 |  |  | 1.23 | (0.95,1.59) | 0.13 |  |
|  | 1963-1973; age 50-59 | 0.97 | (0.76,1.25) | 0.13 |  |  | 1.39 | (1.10,1.76) | 0.12 |  |
|  | 1953-1963; age 60-69 | 1.24 | (0.93,1.65) | 0.15 |  |  | 1.52 | (1.16,2.00) | 0.14 |  |
|  | 1943-1953; age 70-79 | 0.76 | (0.37,1.54) | 0.36 |  |  | 1.28 | (0.75,2.20) | 0.27 |  |
|  | 1943 or earlier; age 80+ | 0.00 | (0.00,0.00) | 0.67 |  |  | 2.87 | (1.26,6.54) | 0.42 |  |
| Gender | (Ref: Male) |  |  |  | <.001 |  |  |  |  | <.001 |
|  | Female | 0.90 | (0.77,1.04) | 0.07 |  |  | 0.94 | (0.82,1.07) | 0.07 |  |
|  | Other | 0.00 | (0.00,0.00) | 0.78 |  |  | 0.00 | (0.00,0.00) | 0.87 |  |
| Religious affiliation | (Ref: No religion/Atheist/Agnostic) |  |  |  | <.001 |  |  |  |  | <.001 |
|  | Buddhism | 1.62 | (1.17,2.24) | 0.17 |  |  | 3.10 | (2.16,4.43) | 0.18 |  |
|  | Chinese folk/traditional religion | 1.29 | (0.84,1.99) | 0.22 |  |  | 3.16 | (2.04,4.91) | 0.22 |  |
|  | Christianity | 1.53 | (1.15,2.03) | 0.15 |  |  | 3.51 | (2.43,5.09) | 0.19 |  |
|  | Collapsed affiliations with prevalence<3% | 2.16 | (1.57,2.97) | 0.16 |  |  | 3.68 | (2.53,5.35) | 0.19 |  |
| Race/ethnicity | (Ref: Plurality group) |  |  |  | 0.570 |  |  |  |  | 0.806 |
|  | Non-plurality groups | 1.04 | (0.88,1.23) | 0.09 |  |  | 1.01 | (0.86,1.18) | 0.08 |  |

Table S6c. Sensitivity of the regression estimates to unmeasured confounding for Hong Kong

|  | | Secular Community Participation | |  | Religious Service Attendance | |
| --- | --- | --- | --- | --- | --- | --- |
| Variable | Category | E-value for Estimate | E-value for 95% CI |  | E-value for Estimate | E-value for 95% CI |
| Relationship with mother | (Ref: Very bad/somewhat bad) |  |  |  |  |  |
|  | Very good/somewhat good | 2.09 | 1.35 |  | 1.43 | 1.00 |
| Relationship with father | (Ref: Very bad/somewhat bad) |  |  |  |  |  |
|  | Very good/somewhat good | 1.33 | 1.00 |  | 1.56 | 1.00 |
| Parent marital status | (Ref: Parents married) |  |  |  |  |  |
|  | Divorced | 2.81 | 1.00 |  | 2.69 | 1.00 |
|  | Parents were never married | 2.08 | 1.00 |  | 1.97 | 1.00 |
|  | One or both parents had died | 2.34 | 1.00 |  | 1.86 | 1.00 |
| Subjective financial status of family growing up | (Ref: Got by) |  |  |  |  |  |
|  | Lived comfortably | 1.07 | 1.00 |  | 1.03 | 1.00 |
|  | Found it difficult | 3.37 | 1.49 |  | 1.21 | 1.00 |
|  | Found it very difficult | 19.95 | 4.28 |  | 1.25 | 1.00 |
| Experienced Abuse | (Ref: No) |  |  |  |  |  |
|  | Yes | 1.87 | 1.20 |  | 1.76 | 1.20 |
| Felt like an outsider in the family | (Ref: No) |  |  |  |  |  |
|  | Yes | 2.43 | 1.96 |  | 2.10 | 1.70 |
| Self-rated health growing up | (Ref: Good) |  |  |  |  |  |
|  | Excellent | 2.01 | 1.01 |  | 1.65 | 1.00 |
|  | Very good | 1.96 | 1.13 |  | 1.79 | 1.11 |
|  | Fair | 2.41 | 1.00 |  | 3.08 | 1.77 |
|  | Poor | 3.11 | 1.00 |  | 2.02 | 1.00 |
| Immigration status | (Ref: Born in this country) |  |  |  |  |  |
|  | Born in another country | 2.52 | 1.37 |  | 1.05 | 1.00 |
| Religious service attendance (age 12) | (Ref: Never) |  |  |  |  |  |
|  | At least 1/week | 10.61 | 6.46 |  | 6.67 | 4.10 |
|  | 1-3/month | 8.01 | 5.05 |  | 4.96 | 3.06 |
|  | < 1/month | 1.74 | 1.00 |  | 1.30 | 1.00 |
| Year of birth | (Ref: 1998-2005; current age: 18-24) |  |  |  |  |  |
|  | 1993-1998; age 25-29 | 1.46 | 1.00 |  | 1.03 | 1.00 |
|  | 1983-1993; age 30-39 | 1.48 | 1.00 |  | 1.86 | 1.00 |
|  | 1973-1983; age 40-49 | 1.58 | 1.00 |  | 1.76 | 1.00 |
|  | 1963-1973; age 50-59 | 1.19 | 1.00 |  | 2.12 | 1.42 |
|  | 1953-1963; age 60-69 | 1.78 | 1.00 |  | 2.41 | 1.58 |
|  | 1943-1953; age 70-79 | 1.98 | 1.00 |  | 1.89 | 1.00 |
|  | 1943 or earlier; age 80+ | 6195062.81 | 1674594.40 |  | 5.18 | 1.82 |
| Gender | (Ref: Male) |  |  |  |  |  |
|  | Female | 1.48 | 1.00 |  | 1.32 | 1.00 |
|  | Other | 1906890.57 | 414143.10 |  | 353437.21 | 63970.36 |
| Religious affiliation | (Ref: No religion/Atheist/Agnostic) |  |  |  |  |  |
|  | Buddhism | 2.62 | 1.61 |  | 5.64 | 3.75 |
|  | Chinese folk/traditional religion | 1.91 | 1.00 |  | 5.78 | 3.50 |
|  | Christianity | 2.43 | 1.56 |  | 6.48 | 4.28 |
|  | Collapsed affiliations with prevalence<3% | 3.74 | 2.52 |  | 6.82 | 4.49 |
| Race/ethnicity | (Ref: Plurality group) |  |  |  |  |  |
|  | Non-plurality groups | 1.23 | 1.00 |  | 1.09 | 1.00 |

Table S7a. Nationally representative descriptive statistics for India

| **Characteristic** | **N = 12,765**^1^ |
| --- | --- |
| **Relationship with mother** |  |
| Very good | 11,465 (90%) |
| Somewhat good | 788 (6.2%) |
| Somewhat bad | 88 (0.7%) |
| Very bad | 73 (0.6%) |
| Does not apply | 269 (2.1%) |
| (Missing) | 82 (0.6%) |
| **Relationship with father** |  |
| Very good | 10,923 (86%) |
| Somewhat good | 995 (7.8%) |
| Somewhat bad | 126 (1.0%) |
| Very bad | 100 (0.8%) |
| Does not apply | 481 (3.8%) |
| (Missing) | 141 (1.1%) |
| **Parent marital status** |  |
| Parents married | 5,578 (44%) |
| Divorced | 236 (1.8%) |
| Parents were never married | 1,055 (8.3%) |
| One or both parents had died | 940 (7.4%) |
| (Missing) | 4,956 (39%) |
| **Subjective financial status of family growing up** |  |
| Lived comfortably | 4,946 (39%) |
| Got by | 3,010 (24%) |
| Found it difficult | 2,703 (21%) |
| Found it very difficult | 2,035 (16%) |
| (Missing) | 70 (0.5%) |
| **Experienced Abuse** |  |
| Yes | 1,468 (11%) |
| No | 10,526 (82%) |
| (Missing) | 771 (6.0%) |
| **Felt like an outsider in the family** |  |
| Yes | 1,926 (15%) |
| No | 10,780 (84%) |
| (Missing) | 59 (0.5%) |
| **Self-rated health growing up** |  |
| Excellent | 2,182 (17%) |
| Very good | 3,882 (30%) |
| Good | 4,028 (32%) |
| Fair | 2,202 (17%) |
| Poor | 424 (3.3%) |
| (Missing) | 47 (0.4%) |
| **Immigration status** |  |
| Born in this country | 12,629 (99%) |
| Born in another country | 110 (0.9%) |
| (Missing) | 26 (0.2%) |
| **Religious service attendance (age 12)** |  |
| At least 1/week | 5,288 (41%) |
| 1-3/month | 2,959 (23%) |
| <1/month | 2,719 (21%) |
| Never | 1,478 (12%) |
| (Missing) | 321 (2.5%) |
| **Year of birth** |  |
| 1998-2005; age 18-24 | 2,543 (20%) |
| 1993-1998; age 25-29 | 1,640 (13%) |
| 1983-1993; age 30-39 | 3,109 (24%) |
| 1973-1983; age 40-49 | 2,275 (18%) |
| 1963-1973; age 50-59 | 1,574 (12%) |
| 1953-1963; age 60-69 | 1,188 (9.3%) |
| 1943-1953; age 70-79 | 370 (2.9%) |
| 1943 or earlier; age 80+ | 67 (0.5%) |
| (Missing) | 0 (0%) |
| **Gender** |  |
| Male | 6,473 (51%) |
| Female | 6,292 (49%) |
| Other | 0 (0%) |
| (Missing) | 0 (0%) |
| **Religious affiliation** |  |
| Christianity | 254 (2.0%) |
| Islam | 1,550 (12%) |
| Hinduism | 10,417 (82%) |
| Buddhism | 180 (1.4%) |
| Judaism | 0 (0%) |
| Sikhism | 126 (1.0%) |
| Baha'i | 0 (0%) |
| Jainism | 9 (<0.1%) |
| Shinto | 4 (<0.1%) |
| Taoism | 0 (0%) |
| Confucianism | 0 (0%) |
| Primal, Animist, or Folk religion | 27 (0.2%) |
| Spiritism | 0 (0%) |
| Umbanda, Candomble, and other African-derived religions | 0 (0%) |
| Chinese folk/traditional religion | 0 (0%) |
| Some other religion | 59 (0.5%) |
| No religion/Atheist/Agnostic | 7 (<0.1%) |
| (Missing) | 131 (1.0%) |
| **Race/Ethnicity** |  |
| General | 3,538 (28%) |
| Other backward caste | 4,177 (33%) |
| Schedule caste | 3,599 (28%) |
| Schedule tribe | 1,185 (9.3%) |
| (Missing) | 267 (2.1%) |
| ^1^n (%) | |

Table S7b. Regression of Community Participation on Childhood Predictors for India

|  | | Secular Community Participation | | | |  | Religious Service Attendance | | | |
| --- | --- | --- | --- | --- | --- | --- | --- | --- | --- | --- |
| Variable | Category | Risk-Ratio | RR 95% CI | log(RR) SE | Global p-value |  | Risk-Ratio | RR 95% CI | log(RR) SE | Global p-value |
| Relationship with mother | (Ref: Very bad/somewhat bad) |  |  |  | 0.464 |  |  |  |  | 0.569 |
|  | Very good/somewhat good | 0.88 | (0.62,1.26) | 0.18 |  |  | 1.05 | (0.88,1.25) | 0.09 |  |
| Relationship with father | (Ref: Very bad/somewhat bad) |  |  |  | 0.240 |  |  |  |  | 0.246 |
|  | Very good/somewhat good | 1.27 | (0.84,1.91) | 0.21 |  |  | 1.09 | (0.94,1.27) | 0.08 |  |
| Parent marital status | (Ref: Parents married) |  |  |  | <.001 |  |  |  |  | 0.072 |
|  | Divorced | 1.23 | (0.81,1.85) | 0.21 |  |  | 0.94 | (0.78,1.12) | 0.09 |  |
|  | Parents were never married | 0.96 | (0.82,1.13) | 0.08 |  |  | 0.97 | (0.90,1.04) | 0.04 |  |
|  | One or both parents had died | 0.58 | (0.46,0.73) | 0.12 |  |  | 0.91 | (0.84,0.99) | 0.04 |  |
| Subjective financial status of family growing up | (Ref: Got by) |  |  |  | 0.230 |  |  |  |  | 0.106 |
|  | Lived comfortably | 1.11 | (0.97,1.28) | 0.07 |  |  | 1.05 | (0.98,1.11) | 0.03 |  |
|  | Found it difficult | 0.98 | (0.84,1.15) | 0.08 |  |  | 0.99 | (0.92,1.06) | 0.03 |  |
|  | Found it very difficult | 1.04 | (0.86,1.26) | 0.10 |  |  | 0.96 | (0.89,1.04) | 0.04 |  |
| Experienced Abuse | (Ref: No) |  |  |  | <.001 |  |  |  |  | <.001 |
|  | Yes | 1.53 | (1.33,1.76) | 0.07 |  |  | 1.14 | (1.06,1.21) | 0.03 |  |
| Felt like an outsider in the family | (Ref: No) |  |  |  | 0.682 |  |  |  |  | 0.343 |
|  | Yes | 0.97 | (0.83,1.13) | 0.08 |  |  | 1.03 | (0.97,1.11) | 0.03 |  |
| Self-rated health growing up | (Ref: Good) |  |  |  | 0.959 |  |  |  |  | 0.322 |
|  | Excellent | 1.00 | (0.84,1.18) | 0.09 |  |  | 1.06 | (1.00,1.13) | 0.03 |  |
|  | Very good | 1.02 | (0.90,1.17) | 0.07 |  |  | 1.02 | (0.96,1.07) | 0.03 |  |
|  | Fair | 0.98 | (0.83,1.15) | 0.08 |  |  | 1.02 | (0.95,1.10) | 0.04 |  |
|  | Poor | 0.93 | (0.66,1.30) | 0.17 |  |  | 1.08 | (0.96,1.22) | 0.06 |  |
| Immigration status | (Ref: Born in this country) |  |  |  | 0.375 |  |  |  |  | 0.109 |
|  | Born in another country | 1.31 | (0.72,2.38) | 0.30 |  |  | 1.16 | (0.97,1.39) | 0.09 |  |
| Religious service attendance (age 12) | (Ref: Never) |  |  |  | 0.028 |  |  |  |  | <.001 |
|  | At least 1/week | 1.34 | (1.10,1.62) | 0.10 |  |  | 1.36 | (1.25,1.48) | 0.04 |  |
|  | 1-3/month | 1.26 | (1.02,1.56) | 0.11 |  |  | 1.20 | (1.10,1.32) | 0.05 |  |
|  | < 1/month | 1.25 | (1.02,1.54) | 0.10 |  |  | 1.13 | (1.04,1.23) | 0.04 |  |
| Year of birth | (Ref: 1998-2005; current age: 18-24) |  |  |  | <.001 |  |  |  |  | <.001 |
|  | 1993-1998; age 25-29 | 0.97 | (0.82,1.14) | 0.08 |  |  | 1.08 | (1.00,1.16) | 0.04 |  |
|  | 1983-1993; age 30-39 | 0.83 | (0.72,0.96) | 0.07 |  |  | 1.09 | (1.02,1.17) | 0.04 |  |
|  | 1973-1983; age 40-49 | 0.67 | (0.56,0.79) | 0.09 |  |  | 1.12 | (1.04,1.20) | 0.04 |  |
|  | 1963-1973; age 50-59 | 0.73 | (0.60,0.89) | 0.10 |  |  | 1.17 | (1.07,1.28) | 0.05 |  |
|  | 1953-1963; age 60-69 | 0.77 | (0.60,0.98) | 0.13 |  |  | 1.23 | (1.11,1.35) | 0.05 |  |
|  | 1943-1953; age 70-79 | 0.73 | (0.48,1.11) | 0.21 |  |  | 1.26 | (1.10,1.45) | 0.07 |  |
|  | 1943 or earlier; age 80+ | 1.02 | (0.49,2.12) | 0.37 |  |  | 1.38 | (1.06,1.81) | 0.14 |  |
| Gender | (Ref: Male) |  |  |  | <.001 |  |  |  |  | 0.050 |
|  | Female | 0.62 | (0.55,0.70) | 0.06 |  |  | 0.95 | (0.91,1.00) | 0.02 |  |
| Religious affiliation | (Ref: Hinduism) |  |  |  | 0.471 |  |  |  |  | <.001 |
|  | Islam | 0.89 | (0.73,1.09) | 0.10 |  |  | 1.25 | (1.16,1.34) | 0.04 |  |
|  | Collapsed affiliations with prevalence<3% | 1.04 | (0.83,1.31) | 0.12 |  |  | 1.14 | (1.04,1.26) | 0.05 |  |
| Race/ethnicity | (Ref: Plurality group) |  |  |  | 0.757 |  |  |  |  | 0.209 |
|  | Non-plurality groups | 1.02 | (0.90,1.14) | 0.06 |  |  | 0.97 | (0.92,1.02) | 0.03 |  |

Table S7c. Sensitivity of the regression estimates to unmeasured confounding for India

|  | | Secular Community Participation | |  | Religious Service Attendance | |
| --- | --- | --- | --- | --- | --- | --- |
| Variable | Category | E-value for Estimate | E-value for 95% CI |  | E-value for Estimate | E-value for 95% CI |
| Relationship with mother | (Ref: Very bad/somewhat bad) |  |  |  |  |  |
|  | Very good/somewhat good | 1.53 | 1.00 |  | 1.27 | 1.00 |
| Relationship with father | (Ref: Very bad/somewhat bad) |  |  |  |  |  |
|  | Very good/somewhat good | 1.85 | 1.00 |  | 1.41 | 1.00 |
| Parent marital status | (Ref: Parents married) |  |  |  |  |  |
|  | Divorced | 1.76 | 1.00 |  | 1.33 | 1.00 |
|  | Parents were never married | 1.24 | 1.00 |  | 1.21 | 1.00 |
|  | One or both parents had died | 2.83 | 2.08 |  | 1.41 | 1.11 |
| Subjective financial status of family growing up | (Ref: Got by) |  |  |  |  |  |
|  | Lived comfortably | 1.46 | 1.00 |  | 1.27 | 1.00 |
|  | Found it difficult | 1.15 | 1.00 |  | 1.12 | 1.00 |
|  | Found it very difficult | 1.25 | 1.00 |  | 1.24 | 1.00 |
| Experienced Abuse | (Ref: No) |  |  |  |  |  |
|  | Yes | 2.42 | 1.98 |  | 1.53 | 1.32 |
| Felt like an outsider in the family | (Ref: No) |  |  |  |  |  |
|  | Yes | 1.21 | 1.00 |  | 1.22 | 1.00 |
| Self-rated health growing up | (Ref: Good) |  |  |  |  |  |
|  | Excellent | 1.06 | 1.00 |  | 1.32 | 1.00 |
|  | Very good | 1.17 | 1.00 |  | 1.15 | 1.00 |
|  | Fair | 1.17 | 1.00 |  | 1.17 | 1.00 |
|  | Poor | 1.37 | 1.00 |  | 1.38 | 1.00 |
| Immigration status | (Ref: Born in this country) |  |  |  |  |  |
|  | Born in another country | 1.95 | 1.00 |  | 1.59 | 1.00 |
| Religious service attendance (age 12) | (Ref: Never) |  |  |  |  |  |
|  | At least 1/week | 2.01 | 1.44 |  | 2.06 | 1.82 |
|  | 1-3/month | 1.83 | 1.15 |  | 1.69 | 1.42 |
|  | < 1/month | 1.82 | 1.18 |  | 1.52 | 1.24 |
| Year of birth | (Ref: 1998-2005; current age: 18-24) |  |  |  |  |  |
|  | 1993-1998; age 25-29 | 1.22 | 1.00 |  | 1.36 | 1.00 |
|  | 1983-1993; age 30-39 | 1.70 | 1.26 |  | 1.41 | 1.16 |
|  | 1973-1983; age 40-49 | 2.37 | 1.84 |  | 1.48 | 1.24 |
|  | 1963-1973; age 50-59 | 2.08 | 1.49 |  | 1.62 | 1.34 |
|  | 1953-1963; age 60-69 | 1.93 | 1.15 |  | 1.75 | 1.46 |
|  | 1943-1953; age 70-79 | 2.09 | 1.00 |  | 1.84 | 1.44 |
|  | 1943 or earlier; age 80+ | 1.17 | 1.00 |  | 2.11 | 1.31 |
| Gender | (Ref: Male) |  |  |  |  |  |
|  | Female | 2.60 | 2.21 |  | 1.27 | 1.00 |
| Religious affiliation | (Ref: Hinduism) |  |  |  |  |  |
|  | Islam | 1.49 | 1.00 |  | 1.80 | 1.59 |
|  | Collapsed affiliations with prevalence<3% | 1.25 | 1.00 |  | 1.55 | 1.23 |
| Race/ethnicity | (Ref: Plurality group) |  |  |  |  |  |
|  | Non-plurality groups | 1.15 | 1.00 |  | 1.22 | 1.00 |

Table S8a. Nationally representative descriptive statistics for Indonesia

| **Characteristic** | **N = 6,992**^1^ |
| --- | --- |
| **Relationship with mother** |  |
| Very good | 6,238 (89%) |
| Somewhat good | 583 (8.3%) |
| Somewhat bad | 50 (0.7%) |
| Very bad | 26 (0.4%) |
| Does not apply | 68 (1.0%) |
| (Missing) | 27 (0.4%) |
| **Relationship with father** |  |
| Very good | 6,067 (87%) |
| Somewhat good | 628 (9.0%) |
| Somewhat bad | 68 (1.0%) |
| Very bad | 52 (0.7%) |
| Does not apply | 115 (1.6%) |
| (Missing) | 61 (0.9%) |
| **Parent marital status** |  |
| Parents married | 5,557 (79%) |
| Divorced | 448 (6.4%) |
| Parents were never married | 47 (0.7%) |
| One or both parents had died | 735 (11%) |
| (Missing) | 205 (2.9%) |
| **Subjective financial status of family growing up** |  |
| Lived comfortably | 3,408 (49%) |
| Got by | 2,955 (42%) |
| Found it difficult | 439 (6.3%) |
| Found it very difficult | 181 (2.6%) |
| (Missing) | 9 (0.1%) |
| **Experienced Abuse** |  |
| Yes | 486 (6.9%) |
| No | 6,427 (92%) |
| (Missing) | 79 (1.1%) |
| **Felt like an outsider in the family** |  |
| Yes | 343 (4.9%) |
| No | 6,639 (95%) |
| (Missing) | 10 (0.1%) |
| **Self-rated health growing up** |  |
| Excellent | 1,246 (18%) |
| Very good | 1,968 (28%) |
| Good | 2,490 (36%) |
| Fair | 1,233 (18%) |
| Poor | 55 (0.8%) |
| (Missing) | 1 (<0.1%) |
| **Immigration status** |  |
| Born in this country | 6,958 (100%) |
| Born in another country | 34 (0.5%) |
| (Missing) | 0 (0%) |
| **Religious service attendance (age 12)** |  |
| At least 1/week | 5,363 (77%) |
| 1-3/month | 973 (14%) |
| <1/month | 329 (4.7%) |
| Never | 275 (3.9%) |
| (Missing) | 51 (0.7%) |
| **Year of birth** |  |
| 1998-2005; age 18-24 | 1,216 (17%) |
| 1993-1998; age 25-29 | 849 (12%) |
| 1983-1993; age 30-39 | 1,591 (23%) |
| 1973-1983; age 40-49 | 1,576 (23%) |
| 1963-1973; age 50-59 | 1,169 (17%) |
| 1953-1963; age 60-69 | 490 (7.0%) |
| 1943-1953; age 70-79 | 83 (1.2%) |
| 1943 or earlier; age 80+ | 17 (0.2%) |
| (Missing) | 0 (0%) |
| **Gender** |  |
| Male | 3,461 (50%) |
| Female | 3,513 (50%) |
| Other | 7 (<0.1%) |
| (Missing) | 11 (0.2%) |
| **Religious affiliation** |  |
| Christianity | 528 (7.6%) |
| Islam | 6,373 (91%) |
| Hinduism | 75 (1.1%) |
| Buddhism | 5 (<0.1%) |
| Judaism | 0 (0%) |
| Sikhism | 0 (0%) |
| Baha'i | 0 (0%) |
| Jainism | 1 (<0.1%) |
| Shinto | 0 (0%) |
| Taoism | 0 (<0.1%) |
| Confucianism | 1 (<0.1%) |
| Primal, Animist, or Folk religion | 1 (<0.1%) |
| Spiritism | 0 (0%) |
| Umbanda, Candomble, and other African-derived religions | 0 (0%) |
| Chinese folk/traditional religion | 0 (0%) |
| Some other religion | 0 (0%) |
| No religion/Atheist/Agnostic | 2 (<0.1%) |
| (Missing) | 8 (0.1%) |
| **Race/Ethnicity** |  |
| Bali | 69 (1.0%) |
| Banjar/Melayu Banjar | 320 (4.6%) |
| Batak | 165 (2.4%) |
| Betawi | 251 (3.6%) |
| Bugis | 243 (3.5%) |
| Jawa | 2,846 (41%) |
| Madura | 262 (3.7%) |
| Makasar | 91 (1.3%) |
| Minangkabau | 273 (3.9%) |
| Other | 1,262 (18%) |
| Sunda/Parahyangan | 1,172 (17%) |
| (Missing) | 38 (0.5%) |
| ^1^n (%) | |

Table S8b. Regression of Community Participation on Childhood Predictors for Indonesia

|  | | Secular Community Participation | | | |  | Religious Service Attendance | | | |
| --- | --- | --- | --- | --- | --- | --- | --- | --- | --- | --- |
| Variable | Category | Risk-Ratio | RR 95% CI | log(RR) SE | Global  p-value |  | Risk-Ratio | RR 95% CI | log(RR) SE | Global  p-value |
| Relationship with mother | (Ref: Very bad/somewhat bad) |  |  |  | 0.587 |  |  |  |  | 0.893 |
|  | Very good/somewhat good | 1.14 | (0.70,1.86) | 0.25 |  |  | 0.99 | (0.83,1.19) | 0.09 |  |
| Relationship with father | (Ref: Very bad/somewhat bad) |  |  |  | 0.880 |  |  |  |  | 0.727 |
|  | Very good/somewhat good | 1.02 | (0.74,1.41) | 0.17 |  |  | 1.02 | (0.92,1.13) | 0.05 |  |
| Parent marital status | (Ref: Parents married) |  |  |  | 0.825 |  |  |  |  | 0.408 |
|  | Divorced | 1.05 | (0.89,1.24) | 0.08 |  |  | 0.96 | (0.88,1.04) | 0.04 |  |
|  | Parents were never married | 1.14 | (0.68,1.90) | 0.26 |  |  | 0.82 | (0.62,1.09) | 0.14 |  |
|  | One or both parents had died | 0.98 | (0.82,1.18) | 0.09 |  |  | 1.00 | (0.94,1.06) | 0.03 |  |
| Subjective financial status of family growing up | (Ref: Got by) |  |  |  | 0.010 |  |  |  |  | 0.925 |
|  | Lived comfortably | 1.15 | (1.05,1.26) | 0.05 |  |  | 1.00 | (0.96,1.04) | 0.02 |  |
|  | Found it difficult | 1.03 | (0.83,1.27) | 0.11 |  |  | 0.97 | (0.89,1.07) | 0.05 |  |
|  | Found it very difficult | 0.76 | (0.51,1.15) | 0.21 |  |  | 1.02 | (0.91,1.15) | 0.06 |  |
| Experienced Abuse | (Ref: No) |  |  |  | 0.096 |  |  |  |  | 0.084 |
|  | Yes | 1.14 | (0.98,1.33) | 0.08 |  |  | 0.94 | (0.87,1.01) | 0.04 |  |
| Felt like an outsider in the family | (Ref: No) |  |  |  | 0.492 |  |  |  |  | 0.069 |
|  | Yes | 0.92 | (0.74,1.16) | 0.11 |  |  | 0.91 | (0.83,1.01) | 0.05 |  |
| Self-rated health growing up | (Ref: Good) |  |  |  | <.001 |  |  |  |  | 0.064 |
|  | Excellent | 1.38 | (1.23,1.56) | 0.06 |  |  | 1.05 | (1.00,1.11) | 0.03 |  |
|  | Very good | 1.20 | (1.07,1.35) | 0.06 |  |  | 1.06 | (1.02,1.11) | 0.02 |  |
|  | Fair | 1.05 | (0.92,1.20) | 0.07 |  |  | 1.05 | (1.00,1.11) | 0.03 |  |
|  | Poor | 1.82 | (1.20,2.76) | 0.21 |  |  | 1.03 | (0.81,1.30) | 0.12 |  |
| Immigration status | (Ref: Born in this country) |  |  |  | 0.395 |  |  |  |  | 0.294 |
|  | Born in another country | 0.80 | (0.47,1.34) | 0.27 |  |  | 0.86 | (0.64,1.15) | 0.15 |  |
| Religious service attendance (age 12) | (Ref: Never) |  |  |  | 0.046 |  |  |  |  | <.001 |
|  | At least 1/week | 1.23 | (0.96,1.57) | 0.12 |  |  | 1.30 | (1.15,1.46) | 0.06 |  |
|  | 1-3/month | 1.15 | (0.88,1.51) | 0.14 |  |  | 1.09 | (0.95,1.24) | 0.07 |  |
|  | < 1/month | 0.96 | (0.68,1.36) | 0.18 |  |  | 0.92 | (0.78,1.09) | 0.09 |  |
| Year of birth | (Ref: 1998-2005; current age: 18-24) |  |  |  | <.001 |  |  |  |  | <.001 |
|  | 1993-1998; age 25-29 | 0.86 | (0.74,0.99) | 0.07 |  |  | 1.00 | (0.95,1.05) | 0.03 |  |
|  | 1983-1993; age 30-39 | 0.84 | (0.74,0.95) | 0.06 |  |  | 1.00 | (0.95,1.05) | 0.03 |  |
|  | 1973-1983; age 40-49 | 0.84 | (0.74,0.94) | 0.06 |  |  | 1.05 | (1.00,1.11) | 0.03 |  |
|  | 1963-1973; age 50-59 | 0.82 | (0.70,0.96) | 0.08 |  |  | 1.06 | (1.00,1.12) | 0.03 |  |
|  | 1953-1963; age 60-69 | 0.76 | (0.60,0.96) | 0.12 |  |  | 1.11 | (1.02,1.20) | 0.04 |  |
|  | 1943-1953; age 70-79 | 0.46 | (0.23,0.92) | 0.36 |  |  | 0.93 | (0.73,1.18) | 0.12 |  |
|  | 1943 or earlier; age 80+ | 0.00 | (0.00,0.00) | 0.41 |  |  | 1.43 | (1.23,1.67) | 0.08 |  |
| Gender | (Ref: Male) |  |  |  | <.001 |  |  |  |  | <.001 |
|  | Female | 0.80 | (0.74,0.87) | 0.04 |  |  | 0.88 | (0.85,0.91) | 0.02 |  |
|  | Other | 1.54 | (0.73,3.24) | 0.38 |  |  | 1.09 | (0.93,1.28) | 0.08 |  |
| Religious affiliation | (Ref: Islam) |  |  |  | 0.785 |  |  |  |  | 0.569 |
|  | Christianity | 1.02 | (0.83,1.27) | 0.11 |  |  | 1.02 | (0.96,1.09) | 0.03 |  |
|  | Collapsed affiliations with prevalence<3% | 0.88 | (0.60,1.29) | 0.20 |  |  | 0.93 | (0.78,1.11) | 0.09 |  |
| Race/ethnicity | (Ref: Plurality group) |  |  |  | 0.594 |  |  |  |  | 0.629 |
|  | Non-plurality groups | 0.98 | (0.89,1.08) | 0.05 |  |  | 1.00 | (0.96,1.05) | 0.02 |  |

Table S8c. Sensitivity of the regression estimates to unmeasured confounding for Indonesia

|  | | Secular Community Participation | |  | Religious Service Attendance | |
| --- | --- | --- | --- | --- | --- | --- |
| Variable | Category | E-value for Estimate | E-value for 95% CI |  | E-value for Estimate | E-value for 95% CI |
| Relationship with mother | (Ref: Very bad/somewhat bad) |  |  |  |  |  |
|  | Very good/somewhat good | 1.55 | 1.00 |  | 1.11 | 1.00 |
| Relationship with father | (Ref: Very bad/somewhat bad) |  |  |  |  |  |
|  | Very good/somewhat good | 1.16 | 1.00 |  | 1.15 | 1.00 |
| Parent marital status | (Ref: Parents married) |  |  |  |  |  |
|  | Divorced | 1.28 | 1.00 |  | 1.27 | 1.00 |
|  | Parents were never married | 1.54 | 1.00 |  | 1.73 | 1.00 |
|  | One or both parents had died | 1.15 | 1.00 |  | 1.07 | 1.00 |
| Subjective financial status of family growing up | (Ref: Got by) |  |  |  |  |  |
|  | Lived comfortably | 1.56 | 1.28 |  | 1.02 | 1.00 |
|  | Found it difficult | 1.19 | 1.00 |  | 1.20 | 1.00 |
|  | Found it very difficult | 1.95 | 1.00 |  | 1.16 | 1.00 |
| Experienced Abuse | (Ref: No) |  |  |  |  |  |
|  | Yes | 1.54 | 1.00 |  | 1.33 | 1.00 |
| Felt like an outsider in the family | (Ref: No) |  |  |  |  |  |
|  | Yes | 1.38 | 1.00 |  | 1.41 | 1.00 |
| Self-rated health growing up | (Ref: Good) |  |  |  |  |  |
|  | Excellent | 2.11 | 1.75 |  | 1.29 | 1.03 |
|  | Very good | 1.69 | 1.35 |  | 1.31 | 1.14 |
|  | Fair | 1.27 | 1.00 |  | 1.29 | 1.03 |
|  | Poor | 3.04 | 1.69 |  | 1.19 | 1.00 |
| Immigration status | (Ref: Born in this country) |  |  |  |  |  |
|  | Born in another country | 1.82 | 1.00 |  | 1.61 | 1.00 |
| Religious service attendance (age 12) | (Ref: Never) |  |  |  |  |  |
|  | At least 1/week | 1.76 | 1.00 |  | 1.92 | 1.57 |
|  | 1-3/month | 1.58 | 1.00 |  | 1.40 | 1.00 |
|  | < 1/month | 1.24 | 1.00 |  | 1.40 | 1.00 |
| Year of birth | (Ref: 1998-2005; current age: 18-24) |  |  |  |  |  |
|  | 1993-1998; age 25-29 | 1.61 | 1.13 |  | 1.04 | 1.00 |
|  | 1983-1993; age 30-39 | 1.67 | 1.28 |  | 1.03 | 1.00 |
|  | 1973-1983; age 40-49 | 1.68 | 1.31 |  | 1.28 | 1.00 |
|  | 1963-1973; age 50-59 | 1.73 | 1.24 |  | 1.31 | 1.07 |
|  | 1953-1963; age 60-69 | 1.96 | 1.24 |  | 1.45 | 1.15 |
|  | 1943-1953; age 70-79 | 3.81 | 1.39 |  | 1.37 | 1.00 |
|  | 1943 or earlier; age 80+ | 953911.58 | 428998.53 |  | 2.21 | 1.75 |
| Gender | (Ref: Male) |  |  |  |  |  |
|  | Female | 1.80 | 1.55 |  | 1.53 | 1.41 |
|  | Other | 2.44 | 1.00 |  | 1.41 | 1.00 |
| Religious affiliation | (Ref: Islam) |  |  |  |  |  |
|  | Christianity | 1.18 | 1.00 |  | 1.17 | 1.00 |
|  | Collapsed affiliations with prevalence<3% | 1.53 | 1.00 |  | 1.37 | 1.00 |
| Race/ethnicity | (Ref: Plurality group) |  |  |  |  |  |
|  | Non-plurality groups | 1.17 | 1.00 |  | 1.07 | 1.00 |

Table S9a. Nationally representative descriptive statistics for Israel

| **Characteristic** | **N = 3,669**^1^ |
| --- | --- |
| **Relationship with mother** |  |
| Very good | 2,686 (73%) |
| Somewhat good | 793 (22%) |
| Somewhat bad | 110 (3.0%) |
| Very bad | 18 (0.5%) |
| Does not apply | 45 (1.2%) |
| (Missing) | 17 (0.5%) |
| **Relationship with father** |  |
| Very good | 2,290 (62%) |
| Somewhat good | 912 (25%) |
| Somewhat bad | 234 (6.4%) |
| Very bad | 37 (1.0%) |
| Does not apply | 171 (4.7%) |
| (Missing) | 25 (0.7%) |
| **Parent marital status** |  |
| Parents married | 3,172 (86%) |
| Divorced | 284 (7.8%) |
| Parents were never married | 36 (1.0%) |
| One or both parents had died | 130 (3.5%) |
| (Missing) | 47 (1.3%) |
| **Subjective financial status of family growing up** |  |
| Lived comfortably | 923 (25%) |
| Got by | 1,822 (50%) |
| Found it difficult | 667 (18%) |
| Found it very difficult | 239 (6.5%) |
| (Missing) | 17 (0.5%) |
| **Experienced Abuse** |  |
| Yes | 0 (0%) |
| No | 0 (0%) |
| (Missing) | 3,669 (100%) |
| **Felt like an outsider in the family** |  |
| Yes | 371 (10%) |
| No | 3,228 (88%) |
| (Missing) | 70 (1.9%) |
| **Self-rated health growing up** |  |
| Excellent | 1,785 (49%) |
| Very good | 1,284 (35%) |
| Good | 480 (13%) |
| Fair | 105 (2.9%) |
| Poor | 6 (0.2%) |
| (Missing) | 8 (0.2%) |
| **Immigration status** |  |
| Born in this country | 2,796 (76%) |
| Born in another country | 868 (24%) |
| (Missing) | 5 (0.1%) |
| **Religious service attendance (age 12)** |  |
| At least 1/week | 867 (24%) |
| 1-3/month | 435 (12%) |
| <1/month | 810 (22%) |
| Never | 1,539 (42%) |
| (Missing) | 17 (0.5%) |
| **Year of birth** |  |
| 1998-2005; age 18-24 | 553 (15%) |
| 1993-1998; age 25-29 | 407 (11%) |
| 1983-1993; age 30-39 | 666 (18%) |
| 1973-1983; age 40-49 | 616 (17%) |
| 1963-1973; age 50-59 | 542 (15%) |
| 1953-1963; age 60-69 | 469 (13%) |
| 1943-1953; age 70-79 | 336 (9.2%) |
| 1943 or earlier; age 80+ | 79 (2.2%) |
| (Missing) | 0 (0%) |
| **Gender** |  |
| Male | 1,791 (49%) |
| Female | 1,872 (51%) |
| Other | 0 (<0.1%) |
| (Missing) | 6 (0.2%) |
| **Religious affiliation** |  |
| Christianity | 60 (1.6%) |
| Islam | 647 (18%) |
| Hinduism | 0 (0%) |
| Buddhism | 0 (0%) |
| Judaism | 2,873 (78%) |
| Sikhism | 1 (<0.1%) |
| Baha'i | 1 (<0.1%) |
| Jainism | 0 (0%) |
| Shinto | 0 (0%) |
| Taoism | 0 (0%) |
| Confucianism | 0 (0%) |
| Primal, Animist, or Folk religion | 3 (<0.1%) |
| Spiritism | 0 (0%) |
| Umbanda, Candomble, and other African-derived religions | 0 (0%) |
| Chinese folk/traditional religion | 0 (0%) |
| Some other religion | 5 (0.1%) |
| No religion/Atheist/Agnostic | 69 (1.9%) |
| (Missing) | 10 (0.3%) |
| **Race/Ethnicity** |  |
| Arab | 674 (18%) |
| Jewish | 2,926 (80%) |
| Other | 39 (1.1%) |
| (Missing) | 30 (0.8%) |
| ^1^n (%) | |

Table S9b. Regression of Community Participation on Childhood Predictors for Israel

|  | | Secular Community Participation | | | |  | Religious Service Attendance | | | |
| --- | --- | --- | --- | --- | --- | --- | --- | --- | --- | --- |
| Variable | Category | Risk-Ratio | RR 95% CI | log(RR) SE | Global  p-value |  | Risk-Ratio | RR 95% CI | log(RR) SE | Global p-value |
| Relationship with mother | (Ref: Very bad/somewhat bad) |  |  |  | 0.513 |  |  |  |  | 0.455 |
|  | Very good/somewhat good | 1.16 | (0.75,1.79) | 0.22 |  |  | 0.90 | (0.68,1.20) | 0.14 |  |
| Relationship with father | (Ref: Very bad/somewhat bad) |  |  |  | 0.808 |  |  |  |  | 0.255 |
|  | Very good/somewhat good | 1.04 | (0.74,1.46) | 0.17 |  |  | 0.88 | (0.71,1.09) | 0.11 |  |
| Parent marital status | (Ref: Parents married) |  |  |  | 0.181 |  |  |  |  | <.001 |
|  | Divorced | 1.01 | (0.78,1.30) | 0.13 |  |  | 0.65 | (0.49,0.87) | 0.15 |  |
|  | Parents were never married | 1.61 | (0.99,2.63) | 0.25 |  |  | 0.38 | (0.11,1.27) | 0.62 |  |
|  | One or both parents had died | 0.83 | (0.52,1.35) | 0.25 |  |  | 0.48 | (0.29,0.81) | 0.26 |  |
| Subjective financial status of family growing up | (Ref: Got by) |  |  |  | 0.087 |  |  |  |  | 0.091 |
|  | Lived comfortably | 1.09 | (0.88,1.36) | 0.11 |  |  | 0.96 | (0.84,1.10) | 0.07 |  |
|  | Found it difficult | 0.76 | (0.60,0.96) | 0.12 |  |  | 1.19 | (1.03,1.37) | 0.07 |  |
|  | Found it very difficult | 0.85 | (0.62,1.17) | 0.16 |  |  | 1.02 | (0.80,1.30) | 0.13 |  |
| Felt like an outsider in the family | (Ref: No) |  |  |  | 0.277 |  |  |  |  | 0.133 |
|  | Yes | 1.14 | (0.89,1.46) | 0.13 |  |  | 0.87 | (0.72,1.06) | 0.10 |  |
| Self-rated health growing up | (Ref: Good) |  |  |  | 0.006 |  |  |  |  | <.001 |
|  | Excellent | 0.81 | (0.62,1.06) | 0.14 |  |  | 1.41 | (1.16,1.71) | 0.10 |  |
|  | Very good | 0.72 | (0.55,0.94) | 0.14 |  |  | 1.32 | (1.11,1.58) | 0.09 |  |
|  | Fair | 1.18 | (0.75,1.86) | 0.23 |  |  | 0.73 | (0.44,1.19) | 0.25 |  |
|  | Poor | 1.61 | (0.84,3.12) | 0.34 |  |  | 4.81 | (2.35,9.87) | 0.37 |  |
| Immigration status | (Ref: Born in this country) |  |  |  | 0.332 |  |  |  |  | 0.101 |
|  | Born in another country | 1.12 | (0.89,1.40) | 0.11 |  |  | 0.86 | (0.73,1.03) | 0.09 |  |
| Religious service attendance (age 12) | (Ref: Never) |  |  |  | 0.055 |  |  |  |  | <.001 |
|  | At least 1/week | 0.90 | (0.72,1.12) | 0.11 |  |  | 4.63 | (3.79,5.67) | 0.10 |  |
|  | 1-3/month | 0.69 | (0.52,0.91) | 0.15 |  |  | 3.79 | (3.02,4.75) | 0.12 |  |
|  | < 1/month | 0.78 | (0.62,0.99) | 0.12 |  |  | 1.89 | (1.48,2.41) | 0.12 |  |
| Year of birth | (Ref: 1998-2005; current age: 18-24) |  |  |  | <.001 |  |  |  |  | 0.055 |
|  | 1993-1998; age 25-29 | 0.86 | (0.66,1.12) | 0.13 |  |  | 0.88 | (0.77,1.01) | 0.07 |  |
|  | 1983-1993; age 30-39 | 0.68 | (0.54,0.86) | 0.12 |  |  | 0.80 | (0.69,0.93) | 0.08 |  |
|  | 1973-1983; age 40-49 | 0.70 | (0.53,0.92) | 0.14 |  |  | 0.79 | (0.66,0.95) | 0.10 |  |
|  | 1963-1973; age 50-59 | 0.75 | (0.55,1.02) | 0.16 |  |  | 0.90 | (0.77,1.07) | 0.08 |  |
|  | 1953-1963; age 60-69 | 0.55 | (0.42,0.72) | 0.14 |  |  | 0.85 | (0.71,1.00) | 0.09 |  |
|  | 1943-1953; age 70-79 | 0.87 | (0.60,1.27) | 0.19 |  |  | 0.81 | (0.65,1.00) | 0.11 |  |
|  | 1943 or earlier; age 80+ | 0.74 | (0.42,1.30) | 0.29 |  |  | 0.60 | (0.36,0.98) | 0.25 |  |
| Gender | (Ref: Male) |  |  |  | <.001 |  |  |  |  | <.001 |
|  | Female | 0.82 | (0.70,0.96) | 0.08 |  |  | 0.92 | (0.82,1.04) | 0.06 |  |
|  | Other | 0.00 | (0.00,0.00) | 1.03 |  |  | 0.00 | (0.00,468.33) | 4.70 |  |
| Religious affiliation | (Ref: Judaism) |  |  |  | 0.423 |  |  |  |  | 0.203 |
|  | Islam | 0.88 | (0.30,2.59) | 0.55 |  |  | 1.37 | (0.53,3.53) | 0.48 |  |
|  | Collapsed affiliations with prevalence<3% | 0.73 | (0.45,1.19) | 0.25 |  |  | 0.79 | (0.41,1.51) | 0.33 |  |
| Race/ethnicity | (Ref: Plurality group) |  |  |  | 0.441 |  |  |  |  | 0.558 |
|  | Non-plurality groups | 1.44 | (0.54,3.87) | 0.50 |  |  | 0.76 | (0.30,1.95) | 0.48 |  |

Table S9c. Sensitivity of the regression estimates to unmeasured confounding for Israel

|  | | Secular Community Participation | |  | Religious Service Attendance | |
| --- | --- | --- | --- | --- | --- | --- |
| Variable | Category | E-value for Estimate | E-value for 95% CI |  | E-value for Estimate | E-value for 95% CI |
| Relationship with mother | (Ref: Very bad/somewhat bad) |  |  |  |  |  |
|  | Very good/somewhat good | 1.58 | 1.00 |  | 1.46 | 1.00 |
| Relationship with father | (Ref: Very bad/somewhat bad) |  |  |  |  |  |
|  | Very good/somewhat good | 1.23 | 1.00 |  | 1.52 | 1.00 |
| Parent marital status | (Ref: Parents married) |  |  |  |  |  |
|  | Divorced | 1.09 | 1.00 |  | 2.43 | 1.57 |
|  | Parents were never married | 2.61 | 1.00 |  | 4.75 | 1.00 |
|  | One or both parents had died | 1.69 | 1.00 |  | 3.55 | 1.79 |
| Subjective financial status of family growing up | (Ref: Got by) |  |  |  |  |  |
|  | Lived comfortably | 1.41 | 1.00 |  | 1.24 | 1.00 |
|  | Found it difficult | 1.98 | 1.25 |  | 1.67 | 1.22 |
|  | Found it very difficult | 1.62 | 1.00 |  | 1.16 | 1.00 |
| Felt like an outsider in the family | (Ref: No) |  |  |  |  |  |
|  | Yes | 1.55 | 1.00 |  | 1.57 | 1.00 |
| Self-rated health growing up | (Ref: Good) |  |  |  |  |  |
|  | Excellent | 1.78 | 1.00 |  | 2.16 | 1.59 |
|  | Very good | 2.13 | 1.34 |  | 1.98 | 1.45 |
|  | Fair | 1.63 | 1.00 |  | 2.10 | 1.00 |
|  | Poor | 2.61 | 1.00 |  | 9.10 | 4.13 |
| Immigration status | (Ref: Born in this country) |  |  |  |  |  |
|  | Born in another country | 1.48 | 1.00 |  | 1.58 | 1.00 |
| Religious service attendance (age 12) | (Ref: Never) |  |  |  |  |  |
|  | At least 1/week | 1.47 | 1.00 |  | 8.74 | 7.04 |
|  | 1-3/month | 2.27 | 1.42 |  | 7.03 | 5.48 |
|  | < 1/month | 1.88 | 1.09 |  | 3.18 | 2.32 |
| Year of birth | (Ref: 1998-2005; current age: 18-24) |  |  |  |  |  |
|  | 1993-1998; age 25-29 | 1.59 | 1.00 |  | 1.52 | 1.00 |
|  | 1983-1993; age 30-39 | 2.29 | 1.59 |  | 1.82 | 1.37 |
|  | 1973-1983; age 40-49 | 2.22 | 1.41 |  | 1.84 | 1.27 |
|  | 1963-1973; age 50-59 | 2.02 | 1.00 |  | 1.45 | 1.00 |
|  | 1953-1963; age 60-69 | 3.06 | 2.14 |  | 1.64 | 1.00 |
|  | 1943-1953; age 70-79 | 1.55 | 1.00 |  | 1.79 | 1.00 |
|  | 1943 or earlier; age 80+ | 2.05 | 1.00 |  | 2.74 | 1.16 |
| Gender | (Ref: Male) |  |  |  |  |  |
|  | Female | 1.73 | 1.25 |  | 1.39 | 1.00 |
|  | Other | 24806.99 | 3313.28 |  | 1528.40 | 1.00 |
| Religious affiliation | (Ref: Judaism) |  |  |  |  |  |
|  | Islam | 1.52 | 1.00 |  | 2.08 | 1.00 |
|  | Collapsed affiliations with prevalence<3% | 2.09 | 1.00 |  | 1.86 | 1.00 |
| Race/ethnicity | (Ref: Plurality group) |  |  |  |  |  |
|  | Non-plurality groups | 2.25 | 1.00 |  | 1.95 | 1.00 |

Table S10a. Nationally representative descriptive statistics for Japan

| **Characteristic** | **N = 20,543**^1^ |
| --- | --- |
| **Relationship with mother** |  |
| Very good | 5,630 (27%) |
| Somewhat good | 9,461 (46%) |
| Somewhat bad | 2,750 (13%) |
| Very bad | 799 (3.9%) |
| Does not apply | 1,838 (8.9%) |
| (Missing) | 66 (0.3%) |
| **Relationship with father** |  |
| Very good | 4,156 (20%) |
| Somewhat good | 9,081 (44%) |
| Somewhat bad | 3,446 (17%) |
| Very bad | 1,223 (6.0%) |
| Does not apply | 2,580 (13%) |
| (Missing) | 57 (0.3%) |
| **Parent marital status** |  |
| Parents married | 17,713 (86%) |
| Divorced | 1,127 (5.5%) |
| Parents were never married | 591 (2.9%) |
| One or both parents had died | 754 (3.7%) |
| (Missing) | 359 (1.7%) |
| **Subjective financial status of family growing up** |  |
| Lived comfortably | 8,320 (41%) |
| Got by | 8,799 (43%) |
| Found it difficult | 2,398 (12%) |
| Found it very difficult | 973 (4.7%) |
| (Missing) | 52 (0.3%) |
| **Experienced Abuse** |  |
| Yes | 1,482 (7.2%) |
| No | 18,964 (92%) |
| (Missing) | 96 (0.5%) |
| **Felt like an outsider in the family** |  |
| Yes | 1,963 (9.6%) |
| No | 17,136 (83%) |
| (Missing) | 1,444 (7.0%) |
| **Self-rated health growing up** |  |
| Excellent | 2,711 (13%) |
| Very good | 7,106 (35%) |
| Good | 6,689 (33%) |
| Fair | 3,199 (16%) |
| Poor | 758 (3.7%) |
| (Missing) | 80 (0.4%) |
| **Immigration status** |  |
| Born in this country | 19,548 (95%) |
| Born in another country | 158 (0.8%) |
| (Missing) | 837 (4.1%) |
| **Religious service attendance (age 12)** |  |
| At least 1/week | 398 (1.9%) |
| 1-3/month | 883 (4.3%) |
| <1/month | 5,023 (24%) |
| Never | 14,117 (69%) |
| (Missing) | 123 (0.6%) |
| **Year of birth** |  |
| 1998-2005; age 18-24 | 1,589 (7.7%) |
| 1993-1998; age 25-29 | 806 (3.9%) |
| 1983-1993; age 30-39 | 2,851 (14%) |
| 1973-1983; age 40-49 | 3,363 (16%) |
| 1963-1973; age 50-59 | 3,770 (18%) |
| 1953-1963; age 60-69 | 4,118 (20%) |
| 1943-1953; age 70-79 | 3,554 (17%) |
| 1943 or earlier; age 80+ | 493 (2.4%) |
| (Missing) | 0 (0%) |
| **Gender** |  |
| Male | 9,847 (48%) |
| Female | 10,602 (52%) |
| Other | 28 (0.1%) |
| (Missing) | 66 (0.3%) |
| **Religious affiliation** |  |
| Christianity | 343 (1.7%) |
| Islam | 7 (<0.1%) |
| Hinduism | 4 (<0.1%) |
| Buddhism | 6,536 (32%) |
| Judaism | 0 (0%) |
| Sikhism | 0 (0%) |
| Baha'i | 7 (<0.1%) |
| Jainism | 1 (<0.1%) |
| Shinto | 382 (1.9%) |
| Taoism | 14 (<0.1%) |
| Confucianism | 25 (0.1%) |
| Primal, Animist, or Folk religion | 13 (<0.1%) |
| Spiritism | 0 (0%) |
| Umbanda, Candomble, and other African-derived religions | 0 (0%) |
| Chinese folk/traditional religion | 0 (0%) |
| Some other religion | 46 (0.2%) |
| No religion/Atheist/Agnostic | 12,950 (63%) |
| (Missing) | 215 (1.0%) |
| ^1^n (%) | |

Table S10b. Regression of Community Participation on Childhood Predictors for Japan

|  | | Secular Community Participation | | | |  | Religious Service Attendance | | | |
| --- | --- | --- | --- | --- | --- | --- | --- | --- | --- | --- |
| Variable | Category | Risk-Ratio | RR 95% CI | log(RR) SE | Global  p-value |  | Risk-Ratio | RR 95% CI | log(RR) SE | Global  p-value |
| Relationship with mother | (Ref: Very bad/somewhat bad) |  |  |  | 0.235 |  |  |  |  | 0.125 |
|  | Very good/somewhat good | 0.91 | (0.77,1.07) | 0.08 |  |  | 1.23 | (0.94,1.62) | 0.14 |  |
| Relationship with father | (Ref: Very bad/somewhat bad) |  |  |  | 0.021 |  |  |  |  | <.001 |
|  | Very good/somewhat good | 1.18 | (1.03,1.36) | 0.07 |  |  | 1.49 | (1.18,1.88) | 0.12 |  |
| Parent marital status | (Ref: Parents married) |  |  |  | 0.065 |  |  |  |  | 0.437 |
|  | Divorced | 0.77 | (0.58,1.02) | 0.15 |  |  | 1.07 | (0.69,1.65) | 0.22 |  |
|  | Parents were never married | 1.09 | (0.83,1.44) | 0.14 |  |  | 1.26 | (0.89,1.80) | 0.18 |  |
|  | One or both parents had died | 1.25 | (0.95,1.64) | 0.14 |  |  | 1.27 | (0.80,2.03) | 0.24 |  |
| Subjective financial status of family growing up | (Ref: Got by) |  |  |  | 0.796 |  |  |  |  | 0.392 |
|  | Lived comfortably | 1.05 | (0.93,1.18) | 0.06 |  |  | 0.98 | (0.80,1.20) | 0.10 |  |
|  | Found it difficult | 1.07 | (0.91,1.27) | 0.09 |  |  | 1.20 | (0.92,1.56) | 0.13 |  |
|  | Found it very difficult | 1.02 | (0.77,1.36) | 0.15 |  |  | 0.88 | (0.58,1.33) | 0.21 |  |
| Experienced Abuse | (Ref: No) |  |  |  | <.001 |  |  |  |  | 0.737 |
|  | Yes | 1.43 | (1.18,1.72) | 0.10 |  |  | 0.96 | (0.72,1.28) | 0.15 |  |
| Felt like an outsider in the family | (Ref: No) |  |  |  | <.001 |  |  |  |  | <.001 |
|  | Yes | 1.36 | (1.13,1.63) | 0.09 |  |  | 2.22 | (1.74,2.85) | 0.13 |  |
| Self-rated health growing up | (Ref: Good) |  |  |  | <.001 |  |  |  |  | 0.209 |
|  | Excellent | 1.39 | (1.19,1.64) | 0.08 |  |  | 1.28 | (0.98,1.68) | 0.14 |  |
|  | Very good | 1.22 | (1.08,1.38) | 0.06 |  |  | 1.22 | (0.99,1.51) | 0.11 |  |
|  | Fair | 0.74 | (0.61,0.89) | 0.09 |  |  | 0.98 | (0.72,1.35) | 0.16 |  |
|  | Poor | 0.75 | (0.53,1.04) | 0.17 |  |  | 0.97 | (0.58,1.61) | 0.26 |  |
| Immigration status | (Ref: Born in this country) |  |  |  | 0.914 |  |  |  |  | 0.416 |
|  | Born in another country | 0.97 | (0.52,1.81) | 0.32 |  |  | 0.75 | (0.26,2.17) | 0.53 |  |
| Religious service attendance (age 12) | (Ref: Never) |  |  |  | <.001 |  |  |  |  | <.001 |
|  | At least 1/week | 2.78 | (2.16,3.56) | 0.13 |  |  | 18.05 | (13.57,23.99) | 0.15 |  |
|  | 1-3/month | 3.42 | (2.90,4.02) | 0.08 |  |  | 11.16 | (8.41,14.80) | 0.14 |  |
|  | < 1/month | 1.50 | (1.32,1.69) | 0.06 |  |  | 2.44 | (1.87,3.18) | 0.14 |  |
| Year of birth | (Ref: 1998-2005; current age: 18-24) |  |  |  | <.001 |  |  |  |  | 0.037 |
|  | 1993-1998; age 25-29 | 0.65 | (0.47,0.89) | 0.17 |  |  | 1.36 | (0.86,2.17) | 0.24 |  |
|  | 1983-1993; age 30-39 | 0.52 | (0.41,0.66) | 0.12 |  |  | 1.19 | (0.79,1.78) | 0.21 |  |
|  | 1973-1983; age 40-49 | 0.42 | (0.34,0.54) | 0.12 |  |  | 0.98 | (0.64,1.48) | 0.21 |  |
|  | 1963-1973; age 50-59 | 0.43 | (0.34,0.54) | 0.12 |  |  | 0.82 | (0.54,1.25) | 0.21 |  |
|  | 1953-1963; age 60-69 | 0.81 | (0.67,0.99) | 0.10 |  |  | 1.31 | (0.88,1.96) | 0.20 |  |
|  | 1943-1953; age 70-79 | 1.29 | (1.06,1.56) | 0.10 |  |  | 1.27 | (0.85,1.88) | 0.20 |  |
|  | 1943 or earlier; age 80+ | 1.27 | (0.96,1.68) | 0.14 |  |  | 1.29 | (0.75,2.24) | 0.28 |  |
| Gender | (Ref: Male) |  |  |  | 0.007 |  |  |  |  | 0.006 |
|  | Female | 1.16 | (1.05,1.28) | 0.05 |  |  | 0.76 | (0.64,0.90) | 0.09 |  |
|  | Other | 0.42 | (0.10,1.78) | 0.74 |  |  | 1.37 | (0.35,5.33) | 0.69 |  |
| Religious affiliation | (Ref: No religion/Atheist/Agnostic) |  |  |  | 0.731 |  |  |  |  | <.001 |
|  | Buddhism | 0.98 | (0.87,1.10) | 0.06 |  |  | 2.01 | (1.61,2.52) | 0.11 |  |
|  | Collapsed affiliations with prevalence<3% | 0.91 | (0.72,1.15) | 0.12 |  |  | 2.27 | (1.69,3.05) | 0.15 |  |

Table S10c. Sensitivity of the regression estimates to unmeasured confounding for Japan

|  | | Secular Community Participation | |  | Religious Service Attendance | |
| --- | --- | --- | --- | --- | --- | --- |
| Variable | Category | E-value for Estimate | E-value for 95% CI |  | E-value for Estimate | E-value for 95% CI |
| Relationship with mother | (Ref: Very bad/somewhat bad) |  |  |  |  |  |
|  | Very good/somewhat good | 1.44 | 1.00 |  | 1.77 | 1.00 |
| Relationship with father | (Ref: Very bad/somewhat bad) |  |  |  |  |  |
|  | Very good/somewhat good | 1.64 | 1.19 |  | 2.34 | 1.63 |
| Parent marital status | (Ref: Parents married) |  |  |  |  |  |
|  | Divorced | 1.94 | 1.00 |  | 1.33 | 1.00 |
|  | Parents were never married | 1.41 | 1.00 |  | 1.84 | 1.00 |
|  | One or both parents had died | 1.81 | 1.00 |  | 1.86 | 1.00 |
| Subjective financial status of family growing up | (Ref: Got by) |  |  |  |  |  |
|  | Lived comfortably | 1.28 | 1.00 |  | 1.15 | 1.00 |
|  | Found it difficult | 1.35 | 1.00 |  | 1.69 | 1.00 |
|  | Found it very difficult | 1.17 | 1.00 |  | 1.53 | 1.00 |
| Experienced Abuse | (Ref: No) |  |  |  |  |  |
|  | Yes | 2.21 | 1.65 |  | 1.26 | 1.00 |
| Felt like an outsider in the family | (Ref: No) |  |  |  |  |  |
|  | Yes | 2.06 | 1.53 |  | 3.87 | 2.87 |
| Self-rated health growing up | (Ref: Good) |  |  |  |  |  |
|  | Excellent | 2.14 | 1.66 |  | 1.88 | 1.00 |
|  | Very good | 1.73 | 1.36 |  | 1.74 | 1.00 |
|  | Fair | 2.06 | 1.51 |  | 1.15 | 1.00 |
|  | Poor | 2.02 | 1.00 |  | 1.22 | 1.00 |
| Immigration status | (Ref: Born in this country) |  |  |  |  |  |
|  | Born in another country | 1.20 | 1.00 |  | 2.02 | 1.00 |
| Religious service attendance (age 12) | (Ref: Never) |  |  |  |  |  |
|  | At least 1/week | 5.00 | 3.75 |  | 35.59 | 26.64 |
|  | 1-3/month | 6.29 | 5.25 |  | 21.80 | 16.30 |
|  | < 1/month | 2.36 | 1.98 |  | 4.31 | 3.14 |
| Year of birth | (Ref: 1998-2005; current age: 18-24) |  |  |  |  |  |
|  | 1993-1998; age 25-29 | 2.47 | 1.48 |  | 2.07 | 1.00 |
|  | 1983-1993; age 30-39 | 3.25 | 2.40 |  | 1.66 | 1.00 |
|  | 1973-1983; age 40-49 | 4.14 | 3.12 |  | 1.18 | 1.00 |
|  | 1963-1973; age 50-59 | 4.06 | 3.08 |  | 1.72 | 1.00 |
|  | 1953-1963; age 60-69 | 1.77 | 1.13 |  | 1.96 | 1.00 |
|  | 1943-1953; age 70-79 | 1.90 | 1.33 |  | 1.85 | 1.00 |
|  | 1943 or earlier; age 80+ | 1.86 | 1.00 |  | 1.91 | 1.00 |
| Gender | (Ref: Male) |  |  |  |  |  |
|  | Female | 1.59 | 1.27 |  | 1.95 | 1.45 |
|  | Other | 4.19 | 1.00 |  | 2.09 | 1.00 |
| Religious affiliation | (Ref: No religion/Atheist/Agnostic) |  |  |  |  |  |
|  | Buddhism | 1.17 | 1.00 |  | 3.45 | 2.61 |
|  | Collapsed affiliations with prevalence<3% | 1.42 | 1.00 |  | 3.97 | 2.76 |

Table S11a. Nationally representative descriptive statistics for Kenya

| **Characteristic** | **N = 11,389**^1^ |
| --- | --- |
| **Relationship with mother** |  |
| Very good | 9,418 (83%) |
| Somewhat good | 1,435 (13%) |
| Somewhat bad | 130 (1.1%) |
| Very bad | 100 (0.9%) |
| Does not apply | 240 (2.1%) |
| (Missing) | 66 (0.6%) |
| **Relationship with father** |  |
| Very good | 7,958 (70%) |
| Somewhat good | 1,896 (17%) |
| Somewhat bad | 216 (1.9%) |
| Very bad | 220 (1.9%) |
| Does not apply | 967 (8.5%) |
| (Missing) | 132 (1.2%) |
| **Parent marital status** |  |
| Parents married | 9,238 (81%) |
| Divorced | 697 (6.1%) |
| Parents were never married | 681 (6.0%) |
| One or both parents had died | 471 (4.1%) |
| (Missing) | 301 (2.6%) |
| **Subjective financial status of family growing up** |  |
| Lived comfortably | 3,026 (27%) |
| Got by | 3,279 (29%) |
| Found it difficult | 4,071 (36%) |
| Found it very difficult | 994 (8.7%) |
| (Missing) | 19 (0.2%) |
| **Experienced Abuse** |  |
| Yes | 1,300 (11%) |
| No | 10,039 (88%) |
| (Missing) | 49 (0.4%) |
| **Felt like an outsider in the family** |  |
| Yes | 1,223 (11%) |
| No | 10,114 (89%) |
| (Missing) | 52 (0.5%) |
| **Self-rated health growing up** |  |
| Excellent | 4,449 (39%) |
| Very good | 2,598 (23%) |
| Good | 2,582 (23%) |
| Fair | 1,384 (12%) |
| Poor | 349 (3.1%) |
| (Missing) | 26 (0.2%) |
| **Immigration status** |  |
| Born in this country | 11,270 (99%) |
| Born in another country | 117 (1.0%) |
| (Missing) | 2 (<0.1%) |
| **Religious service attendance (age 12)** |  |
| At least 1/week | 9,189 (81%) |
| 1-3/month | 1,687 (15%) |
| <1/month | 236 (2.1%) |
| Never | 198 (1.7%) |
| (Missing) | 79 (0.7%) |
| **Year of birth** |  |
| 1998-2005; age 18-24 | 2,868 (25%) |
| 1993-1998; age 25-29 | 2,035 (18%) |
| 1983-1993; age 30-39 | 2,564 (23%) |
| 1973-1983; age 40-49 | 1,708 (15%) |
| 1963-1973; age 50-59 | 1,072 (9.4%) |
| 1953-1963; age 60-69 | 710 (6.2%) |
| 1943-1953; age 70-79 | 360 (3.2%) |
| 1943 or earlier; age 80+ | 67 (0.6%) |
| (Missing) | 5 (<0.1%) |
| **Gender** |  |
| Male | 5,567 (49%) |
| Female | 5,813 (51%) |
| Other | 2 (<0.1%) |
| (Missing) | 7 (<0.1%) |
| **Religious affiliation** |  |
| Christianity | 10,369 (91%) |
| Islam | 916 (8.0%) |
| Hinduism | 0 (0%) |
| Buddhism | 5 (<0.1%) |
| Judaism | 6 (<0.1%) |
| Sikhism | 0 (<0.1%) |
| Baha'i | 3 (<0.1%) |
| Jainism | 1 (<0.1%) |
| Shinto | 0 (0%) |
| Taoism | 0 (0%) |
| Confucianism | 0 (0%) |
| Primal, Animist, or Folk religion | 13 (0.1%) |
| Spiritism | 0 (0%) |
| Umbanda, Candomble, and other African-derived religions | 0 (0%) |
| Chinese folk/traditional religion | 0 (0%) |
| Some other religion | 0 (<0.1%) |
| No religion/Atheist/Agnostic | 67 (0.6%) |
| (Missing) | 9 (<0.1%) |
| **Race/Ethnicity** |  |
| Embu | 197 (1.7%) |
| Kalenjin | 1,377 (12%) |
| Kamba | 1,299 (11%) |
| Kenyan Somali/Somali | 396 (3.5%) |
| Kikuyu | 2,119 (19%) |
| Kisii | 789 (6.9%) |
| Luhya | 1,943 (17%) |
| Luo | 1,120 (9.8%) |
| Maasai | 237 (2.1%) |
| Meru | 630 (5.5%) |
| Miji Kenda tribes | 708 (6.2%) |
| Other | 548 (4.8%) |
| (Missing) | 27 (0.2%) |
| ^1^n (%) | |

Table S11b. Regression of Community Participation on Childhood Predictors for Kenya

|  | | Secular Community Participation | | | |  | Religious Service Attendance | | | |
| --- | --- | --- | --- | --- | --- | --- | --- | --- | --- | --- |
| Variable | Category | Risk-Ratio | RR 95% CI | log(RR) SE | Global  p-value |  | Risk-Ratio | RR 95% CI | log(RR) SE | Global  p-value |
| Relationship with mother | (Ref: Very bad/somewhat bad) |  |  |  | 0.535 |  |  |  |  | 0.314 |
|  | Very good/somewhat good | 1.07 | (0.85,1.35) | 0.12 |  |  | 1.04 | (0.96,1.14) | 0.04 |  |
| Relationship with father | (Ref: Very bad/somewhat bad) |  |  |  | 0.159 |  |  |  |  | 0.429 |
|  | Very good/somewhat good | 1.14 | (0.95,1.36) | 0.09 |  |  | 1.02 | (0.97,1.09) | 0.03 |  |
| Parent marital status | (Ref: Parents married) |  |  |  | 0.120 |  |  |  |  | 0.203 |
|  | Divorced | 0.96 | (0.84,1.10) | 0.07 |  |  | 0.97 | (0.91,1.02) | 0.03 |  |
|  | Parents were never married | 0.98 | (0.84,1.16) | 0.08 |  |  | 0.95 | (0.89,1.01) | 0.03 |  |
|  | One or both parents had died | 0.78 | (0.64,0.96) | 0.10 |  |  | 0.97 | (0.91,1.04) | 0.03 |  |
| Subjective financial status of family growing up | (Ref: Got by) |  |  |  | 0.559 |  |  |  |  | 0.951 |
|  | Lived comfortably | 1.02 | (0.93,1.11) | 0.04 |  |  | 1.01 | (0.97,1.04) | 0.02 |  |
|  | Found it difficult | 0.95 | (0.87,1.04) | 0.05 |  |  | 1.00 | (0.97,1.04) | 0.02 |  |
|  | Found it very difficult | 0.98 | (0.84,1.13) | 0.08 |  |  | 1.01 | (0.96,1.06) | 0.02 |  |
| Experienced Abuse | (Ref: No) |  |  |  | <.001 |  |  |  |  | 0.018 |
|  | Yes | 1.30 | (1.17,1.44) | 0.05 |  |  | 0.95 | (0.91,0.99) | 0.02 |  |
| Felt like an outsider in the family | (Ref: No) |  |  |  | 0.554 |  |  |  |  | 0.020 |
|  | Yes | 0.96 | (0.84,1.10) | 0.07 |  |  | 0.95 | (0.91,0.99) | 0.02 |  |
| Self-rated health growing up | (Ref: Good) |  |  |  | 0.164 |  |  |  |  | 0.067 |
|  | Excellent | 1.09 | (0.99,1.21) | 0.05 |  |  | 1.01 | (0.98,1.05) | 0.02 |  |
|  | Very good | 0.98 | (0.88,1.09) | 0.06 |  |  | 0.97 | (0.93,1.01) | 0.02 |  |
|  | Fair | 1.01 | (0.90,1.15) | 0.06 |  |  | 0.98 | (0.95,1.02) | 0.02 |  |
|  | Poor | 1.06 | (0.84,1.33) | 0.12 |  |  | 1.00 | (0.92,1.09) | 0.04 |  |
| Immigration status | (Ref: Born in this country) |  |  |  | 0.777 |  |  |  |  | 0.558 |
|  | Born in another country | 0.95 | (0.68,1.34) | 0.17 |  |  | 1.03 | (0.93,1.15) | 0.05 |  |
| Religious service attendance (age 12) | (Ref: Never) |  |  |  | 0.007 |  |  |  |  | <.001 |
|  | At least 1/week | 1.07 | (0.78,1.48) | 0.16 |  |  | 1.15 | (1.02,1.30) | 0.06 |  |
|  | 1-3/month | 0.89 | (0.65,1.23) | 0.16 |  |  | 1.01 | (0.89,1.15) | 0.06 |  |
|  | < 1/month | 1.07 | (0.73,1.55) | 0.19 |  |  | 1.11 | (0.95,1.30) | 0.08 |  |
| Year of birth | (Ref: 1998-2005; current age: 18-24) |  |  |  | <.001 |  |  |  |  | <.001 |
|  | 1993-1998; age 25-29 | 0.86 | (0.78,0.94) | 0.05 |  |  | 1.00 | (0.97,1.04) | 0.02 |  |
|  | 1983-1993; age 30-39 | 0.85 | (0.78,0.93) | 0.04 |  |  | 1.04 | (1.01,1.07) | 0.02 |  |
|  | 1973-1983; age 40-49 | 0.80 | (0.70,0.91) | 0.07 |  |  | 1.07 | (1.03,1.11) | 0.02 |  |
|  | 1963-1973; age 50-59 | 0.77 | (0.66,0.91) | 0.08 |  |  | 1.15 | (1.10,1.20) | 0.02 |  |
|  | 1953-1963; age 60-69 | 0.72 | (0.56,0.92) | 0.13 |  |  | 1.10 | (1.04,1.17) | 0.03 |  |
|  | 1943-1953; age 70-79 | 0.57 | (0.39,0.85) | 0.20 |  |  | 1.04 | (0.94,1.15) | 0.05 |  |
|  | 1943 or earlier; age 80+ | 0.96 | (0.49,1.88) | 0.34 |  |  | 1.00 | (0.80,1.25) | 0.11 |  |
| Gender | (Ref: Male) |  |  |  | <.001 |  |  |  |  | <.001 |
|  | Female | 0.69 | (0.64,0.74) | 0.04 |  |  | 1.15 | (1.12,1.18) | 0.01 |  |
|  | Other | 0.00 | (0.00,0.00) | 0.87 |  |  | 1.48 | (1.39,1.57) | 0.03 |  |
| Religious affiliation | (Ref: Christianity) |  |  |  | 0.930 |  |  |  |  | 0.058 |
|  | Islam | 1.01 | (0.87,1.19) | 0.08 |  |  | 1.04 | (0.99,1.10) | 0.03 |  |
|  | Collapsed affiliations with prevalence<3% | 0.91 | (0.52,1.58) | 0.28 |  |  | 0.80 | (0.63,1.01) | 0.12 |  |
| Race/ethnicity | (Ref: Plurality group) |  |  |  | <.001 |  |  |  |  | 0.001 |
|  | Non-plurality groups | 1.41 | (1.24,1.59) | 0.06 |  |  | 1.07 | (1.03,1.12) | 0.02 |  |

Table S11c. Sensitivity of the regression estimates to unmeasured confounding for Kenya

|  | | Secular Community Participation | |  | Religious Service Attendance | |
| --- | --- | --- | --- | --- | --- | --- |
| Variable | Category | E-value for Estimate | E-value for 95% CI |  | E-value for Estimate | E-value for 95% CI |
| Relationship with mother | (Ref: Very bad/somewhat bad) |  |  |  |  |  |
|  | Very good/somewhat good | 1.35 | 1.00 |  | 1.26 | 1.00 |
| Relationship with father | (Ref: Very bad/somewhat bad) |  |  |  |  |  |
|  | Very good/somewhat good | 1.53 | 1.00 |  | 1.18 | 1.00 |
| Parent marital status | (Ref: Parents married) |  |  |  |  |  |
|  | Divorced | 1.23 | 1.00 |  | 1.23 | 1.00 |
|  | Parents were never married | 1.14 | 1.00 |  | 1.28 | 1.00 |
|  | One or both parents had died | 1.87 | 1.25 |  | 1.20 | 1.00 |
| Subjective financial status of family growing up | (Ref: Got by) |  |  |  |  |  |
|  | Lived comfortably | 1.14 | 1.00 |  | 1.10 | 1.00 |
|  | Found it difficult | 1.27 | 1.00 |  | 1.06 | 1.00 |
|  | Found it very difficult | 1.19 | 1.00 |  | 1.12 | 1.00 |
| Experienced Abuse | (Ref: No) |  |  |  |  |  |
|  | Yes | 1.91 | 1.61 |  | 1.29 | 1.11 |
| Felt like an outsider in the family | (Ref: No) |  |  |  |  |  |
|  | Yes | 1.25 | 1.00 |  | 1.29 | 1.10 |
| Self-rated health growing up | (Ref: Good) |  |  |  |  |  |
|  | Excellent | 1.42 | 1.00 |  | 1.12 | 1.00 |
|  | Very good | 1.16 | 1.00 |  | 1.21 | 1.00 |
|  | Fair | 1.14 | 1.00 |  | 1.15 | 1.00 |
|  | Poor | 1.30 | 1.00 |  | 1.07 | 1.00 |
| Immigration status | (Ref: Born in this country) |  |  |  |  |  |
|  | Born in another country | 1.28 | 1.00 |  | 1.21 | 1.00 |
| Religious service attendance (age 12) | (Ref: Never) |  |  |  |  |  |
|  | At least 1/week | 1.36 | 1.00 |  | 1.57 | 1.17 |
|  | 1-3/month | 1.48 | 1.00 |  | 1.12 | 1.00 |
|  | < 1/month | 1.33 | 1.00 |  | 1.46 | 1.00 |
| Year of birth | (Ref: 1998-2005; current age: 18-24) |  |  |  |  |  |
|  | 1993-1998; age 25-29 | 1.60 | 1.32 |  | 1.07 | 1.00 |
|  | 1983-1993; age 30-39 | 1.62 | 1.36 |  | 1.25 | 1.09 |
|  | 1973-1983; age 40-49 | 1.80 | 1.43 |  | 1.34 | 1.21 |
|  | 1963-1973; age 50-59 | 1.91 | 1.44 |  | 1.56 | 1.42 |
|  | 1953-1963; age 60-69 | 2.13 | 1.39 |  | 1.44 | 1.23 |
|  | 1943-1953; age 70-79 | 2.88 | 1.62 |  | 1.24 | 1.00 |
|  | 1943 or earlier; age 80+ | 1.25 | 1.00 |  | 1.04 | 1.00 |
| Gender | (Ref: Male) |  |  |  |  |  |
|  | Female | 2.26 | 2.03 |  | 1.56 | 1.48 |
|  | Other | 132209.82 | 24035.51 |  | 2.32 | 2.13 |
| Religious affiliation | (Ref: Christianity) |  |  |  |  |  |
|  | Islam | 1.13 | 1.00 |  | 1.24 | 1.00 |
|  | Collapsed affiliations with prevalence<3% | 1.44 | 1.00 |  | 1.82 | 1.00 |
| Race/ethnicity | (Ref: Plurality group) |  |  |  |  |  |
|  | Non-plurality groups | 2.16 | 1.79 |  | 1.35 | 1.19 |

Table S12a. Nationally representative descriptive statistics for Mexico

| **Characteristic** | **N = 5,776**^1^ |
| --- | --- |
| **Relationship with mother** |  |
| Very good | 3,912 (68%) |
| Somewhat good | 1,340 (23%) |
| Somewhat bad | 177 (3.1%) |
| Very bad | 90 (1.6%) |
| Does not apply | 177 (3.1%) |
| (Missing) | 80 (1.4%) |
| **Relationship with father** |  |
| Very good | 3,089 (53%) |
| Somewhat good | 1,556 (27%) |
| Somewhat bad | 335 (5.8%) |
| Very bad | 267 (4.6%) |
| Does not apply | 470 (8.1%) |
| (Missing) | 60 (1.0%) |
| **Parent marital status** |  |
| Parents married | 3,999 (69%) |
| Divorced | 341 (5.9%) |
| Parents were never married | 827 (14%) |
| One or both parents had died | 176 (3.0%) |
| (Missing) | 432 (7.5%) |
| **Subjective financial status of family growing up** |  |
| Lived comfortably | 1,775 (31%) |
| Got by | 1,872 (32%) |
| Found it difficult | 1,712 (30%) |
| Found it very difficult | 369 (6.4%) |
| (Missing) | 48 (0.8%) |
| **Experienced Abuse** |  |
| Yes | 905 (16%) |
| No | 4,604 (80%) |
| (Missing) | 267 (4.6%) |
| **Felt like an outsider in the family** |  |
| Yes | 772 (13%) |
| No | 4,897 (85%) |
| (Missing) | 107 (1.9%) |
| **Self-rated health growing up** |  |
| Excellent | 1,860 (32%) |
| Very good | 1,350 (23%) |
| Good | 1,677 (29%) |
| Fair | 743 (13%) |
| Poor | 133 (2.3%) |
| (Missing) | 14 (0.2%) |
| **Immigration status** |  |
| Born in this country | 5,517 (96%) |
| Born in another country | 108 (1.9%) |
| (Missing) | 151 (2.6%) |
| **Religious service attendance (age 12)** |  |
| At least 1/week | 2,514 (44%) |
| 1-3/month | 1,162 (20%) |
| <1/month | 1,087 (19%) |
| Never | 944 (16%) |
| (Missing) | 69 (1.2%) |
| **Year of birth** |  |
| 1998-2005; age 18-24 | 986 (17%) |
| 1993-1998; age 25-29 | 623 (11%) |
| 1983-1993; age 30-39 | 1,312 (23%) |
| 1973-1983; age 40-49 | 1,027 (18%) |
| 1963-1973; age 50-59 | 873 (15%) |
| 1953-1963; age 60-69 | 611 (11%) |
| 1943-1953; age 70-79 | 277 (4.8%) |
| 1943 or earlier; age 80+ | 68 (1.2%) |
| (Missing) | 0 (0%) |
| **Gender** |  |
| Male | 2,755 (48%) |
| Female | 2,997 (52%) |
| Other | 3 (<0.1%) |
| (Missing) | 21 (0.4%) |
| **Religious affiliation** |  |
| Christianity | 5,337 (92%) |
| Islam | 6 (<0.1%) |
| Hinduism | 1 (<0.1%) |
| Buddhism | 1 (<0.1%) |
| Judaism | 8 (0.1%) |
| Sikhism | 4 (<0.1%) |
| Baha'i | 1 (<0.1%) |
| Jainism | 0 (0%) |
| Shinto | 2 (<0.1%) |
| Taoism | 5 (<0.1%) |
| Confucianism | 0 (0%) |
| Primal, Animist, or Folk religion | 2 (<0.1%) |
| Spiritism | 0 (0%) |
| Umbanda, Candomble, and other African-derived religions | 0 (0%) |
| Chinese folk/traditional religion | 0 (0%) |
| Some other religion | 7 (0.1%) |
| No religion/Atheist/Agnostic | 328 (5.7%) |
| (Missing) | 74 (1.3%) |
| **Race/Ethnicity** |  |
| Black | 108 (1.9%) |
| Indigenous | 594 (10%) |
| Mestizo | 2,762 (48%) |
| Mulatto | 63 (1.1%) |
| Other | 339 (5.9%) |
| White | 1,116 (19%) |
| (Missing) | 794 (14%) |
| ^1^n (%) | |

Table S12b. Regression of Community Participation on Childhood Predictors for Mexico

|  | | Secular Community Participation | | | |  | Religious Service Attendance | | | |
| --- | --- | --- | --- | --- | --- | --- | --- | --- | --- | --- |
| Variable | Category | Risk-Ratio | RR 95% CI | log(RR) SE | Global  p-value |  | Risk-Ratio | RR 95% CI | log(RR) SE | Global  p-value |
| Relationship with mother | (Ref: Very bad/somewhat bad) |  |  |  | 0.378 |  |  |  |  | 0.280 |
|  | Very good/somewhat good | 1.15 | (0.83,1.61) | 0.17 |  |  | 0.91 | (0.74,1.11) | 0.10 |  |
| Relationship with father | (Ref: Very bad/somewhat bad) |  |  |  | 0.829 |  |  |  |  | 0.173 |
|  | Very good/somewhat good | 0.98 | (0.78,1.23) | 0.12 |  |  | 1.11 | (0.95,1.29) | 0.08 |  |
| Parent marital status | (Ref: Parents married) |  |  |  | 0.057 |  |  |  |  | 0.128 |
|  | Divorced | 1.30 | (1.00,1.69) | 0.13 |  |  | 0.96 | (0.77,1.21) | 0.12 |  |
|  | Parents were never married | 0.87 | (0.69,1.10) | 0.12 |  |  | 0.88 | (0.76,1.03) | 0.08 |  |
|  | One or both parents had died | 0.88 | (0.57,1.37) | 0.22 |  |  | 0.76 | (0.55,1.05) | 0.16 |  |
| Subjective financial status of family growing up | (Ref: Got by) |  |  |  | 0.030 |  |  |  |  | 0.025 |
|  | Lived comfortably | 1.23 | (1.02,1.48) | 0.09 |  |  | 1.05 | (0.93,1.19) | 0.06 |  |
|  | Found it difficult | 0.94 | (0.76,1.16) | 0.11 |  |  | 1.12 | (0.99,1.26) | 0.06 |  |
|  | Found it very difficult | 0.96 | (0.69,1.36) | 0.17 |  |  | 1.30 | (1.09,1.56) | 0.09 |  |
| Experienced Abuse | (Ref: No) |  |  |  | 0.247 |  |  |  |  | 0.289 |
|  | Yes | 1.12 | (0.91,1.38) | 0.11 |  |  | 0.93 | (0.81,1.07) | 0.07 |  |
| Felt like an outsider in the family | (Ref: No) |  |  |  | 0.136 |  |  |  |  | 0.627 |
|  | Yes | 1.15 | (0.95,1.39) | 0.10 |  |  | 1.03 | (0.89,1.18) | 0.07 |  |
| Self-rated health growing up | (Ref: Good) |  |  |  | 0.081 |  |  |  |  | 0.335 |
|  | Excellent | 1.29 | (1.04,1.60) | 0.11 |  |  | 0.92 | (0.80,1.04) | 0.07 |  |
|  | Very good | 1.26 | (1.00,1.59) | 0.12 |  |  | 0.97 | (0.84,1.10) | 0.07 |  |
|  | Fair | 0.99 | (0.73,1.34) | 0.15 |  |  | 1.04 | (0.89,1.20) | 0.07 |  |
|  | Poor | 1.22 | (0.76,1.97) | 0.24 |  |  | 0.83 | (0.60,1.13) | 0.16 |  |
| Immigration status | (Ref: Born in this country) |  |  |  | 0.278 |  |  |  |  | 0.005 |
|  | Born in another country | 0.74 | (0.41,1.35) | 0.30 |  |  | 0.46 | (0.26,0.81) | 0.29 |  |
| Religious service attendance (age 12) | (Ref: Never) |  |  |  | <.001 |  |  |  |  | <.001 |
|  | At least 1/week | 3.05 | (2.27,4.09) | 0.15 |  |  | 2.23 | (1.85,2.69) | 0.10 |  |
|  | 1-3/month | 2.29 | (1.69,3.12) | 0.16 |  |  | 1.42 | (1.15,1.75) | 0.11 |  |
|  | < 1/month | 1.74 | (1.24,2.45) | 0.17 |  |  | 0.80 | (0.62,1.02) | 0.12 |  |
| Year of birth | (Ref: 1998-2005; current age: 18-24) |  |  |  | 0.074 |  |  |  |  | <.001 |
|  | 1993-1998; age 25-29 | 0.99 | (0.78,1.26) | 0.12 |  |  | 1.15 | (0.95,1.39) | 0.10 |  |
|  | 1983-1993; age 30-39 | 0.96 | (0.78,1.19) | 0.11 |  |  | 1.34 | (1.13,1.59) | 0.09 |  |
|  | 1973-1983; age 40-49 | 0.76 | (0.59,0.97) | 0.13 |  |  | 1.48 | (1.25,1.77) | 0.09 |  |
|  | 1963-1973; age 50-59 | 0.76 | (0.57,1.01) | 0.15 |  |  | 1.65 | (1.39,1.95) | 0.09 |  |
|  | 1953-1963; age 60-69 | 0.67 | (0.48,0.93) | 0.17 |  |  | 1.56 | (1.28,1.90) | 0.10 |  |
|  | 1943-1953; age 70-79 | 0.73 | (0.45,1.18) | 0.25 |  |  | 1.70 | (1.37,2.11) | 0.11 |  |
|  | 1943 or earlier; age 80+ | 0.98 | (0.52,1.84) | 0.32 |  |  | 2.29 | (1.71,3.07) | 0.15 |  |
| Gender | (Ref: Male) |  |  |  | <.001 |  |  |  |  | <.001 |
|  | Female | 0.61 | (0.52,0.71) | 0.08 |  |  | 1.18 | (1.07,1.30) | 0.05 |  |
|  | Other | 1.77 | (0.71,4.43) | 0.47 |  |  | 0.31 | (0.08,1.28) | 0.72 |  |
| Religious affiliation | (Ref: No religion/Atheist/Agnostic) |  |  |  | 0.001 |  |  |  |  | <.001 |
|  | Christianity | 0.72 | (0.54,0.97) | 0.15 |  |  | 1.20 | (0.93,1.54) | 0.13 |  |
|  | Collapsed affiliations with prevalence<3% | 1.40 | (0.86,2.28) | 0.25 |  |  | 2.10 | (1.41,3.13) | 0.20 |  |
| Race/ethnicity | (Ref: Plurality group) |  |  |  | 0.238 |  |  |  |  | <.001 |
|  | Non-plurality groups | 1.09 | (0.94,1.28) | 0.08 |  |  | 1.21 | (1.10,1.34) | 0.05 |  |

Table S12c. Sensitivity of the regression estimates to unmeasured confounding for Mexico

|  | | Secular Community Participation | |  | Religious Service Attendance | |
| --- | --- | --- | --- | --- | --- | --- |
| Variable | Category | E-value for Estimate | E-value for 95% CI |  | E-value for Estimate | E-value for 95% CI |
| Relationship with mother | (Ref: Very bad/somewhat bad) |  |  |  |  |  |
|  | Very good/somewhat good | 1.58 | 1.00 |  | 1.44 | 1.00 |
| Relationship with father | (Ref: Very bad/somewhat bad) |  |  |  |  |  |
|  | Very good/somewhat good | 1.18 | 1.00 |  | 1.46 | 1.00 |
| Parent marital status | (Ref: Parents married) |  |  |  |  |  |
|  | Divorced | 1.93 | 1.04 |  | 1.24 | 1.00 |
|  | Parents were never married | 1.57 | 1.00 |  | 1.52 | 1.00 |
|  | One or both parents had died | 1.53 | 1.00 |  | 1.96 | 1.00 |
| Subjective financial status of family growing up | (Ref: Got by) |  |  |  |  |  |
|  | Lived comfortably | 1.75 | 1.16 |  | 1.28 | 1.00 |
|  | Found it difficult | 1.31 | 1.00 |  | 1.49 | 1.00 |
|  | Found it very difficult | 1.23 | 1.00 |  | 1.93 | 1.40 |
| Experienced Abuse | (Ref: No) |  |  |  |  |  |
|  | Yes | 1.49 | 1.00 |  | 1.34 | 1.00 |
| Felt like an outsider in the family | (Ref: No) |  |  |  |  |  |
|  | Yes | 1.57 | 1.00 |  | 1.19 | 1.00 |
| Self-rated health growing up | (Ref: Good) |  |  |  |  |  |
|  | Excellent | 1.91 | 1.25 |  | 1.41 | 1.00 |
|  | Very good | 1.83 | 1.05 |  | 1.23 | 1.00 |
|  | Fair | 1.10 | 1.00 |  | 1.23 | 1.00 |
|  | Poor | 1.74 | 1.00 |  | 1.72 | 1.00 |
| Immigration status | (Ref: Born in this country) |  |  |  |  |  |
|  | Born in another country | 2.03 | 1.00 |  | 3.76 | 1.78 |
| Religious service attendance (age 12) | (Ref: Never) |  |  |  |  |  |
|  | At least 1/week | 5.55 | 3.97 |  | 3.88 | 3.09 |
|  | 1-3/month | 4.02 | 2.76 |  | 2.19 | 1.57 |
|  | < 1/month | 2.88 | 1.78 |  | 1.82 | 1.00 |
| Year of birth | (Ref: 1998-2005; current age: 18-24) |  |  |  |  |  |
|  | 1993-1998; age 25-29 | 1.10 | 1.00 |  | 1.55 | 1.00 |
|  | 1983-1993; age 30-39 | 1.25 | 1.00 |  | 2.02 | 1.52 |
|  | 1973-1983; age 40-49 | 1.98 | 1.21 |  | 2.33 | 1.81 |
|  | 1963-1973; age 50-59 | 1.96 | 1.00 |  | 2.68 | 2.13 |
|  | 1953-1963; age 60-69 | 2.35 | 1.35 |  | 2.49 | 1.87 |
|  | 1943-1953; age 70-79 | 2.10 | 1.00 |  | 2.79 | 2.08 |
|  | 1943 or earlier; age 80+ | 1.16 | 1.00 |  | 4.01 | 2.81 |
| Gender | (Ref: Male) |  |  |  |  |  |
|  | Female | 2.66 | 2.15 |  | 1.64 | 1.34 |
|  | Other | 2.95 | 1.00 |  | 5.84 | 1.00 |
| Religious affiliation | (Ref: No religion/Atheist/Agnostic) |  |  |  |  |  |
|  | Christianity | 2.12 | 1.21 |  | 1.68 | 1.00 |
|  | Collapsed affiliations with prevalence<3% | 2.14 | 1.00 |  | 3.63 | 2.18 |
| Race/ethnicity | (Ref: Plurality group) |  |  |  |  |  |
|  | Non-plurality groups | 1.42 | 1.00 |  | 1.72 | 1.42 |

Table S13a. Nationally representative descriptive statistics for Nigeria

| **Characteristic** | **N = 6,827**^1^ |
| --- | --- |
| **Relationship with mother** |  |
| Very good | 5,986 (88%) |
| Somewhat good | 648 (9.5%) |
| Somewhat bad | 62 (0.9%) |
| Very bad | 18 (0.3%) |
| Does not apply | 104 (1.5%) |
| (Missing) | 9 (0.1%) |
| **Relationship with father** |  |
| Very good | 5,578 (82%) |
| Somewhat good | 924 (14%) |
| Somewhat bad | 76 (1.1%) |
| Very bad | 43 (0.6%) |
| Does not apply | 177 (2.6%) |
| (Missing) | 29 (0.4%) |
| **Parent marital status** |  |
| Parents married | 5,568 (82%) |
| Divorced | 307 (4.5%) |
| Parents were never married | 335 (4.9%) |
| One or both parents had died | 462 (6.8%) |
| (Missing) | 154 (2.3%) |
| **Subjective financial status of family growing up** |  |
| Lived comfortably | 2,192 (32%) |
| Got by | 2,381 (35%) |
| Found it difficult | 1,661 (24%) |
| Found it very difficult | 563 (8.3%) |
| (Missing) | 29 (0.4%) |
| **Experienced Abuse** |  |
| Yes | 880 (13%) |
| No | 5,851 (86%) |
| (Missing) | 96 (1.4%) |
| **Felt like an outsider in the family** |  |
| Yes | 669 (9.8%) |
| No | 6,059 (89%) |
| (Missing) | 99 (1.5%) |
| **Self-rated health growing up** |  |
| Excellent | 2,644 (39%) |
| Very good | 2,613 (38%) |
| Good | 1,152 (17%) |
| Fair | 306 (4.5%) |
| Poor | 98 (1.4%) |
| (Missing) | 14 (0.2%) |
| **Immigration status** |  |
| Born in this country | 6,779 (99%) |
| Born in another country | 47 (0.7%) |
| (Missing) | 1 (<0.1%) |
| **Religious service attendance (age 12)** |  |
| At least 1/week | 5,907 (87%) |
| 1-3/month | 600 (8.8%) |
| <1/month | 136 (2.0%) |
| Never | 138 (2.0%) |
| (Missing) | 45 (0.7%) |
| **Year of birth** |  |
| 1998-2005; age 18-24 | 1,533 (22%) |
| 1993-1998; age 25-29 | 1,193 (17%) |
| 1983-1993; age 30-39 | 1,943 (28%) |
| 1973-1983; age 40-49 | 1,059 (16%) |
| 1963-1973; age 50-59 | 619 (9.1%) |
| 1953-1963; age 60-69 | 296 (4.3%) |
| 1943-1953; age 70-79 | 133 (2.0%) |
| 1943 or earlier; age 80+ | 50 (0.7%) |
| (Missing) | 0 (0%) |
| **Gender** |  |
| Male | 3,371 (49%) |
| Female | 3,456 (51%) |
| Other | 0 (<0.1%) |
| (Missing) | 0 (0%) |
| **Religious affiliation** |  |
| Christianity | 3,463 (51%) |
| Islam | 3,314 (49%) |
| Hinduism | 0 (0%) |
| Buddhism | 0 (<0.1%) |
| Judaism | 0 (0%) |
| Sikhism | 0 (0%) |
| Baha'i | 0 (0%) |
| Jainism | 0 (0%) |
| Shinto | 0 (0%) |
| Taoism | 0 (0%) |
| Confucianism | 0 (<0.1%) |
| Primal, Animist, or Folk religion | 17 (0.3%) |
| Spiritism | 0 (0%) |
| Umbanda, Candomble, and other African-derived religions | 0 (0%) |
| Chinese folk/traditional religion | 0 (0%) |
| Some other religion | 0 (0%) |
| No religion/Atheist/Agnostic | 19 (0.3%) |
| (Missing) | 14 (0.2%) |
| **Race/Ethnicity** |  |
| Edo | 116 (1.7%) |
| Efik | 48 (0.7%) |
| Fulani | 266 (3.9%) |
| Hausa | 2,342 (34%) |
| Ibibio | 180 (2.6%) |
| Idoma | 61 (0.9%) |
| Igala | 77 (1.1%) |
| Igbo (Ibo) | 1,111 (16%) |
| Ijaw | 110 (1.6%) |
| Kanuri | 31 (0.5%) |
| Other | 1,014 (15%) |
| Tiv | 198 (2.9%) |
| Urhobo | 38 (0.6%) |
| Yoruba | 1,230 (18%) |
| (Missing) | 4 (<0.1%) |
| ^1^n (%) | |

Table S13b. Regression of Community Participation on Childhood Predictors for Nigeria

|  | | Secular Community Participation | | | |  | Religious Service Attendance | | | |
| --- | --- | --- | --- | --- | --- | --- | --- | --- | --- | --- |
| Variable | Category | Risk-Ratio | RR 95% CI | log(RR) SE | Global  p-value |  | Risk-Ratio | RR 95% CI | log(RR) SE | Global  p-value |
| Relationship with mother | (Ref: Very bad/somewhat bad) |  |  |  | 0.317 |  |  |  |  | 0.715 |
|  | Very good/somewhat good | 1.18 | (0.85,1.64) | 0.17 |  |  | 1.01 | (0.93,1.10) | 0.04 |  |
| Relationship with father | (Ref: Very bad/somewhat bad) |  |  |  | 0.780 |  |  |  |  | 0.224 |
|  | Very good/somewhat good | 1.04 | (0.79,1.35) | 0.14 |  |  | 1.06 | (0.96,1.17) | 0.05 |  |
| Parent marital status | (Ref: Parents married) |  |  |  | <.001 |  |  |  |  | 0.003 |
|  | Divorced | 1.67 | (1.41,1.96) | 0.08 |  |  | 0.91 | (0.84,0.99) | 0.04 |  |
|  | Parents were never married | 1.03 | (0.84,1.27) | 0.11 |  |  | 0.93 | (0.87,0.99) | 0.03 |  |
|  | One or both parents had died | 1.17 | (0.98,1.40) | 0.09 |  |  | 0.93 | (0.87,1.00) | 0.04 |  |
| Subjective financial status of family growing up | (Ref: Got by) |  |  |  | 0.023 |  |  |  |  | 0.502 |
|  | Lived comfortably | 1.10 | (0.97,1.24) | 0.06 |  |  | 1.01 | (0.98,1.04) | 0.02 |  |
|  | Found it difficult | 0.97 | (0.85,1.10) | 0.07 |  |  | 0.98 | (0.94,1.02) | 0.02 |  |
|  | Found it very difficult | 0.82 | (0.65,1.03) | 0.12 |  |  | 0.99 | (0.94,1.05) | 0.03 |  |
| Experienced Abuse | (Ref: No) |  |  |  | 0.042 |  |  |  |  | 0.004 |
|  | Yes | 1.13 | (1.00,1.27) | 0.06 |  |  | 0.93 | (0.89,0.98) | 0.02 |  |
| Felt like an outsider in the family | (Ref: No) |  |  |  | 0.080 |  |  |  |  | 0.584 |
|  | Yes | 0.88 | (0.76,1.02) | 0.07 |  |  | 1.01 | (0.97,1.05) | 0.02 |  |
| Self-rated health growing up | (Ref: Good) |  |  |  | 0.950 |  |  |  |  | 0.691 |
|  | Excellent | 1.03 | (0.89,1.19) | 0.07 |  |  | 1.01 | (0.97,1.05) | 0.02 |  |
|  | Very good | 0.99 | (0.86,1.13) | 0.07 |  |  | 1.01 | (0.97,1.05) | 0.02 |  |
|  | Fair | 0.95 | (0.71,1.27) | 0.15 |  |  | 0.98 | (0.91,1.06) | 0.04 |  |
|  | Poor | 0.99 | (0.61,1.60) | 0.25 |  |  | 0.94 | (0.83,1.06) | 0.06 |  |
| Immigration status | (Ref: Born in this country) |  |  |  | 0.870 |  |  |  |  | 0.473 |
|  | Born in another country | 1.05 | (0.57,1.93) | 0.31 |  |  | 0.93 | (0.76,1.13) | 0.10 |  |
| Religious service attendance (age 12) | (Ref: Never) |  |  |  | 0.015 |  |  |  |  | 0.034 |
|  | At least 1/week | 1.35 | (0.80,2.28) | 0.27 |  |  | 1.00 | (0.90,1.11) | 0.05 |  |
|  | 1-3/month | 1.40 | (0.82,2.40) | 0.27 |  |  | 0.96 | (0.86,1.08) | 0.06 |  |
|  | < 1/month | 0.89 | (0.48,1.64) | 0.31 |  |  | 0.83 | (0.70,0.99) | 0.09 |  |
| Year of birth | (Ref: 1998-2005; current age: 18-24) |  |  |  | 0.172 |  |  |  |  | 0.920 |
|  | 1993-1998; age 25-29 | 1.05 | (0.93,1.18) | 0.06 |  |  | 1.01 | (0.98,1.05) | 0.02 |  |
|  | 1983-1993; age 30-39 | 1.05 | (0.92,1.20) | 0.07 |  |  | 1.02 | (0.99,1.05) | 0.02 |  |
|  | 1973-1983; age 40-49 | 1.06 | (0.91,1.24) | 0.08 |  |  | 1.02 | (0.98,1.06) | 0.02 |  |
|  | 1963-1973; age 50-59 | 0.77 | (0.57,1.03) | 0.15 |  |  | 1.02 | (0.96,1.09) | 0.03 |  |
|  | 1953-1963; age 60-69 | 0.69 | (0.48,1.00) | 0.19 |  |  | 1.02 | (0.94,1.11) | 0.04 |  |
|  | 1943-1953; age 70-79 | 1.12 | (0.67,1.88) | 0.26 |  |  | 0.98 | (0.85,1.12) | 0.07 |  |
|  | 1943 or earlier; age 80+ | 0.97 | (0.32,2.87) | 0.56 |  |  | 1.05 | (0.90,1.23) | 0.08 |  |
| Gender | (Ref: Male) |  |  |  | <.001 |  |  |  |  | <.001 |
|  | Female | 0.77 | (0.71,0.84) | 0.04 |  |  | 1.00 | (0.97,1.03) | 0.02 |  |
|  | Other | 0.00 | (0.00,0.00) | 1.01 |  |  | 1.19 | (1.13,1.25) | 0.03 |  |
| Religious affiliation | (Ref: Christianity) |  |  |  | 0.444 |  |  |  |  | 0.097 |
|  | Islam | 1.03 | (0.85,1.25) | 0.10 |  |  | 1.04 | (1.00,1.08) | 0.02 |  |
|  | Collapsed affiliations with prevalence<3% | 1.40 | (0.78,2.54) | 0.30 |  |  | 0.93 | (0.65,1.32) | 0.18 |  |
| Race/ethnicity | (Ref: Plurality group) |  |  |  | 0.063 |  |  |  |  | 0.129 |
|  | Non-plurality groups | 1.15 | (0.93,1.41) | 0.10 |  |  | 1.03 | (0.99,1.07) | 0.02 |  |

Table S13c. Sensitivity of the regression estimates to unmeasured confounding for Nigeria

|  | | Secular Community Participation | |  | Religious Service Attendance | |
| --- | --- | --- | --- | --- | --- | --- |
| Variable | Category | E-value for Estimate | E-value for 95% CI |  | E-value for Estimate | E-value for 95% CI |
| Relationship with mother | (Ref: Very bad/somewhat bad) |  |  |  |  |  |
|  | Very good/somewhat good | 1.64 | 1.00 |  | 1.14 | 1.00 |
| Relationship with father | (Ref: Very bad/somewhat bad) |  |  |  |  |  |
|  | Very good/somewhat good | 1.23 | 1.00 |  | 1.31 | 1.00 |
| Parent marital status | (Ref: Parents married) |  |  |  |  |  |
|  | Divorced | 2.72 | 2.18 |  | 1.42 | 1.14 |
|  | Parents were never married | 1.22 | 1.00 |  | 1.36 | 1.13 |
|  | One or both parents had died | 1.63 | 1.00 |  | 1.37 | 1.08 |
| Subjective financial status of family growing up | (Ref: Got by) |  |  |  |  |  |
|  | Lived comfortably | 1.42 | 1.00 |  | 1.10 | 1.00 |
|  | Found it difficult | 1.22 | 1.00 |  | 1.17 | 1.00 |
|  | Found it very difficult | 1.74 | 1.00 |  | 1.08 | 1.00 |
| Experienced Abuse | (Ref: No) |  |  |  |  |  |
|  | Yes | 1.50 | 1.00 |  | 1.34 | 1.17 |
| Felt like an outsider in the family | (Ref: No) |  |  |  |  |  |
|  | Yes | 1.52 | 1.00 |  | 1.11 | 1.00 |
| Self-rated health growing up | (Ref: Good) |  |  |  |  |  |
|  | Excellent | 1.19 | 1.00 |  | 1.10 | 1.00 |
|  | Very good | 1.13 | 1.00 |  | 1.09 | 1.00 |
|  | Fair | 1.30 | 1.00 |  | 1.17 | 1.00 |
|  | Poor | 1.13 | 1.00 |  | 1.32 | 1.00 |
| Immigration status | (Ref: Born in this country) |  |  |  |  |  |
|  | Born in another country | 1.28 | 1.00 |  | 1.36 | 1.00 |
| Religious service attendance (age 12) | (Ref: Never) |  |  |  |  |  |
|  | At least 1/week | 2.03 | 1.00 |  | 1.06 | 1.00 |
|  | 1-3/month | 2.15 | 1.00 |  | 1.25 | 1.00 |
|  | < 1/month | 1.49 | 1.00 |  | 1.70 | 1.10 |
| Year of birth | (Ref: 1998-2005; current age: 18-24) |  |  |  |  |  |
|  | 1993-1998; age 25-29 | 1.28 | 1.00 |  | 1.12 | 1.00 |
|  | 1983-1993; age 30-39 | 1.29 | 1.00 |  | 1.15 | 1.00 |
|  | 1973-1983; age 40-49 | 1.32 | 1.00 |  | 1.16 | 1.00 |
|  | 1963-1973; age 50-59 | 1.93 | 1.00 |  | 1.18 | 1.00 |
|  | 1953-1963; age 60-69 | 2.25 | 1.00 |  | 1.17 | 1.00 |
|  | 1943-1953; age 70-79 | 1.49 | 1.00 |  | 1.19 | 1.00 |
|  | 1943 or earlier; age 80+ | 1.23 | 1.00 |  | 1.29 | 1.00 |
| Gender | (Ref: Male) |  |  |  |  |  |
|  | Female | 1.92 | 1.67 |  | 1.05 | 1.00 |
|  | Other | 25493.50 | 3538.77 |  | 1.66 | 1.50 |
| Religious affiliation | (Ref: Christianity) |  |  |  |  |  |
|  | Islam | 1.20 | 1.00 |  | 1.24 | 1.04 |
|  | Collapsed affiliations with prevalence<3% | 2.16 | 1.00 |  | 1.37 | 1.00 |
| Race/ethnicity | (Ref: Plurality group) |  |  |  |  |  |
|  | Non-plurality groups | 1.56 | 1.00 |  | 1.21 | 1.00 |

Table S14a. Nationally representative descriptive statistics for Philippines

| **Characteristic** | **N = 5,292**^1^ |
| --- | --- |
| **Relationship with mother** |  |
| Very good | 3,333 (63%) |
| Somewhat good | 1,703 (32%) |
| Somewhat bad | 124 (2.3%) |
| Very bad | 39 (0.7%) |
| Does not apply | 59 (1.1%) |
| (Missing) | 35 (0.7%) |
| **Relationship with father** |  |
| Very good | 3,443 (65%) |
| Somewhat good | 1,429 (27%) |
| Somewhat bad | 159 (3.0%) |
| Very bad | 58 (1.1%) |
| Does not apply | 108 (2.0%) |
| (Missing) | 95 (1.8%) |
| **Parent marital status** |  |
| Parents married | 4,575 (86%) |
| Divorced | 64 (1.2%) |
| Parents were never married | 517 (9.8%) |
| One or both parents had died | 51 (1.0%) |
| (Missing) | 85 (1.6%) |
| **Subjective financial status of family growing up** |  |
| Lived comfortably | 937 (18%) |
| Got by | 3,006 (57%) |
| Found it difficult | 1,055 (20%) |
| Found it very difficult | 291 (5.5%) |
| (Missing) | 3 (<0.1%) |
| **Experienced Abuse** |  |
| Yes | 420 (7.9%) |
| No | 4,837 (91%) |
| (Missing) | 35 (0.7%) |
| **Felt like an outsider in the family** |  |
| Yes | 395 (7.5%) |
| No | 4,884 (92%) |
| (Missing) | 13 (0.2%) |
| **Self-rated health growing up** |  |
| Excellent | 1,041 (20%) |
| Very good | 559 (11%) |
| Good | 2,174 (41%) |
| Fair | 1,246 (24%) |
| Poor | 272 (5.1%) |
| (Missing) | 0 (<0.1%) |
| **Immigration status** |  |
| Born in this country | 5,284 (100%) |
| Born in another country | 8 (0.1%) |
| (Missing) | 0 (0%) |
| **Religious service attendance (age 12)** |  |
| At least 1/week | 2,453 (46%) |
| 1-3/month | 1,699 (32%) |
| <1/month | 892 (17%) |
| Never | 201 (3.8%) |
| (Missing) | 47 (0.9%) |
| **Year of birth** |  |
| 1998-2005; age 18-24 | 1,073 (20%) |
| 1993-1998; age 25-29 | 695 (13%) |
| 1983-1993; age 30-39 | 1,160 (22%) |
| 1973-1983; age 40-49 | 972 (18%) |
| 1963-1973; age 50-59 | 732 (14%) |
| 1953-1963; age 60-69 | 495 (9.4%) |
| 1943-1953; age 70-79 | 143 (2.7%) |
| 1943 or earlier; age 80+ | 23 (0.4%) |
| (Missing) | 0 (0%) |
| **Gender** |  |
| Male | 2,625 (50%) |
| Female | 2,643 (50%) |
| Other | 13 (0.2%) |
| (Missing) | 11 (0.2%) |
| **Religious affiliation** |  |
| Christianity | 4,968 (94%) |
| Islam | 276 (5.2%) |
| Hinduism | 0 (0%) |
| Buddhism | 1 (<0.1%) |
| Judaism | 0 (0%) |
| Sikhism | 4 (<0.1%) |
| Baha'i | 1 (<0.1%) |
| Jainism | 0 (0%) |
| Shinto | 0 (0%) |
| Taoism | 0 (0%) |
| Confucianism | 0 (0%) |
| Primal, Animist, or Folk religion | 14 (0.3%) |
| Spiritism | 0 (0%) |
| Umbanda, Candomble, and other African-derived religions | 0 (0%) |
| Chinese folk/traditional religion | 0 (0%) |
| Some other religion | 9 (0.2%) |
| No religion/Atheist/Agnostic | 9 (0.2%) |
| (Missing) | 11 (0.2%) |
| **Race/Ethnicity** |  |
| Aeta | 1 (<0.1%) |
| Badjao | 2 (<0.1%) |
| Bicolano/Bikolano | 300 (5.7%) |
| Cebuano | 656 (12%) |
| Chinese-Filipino | 3 (<0.1%) |
| Igorot | 42 (0.8%) |
| Ilocano/Ilokano | 429 (8.1%) |
| Ilonggo/Hiligaynon | 428 (8.1%) |
| Kapampangan | 107 (2.0%) |
| Maguindanaoan | 84 (1.6%) |
| Mangyan | 2 (<0.1%) |
| Maranao | 39 (0.7%) |
| Masbateno | 54 (1.0%) |
| Other | 244 (4.6%) |
| Pangasinense | 107 (2.0%) |
| Tagalog | 1,691 (32%) |
| Tausug | 94 (1.8%) |
| Visayan/Bisaya | 739 (14%) |
| Waray | 216 (4.1%) |
| Zamboangueno | 51 (1.0%) |
| (Missing) | 3 (<0.1%) |
| ^1^n (%) | |

Table S14b. Regression of Community Participation on Childhood Predictors for Philippines

|  | | Secular Community Participation | | | |  | Religious Service Attendance | | | |
| --- | --- | --- | --- | --- | --- | --- | --- | --- | --- | --- |
| Variable | Category | Risk-Ratio | RR 95% CI | log(RR) SE | Global  p-value |  | Risk-Ratio | RR 95% CI | log(RR) SE | Global  p-value |
| Relationship with mother | (Ref: Very bad/somewhat bad) |  |  |  | 0.258 |  |  |  |  | 0.540 |
|  | Very good/somewhat good | 0.80 | (0.53,1.19) | 0.20 |  |  | 1.06 | (0.88,1.28) | 0.10 |  |
| Relationship with father | (Ref: Very bad/somewhat bad) |  |  |  | 0.207 |  |  |  |  | 0.110 |
|  | Very good/somewhat good | 1.25 | (0.88,1.77) | 0.18 |  |  | 1.14 | (0.97,1.35) | 0.08 |  |
| Parent marital status | (Ref: Parents married) |  |  |  | 0.452 |  |  |  |  | 0.615 |
|  | Divorced | 0.82 | (0.43,1.56) | 0.33 |  |  | 0.82 | (0.61,1.11) | 0.15 |  |
|  | Parents were never married | 1.09 | (0.85,1.40) | 0.13 |  |  | 1.01 | (0.90,1.13) | 0.06 |  |
|  | One or both parents had died | 1.46 | (0.83,2.57) | 0.29 |  |  | 0.97 | (0.71,1.33) | 0.16 |  |
| Subjective financial status of family growing up | (Ref: Got by) |  |  |  | 0.024 |  |  |  |  | 0.055 |
|  | Lived comfortably | 0.90 | (0.73,1.11) | 0.11 |  |  | 1.08 | (1.00,1.17) | 0.04 |  |
|  | Found it difficult | 0.89 | (0.74,1.06) | 0.09 |  |  | 0.95 | (0.87,1.04) | 0.05 |  |
|  | Found it very difficult | 1.38 | (1.04,1.83) | 0.14 |  |  | 0.99 | (0.88,1.11) | 0.06 |  |
| Experienced Abuse | (Ref: No) |  |  |  | 0.089 |  |  |  |  | 0.322 |
|  | Yes | 1.22 | (0.97,1.55) | 0.12 |  |  | 0.94 | (0.83,1.07) | 0.07 |  |
| Felt like an outsider in the family | (Ref: No) |  |  |  | 0.507 |  |  |  |  | 0.327 |
|  | Yes | 1.09 | (0.84,1.41) | 0.13 |  |  | 1.06 | (0.94,1.19) | 0.06 |  |
| Self-rated health growing up | (Ref: Good) |  |  |  | 0.011 |  |  |  |  | 0.016 |
|  | Excellent | 1.09 | (0.90,1.33) | 0.10 |  |  | 1.06 | (0.98,1.14) | 0.04 |  |
|  | Very good | 1.22 | (0.98,1.52) | 0.11 |  |  | 1.02 | (0.90,1.14) | 0.06 |  |
|  | Fair | 0.80 | (0.66,0.98) | 0.10 |  |  | 0.92 | (0.85,0.99) | 0.04 |  |
|  | Poor | 0.92 | (0.65,1.29) | 0.17 |  |  | 1.12 | (0.99,1.26) | 0.06 |  |
| Immigration status | (Ref: Born in this country) |  |  |  | 0.432 |  |  |  |  | 0.657 |
|  | Born in another country | 1.60 | (0.50,5.13) | 0.60 |  |  | 0.84 | (0.38,1.84) | 0.40 |  |
| Religious service attendance (age 12) | (Ref: Never) |  |  |  | <.001 |  |  |  |  | <.001 |
|  | At least 1/week | 2.09 | (1.27,3.46) | 0.26 |  |  | 1.58 | (1.26,1.98) | 0.11 |  |
|  | 1-3/month | 1.89 | (1.14,3.14) | 0.26 |  |  | 1.17 | (0.93,1.46) | 0.11 |  |
|  | < 1/month | 1.29 | (0.76,2.18) | 0.27 |  |  | 0.90 | (0.71,1.15) | 0.12 |  |
| Year of birth | (Ref: 1998-2005; current age: 18-24) |  |  |  | 0.006 |  |  |  |  | 0.089 |
|  | 1993-1998; age 25-29 | 0.87 | (0.67,1.12) | 0.13 |  |  | 0.94 | (0.84,1.05) | 0.06 |  |
|  | 1983-1993; age 30-39 | 0.70 | (0.56,0.87) | 0.11 |  |  | 0.98 | (0.90,1.08) | 0.05 |  |
|  | 1973-1983; age 40-49 | 0.78 | (0.62,0.98) | 0.12 |  |  | 1.04 | (0.94,1.15) | 0.05 |  |
|  | 1963-1973; age 50-59 | 0.99 | (0.80,1.23) | 0.11 |  |  | 1.05 | (0.94,1.17) | 0.06 |  |
|  | 1953-1963; age 60-69 | 0.79 | (0.57,1.09) | 0.16 |  |  | 1.15 | (1.02,1.30) | 0.06 |  |
|  | 1943-1953; age 70-79 | 0.82 | (0.50,1.35) | 0.25 |  |  | 1.14 | (0.93,1.39) | 0.10 |  |
|  | 1943 or earlier; age 80+ | 0.32 | (0.12,0.87) | 0.51 |  |  | 0.99 | (0.62,1.58) | 0.24 |  |
| Gender | (Ref: Male) |  |  |  | <.001 |  |  |  |  | 0.011 |
|  | Female | 0.65 | (0.56,0.76) | 0.08 |  |  | 1.10 | (1.03,1.17) | 0.03 |  |
|  | Other | 1.19 | (0.50,2.88) | 0.45 |  |  | 0.79 | (0.42,1.49) | 0.32 |  |
| Religious affiliation | (Ref: Christianity) |  |  |  | 0.025 |  |  |  |  | <.001 |
|  | Islam | 1.37 | (1.09,1.72) | 0.12 |  |  | 1.47 | (1.35,1.60) | 0.04 |  |
|  | Collapsed affiliations with prevalence<3% | 1.12 | (0.47,2.66) | 0.44 |  |  | 1.04 | (0.70,1.55) | 0.20 |  |
| Race/ethnicity | (Ref: Plurality group) |  |  |  | 0.199 |  |  |  |  | 0.074 |
|  | Non-plurality groups | 0.91 | (0.78,1.06) | 0.08 |  |  | 1.07 | (0.99,1.15) | 0.04 |  |

Table S14c. Sensitivity of the regression estimates to unmeasured confounding for Philippines

|  | | Secular Community Participation | |  | Religious Service Attendance | |
| --- | --- | --- | --- | --- | --- | --- |
| Variable | Category | E-value for Estimate | E-value for 95% CI |  | E-value for Estimate | E-value for 95% CI |
| Relationship with mother | (Ref: Very bad/somewhat bad) |  |  |  |  |  |
|  | Very good/somewhat good | 1.82 | 1.00 |  | 1.31 | 1.00 |
| Relationship with father | (Ref: Very bad/somewhat bad) |  |  |  |  |  |
|  | Very good/somewhat good | 1.80 | 1.00 |  | 1.54 | 1.00 |
| Parent marital status | (Ref: Parents married) |  |  |  |  |  |
|  | Divorced | 1.73 | 1.00 |  | 1.72 | 1.00 |
|  | Parents were never married | 1.41 | 1.00 |  | 1.08 | 1.00 |
|  | One or both parents had died | 2.29 | 1.00 |  | 1.20 | 1.00 |
| Subjective financial status of family growing up | (Ref: Got by) |  |  |  |  |  |
|  | Lived comfortably | 1.45 | 1.00 |  | 1.38 | 1.06 |
|  | Found it difficult | 1.51 | 1.00 |  | 1.29 | 1.00 |
|  | Found it very difficult | 2.11 | 1.24 |  | 1.13 | 1.00 |
| Experienced Abuse | (Ref: No) |  |  |  |  |  |
|  | Yes | 1.75 | 1.00 |  | 1.33 | 1.00 |
| Felt like an outsider in the family | (Ref: No) |  |  |  |  |  |
|  | Yes | 1.40 | 1.00 |  | 1.31 | 1.00 |
| Self-rated health growing up | (Ref: Good) |  |  |  |  |  |
|  | Excellent | 1.41 | 1.00 |  | 1.30 | 1.00 |
|  | Very good | 1.75 | 1.00 |  | 1.14 | 1.00 |
|  | Fair | 1.80 | 1.19 |  | 1.40 | 1.10 |
|  | Poor | 1.41 | 1.00 |  | 1.47 | 1.00 |
| Immigration status | (Ref: Born in this country) |  |  |  |  |  |
|  | Born in another country | 2.57 | 1.00 |  | 1.68 | 1.00 |
| Religious service attendance (age 12) | (Ref: Never) |  |  |  |  |  |
|  | At least 1/week | 3.61 | 1.85 |  | 2.54 | 1.84 |
|  | 1-3/month | 3.19 | 1.54 |  | 1.60 | 1.00 |
|  | < 1/month | 1.89 | 1.00 |  | 1.45 | 1.00 |
| Year of birth | (Ref: 1998-2005; current age: 18-24) |  |  |  |  |  |
|  | 1993-1998; age 25-29 | 1.57 | 1.00 |  | 1.33 | 1.00 |
|  | 1983-1993; age 30-39 | 2.22 | 1.56 |  | 1.14 | 1.00 |
|  | 1973-1983; age 40-49 | 1.89 | 1.17 |  | 1.23 | 1.00 |
|  | 1963-1973; age 50-59 | 1.09 | 1.00 |  | 1.28 | 1.00 |
|  | 1953-1963; age 60-69 | 1.85 | 1.00 |  | 1.56 | 1.14 |
|  | 1943-1953; age 70-79 | 1.73 | 1.00 |  | 1.53 | 1.00 |
|  | 1943 or earlier; age 80+ | 5.75 | 1.57 |  | 1.11 | 1.00 |
| Gender | (Ref: Male) |  |  |  |  |  |
|  | Female | 2.43 | 1.94 |  | 1.42 | 1.21 |
|  | Other | 1.67 | 1.00 |  | 1.83 | 1.00 |
| Religious affiliation | (Ref: Christianity) |  |  |  |  |  |
|  | Islam | 2.08 | 1.40 |  | 2.30 | 2.03 |
|  | Collapsed affiliations with prevalence<3% | 1.50 | 1.00 |  | 1.24 | 1.00 |
| Race/ethnicity | (Ref: Plurality group) |  |  |  |  |  |
|  | Non-plurality groups | 1.44 | 1.00 |  | 1.33 | 1.00 |

Table S15a. Nationally representative descriptive statistics for Poland

| **Characteristic** | **N = 10,389**^1^ |
| --- | --- |
| **Relationship with mother** |  |
| Very good | 4,879 (47%) |
| Somewhat good | 4,973 (48%) |
| Somewhat bad | 285 (2.7%) |
| Very bad | 58 (0.6%) |
| Does not apply | 80 (0.8%) |
| (Missing) | 112 (1.1%) |
| **Relationship with father** |  |
| Very good | 4,231 (41%) |
| Somewhat good | 4,984 (48%) |
| Somewhat bad | 516 (5.0%) |
| Very bad | 78 (0.7%) |
| Does not apply | 407 (3.9%) |
| (Missing) | 173 (1.7%) |
| **Parent marital status** |  |
| Parents married | 8,972 (86%) |
| Divorced | 587 (5.7%) |
| Parents were never married | 193 (1.9%) |
| One or both parents had died | 313 (3.0%) |
| (Missing) | 324 (3.1%) |
| **Subjective financial status of family growing up** |  |
| Lived comfortably | 1,384 (13%) |
| Got by | 6,257 (60%) |
| Found it difficult | 2,133 (21%) |
| Found it very difficult | 509 (4.9%) |
| (Missing) | 106 (1.0%) |
| **Experienced Abuse** |  |
| Yes | 325 (3.1%) |
| No | 10,009 (96%) |
| (Missing) | 55 (0.5%) |
| **Felt like an outsider in the family** |  |
| Yes | 490 (4.7%) |
| No | 9,615 (93%) |
| (Missing) | 284 (2.7%) |
| **Self-rated health growing up** |  |
| Excellent | 2,676 (26%) |
| Very good | 5,371 (52%) |
| Good | 1,779 (17%) |
| Fair | 406 (3.9%) |
| Poor | 123 (1.2%) |
| (Missing) | 34 (0.3%) |
| **Immigration status** |  |
| Born in this country | 10,258 (99%) |
| Born in another country | 108 (1.0%) |
| (Missing) | 23 (0.2%) |
| **Religious service attendance (age 12)** |  |
| At least 1/week | 4,751 (46%) |
| 1-3/month | 2,689 (26%) |
| <1/month | 2,161 (21%) |
| Never | 354 (3.4%) |
| (Missing) | 434 (4.2%) |
| **Year of birth** |  |
| 1998-2005; age 18-24 | 955 (9.2%) |
| 1993-1998; age 25-29 | 761 (7.3%) |
| 1983-1993; age 30-39 | 2,159 (21%) |
| 1973-1983; age 40-49 | 1,956 (19%) |
| 1963-1973; age 50-59 | 1,670 (16%) |
| 1953-1963; age 60-69 | 1,909 (18%) |
| 1943-1953; age 70-79 | 833 (8.0%) |
| 1943 or earlier; age 80+ | 145 (1.4%) |
| (Missing) | 1 (<0.1%) |
| **Gender** |  |
| Male | 4,974 (48%) |
| Female | 5,387 (52%) |
| Other | 3 (<0.1%) |
| (Missing) | 26 (0.2%) |
| **Religious affiliation** |  |
| Christianity | 9,861 (95%) |
| Islam | 3 (<0.1%) |
| Hinduism | 0 (0%) |
| Buddhism | 2 (<0.1%) |
| Judaism | 0 (0%) |
| Sikhism | 1 (<0.1%) |
| Baha'i | 0 (0%) |
| Jainism | 0 (0%) |
| Shinto | 0 (0%) |
| Taoism | 0 (0%) |
| Confucianism | 0 (0%) |
| Primal, Animist, or Folk religion | 5 (<0.1%) |
| Spiritism | 0 (0%) |
| Umbanda, Candomble, and other African-derived religions | 0 (0%) |
| Chinese folk/traditional religion | 0 (0%) |
| Some other religion | 0 (0%) |
| No religion/Atheist/Agnostic | 482 (4.6%) |
| (Missing) | 35 (0.3%) |
| **Race/Ethnicity** |  |
| Belarussian | 2 (<0.1%) |
| German | 4 (<0.1%) |
| Kashubians | 3 (<0.1%) |
| Other | 4 (<0.1%) |
| Polish | 10,309 (99%) |
| Silesia | 14 (0.1%) |
| Ukrainian | 38 (0.4%) |
| (Missing) | 14 (0.1%) |
| ^1^n (%) | |

Table S15b. Regression of Community Participation on Childhood Predictors for Poland

|  | | Secular Community Participation | | | |  | Religious Service Attendance | | | |
| --- | --- | --- | --- | --- | --- | --- | --- | --- | --- | --- |
| Variable | Category | Risk-Ratio | RR 95% CI | log(RR) SE | Global  p-value |  | Risk-Ratio | RR 95% CI | log(RR) SE | Global  p-value |
| Relationship with mother | (Ref: Very bad/somewhat bad) |  |  |  | 0.391 |  |  |  |  | 0.398 |
|  | Very good/somewhat good | 0.80 | (0.47,1.36) | 0.27 |  |  | 1.16 | (0.81,1.65) | 0.18 |  |
| Relationship with father | (Ref: Very bad/somewhat bad) |  |  |  | 0.718 |  |  |  |  | 0.150 |
|  | Very good/somewhat good | 1.10 | (0.66,1.83) | 0.26 |  |  | 1.24 | (0.92,1.67) | 0.15 |  |
| Parent marital status | (Ref: Parents married) |  |  |  | <.001 |  |  |  |  | 0.470 |
|  | Divorced | 1.51 | (1.07,2.12) | 0.17 |  |  | 0.82 | (0.65,1.05) | 0.12 |  |
|  | Parents were never married | 2.29 | (1.59,3.32) | 0.19 |  |  | 0.99 | (0.73,1.36) | 0.16 |  |
|  | One or both parents had died | 1.73 | (1.01,2.95) | 0.27 |  |  | 0.97 | (0.76,1.24) | 0.12 |  |
| Subjective financial status of family growing up | (Ref: Got by) |  |  |  | 0.111 |  |  |  |  | 0.046 |
|  | Lived comfortably | 1.23 | (0.93,1.64) | 0.14 |  |  | 0.87 | (0.76,1.00) | 0.07 |  |
|  | Found it difficult | 1.20 | (0.91,1.58) | 0.14 |  |  | 1.07 | (0.98,1.16) | 0.04 |  |
|  | Found it very difficult | 1.69 | (1.01,2.83) | 0.26 |  |  | 0.97 | (0.79,1.18) | 0.10 |  |
| Experienced Abuse | (Ref: No) |  |  |  | 0.593 |  |  |  |  | 0.009 |
|  | Yes | 1.10 | (0.78,1.55) | 0.18 |  |  | 0.70 | (0.54,0.92) | 0.13 |  |
| Felt like an outsider in the family | (Ref: No) |  |  |  | 0.638 |  |  |  |  | 0.803 |
|  | Yes | 1.09 | (0.76,1.56) | 0.18 |  |  | 1.00 | (0.80,1.25) | 0.11 |  |
| Self-rated health growing up | (Ref: Good) |  |  |  | <.001 |  |  |  |  | 0.010 |
|  | Excellent | 0.56 | (0.42,0.76) | 0.15 |  |  | 0.89 | (0.78,1.03) | 0.07 |  |
|  | Very good | 0.79 | (0.65,0.97) | 0.10 |  |  | 1.07 | (0.95,1.20) | 0.06 |  |
|  | Fair | 1.03 | (0.70,1.52) | 0.20 |  |  | 0.89 | (0.72,1.11) | 0.11 |  |
|  | Poor | 1.82 | (0.96,3.45) | 0.33 |  |  | 0.72 | (0.45,1.16) | 0.24 |  |
| Immigration status | (Ref: Born in this country) |  |  |  | 0.001 |  |  |  |  | 0.323 |
|  | Born in another country | 2.68 | (1.45,4.94) | 0.31 |  |  | 1.25 | (0.79,1.98) | 0.23 |  |
| Religious service attendance (age 12) | (Ref: Never) |  |  |  | 0.667 |  |  |  |  | <.001 |
|  | At least 1/week | 0.88 | (0.54,1.45) | 0.25 |  |  | 5.05 | (3.34,7.63) | 0.21 |  |
|  | 1-3/month | 1.01 | (0.59,1.71) | 0.27 |  |  | 2.27 | (1.46,3.51) | 0.22 |  |
|  | < 1/month | 0.91 | (0.57,1.44) | 0.23 |  |  | 1.28 | (0.83,1.98) | 0.22 |  |
| Year of birth | (Ref: 1998-2005; current age: 18-24) |  |  |  | 0.001 |  |  |  |  | <.001 |
|  | 1993-1998; age 25-29 | 0.77 | (0.57,1.05) | 0.15 |  |  | 0.87 | (0.69,1.10) | 0.12 |  |
|  | 1983-1993; age 30-39 | 0.66 | (0.51,0.85) | 0.13 |  |  | 1.08 | (0.87,1.34) | 0.11 |  |
|  | 1973-1983; age 40-49 | 0.58 | (0.44,0.77) | 0.14 |  |  | 1.20 | (0.97,1.49) | 0.11 |  |
|  | 1963-1973; age 50-59 | 0.55 | (0.39,0.77) | 0.17 |  |  | 1.51 | (1.21,1.88) | 0.11 |  |
|  | 1953-1963; age 60-69 | 0.60 | (0.41,0.88) | 0.19 |  |  | 1.83 | (1.45,2.31) | 0.12 |  |
|  | 1943-1953; age 70-79 | 0.66 | (0.39,1.11) | 0.27 |  |  | 1.91 | (1.50,2.44) | 0.13 |  |
|  | 1943 or earlier; age 80+ | 1.17 | (0.61,2.23) | 0.33 |  |  | 1.74 | (1.17,2.58) | 0.20 |  |
| Gender | (Ref: Male) |  |  |  | <.001 |  |  |  |  | <.001 |
|  | Female | 1.02 | (0.86,1.21) | 0.09 |  |  | 1.27 | (1.18,1.37) | 0.04 |  |
|  | Other | 0.00 | (0.00,0.00) | 0.68 |  |  | 1.00 | (0.14,7.36) | 1.02 |  |
| Religious affiliation | (Ref: No religion/Atheist/Agnostic) |  |  |  | 0.490 |  |  |  |  | 0.559 |
|  | Christianity | 1.12 | (0.71,1.77) | 0.23 |  |  | 0.99 | (0.73,1.34) | 0.15 |  |
|  | Collapsed affiliations with prevalence<3% | 2.06 | (0.58,7.31) | 0.65 |  |  | 1.54 | (0.60,3.95) | 0.48 |  |
| Race/ethnicity | (Ref: Plurality group) |  |  |  | 0.005 |  |  |  |  | 0.214 |
|  | Non-plurality groups | 0.17 | (0.05,0.59) | 0.63 |  |  | 0.66 | (0.33,1.30) | 0.35 |  |

Table S15c. Sensitivity of the regression estimates to unmeasured confounding for Poland

|  | | Secular Community Participation | |  | Religious Service Attendance | |
| --- | --- | --- | --- | --- | --- | --- |
| Variable | Category | E-value for Estimate | E-value for 95% CI |  | E-value for Estimate | E-value for 95% CI |
| Relationship with mother | (Ref: Very bad/somewhat bad) |  |  |  |  |  |
|  | Very good/somewhat good | 1.81 | 1.00 |  | 1.59 | 1.00 |
| Relationship with father | (Ref: Very bad/somewhat bad) |  |  |  |  |  |
|  | Very good/somewhat good | 1.42 | 1.00 |  | 1.79 | 1.00 |
| Parent marital status | (Ref: Parents married) |  |  |  |  |  |
|  | Divorced | 2.38 | 1.35 |  | 1.72 | 1.00 |
|  | Parents were never married | 4.02 | 2.55 |  | 1.09 | 1.00 |
|  | One or both parents had died | 2.85 | 1.14 |  | 1.21 | 1.00 |
| Subjective financial status of family growing up | (Ref: Got by) |  |  |  |  |  |
|  | Lived comfortably | 1.77 | 1.00 |  | 1.57 | 1.06 |
|  | Found it difficult | 1.68 | 1.00 |  | 1.33 | 1.00 |
|  | Found it very difficult | 2.76 | 1.09 |  | 1.21 | 1.00 |
| Experienced Abuse | (Ref: No) |  |  |  |  |  |
|  | Yes | 1.43 | 1.00 |  | 2.20 | 1.41 |
| Felt like an outsider in the family | (Ref: No) |  |  |  |  |  |
|  | Yes | 1.39 | 1.00 |  | 1.05 | 1.00 |
| Self-rated health growing up | (Ref: Good) |  |  |  |  |  |
|  | Excellent | 2.96 | 1.97 |  | 1.49 | 1.00 |
|  | Very good | 1.84 | 1.22 |  | 1.34 | 1.00 |
|  | Fair | 1.20 | 1.00 |  | 1.48 | 1.00 |
|  | Poor | 3.05 | 1.00 |  | 2.11 | 1.00 |
| Immigration status | (Ref: Born in this country) |  |  |  |  |  |
|  | Born in another country | 4.80 | 2.26 |  | 1.82 | 1.00 |
| Religious service attendance (age 12) | (Ref: Never) |  |  |  |  |  |
|  | At least 1/week | 1.51 | 1.00 |  | 9.57 | 6.13 |
|  | 1-3/month | 1.09 | 1.00 |  | 3.96 | 2.29 |
|  | < 1/month | 1.44 | 1.00 |  | 1.88 | 1.00 |
| Year of birth | (Ref: 1998-2005; current age: 18-24) |  |  |  |  |  |
|  | 1993-1998; age 25-29 | 1.91 | 1.00 |  | 1.55 | 1.00 |
|  | 1983-1993; age 30-39 | 2.39 | 1.62 |  | 1.37 | 1.00 |
|  | 1973-1983; age 40-49 | 2.85 | 1.94 |  | 1.69 | 1.00 |
|  | 1963-1973; age 50-59 | 3.03 | 1.91 |  | 2.39 | 1.72 |
|  | 1953-1963; age 60-69 | 2.72 | 1.54 |  | 3.06 | 2.26 |
|  | 1943-1953; age 70-79 | 2.40 | 1.00 |  | 3.24 | 2.36 |
|  | 1943 or earlier; age 80+ | 1.62 | 1.00 |  | 2.87 | 1.62 |
| Gender | (Ref: Male) |  |  |  |  |  |
|  | Female | 1.14 | 1.00 |  | 1.86 | 1.64 |
|  | Other | 151131.55 | 39569.62 |  | 1.03 | 1.00 |
| Religious affiliation | (Ref: No religion/Atheist/Agnostic) |  |  |  |  |  |
|  | Christianity | 1.50 | 1.00 |  | 1.11 | 1.00 |
|  | Collapsed affiliations with prevalence<3% | 3.53 | 1.00 |  | 2.45 | 1.00 |
| Race/ethnicity | (Ref: Plurality group) |  |  |  |  |  |
|  | Non-plurality groups | 11.01 | 2.77 |  | 2.40 | 1.00 |

Table S16a. Nationally representative descriptive statistics for South Africa

| **Characteristic** | **N = 2,651**^1^ |
| --- | --- |
| **Relationship with mother** |  |
| Very good | 2,186 (82%) |
| Somewhat good | 263 (9.9%) |
| Somewhat bad | 51 (1.9%) |
| Very bad | 39 (1.5%) |
| Does not apply | 90 (3.4%) |
| (Missing) | 21 (0.8%) |
| **Relationship with father** |  |
| Very good | 1,656 (62%) |
| Somewhat good | 333 (13%) |
| Somewhat bad | 86 (3.3%) |
| Very bad | 159 (6.0%) |
| Does not apply | 331 (12%) |
| (Missing) | 85 (3.2%) |
| **Parent marital status** |  |
| Parents married | 1,321 (50%) |
| Divorced | 131 (5.0%) |
| Parents were never married | 904 (34%) |
| One or both parents had died | 140 (5.3%) |
| (Missing) | 155 (5.8%) |
| **Subjective financial status of family growing up** |  |
| Lived comfortably | 1,050 (40%) |
| Got by | 875 (33%) |
| Found it difficult | 432 (16%) |
| Found it very difficult | 289 (11%) |
| (Missing) | 5 (0.2%) |
| **Experienced Abuse** |  |
| Yes | 450 (17%) |
| No | 2,149 (81%) |
| (Missing) | 52 (2.0%) |
| **Felt like an outsider in the family** |  |
| Yes | 434 (16%) |
| No | 2,211 (83%) |
| (Missing) | 6 (0.2%) |
| **Self-rated health growing up** |  |
| Excellent | 1,225 (46%) |
| Very good | 590 (22%) |
| Good | 370 (14%) |
| Fair | 266 (10%) |
| Poor | 183 (6.9%) |
| (Missing) | 17 (0.6%) |
| **Immigration status** |  |
| Born in this country | 2,511 (95%) |
| Born in another country | 139 (5.2%) |
| (Missing) | 1 (<0.1%) |
| **Religious service attendance (age 12)** |  |
| At least 1/week | 1,681 (63%) |
| 1-3/month | 552 (21%) |
| <1/month | 175 (6.6%) |
| Never | 217 (8.2%) |
| (Missing) | 26 (1.0%) |
| **Year of birth** |  |
| 1998-2005; age 18-24 | 461 (17%) |
| 1993-1998; age 25-29 | 364 (14%) |
| 1983-1993; age 30-39 | 655 (25%) |
| 1973-1983; age 40-49 | 522 (20%) |
| 1963-1973; age 50-59 | 309 (12%) |
| 1953-1963; age 60-69 | 195 (7.4%) |
| 1943-1953; age 70-79 | 120 (4.5%) |
| 1943 or earlier; age 80+ | 17 (0.6%) |
| (Missing) | 9 (0.3%) |
| **Gender** |  |
| Male | 1,288 (49%) |
| Female | 1,356 (51%) |
| Other | 2 (<0.1%) |
| (Missing) | 4 (0.2%) |
| **Religious affiliation** |  |
| Christianity | 2,323 (88%) |
| Islam | 52 (2.0%) |
| Hinduism | 2 (<0.1%) |
| Buddhism | 11 (0.4%) |
| Judaism | 0 (0%) |
| Sikhism | 0 (0%) |
| Baha'i | 0 (0%) |
| Jainism | 0 (0%) |
| Shinto | 2 (<0.1%) |
| Taoism | 1 (<0.1%) |
| Confucianism | 0 (0%) |
| Primal, Animist, or Folk religion | 117 (4.4%) |
| Spiritism | 0 (0%) |
| Umbanda, Candomble, and other African-derived religions | 0 (0%) |
| Chinese folk/traditional religion | 0 (0%) |
| Some other religion | 7 (0.3%) |
| No religion/Atheist/Agnostic | 107 (4.1%) |
| (Missing) | 27 (1.0%) |
| **Race/Ethnicity** |  |
| Asian/Indian | 6 (0.2%) |
| Black | 2,381 (90%) |
| Colored | 252 (9.5%) |
| Other | 1 (<0.1%) |
| White | 8 (0.3%) |
| (Missing) | 3 (0.1%) |
| ^1^n (%) | |

Table S16b. Regression of Community Participation on Childhood Predictors for South Africa

|  | | Secular Community Participation | | | |  | Religious Service Attendance | | | | | |
| --- | --- | --- | --- | --- | --- | --- | --- | --- | --- | --- | --- | --- |
| Variable | Category | Risk-Ratio | RR 95% CI | log(RR) SE | Global  p-value | | |  | Risk-Ratio | RR 95% CI | log(RR) SE | Global  p-value |
| Relationship with mother | (Ref: Very bad/somewhat bad) |  |  |  | 0.849 | | |  |  |  |  | 0.232 |
|  | Very good/somewhat good | 1.03 | (0.66,1.63) | 0.23 |  | | |  | 0.85 | (0.65,1.12) | 0.14 |  |
| Relationship with father | (Ref: Very bad/somewhat bad) |  |  |  | 0.810 | | |  |  |  |  | 0.546 |
|  | Very good/somewhat good | 1.00 | (0.77,1.29) | 0.13 |  | | |  | 1.05 | (0.89,1.24) | 0.08 |  |
| Parent marital status | (Ref: Parents married) |  |  |  | 0.276 | | |  |  |  |  | 0.006 |
|  | Divorced | 1.35 | (0.98,1.87) | 0.17 |  | | |  | 0.87 | (0.69,1.10) | 0.12 |  |
|  | Parents were never married | 1.02 | (0.86,1.22) | 0.09 |  | | |  | 0.82 | (0.73,0.93) | 0.06 |  |
|  | One or both parents had died | 0.94 | (0.61,1.43) | 0.22 |  | | |  | 0.88 | (0.69,1.11) | 0.12 |  |
| Subjective financial status of family growing up | (Ref: Got by) |  |  |  | 0.703 | | |  |  |  |  | 0.952 |
|  | Lived comfortably | 1.07 | (0.90,1.27) | 0.09 |  | | |  | 1.03 | (0.92,1.14) | 0.06 |  |
|  | Found it difficult | 0.92 | (0.69,1.23) | 0.15 |  | | |  | 0.99 | (0.85,1.16) | 0.08 |  |
|  | Found it very difficult | 1.02 | (0.74,1.41) | 0.16 |  | | |  | 1.02 | (0.84,1.23) | 0.10 |  |
| Experienced Abuse | (Ref: No) |  |  |  | <.001 | | |  |  |  |  | 0.046 |
|  | Yes | 1.53 | (1.24,1.90) | 0.11 |  | | |  | 0.85 | (0.72,1.00) | 0.08 |  |
| Felt like an outsider in the family | (Ref: No) |  |  |  | 0.558 | | |  |  |  |  | 0.656 |
|  | Yes | 1.09 | (0.82,1.44) | 0.14 |  | | |  | 0.97 | (0.86,1.10) | 0.06 |  |
| Self-rated health growing up | (Ref: Good) |  |  |  | 0.111 | | |  |  |  |  | 0.376 |
|  | Excellent | 0.85 | (0.67,1.08) | 0.12 |  | | |  | 1.11 | (0.96,1.29) | 0.08 |  |
|  | Very good | 0.84 | (0.63,1.12) | 0.15 |  | | |  | 1.16 | (0.99,1.37) | 0.08 |  |
|  | Fair | 0.59 | (0.40,0.87) | 0.19 |  | | |  | 1.10 | (0.91,1.33) | 0.10 |  |
|  | Poor | 0.90 | (0.55,1.47) | 0.25 |  | | |  | 1.23 | (0.96,1.57) | 0.13 |  |
| Immigration status | (Ref: Born in this country) |  |  |  | 0.087 | | |  |  |  |  | 0.036 |
|  | Born in another country | 0.62 | (0.36,1.07) | 0.27 |  | | |  | 1.23 | (1.01,1.49) | 0.10 |  |
| Religious service attendance (age 12) | (Ref: Never) |  |  |  | 0.324 | | |  |  |  |  | <.001 |
|  | At least 1/week | 1.01 | (0.65,1.59) | 0.23 |  | | |  | 1.09 | (0.83,1.42) | 0.14 |  |
|  | 1-3/month | 1.04 | (0.65,1.68) | 0.24 |  | | |  | 0.89 | (0.68,1.16) | 0.14 |  |
|  | < 1/month | 0.74 | (0.42,1.30) | 0.29 |  | | |  | 0.60 | (0.41,0.87) | 0.19 |  |
| Year of birth | (Ref: 1998-2005; current age: 18-24) |  |  |  | <.001 | | |  |  |  |  | 0.001 |
|  | 1993-1998; age 25-29 | 0.73 | (0.58,0.93) | 0.12 |  | | |  | 0.85 | (0.71,1.01) | 0.09 |  |
|  | 1983-1993; age 30-39 | 0.65 | (0.52,0.82) | 0.12 |  | | |  | 0.85 | (0.72,1.00) | 0.08 |  |
|  | 1973-1983; age 40-49 | 0.58 | (0.44,0.76) | 0.14 |  | | |  | 0.91 | (0.77,1.07) | 0.08 |  |
|  | 1963-1973; age 50-59 | 0.51 | (0.36,0.72) | 0.18 |  | | |  | 1.09 | (0.92,1.29) | 0.09 |  |
|  | 1953-1963; age 60-69 | 0.42 | (0.24,0.73) | 0.28 |  | | |  | 1.10 | (0.91,1.34) | 0.10 |  |
|  | 1943-1953; age 70-79 | 0.74 | (0.41,1.32) | 0.30 |  | | |  | 1.22 | (0.96,1.54) | 0.12 |  |
|  | 1943 or earlier; age 80+ | 1.80 | (0.78,4.16) | 0.43 |  | | |  | 0.81 | (0.34,1.92) | 0.44 |  |
| Gender | (Ref: Male) |  |  |  | <.001 | | |  |  |  |  | <.001 |
|  | Female | 0.84 | (0.71,0.98) | 0.08 |  | | |  | 1.47 | (1.33,1.63) | 0.05 |  |
|  | Other | 5.67 | (4.19,7.68) | 0.15 |  | | |  | 1.78 | (1.19,2.67) | 0.21 |  |
| Religious affiliation | (Ref: No religion/Atheist/Agnostic) |  |  |  | 0.680 | | |  |  |  |  | 0.048 |
|  | Primal, Animist, or Folk religion | 0.76 | (0.33,1.76) | 0.43 |  | | |  | 1.27 | (0.79,2.05) | 0.24 |  |
|  | Christianity | 1.03 | (0.56,1.92) | 0.32 |  | | |  | 1.50 | (1.00,2.28) | 0.21 |  |
|  | Collapsed affiliations with prevalence<3% | 0.88 | (0.32,2.44) | 0.52 |  | | |  | 1.04 | (0.59,1.82) | 0.29 |  |
| Race/ethnicity | (Ref: Plurality group) |  |  |  | 0.721 | | |  |  |  |  | 0.776 |
|  | Non-plurality groups | 1.05 | (0.78,1.41) | 0.15 |  | | |  | 1.03 | (0.85,1.25) | 0.10 |  |

Table S16c. Sensitivity of the regression estimates to unmeasured confounding for South Africa

|  | | Secular Community Participation | |  | Religious Service Attendance | |
| --- | --- | --- | --- | --- | --- | --- |
| Variable | Category | E-value for Estimate | E-value for 95% CI |  | E-value for Estimate | E-value for 95% CI |
| Relationship with mother | (Ref: Very bad/somewhat bad) |  |  |  |  |  |
|  | Very good/somewhat good | 1.22 | 1.00 |  | 1.63 | 1.00 |
| Relationship with father | (Ref: Very bad/somewhat bad) |  |  |  |  |  |
|  | Very good/somewhat good | 1.03 | 1.00 |  | 1.28 | 1.00 |
| Parent marital status | (Ref: Parents married) |  |  |  |  |  |
|  | Divorced | 2.04 | 1.00 |  | 1.57 | 1.00 |
|  | Parents were never married | 1.17 | 1.00 |  | 1.72 | 1.38 |
|  | One or both parents had died | 1.34 | 1.00 |  | 1.54 | 1.00 |
| Subjective financial status of family growing up | (Ref: Got by) |  |  |  |  |  |
|  | Lived comfortably | 1.34 | 1.00 |  | 1.20 | 1.00 |
|  | Found it difficult | 1.41 | 1.00 |  | 1.10 | 1.00 |
|  | Found it very difficult | 1.17 | 1.00 |  | 1.15 | 1.00 |
| Experienced Abuse | (Ref: No) |  |  |  |  |  |
|  | Yes | 2.44 | 1.78 |  | 1.63 | 1.03 |
| Felt like an outsider in the family | (Ref: No) |  |  |  |  |  |
|  | Yes | 1.39 | 1.00 |  | 1.20 | 1.00 |
| Self-rated health growing up | (Ref: Good) |  |  |  |  |  |
|  | Excellent | 1.63 | 1.00 |  | 1.46 | 1.00 |
|  | Very good | 1.67 | 1.00 |  | 1.60 | 1.00 |
|  | Fair | 2.76 | 1.57 |  | 1.43 | 1.00 |
|  | Poor | 1.46 | 1.00 |  | 1.76 | 1.00 |
| Immigration status | (Ref: Born in this country) |  |  |  |  |  |
|  | Born in another country | 2.59 | 1.00 |  | 1.76 | 1.14 |
| Religious service attendance (age 12) | (Ref: Never) |  |  |  |  |  |
|  | At least 1/week | 1.13 | 1.00 |  | 1.40 | 1.00 |
|  | 1-3/month | 1.26 | 1.00 |  | 1.51 | 1.00 |
|  | < 1/month | 2.05 | 1.00 |  | 2.75 | 1.58 |
| Year of birth | (Ref: 1998-2005; current age: 18-24) |  |  |  |  |  |
|  | 1993-1998; age 25-29 | 2.06 | 1.35 |  | 1.64 | 1.00 |
|  | 1983-1993; age 30-39 | 2.44 | 1.74 |  | 1.63 | 1.02 |
|  | 1973-1983; age 40-49 | 2.85 | 1.95 |  | 1.44 | 1.00 |
|  | 1963-1973; age 50-59 | 3.34 | 2.13 |  | 1.40 | 1.00 |
|  | 1953-1963; age 60-69 | 4.17 | 2.06 |  | 1.44 | 1.00 |
|  | 1943-1953; age 70-79 | 2.06 | 1.00 |  | 1.73 | 1.00 |
|  | 1943 or earlier; age 80+ | 3.00 | 1.00 |  | 1.79 | 1.00 |
| Gender | (Ref: Male) |  |  |  |  |  |
|  | Female | 1.68 | 1.14 |  | 2.31 | 2.00 |
|  | Other | 10.81 | 7.84 |  | 2.97 | 1.67 |
| Religious affiliation | (Ref: No religion/Atheist/Agnostic) |  |  |  |  |  |
|  | Primal, Animist, or Folk religion | 1.94 | 1.00 |  | 1.86 | 1.00 |
|  | Christianity | 1.21 | 1.00 |  | 2.38 | 1.00 |
|  | Collapsed affiliations with prevalence<3% | 1.52 | 1.00 |  | 1.24 | 1.00 |
| Race/ethnicity | (Ref: Plurality group) |  |  |  |  |  |
|  | Non-plurality groups | 1.28 | 1.00 |  | 1.20 | 1.00 |

Table S17a. Nationally representative descriptive statistics for Spain

| **Characteristic** | **N = 6,290**^1^ |
| --- | --- |
| **Relationship with mother** |  |
| Very good | 4,557 (72%) |
| Somewhat good | 1,258 (20%) |
| Somewhat bad | 248 (3.9%) |
| Very bad | 92 (1.5%) |
| Does not apply | 107 (1.7%) |
| (Missing) | 28 (0.4%) |
| **Relationship with father** |  |
| Very good | 4,131 (66%) |
| Somewhat good | 1,397 (22%) |
| Somewhat bad | 309 (4.9%) |
| Very bad | 178 (2.8%) |
| Does not apply | 243 (3.9%) |
| (Missing) | 33 (0.5%) |
| **Parent marital status** |  |
| Parents married | 5,285 (84%) |
| Divorced | 378 (6.0%) |
| Parents were never married | 312 (5.0%) |
| One or both parents had died | 126 (2.0%) |
| (Missing) | 188 (3.0%) |
| **Subjective financial status of family growing up** |  |
| Lived comfortably | 2,041 (32%) |
| Got by | 2,956 (47%) |
| Found it difficult | 1,154 (18%) |
| Found it very difficult | 110 (1.7%) |
| (Missing) | 29 (0.5%) |
| **Experienced Abuse** |  |
| Yes | 659 (10%) |
| No | 5,510 (88%) |
| (Missing) | 122 (1.9%) |
| **Felt like an outsider in the family** |  |
| Yes | 579 (9.2%) |
| No | 5,637 (90%) |
| (Missing) | 75 (1.2%) |
| **Self-rated health growing up** |  |
| Excellent | 2,450 (39%) |
| Very good | 2,286 (36%) |
| Good | 1,235 (20%) |
| Fair | 164 (2.6%) |
| Poor | 135 (2.1%) |
| (Missing) | 20 (0.3%) |
| **Immigration status** |  |
| Born in this country | 5,479 (87%) |
| Born in another country | 788 (13%) |
| (Missing) | 23 (0.4%) |
| **Religious service attendance (age 12)** |  |
| At least 1/week | 2,391 (38%) |
| 1-3/month | 1,132 (18%) |
| <1/month | 1,287 (20%) |
| Never | 1,445 (23%) |
| (Missing) | 36 (0.6%) |
| **Year of birth** |  |
| 1998-2005; age 18-24 | 594 (9.4%) |
| 1993-1998; age 25-29 | 450 (7.2%) |
| 1983-1993; age 30-39 | 1,111 (18%) |
| 1973-1983; age 40-49 | 1,396 (22%) |
| 1963-1973; age 50-59 | 1,252 (20%) |
| 1953-1963; age 60-69 | 977 (16%) |
| 1943-1953; age 70-79 | 467 (7.4%) |
| 1943 or earlier; age 80+ | 43 (0.7%) |
| (Missing) | 0 (0%) |
| **Gender** |  |
| Male | 3,142 (50%) |
| Female | 3,119 (50%) |
| Other | 6 (0.1%) |
| (Missing) | 22 (0.4%) |
| **Religious affiliation** |  |
| Christianity | 5,119 (81%) |
| Islam | 132 (2.1%) |
| Hinduism | 5 (<0.1%) |
| Buddhism | 8 (0.1%) |
| Judaism | 5 (<0.1%) |
| Sikhism | 2 (<0.1%) |
| Baha'i | 0 (0%) |
| Jainism | 0 (0%) |
| Shinto | 0 (0%) |
| Taoism | 0 (0%) |
| Confucianism | 1 (<0.1%) |
| Primal, Animist, or Folk religion | 4 (<0.1%) |
| Spiritism | 0 (0%) |
| Umbanda, Candomble, and other African-derived religions | 0 (0%) |
| Chinese folk/traditional religion | 0 (0%) |
| Some other religion | 13 (0.2%) |
| No religion/Atheist/Agnostic | 972 (15%) |
| (Missing) | 29 (0.5%) |
| ^1^n (%) | |

Table S17b. Regression of Community Participation on Childhood Predictors for Spain

|  | | Secular Community Participation | | | |  | Religious Service Attendance | | | |
| --- | --- | --- | --- | --- | --- | --- | --- | --- | --- | --- |
| Variable | Category | Risk-Ratio | RR 95% CI | log(RR) SE | Global  p-value |  | Risk-Ratio | RR 95% CI | log(RR) SE | Global p-value |
| Relationship with mother | (Ref: Very bad/somewhat bad) |  |  |  | 0.020 |  |  |  |  | 0.163 |
|  | Very good/somewhat good | 1.34 | (1.04,1.73) | 0.13 |  |  | 1.25 | (0.90,1.72) | 0.16 |  |
| Relationship with father | (Ref: Very bad/somewhat bad) |  |  |  | 0.351 |  |  |  |  | 0.010 |
|  | Very good/somewhat good | 1.10 | (0.90,1.34) | 0.10 |  |  | 1.41 | (1.08,1.84) | 0.14 |  |
| Parent marital status | (Ref: Parents married) |  |  |  | <.001 |  |  |  |  | 0.038 |
|  | Divorced | 1.36 | (1.11,1.66) | 0.10 |  |  | 1.44 | (1.09,1.90) | 0.14 |  |
|  | Parents were never married | 1.17 | (0.92,1.50) | 0.12 |  |  | 1.22 | (0.96,1.54) | 0.12 |  |
|  | One or both parents had died | 1.67 | (1.19,2.33) | 0.17 |  |  | 0.97 | (0.60,1.57) | 0.24 |  |
| Subjective financial status of family growing up | (Ref: Got by) |  |  |  | 0.688 |  |  |  |  | 0.040 |
|  | Lived comfortably | 1.05 | (0.91,1.21) | 0.07 |  |  | 1.26 | (1.05,1.51) | 0.09 |  |
|  | Found it difficult | 0.98 | (0.82,1.17) | 0.09 |  |  | 0.97 | (0.76,1.24) | 0.12 |  |
|  | Found it very difficult | 0.83 | (0.52,1.34) | 0.24 |  |  | 1.08 | (0.63,1.85) | 0.28 |  |
| Experienced Abuse | (Ref: No) |  |  |  | 0.412 |  |  |  |  | 0.617 |
|  | Yes | 0.93 | (0.77,1.11) | 0.09 |  |  | 0.95 | (0.75,1.19) | 0.12 |  |
| Felt like an outsider in the family | (Ref: No) |  |  |  | 0.001 |  |  |  |  | 0.003 |
|  | Yes | 1.32 | (1.11,1.58) | 0.09 |  |  | 1.32 | (1.09,1.59) | 0.09 |  |
| Self-rated health growing up | (Ref: Good) |  |  |  | 0.061 |  |  |  |  | 0.153 |
|  | Excellent | 1.12 | (0.93,1.35) | 0.09 |  |  | 1.03 | (0.83,1.27) | 0.11 |  |
|  | Very good | 1.27 | (1.05,1.53) | 0.10 |  |  | 1.21 | (0.97,1.52) | 0.11 |  |
|  | Fair | 1.06 | (0.70,1.63) | 0.22 |  |  | 0.66 | (0.32,1.35) | 0.36 |  |
|  | Poor | 1.48 | (0.99,2.22) | 0.21 |  |  | 1.21 | (0.71,2.07) | 0.27 |  |
| Immigration status | (Ref: Born in this country) |  |  |  | 0.019 |  |  |  |  | <.001 |
|  | Born in another country | 0.82 | (0.70,0.97) | 0.08 |  |  | 1.47 | (1.24,1.73) | 0.08 |  |
| Religious service attendance (age 12) | (Ref: Never) |  |  |  | <.001 |  |  |  |  | <.001 |
|  | At least 1/week | 1.41 | (1.16,1.70) | 0.10 |  |  | 11.72 | (7.44,18.46) | 0.23 |  |
|  | 1-3/month | 1.49 | (1.23,1.81) | 0.10 |  |  | 10.93 | (6.91,17.31) | 0.23 |  |
|  | < 1/month | 1.17 | (0.95,1.45) | 0.11 |  |  | 2.69 | (1.56,4.65) | 0.28 |  |
| Year of birth | (Ref: 1998-2005; current age: 18-24) |  |  |  | <.001 |  |  |  |  | 0.191 |
|  | 1993-1998; age 25-29 | 1.05 | (0.85,1.30) | 0.11 |  |  | 0.94 | (0.72,1.24) | 0.14 |  |
|  | 1983-1993; age 30-39 | 0.80 | (0.66,0.96) | 0.10 |  |  | 0.88 | (0.69,1.11) | 0.12 |  |
|  | 1973-1983; age 40-49 | 0.73 | (0.60,0.88) | 0.10 |  |  | 0.81 | (0.63,1.03) | 0.12 |  |
|  | 1963-1973; age 50-59 | 0.65 | (0.52,0.80) | 0.11 |  |  | 0.79 | (0.61,1.03) | 0.13 |  |
|  | 1953-1963; age 60-69 | 0.67 | (0.51,0.88) | 0.14 |  |  | 0.76 | (0.55,1.04) | 0.16 |  |
|  | 1943-1953; age 70-79 | 0.69 | (0.48,0.98) | 0.18 |  |  | 0.87 | (0.57,1.34) | 0.22 |  |
|  | 1943 or earlier; age 80+ | 0.50 | (0.20,1.29) | 0.48 |  |  | 1.64 | (0.87,3.11) | 0.32 |  |
| Gender | (Ref: Male) |  |  |  | 0.105 |  |  |  |  | 0.007 |
|  | Female | 0.89 | (0.79,1.00) | 0.06 |  |  | 0.78 | (0.67,0.91) | 0.08 |  |
|  | Other | 1.47 | (0.55,3.93) | 0.50 |  |  | 0.93 | (0.27,3.19) | 0.63 |  |
| Religious affiliation | (Ref: No religion/Atheist/Agnostic) |  |  |  | <.001 |  |  |  |  | <.001 |
|  | Christianity | 1.04 | (0.86,1.25) | 0.09 |  |  | 1.26 | (0.95,1.68) | 0.15 |  |
|  | Collapsed affiliations with prevalence<3% | 1.82 | (1.40,2.37) | 0.13 |  |  | 2.11 | (1.48,2.99) | 0.18 |  |

Table S17c. Sensitivity of the regression estimates to unmeasured confounding for Spain

|  | | Secular Community Participation | |  | Religious Service Attendance | |
| --- | --- | --- | --- | --- | --- | --- |
| Variable | Category | E-value for Estimate | E-value for 95% CI |  | E-value for Estimate | E-value for 95% CI |
| Relationship with mother | (Ref: Very bad/somewhat bad) |  |  |  |  |  |
|  | Very good/somewhat good | 2.02 | 1.25 |  | 1.80 | 1.00 |
| Relationship with father | (Ref: Very bad/somewhat bad) |  |  |  |  |  |
|  | Very good/somewhat good | 1.42 | 1.00 |  | 2.17 | 1.37 |
| Parent marital status | (Ref: Parents married) |  |  |  |  |  |
|  | Divorced | 2.05 | 1.45 |  | 2.23 | 1.39 |
|  | Parents were never married | 1.63 | 1.00 |  | 1.73 | 1.00 |
|  | One or both parents had died | 2.72 | 1.67 |  | 1.19 | 1.00 |
| Subjective financial status of family growing up | (Ref: Got by) |  |  |  |  |  |
|  | Lived comfortably | 1.28 | 1.00 |  | 1.83 | 1.29 |
|  | Found it difficult | 1.18 | 1.00 |  | 1.20 | 1.00 |
|  | Found it very difficult | 1.69 | 1.00 |  | 1.36 | 1.00 |
| Experienced Abuse | (Ref: No) |  |  |  |  |  |
|  | Yes | 1.37 | 1.00 |  | 1.31 | 1.00 |
| Felt like an outsider in the family | (Ref: No) |  |  |  |  |  |
|  | Yes | 1.98 | 1.47 |  | 1.96 | 1.41 |
| Self-rated health growing up | (Ref: Good) |  |  |  |  |  |
|  | Excellent | 1.49 | 1.00 |  | 1.20 | 1.00 |
|  | Very good | 1.85 | 1.28 |  | 1.72 | 1.00 |
|  | Fair | 1.33 | 1.00 |  | 2.39 | 1.00 |
|  | Poor | 2.33 | 1.00 |  | 1.72 | 1.00 |
| Immigration status | (Ref: Born in this country) |  |  |  |  |  |
|  | Born in another country | 1.73 | 1.22 |  | 2.30 | 1.80 |
| Religious service attendance (age 12) | (Ref: Never) |  |  |  |  |  |
|  | At least 1/week | 2.16 | 1.60 |  | 22.92 | 14.36 |
|  | 1-3/month | 2.35 | 1.76 |  | 21.36 | 13.29 |
|  | < 1/month | 1.62 | 1.00 |  | 4.82 | 2.49 |
| Year of birth | (Ref: 1998-2005; current age: 18-24) |  |  |  |  |  |
|  | 1993-1998; age 25-29 | 1.29 | 1.00 |  | 1.31 | 1.00 |
|  | 1983-1993; age 30-39 | 1.82 | 1.23 |  | 1.54 | 1.00 |
|  | 1973-1983; age 40-49 | 2.08 | 1.52 |  | 1.78 | 1.00 |
|  | 1963-1973; age 50-59 | 2.46 | 1.81 |  | 1.84 | 1.00 |
|  | 1953-1963; age 60-69 | 2.35 | 1.54 |  | 1.98 | 1.00 |
|  | 1943-1953; age 70-79 | 2.27 | 1.15 |  | 1.55 | 1.00 |
|  | 1943 or earlier; age 80+ | 3.38 | 1.00 |  | 2.67 | 1.00 |
| Gender | (Ref: Male) |  |  |  |  |  |
|  | Female | 1.50 | 1.00 |  | 1.87 | 1.42 |
|  | Other | 2.31 | 1.00 |  | 1.35 | 1.00 |
| Religious affiliation | (Ref: No religion/Atheist/Agnostic) |  |  |  |  |  |
|  | Christianity | 1.23 | 1.00 |  | 1.84 | 1.00 |
|  | Collapsed affiliations with prevalence<3% | 3.04 | 2.14 |  | 3.63 | 2.33 |

Table S18a. Nationally representative descriptive statistics for Sweden

| **Characteristic** | **N = 15,068**^1^ |
| --- | --- |
| **Relationship with mother** |  |
| Very good | 8,743 (58%) |
| Somewhat good | 4,513 (30%) |
| Somewhat bad | 1,194 (7.9%) |
| Very bad | 371 (2.5%) |
| Does not apply | 216 (1.4%) |
| (Missing) | 30 (0.2%) |
| **Relationship with father** |  |
| Very good | 7,134 (47%) |
| Somewhat good | 4,885 (32%) |
| Somewhat bad | 1,588 (11%) |
| Very bad | 725 (4.8%) |
| Does not apply | 720 (4.8%) |
| (Missing) | 16 (0.1%) |
| **Parent marital status** |  |
| Parents married | 10,887 (72%) |
| Divorced | 1,927 (13%) |
| Parents were never married | 1,747 (12%) |
| One or both parents had died | 362 (2.4%) |
| (Missing) | 145 (1.0%) |
| **Subjective financial status of family growing up** |  |
| Lived comfortably | 5,951 (39%) |
| Got by | 7,717 (51%) |
| Found it difficult | 1,238 (8.2%) |
| Found it very difficult | 140 (0.9%) |
| (Missing) | 22 (0.1%) |
| **Experienced Abuse** |  |
| Yes | 2,288 (15%) |
| No | 12,735 (85%) |
| (Missing) | 45 (0.3%) |
| **Felt like an outsider in the family** |  |
| Yes | 1,867 (12%) |
| No | 13,034 (86%) |
| (Missing) | 168 (1.1%) |
| **Self-rated health growing up** |  |
| Excellent | 5,733 (38%) |
| Very good | 5,124 (34%) |
| Good | 2,669 (18%) |
| Fair | 1,108 (7.4%) |
| Poor | 397 (2.6%) |
| (Missing) | 38 (0.2%) |
| **Immigration status** |  |
| Born in this country | 13,922 (92%) |
| Born in another country | 1,052 (7.0%) |
| (Missing) | 94 (0.6%) |
| **Religious service attendance (age 12)** |  |
| At least 1/week | 955 (6.3%) |
| 1-3/month | 1,362 (9.0%) |
| <1/month | 6,224 (41%) |
| Never | 6,472 (43%) |
| (Missing) | 54 (0.4%) |
| **Year of birth** |  |
| 1998-2005; age 18-24 | 1,515 (10%) |
| 1993-1998; age 25-29 | 1,399 (9.3%) |
| 1983-1993; age 30-39 | 2,398 (16%) |
| 1973-1983; age 40-49 | 2,221 (15%) |
| 1963-1973; age 50-59 | 2,493 (17%) |
| 1953-1963; age 60-69 | 2,168 (14%) |
| 1943-1953; age 70-79 | 2,253 (15%) |
| 1943 or earlier; age 80+ | 621 (4.1%) |
| (Missing) | 0 (0%) |
| **Gender** |  |
| Male | 7,536 (50%) |
| Female | 7,493 (50%) |
| Other | 27 (0.2%) |
| (Missing) | 12 (<0.1%) |
| **Religious affiliation** |  |
| Christianity | 10,617 (70%) |
| Islam | 462 (3.1%) |
| Hinduism | 16 (0.1%) |
| Buddhism | 41 (0.3%) |
| Judaism | 51 (0.3%) |
| Sikhism | 9 (<0.1%) |
| Baha'i | 3 (<0.1%) |
| Jainism | 0 (0%) |
| Shinto | 1 (<0.1%) |
| Taoism | 0 (0%) |
| Confucianism | 4 (<0.1%) |
| Primal, Animist, or Folk religion | 31 (0.2%) |
| Spiritism | 0 (0%) |
| Umbanda, Candomble, and other African-derived religions | 0 (0%) |
| Chinese folk/traditional religion | 0 (0%) |
| Some other religion | 69 (0.5%) |
| No religion/Atheist/Agnostic | 3,738 (25%) |
| (Missing) | 26 (0.2%) |
| ^1^n (%) | |

Table S18b. Regression of Community Participation on Childhood Predictors for Sweden

|  | | Secular Community Participation | | | |  | Religious Service Attendance | | | |
| --- | --- | --- | --- | --- | --- | --- | --- | --- | --- | --- |
| Variable | Category | Risk-Ratio | RR 95% CI | log(RR) SE | Global  p-value |  | Risk-Ratio | RR 95% CI | log(RR) SE | Global  p-value |
| Relationship with mother | (Ref: Very bad/somewhat bad) |  |  |  | 0.566 |  |  |  |  | 0.285 |
|  | Very good/somewhat good | 1.05 | (0.89,1.24) | 0.08 |  |  | 1.25 | (0.83,1.88) | 0.21 |  |
| Relationship with father | (Ref: Very bad/somewhat bad) |  |  |  | 0.172 |  |  |  |  | 0.120 |
|  | Very good/somewhat good | 1.10 | (0.96,1.26) | 0.07 |  |  | 1.31 | (0.93,1.86) | 0.18 |  |
| Parent marital status | (Ref: Parents married) |  |  |  | 0.261 |  |  |  |  | 0.909 |
|  | Divorced | 1.05 | (0.92,1.20) | 0.07 |  |  | 0.89 | (0.65,1.23) | 0.16 |  |
|  | Parents were never married | 0.89 | (0.77,1.03) | 0.07 |  |  | 0.97 | (0.67,1.40) | 0.19 |  |
|  | One or both parents had died | 0.92 | (0.66,1.29) | 0.17 |  |  | 0.92 | (0.40,2.07) | 0.42 |  |
| Subjective financial status of family growing up | (Ref: Got by) |  |  |  | 0.034 |  |  |  |  | 0.841 |
|  | Lived comfortably | 1.11 | (1.01,1.21) | 0.05 |  |  | 0.98 | (0.80,1.19) | 0.10 |  |
|  | Found it difficult | 1.07 | (0.90,1.28) | 0.09 |  |  | 0.85 | (0.59,1.24) | 0.19 |  |
|  | Found it very difficult | 1.55 | (1.03,2.35) | 0.21 |  |  | 0.84 | (0.39,1.82) | 0.39 |  |
| Experienced Abuse | (Ref: No) |  |  |  | 0.419 |  |  |  |  | 0.007 |
|  | Yes | 1.05 | (0.93,1.20) | 0.07 |  |  | 1.38 | (1.09,1.74) | 0.12 |  |
| Felt like an outsider in the family | (Ref: No) |  |  |  | 0.649 |  |  |  |  | 0.469 |
|  | Yes | 1.03 | (0.89,1.20) | 0.08 |  |  | 1.10 | (0.84,1.44) | 0.14 |  |
| Self-rated health growing up | (Ref: Good) |  |  |  | <.001 |  |  |  |  | 0.153 |
|  | Excellent | 1.28 | (1.12,1.45) | 0.07 |  |  | 0.92 | (0.70,1.20) | 0.14 |  |
|  | Very good | 1.14 | (1.00,1.29) | 0.07 |  |  | 0.99 | (0.76,1.29) | 0.13 |  |
|  | Fair | 0.77 | (0.62,0.95) | 0.11 |  |  | 0.63 | (0.40,0.98) | 0.23 |  |
|  | Poor | 0.76 | (0.55,1.06) | 0.17 |  |  | 0.61 | (0.32,1.17) | 0.33 |  |
| Immigration status | (Ref: Born in this country) |  |  |  | 0.790 |  |  |  |  | 0.190 |
|  | Born in another country | 0.98 | (0.83,1.16) | 0.09 |  |  | 1.18 | (0.92,1.51) | 0.13 |  |
| Religious service attendance (age 12) | (Ref: Never) |  |  |  | <.001 |  |  |  |  | <.001 |
|  | At least 1/week | 1.41 | (1.21,1.65) | 0.08 |  |  | 34.34 | (22.94,51.40) | 0.21 |  |
|  | 1-3/month | 1.33 | (1.15,1.53) | 0.07 |  |  | 12.52 | (8.19,19.12) | 0.22 |  |
|  | < 1/month | 1.17 | (1.07,1.28) | 0.05 |  |  | 2.20 | (1.42,3.41) | 0.22 |  |
| Year of birth | (Ref: 1998-2005; current age: 18-24) |  |  |  | <.001 |  |  |  |  | <.001 |
|  | 1993-1998; age 25-29 | 0.64 | (0.53,0.75) | 0.09 |  |  | 0.89 | (0.67,1.20) | 0.15 |  |
|  | 1983-1993; age 30-39 | 0.63 | (0.55,0.74) | 0.08 |  |  | 0.67 | (0.49,0.91) | 0.16 |  |
|  | 1973-1983; age 40-49 | 0.73 | (0.63,0.85) | 0.08 |  |  | 0.66 | (0.48,0.91) | 0.16 |  |
|  | 1963-1973; age 50-59 | 0.61 | (0.52,0.71) | 0.08 |  |  | 0.65 | (0.47,0.90) | 0.17 |  |
|  | 1953-1963; age 60-69 | 0.57 | (0.49,0.67) | 0.08 |  |  | 0.51 | (0.35,0.73) | 0.19 |  |
|  | 1943-1953; age 70-79 | 0.76 | (0.65,0.89) | 0.08 |  |  | 0.47 | (0.33,0.67) | 0.18 |  |
|  | 1943 or earlier; age 80+ | 0.90 | (0.74,1.10) | 0.10 |  |  | 0.42 | (0.26,0.67) | 0.24 |  |
| Gender | (Ref: Male) |  |  |  | <.001 |  |  |  |  | 0.017 |
|  | Female | 0.86 | (0.79,0.93) | 0.04 |  |  | 0.78 | (0.65,0.94) | 0.09 |  |
|  | Other | 2.06 | (1.13,3.76) | 0.31 |  |  | 2.20 | (0.39,12.57) | 0.89 |  |
| Religious affiliation | (Ref: No religion/Atheist/Agnostic) |  |  |  | 0.040 |  |  |  |  | <.001 |
|  | Islam | 0.99 | (0.75,1.30) | 0.14 |  |  | 3.28 | (1.95,5.52) | 0.27 |  |
|  | Christianity | 1.16 | (1.04,1.29) | 0.06 |  |  | 1.92 | (1.25,2.95) | 0.22 |  |
|  | Collapsed affiliations with prevalence<3% | 1.01 | (0.66,1.53) | 0.22 |  |  | 1.81 | (0.81,4.04) | 0.41 |  |

Table S18c. Sensitivity of the regression estimates to unmeasured confounding for Sweden

|  | | Secular Community Participation | |  | Religious Service Attendance | |
| --- | --- | --- | --- | --- | --- | --- |
| Variable | Category | E-value for Estimate | E-value for 95% CI |  | E-value for Estimate | E-value for 95% CI |
| Relationship with mother | (Ref: Very bad/somewhat bad) |  |  |  |  |  |
|  | Very good/somewhat good | 1.28 | 1.00 |  | 1.81 | 1.00 |
| Relationship with father | (Ref: Very bad/somewhat bad) |  |  |  |  |  |
|  | Very good/somewhat good | 1.43 | 1.00 |  | 1.96 | 1.00 |
| Parent marital status | (Ref: Parents married) |  |  |  |  |  |
|  | Divorced | 1.28 | 1.00 |  | 1.48 | 1.00 |
|  | Parents were never married | 1.50 | 1.00 |  | 1.21 | 1.00 |
|  | One or both parents had died | 1.39 | 1.00 |  | 1.41 | 1.00 |
| Subjective financial status of family growing up | (Ref: Got by) |  |  |  |  |  |
|  | Lived comfortably | 1.45 | 1.13 |  | 1.18 | 1.00 |
|  | Found it difficult | 1.36 | 1.00 |  | 1.62 | 1.00 |
|  | Found it very difficult | 2.48 | 1.20 |  | 1.66 | 1.00 |
| Experienced Abuse | (Ref: No) |  |  |  |  |  |
|  | Yes | 1.29 | 1.00 |  | 2.11 | 1.41 |
| Felt like an outsider in the family | (Ref: No) |  |  |  |  |  |
|  | Yes | 1.22 | 1.00 |  | 1.44 | 1.00 |
| Self-rated health growing up | (Ref: Good) |  |  |  |  |  |
|  | Excellent | 1.87 | 1.50 |  | 1.40 | 1.00 |
|  | Very good | 1.53 | 1.00 |  | 1.10 | 1.00 |
|  | Fair | 1.92 | 1.27 |  | 2.56 | 1.18 |
|  | Poor | 1.95 | 1.00 |  | 2.64 | 1.00 |
| Immigration status | (Ref: Born in this country) |  |  |  |  |  |
|  | Born in another country | 1.17 | 1.00 |  | 1.64 | 1.00 |
| Religious service attendance (age 12) | (Ref: Never) |  |  |  |  |  |
|  | At least 1/week | 2.18 | 1.71 |  | 68.17 | 45.37 |
|  | 1-3/month | 1.98 | 1.55 |  | 24.52 | 15.87 |
|  | < 1/month | 1.61 | 1.33 |  | 3.82 | 2.19 |
| Year of birth | (Ref: 1998-2005; current age: 18-24) |  |  |  |  |  |
|  | 1993-1998; age 25-29 | 2.52 | 1.98 |  | 1.49 | 1.00 |
|  | 1983-1993; age 30-39 | 2.53 | 2.06 |  | 2.36 | 1.43 |
|  | 1973-1983; age 40-49 | 2.08 | 1.64 |  | 2.38 | 1.42 |
|  | 1963-1973; age 50-59 | 2.66 | 2.17 |  | 2.45 | 1.46 |
|  | 1953-1963; age 60-69 | 2.88 | 2.34 |  | 3.37 | 2.07 |
|  | 1943-1953; age 70-79 | 1.96 | 1.51 |  | 3.66 | 2.34 |
|  | 1943 or earlier; age 80+ | 1.46 | 1.00 |  | 4.20 | 2.36 |
| Gender | (Ref: Male) |  |  |  |  |  |
|  | Female | 1.60 | 1.35 |  | 1.88 | 1.32 |
|  | Other | 3.55 | 1.52 |  | 3.83 | 1.00 |
| Religious affiliation | (Ref: No religion/Atheist/Agnostic) |  |  |  |  |  |
|  | Islam | 1.12 | 1.00 |  | 6.01 | 3.31 |
|  | Christianity | 1.60 | 1.26 |  | 3.24 | 1.80 |
|  | Collapsed affiliations with prevalence<3% | 1.09 | 1.00 |  | 3.03 | 1.00 |

Table S19a. Nationally representative descriptive statistics for Tanzania

| **Characteristic** | **N = 9,075**^1^ |
| --- | --- |
| **Relationship with mother** |  |
| Very good | 7,739 (85%) |
| Somewhat good | 796 (8.8%) |
| Somewhat bad | 84 (0.9%) |
| Very bad | 84 (0.9%) |
| Does not apply | 303 (3.3%) |
| (Missing) | 70 (0.8%) |
| **Relationship with father** |  |
| Very good | 6,831 (75%) |
| Somewhat good | 1,101 (12%) |
| Somewhat bad | 203 (2.2%) |
| Very bad | 247 (2.7%) |
| Does not apply | 550 (6.1%) |
| (Missing) | 142 (1.6%) |
| **Parent marital status** |  |
| Parents married | 6,929 (76%) |
| Divorced | 678 (7.5%) |
| Parents were never married | 751 (8.3%) |
| One or both parents had died | 313 (3.4%) |
| (Missing) | 404 (4.4%) |
| **Subjective financial status of family growing up** |  |
| Lived comfortably | 2,611 (29%) |
| Got by | 2,909 (32%) |
| Found it difficult | 2,679 (30%) |
| Found it very difficult | 814 (9.0%) |
| (Missing) | 61 (0.7%) |
| **Experienced Abuse** |  |
| Yes | 716 (7.9%) |
| No | 8,328 (92%) |
| (Missing) | 32 (0.3%) |
| **Felt like an outsider in the family** |  |
| Yes | 734 (8.1%) |
| No | 8,320 (92%) |
| (Missing) | 22 (0.2%) |
| **Self-rated health growing up** |  |
| Excellent | 2,406 (27%) |
| Very good | 2,036 (22%) |
| Good | 2,946 (32%) |
| Fair | 1,177 (13%) |
| Poor | 456 (5.0%) |
| (Missing) | 54 (0.6%) |
| **Immigration status** |  |
| Born in this country | 9,048 (100%) |
| Born in another country | 25 (0.3%) |
| (Missing) | 1 (<0.1%) |
| **Religious service attendance (age 12)** |  |
| At least 1/week | 5,580 (61%) |
| 1-3/month | 2,383 (26%) |
| <1/month | 333 (3.7%) |
| Never | 595 (6.6%) |
| (Missing) | 184 (2.0%) |
| **Year of birth** |  |
| 1998-2005; age 18-24 | 2,284 (25%) |
| 1993-1998; age 25-29 | 1,349 (15%) |
| 1983-1993; age 30-39 | 2,060 (23%) |
| 1973-1983; age 40-49 | 1,503 (17%) |
| 1963-1973; age 50-59 | 912 (10%) |
| 1953-1963; age 60-69 | 575 (6.3%) |
| 1943-1953; age 70-79 | 297 (3.3%) |
| 1943 or earlier; age 80+ | 93 (1.0%) |
| (Missing) | 2 (<0.1%) |
| **Gender** |  |
| Male | 4,299 (47%) |
| Female | 4,776 (53%) |
| Other | 0 (0%) |
| (Missing) | 0 (0%) |
| **Religious affiliation** |  |
| Christianity | 5,651 (62%) |
| Islam | 3,060 (34%) |
| Hinduism | 0 (0%) |
| Buddhism | 0 (0%) |
| Judaism | 0 (0%) |
| Sikhism | 0 (0%) |
| Baha'i | 1 (<0.1%) |
| Jainism | 0 (0%) |
| Shinto | 0 (0%) |
| Taoism | 0 (0%) |
| Confucianism | 0 (0%) |
| Primal, Animist, or Folk religion | 11 (0.1%) |
| Spiritism | 0 (0%) |
| Umbanda, Candomble, and other African-derived religions | 0 (0%) |
| Chinese folk/traditional religion | 0 (0%) |
| Some other religion | 0 (0%) |
| No religion/Atheist/Agnostic | 345 (3.8%) |
| (Missing) | 7 (<0.1%) |
| **Race/Ethnicity** |  |
| African | 9,060 (100%) |
| Arab | 11 (0.1%) |
| Indian | 3 (<0.1%) |
| (Missing) | 2 (<0.1%) |
| ^1^n (%) | |

Table S19b. Regression of Community Participation on Childhood Predictors for Tanzania

|  | | Secular Community Participation | | | |  | Religious Service Attendance | | | |
| --- | --- | --- | --- | --- | --- | --- | --- | --- | --- | --- |
| Variable | Category | Risk-Ratio | RR 95% CI | log(RR) SE | Global  p-value |  | Risk-Ratio | RR 95% CI | log(RR) SE | Global  p-value |
| Relationship with mother | (Ref: Very bad/somewhat bad) |  |  |  | 0.370 |  |  |  |  | 0.091 |
|  | Very good/somewhat good | 0.87 | (0.64,1.20) | 0.16 |  |  | 1.08 | (0.99,1.18) | 0.05 |  |
| Relationship with father | (Ref: Very bad/somewhat bad) |  |  |  | 0.239 |  |  |  |  | 0.437 |
|  | Very good/somewhat good | 1.16 | (0.90,1.48) | 0.13 |  |  | 1.03 | (0.96,1.10) | 0.03 |  |
| Parent marital status | (Ref: Parents married) |  |  |  | 0.966 |  |  |  |  | 0.044 |
|  | Divorced | 0.97 | (0.77,1.21) | 0.11 |  |  | 0.91 | (0.85,0.98) | 0.04 |  |
|  | Parents were never married | 1.03 | (0.85,1.24) | 0.10 |  |  | 0.96 | (0.90,1.01) | 0.03 |  |
|  | One or both parents had died | 1.02 | (0.75,1.39) | 0.16 |  |  | 0.96 | (0.89,1.05) | 0.04 |  |
| Subjective financial status of family growing up | (Ref: Got by) |  |  |  | 0.229 |  |  |  |  | 0.237 |
|  | Lived comfortably | 1.11 | (0.97,1.26) | 0.07 |  |  | 0.97 | (0.94,1.01) | 0.02 |  |
|  | Found it difficult | 1.00 | (0.87,1.15) | 0.07 |  |  | 0.98 | (0.95,1.02) | 0.02 |  |
|  | Found it very difficult | 0.94 | (0.77,1.14) | 0.10 |  |  | 1.03 | (0.98,1.09) | 0.03 |  |
| Experienced Abuse | (Ref: No) |  |  |  | 0.197 |  |  |  |  | 0.325 |
|  | Yes | 1.13 | (0.94,1.35) | 0.09 |  |  | 0.97 | (0.92,1.03) | 0.03 |  |
| Felt like an outsider in the family | (Ref: No) |  |  |  | 0.120 |  |  |  |  | 0.180 |
|  | Yes | 0.86 | (0.70,1.04) | 0.10 |  |  | 0.96 | (0.90,1.02) | 0.03 |  |
| Self-rated health growing up | (Ref: Good) |  |  |  | 0.019 |  |  |  |  | 0.861 |
|  | Excellent | 1.14 | (0.98,1.33) | 0.08 |  |  | 0.99 | (0.95,1.03) | 0.02 |  |
|  | Very good | 1.08 | (0.93,1.26) | 0.08 |  |  | 1.00 | (0.96,1.04) | 0.02 |  |
|  | Fair | 0.91 | (0.75,1.11) | 0.10 |  |  | 1.01 | (0.96,1.06) | 0.03 |  |
|  | Poor | 1.31 | (1.00,1.73) | 0.14 |  |  | 0.98 | (0.90,1.06) | 0.04 |  |
| Immigration status | (Ref: Born in this country) |  |  |  | 0.061 |  |  |  |  | 0.807 |
|  | Born in another country | 0.32 | (0.10,1.05) | 0.60 |  |  | 0.96 | (0.71,1.31) | 0.16 |  |
| Religious service attendance (age 12) | (Ref: Never) |  |  |  | 0.014 |  |  |  |  | <.001 |
|  | At least 1/week | 0.97 | (0.73,1.29) | 0.15 |  |  | 1.09 | (0.99,1.20) | 0.05 |  |
|  | 1-3/month | 1.19 | (0.88,1.62) | 0.16 |  |  | 1.02 | (0.93,1.13) | 0.05 |  |
|  | < 1/month | 0.94 | (0.64,1.37) | 0.19 |  |  | 0.93 | (0.82,1.06) | 0.07 |  |
| Year of birth | (Ref: 1998-2005; current age: 18-24) |  |  |  | 0.939 |  |  |  |  | 0.002 |
|  | 1993-1998; age 25-29 | 0.94 | (0.80,1.11) | 0.08 |  |  | 1.00 | (0.95,1.05) | 0.03 |  |
|  | 1983-1993; age 30-39 | 0.97 | (0.85,1.11) | 0.07 |  |  | 1.03 | (0.98,1.07) | 0.02 |  |
|  | 1973-1983; age 40-49 | 0.93 | (0.78,1.10) | 0.09 |  |  | 1.03 | (0.98,1.08) | 0.02 |  |
|  | 1963-1973; age 50-59 | 0.91 | (0.77,1.07) | 0.08 |  |  | 1.09 | (1.04,1.15) | 0.03 |  |
|  | 1953-1963; age 60-69 | 0.98 | (0.76,1.26) | 0.13 |  |  | 1.09 | (1.03,1.16) | 0.03 |  |
|  | 1943-1953; age 70-79 | 0.90 | (0.62,1.30) | 0.19 |  |  | 1.12 | (1.02,1.22) | 0.04 |  |
|  | 1943 or earlier; age 80+ | 0.70 | (0.32,1.54) | 0.40 |  |  | 1.14 | (0.97,1.33) | 0.08 |  |
| Gender | (Ref: Male) |  |  |  | <.001 |  |  |  |  | <.001 |
|  | Female | 0.72 | (0.64,0.80) | 0.06 |  |  | 1.08 | (1.04,1.11) | 0.01 |  |
| Religious affiliation | (Ref: No religion/Atheist/Agnostic) |  |  |  | <.001 |  |  |  |  | 0.048 |
|  | Islam | 1.31 | (0.82,2.08) | 0.24 |  |  | 1.13 | (1.00,1.29) | 0.07 |  |
|  | Christianity | 1.17 | (0.75,1.82) | 0.23 |  |  | 1.10 | (0.97,1.25) | 0.07 |  |
|  | Collapsed affiliations with prevalence<3% | 0.00 | (0.00,0.00) | 0.44 |  |  | 0.67 | (0.36,1.25) | 0.32 |  |
| Race/ethnicity | (Ref: Plurality group) |  |  |  | 0.751 |  |  |  |  | <.001 |
|  | Non-plurality groups | 1.14 | (0.51,2.54) | 0.41 |  |  | 1.27 | (1.14,1.42) | 0.06 |  |

Table S19c. Sensitivity of the regression estimates to unmeasured confounding for Tanzania

|  | | Secular Community Participation | |  | Religious Service Attendance | |
| --- | --- | --- | --- | --- | --- | --- |
| Variable | Category | E-value for Estimate | E-value for 95% CI |  | E-value for Estimate | E-value for 95% CI |
| Relationship with mother | (Ref: Very bad/somewhat bad) |  |  |  |  |  |
|  | Very good/somewhat good | 1.55 | 1.00 |  | 1.37 | 1.00 |
| Relationship with father | (Ref: Very bad/somewhat bad) |  |  |  |  |  |
|  | Very good/somewhat good | 1.58 | 1.00 |  | 1.19 | 1.00 |
| Parent marital status | (Ref: Parents married) |  |  |  |  |  |
|  | Divorced | 1.23 | 1.00 |  | 1.43 | 1.16 |
|  | Parents were never married | 1.19 | 1.00 |  | 1.27 | 1.00 |
|  | One or both parents had died | 1.17 | 1.00 |  | 1.24 | 1.00 |
| Subjective financial status of family growing up | (Ref: Got by) |  |  |  |  |  |
|  | Lived comfortably | 1.45 | 1.00 |  | 1.19 | 1.00 |
|  | Found it difficult | 1.04 | 1.00 |  | 1.15 | 1.00 |
|  | Found it very difficult | 1.34 | 1.00 |  | 1.21 | 1.00 |
| Experienced Abuse | (Ref: No) |  |  |  |  |  |
|  | Yes | 1.50 | 1.00 |  | 1.20 | 1.00 |
| Felt like an outsider in the family | (Ref: No) |  |  |  |  |  |
|  | Yes | 1.61 | 1.00 |  | 1.25 | 1.00 |
| Self-rated health growing up | (Ref: Good) |  |  |  |  |  |
|  | Excellent | 1.54 | 1.00 |  | 1.13 | 1.00 |
|  | Very good | 1.38 | 1.00 |  | 1.02 | 1.00 |
|  | Fair | 1.42 | 1.00 |  | 1.10 | 1.00 |
|  | Poor | 1.95 | 1.00 |  | 1.17 | 1.00 |
| Immigration status | (Ref: Born in this country) |  |  |  |  |  |
|  | Born in another country | 5.61 | 1.00 |  | 1.24 | 1.00 |
| Religious service attendance (age 12) | (Ref: Never) |  |  |  |  |  |
|  | At least 1/week | 1.22 | 1.00 |  | 1.42 | 1.00 |
|  | 1-3/month | 1.67 | 1.00 |  | 1.18 | 1.00 |
|  | < 1/month | 1.34 | 1.00 |  | 1.36 | 1.00 |
| Year of birth | (Ref: 1998-2005; current age: 18-24) |  |  |  |  |  |
|  | 1993-1998; age 25-29 | 1.31 | 1.00 |  | 1.04 | 1.00 |
|  | 1983-1993; age 30-39 | 1.21 | 1.00 |  | 1.19 | 1.00 |
|  | 1973-1983; age 40-49 | 1.36 | 1.00 |  | 1.19 | 1.00 |
|  | 1963-1973; age 50-59 | 1.44 | 1.00 |  | 1.41 | 1.24 |
|  | 1953-1963; age 60-69 | 1.15 | 1.00 |  | 1.41 | 1.20 |
|  | 1943-1953; age 70-79 | 1.47 | 1.00 |  | 1.48 | 1.18 |
|  | 1943 or earlier; age 80+ | 2.19 | 1.00 |  | 1.53 | 1.00 |
| Gender | (Ref: Male) |  |  |  |  |  |
|  | Female | 2.13 | 1.80 |  | 1.36 | 1.26 |
| Religious affiliation | (Ref: No religion/Atheist/Agnostic) |  |  |  |  |  |
|  | Islam | 1.94 | 1.00 |  | 1.52 | 1.00 |
|  | Christianity | 1.62 | 1.00 |  | 1.44 | 1.00 |
|  | Collapsed affiliations with prevalence<3% | 585472.16 | 246274.37 |  | 2.34 | 1.00 |
| Race/ethnicity | (Ref: Plurality group) |  |  |  |  |  |
|  | Non-plurality groups | 1.54 | 1.00 |  | 1.86 | 1.53 |

Table S20a. Nationally representative descriptive statistics for Turkey

| **Characteristic** | **N = 1,473**^1^ |
| --- | --- |
| **Relationship with mother** |  |
| Very good | 970 (66%) |
| Somewhat good | 401 (27%) |
| Somewhat bad | 48 (3.2%) |
| Very bad | 26 (1.8%) |
| Does not apply | 21 (1.4%) |
| (Missing) | 7 (0.5%) |
| **Relationship with father** |  |
| Very good | 795 (54%) |
| Somewhat good | 425 (29%) |
| Somewhat bad | 73 (5.0%) |
| Very bad | 95 (6.5%) |
| Does not apply | 60 (4.1%) |
| (Missing) | 25 (1.7%) |
| **Parent marital status** |  |
| Parents married | 1,325 (90%) |
| Divorced | 57 (3.9%) |
| Parents were never married | 7 (0.5%) |
| One or both parents had died | 61 (4.1%) |
| (Missing) | 23 (1.5%) |
| **Subjective financial status of family growing up** |  |
| Lived comfortably | 498 (34%) |
| Got by | 647 (44%) |
| Found it difficult | 218 (15%) |
| Found it very difficult | 108 (7.3%) |
| (Missing) | 2 (0.1%) |
| **Experienced Abuse** |  |
| Yes | 158 (11%) |
| No | 1,290 (88%) |
| (Missing) | 25 (1.7%) |
| **Felt like an outsider in the family** |  |
| Yes | 157 (11%) |
| No | 1,306 (89%) |
| (Missing) | 9 (0.6%) |
| **Self-rated health growing up** |  |
| Excellent | 377 (26%) |
| Very good | 410 (28%) |
| Good | 419 (28%) |
| Fair | 220 (15%) |
| Poor | 47 (3.2%) |
| (Missing) | 0 (<0.1%) |
| **Immigration status** |  |
| Born in this country | 1,415 (96%) |
| Born in another country | 58 (4.0%) |
| (Missing) | 0 (0%) |
| **Religious service attendance (age 12)** |  |
| At least 1/week | 609 (41%) |
| 1-3/month | 238 (16%) |
| <1/month | 225 (15%) |
| Never | 383 (26%) |
| (Missing) | 18 (1.2%) |
| **Year of birth** |  |
| 1998-2005; age 18-24 | 222 (15%) |
| 1993-1998; age 25-29 | 152 (10%) |
| 1983-1993; age 30-39 | 315 (21%) |
| 1973-1983; age 40-49 | 312 (21%) |
| 1963-1973; age 50-59 | 225 (15%) |
| 1953-1963; age 60-69 | 164 (11%) |
| 1943-1953; age 70-79 | 65 (4.4%) |
| 1943 or earlier; age 80+ | 18 (1.2%) |
| (Missing) | 0 (0%) |
| **Gender** |  |
| Male | 754 (51%) |
| Female | 719 (49%) |
| Other | 0 (0%) |
| (Missing) | 0 (0%) |
| **Religious affiliation** |  |
| Christianity | 1 (<0.1%) |
| Islam | 1,439 (98%) |
| Hinduism | 0 (0%) |
| Buddhism | 0 (0%) |
| Judaism | 1 (<0.1%) |
| Sikhism | 0 (0%) |
| Baha'i | 0 (0%) |
| Jainism | 0 (0%) |
| Shinto | 0 (0%) |
| Taoism | 0 (0%) |
| Confucianism | 0 (0%) |
| Primal, Animist, or Folk religion | 0 (0%) |
| Spiritism | 0 (0%) |
| Umbanda, Candomble, and other African-derived religions | 0 (0%) |
| Chinese folk/traditional religion | 0 (0%) |
| Some other religion | 0 (0%) |
| No religion/Atheist/Agnostic | 13 (0.9%) |
| (Missing) | 19 (1.3%) |
| **Race/Ethnicity** |  |
| Albanian | 8 (0.5%) |
| Arab | 51 (3.5%) |
| Armenian | 1 (<0.1%) |
| Azeri | 9 (0.6%) |
| Bosnian | 5 (0.3%) |
| Circassian | 19 (1.3%) |
| Georgian | 4 (0.3%) |
| Greek | 1 (<0.1%) |
| Kurdish/Zaza | 252 (17%) |
| Laz | 25 (1.7%) |
| Other | 58 (3.9%) |
| Turkish | 1,030 (70%) |
| Uyghur | 1 (<0.1%) |
| (Missing) | 9 (0.6%) |
| ^1^n (%) | |

Table S20b. Regression of Community Participation on Childhood Predictors for Turkey

|  | | Secular Community Participation | | | |  | Religious Service Attendance | | | |
| --- | --- | --- | --- | --- | --- | --- | --- | --- | --- | --- |
| Variable | Category | Risk-Ratio | RR 95% CI | log(RR) SE | Global p-value |  | Risk-Ratio | RR 95% CI | log(RR) SE | Global p-value |
| Relationship with mother | (Ref: Very bad/somewhat bad) |  |  |  | 0.287 |  |  |  |  | 0.192 |
|  | Very good/somewhat good | 0.78 | (0.49,1.24) | 0.24 |  |  | 0.86 | (0.68,1.09) | 0.12 |  |
| Relationship with father | (Ref: Very bad/somewhat bad) |  |  |  | 0.930 |  |  |  |  | 0.212 |
|  | Very good/somewhat good | 1.01 | (0.71,1.45) | 0.18 |  |  | 1.14 | (0.92,1.41) | 0.11 |  |
| Parent marital status | (Ref: Parents married) |  |  |  | 0.528 |  |  |  |  | 0.910 |
|  | Divorced | 0.95 | (0.55,1.65) | 0.28 |  |  | 0.99 | (0.72,1.35) | 0.16 |  |
|  | Parents were never married | 0.32 | (0.03,3.09) | 1.10 |  |  | 0.78 | (0.39,1.57) | 0.36 |  |
|  | One or both parents had died | 1.21 | (0.66,2.22) | 0.31 |  |  | 1.01 | (0.71,1.42) | 0.17 |  |
| Subjective financial status of family growing up | (Ref: Got by) |  |  |  | 0.443 |  |  |  |  | 0.155 |
|  | Lived comfortably | 1.14 | (0.89,1.46) | 0.13 |  |  | 1.08 | (0.95,1.24) | 0.07 |  |
|  | Found it difficult | 0.99 | (0.68,1.43) | 0.19 |  |  | 1.09 | (0.93,1.29) | 0.08 |  |
|  | Found it very difficult | 1.39 | (0.84,2.29) | 0.25 |  |  | 1.28 | (1.03,1.59) | 0.11 |  |
| Experienced Abuse | (Ref: No) |  |  |  | 0.537 |  |  |  |  | 0.013 |
|  | Yes | 0.90 | (0.64,1.27) | 0.18 |  |  | 0.73 | (0.57,0.94) | 0.13 |  |
| Felt like an outsider in the family | (Ref: No) |  |  |  | 0.320 |  |  |  |  | 0.768 |
|  | Yes | 1.19 | (0.84,1.70) | 0.18 |  |  | 0.97 | (0.80,1.19) | 0.10 |  |
| Self-rated health growing up | (Ref: Good) |  |  |  | 0.999 |  |  |  |  | 0.031 |
|  | Excellent | 0.97 | (0.73,1.29) | 0.15 |  |  | 0.77 | (0.65,0.91) | 0.08 |  |
|  | Very good | 0.96 | (0.70,1.33) | 0.17 |  |  | 0.90 | (0.77,1.05) | 0.08 |  |
|  | Fair | 0.95 | (0.65,1.39) | 0.19 |  |  | 0.92 | (0.77,1.11) | 0.09 |  |
|  | Poor | 0.96 | (0.43,2.15) | 0.41 |  |  | 0.74 | (0.47,1.16) | 0.23 |  |
| Immigration status | (Ref: Born in this country) |  |  |  | 0.837 |  |  |  |  | 0.610 |
|  | Born in another country | 0.98 | (0.53,1.81) | 0.31 |  |  | 1.08 | (0.80,1.46) | 0.15 |  |
| Religious service attendance (age 12) | (Ref: Never) |  |  |  | 0.061 |  |  |  |  | <.001 |
|  | At least 1/week | 1.29 | (0.93,1.79) | 0.17 |  |  | 1.67 | (1.41,1.99) | 0.09 |  |
|  | 1-3/month | 1.58 | (1.07,2.33) | 0.20 |  |  | 1.42 | (1.15,1.74) | 0.11 |  |
|  | < 1/month | 1.56 | (1.06,2.29) | 0.20 |  |  | 0.98 | (0.76,1.26) | 0.13 |  |
| Year of birth | (Ref: 1998-2005; current age: 18-24) |  |  |  | <.001 |  |  |  |  | 0.010 |
|  | 1993-1998; age 25-29 | 0.76 | (0.53,1.09) | 0.18 |  |  | 1.27 | (1.03,1.56) | 0.11 |  |
|  | 1983-1993; age 30-39 | 0.66 | (0.49,0.90) | 0.16 |  |  | 1.20 | (1.00,1.45) | 0.09 |  |
|  | 1973-1983; age 40-49 | 0.50 | (0.36,0.69) | 0.17 |  |  | 1.12 | (0.92,1.36) | 0.10 |  |
|  | 1963-1973; age 50-59 | 0.51 | (0.34,0.77) | 0.21 |  |  | 1.48 | (1.21,1.80) | 0.10 |  |
|  | 1953-1963; age 60-69 | 0.68 | (0.42,1.09) | 0.24 |  |  | 1.37 | (1.07,1.76) | 0.13 |  |
|  | 1943-1953; age 70-79 | 0.47 | (0.17,1.30) | 0.52 |  |  | 1.27 | (0.85,1.91) | 0.21 |  |
|  | 1943 or earlier; age 80+ | 0.60 | (0.09,3.87) | 0.95 |  |  | 1.40 | (0.75,2.61) | 0.32 |  |
| Gender | (Ref: Male) |  |  |  | <.001 |  |  |  |  | 0.210 |
|  | Female | 0.61 | (0.48,0.79) | 0.13 |  |  | 0.92 | (0.82,1.05) | 0.06 |  |
| Religious affiliation | (Ref: Islam) |  |  |  | 0.445 |  |  |  |  | 0.023 |
|  | Collapsed affiliations with prevalence<3% | 1.25 | (0.65,2.40) | 0.33 |  |  | 0.21 | (0.05,0.84) | 0.71 |  |
| Race/ethnicity | (Ref: Plurality group) |  |  |  | 0.176 |  |  |  |  | 0.003 |
|  | Non-plurality groups | 1.18 | (0.91,1.54) | 0.13 |  |  | 1.19 | (1.06,1.34) | 0.06 |  |

Table S20c. Sensitivity of the regression estimates to unmeasured confounding for Turkey

|  | | Secular Community Participation | |  | Religious Service Attendance | |
| --- | --- | --- | --- | --- | --- | --- |
| Variable | Category | E-value for Estimate | E-value for 95% CI |  | E-value for Estimate | E-value for 95% CI |
| Relationship with mother | (Ref: Very bad/somewhat bad) |  |  |  |  |  |
|  | Very good/somewhat good | 1.89 | 1.00 |  | 1.61 | 1.00 |
| Relationship with father | (Ref: Very bad/somewhat bad) |  |  |  |  |  |
|  | Very good/somewhat good | 1.12 | 1.00 |  | 1.54 | 1.00 |
| Parent marital status | (Ref: Parents married) |  |  |  |  |  |
|  | Divorced | 1.28 | 1.00 |  | 1.12 | 1.00 |
|  | Parents were never married | 5.65 | 1.00 |  | 1.88 | 1.00 |
|  | One or both parents had died | 1.72 | 1.00 |  | 1.09 | 1.00 |
| Subjective financial status of family growing up | (Ref: Got by) |  |  |  |  |  |
|  | Lived comfortably | 1.54 | 1.00 |  | 1.39 | 1.00 |
|  | Found it difficult | 1.11 | 1.00 |  | 1.42 | 1.00 |
|  | Found it very difficult | 2.13 | 1.00 |  | 1.87 | 1.19 |
| Experienced Abuse | (Ref: No) |  |  |  |  |  |
|  | Yes | 1.46 | 1.00 |  | 2.08 | 1.34 |
| Felt like an outsider in the family | (Ref: No) |  |  |  |  |  |
|  | Yes | 1.67 | 1.00 |  | 1.21 | 1.00 |
| Self-rated health growing up | (Ref: Good) |  |  |  |  |  |
|  | Excellent | 1.20 | 1.00 |  | 1.92 | 1.44 |
|  | Very good | 1.24 | 1.00 |  | 1.45 | 1.00 |
|  | Fair | 1.27 | 1.00 |  | 1.38 | 1.00 |
|  | Poor | 1.24 | 1.00 |  | 2.06 | 1.00 |
| Immigration status | (Ref: Born in this country) |  |  |  |  |  |
|  | Born in another country | 1.18 | 1.00 |  | 1.38 | 1.00 |
| Religious service attendance (age 12) | (Ref: Never) |  |  |  |  |  |
|  | At least 1/week | 1.90 | 1.00 |  | 2.74 | 2.16 |
|  | 1-3/month | 2.53 | 1.34 |  | 2.18 | 1.57 |
|  | < 1/month | 2.49 | 1.31 |  | 1.18 | 1.00 |
| Year of birth | (Ref: 1998-2005; current age: 18-24) |  |  |  |  |  |
|  | 1993-1998; age 25-29 | 1.95 | 1.00 |  | 1.85 | 1.20 |
|  | 1983-1993; age 30-39 | 2.38 | 1.47 |  | 1.69 | 1.00 |
|  | 1973-1983; age 40-49 | 3.43 | 2.26 |  | 1.48 | 1.00 |
|  | 1963-1973; age 50-59 | 3.31 | 1.91 |  | 2.31 | 1.71 |
|  | 1953-1963; age 60-69 | 2.29 | 1.00 |  | 2.09 | 1.34 |
|  | 1943-1953; age 70-79 | 3.67 | 1.00 |  | 1.87 | 1.00 |
|  | 1943 or earlier; age 80+ | 2.70 | 1.00 |  | 2.14 | 1.00 |
| Gender | (Ref: Male) |  |  |  |  |  |
|  | Female | 2.64 | 1.85 |  | 1.38 | 1.00 |
| Religious affiliation | (Ref: Islam) |  |  |  |  |  |
|  | Collapsed affiliations with prevalence<3% | 1.81 | 1.00 |  | 9.12 | 1.66 |
| Race/ethnicity | (Ref: Plurality group) |  |  |  |  |  |
|  | Non-plurality groups | 1.65 | 1.00 |  | 1.66 | 1.30 |

Table S21a. Nationally representative descriptive statistics for United Kingdom

| **Characteristic** | **N = 5,368**^1^ |
| --- | --- |
| **Relationship with mother** |  |
| Very good | 3,435 (64%) |
| Somewhat good | 1,338 (25%) |
| Somewhat bad | 325 (6.1%) |
| Very bad | 150 (2.8%) |
| Does not apply | 92 (1.7%) |
| (Missing) | 27 (0.5%) |
| **Relationship with father** |  |
| Very good | 2,907 (54%) |
| Somewhat good | 1,383 (26%) |
| Somewhat bad | 407 (7.6%) |
| Very bad | 321 (6.0%) |
| Does not apply | 321 (6.0%) |
| (Missing) | 29 (0.5%) |
| **Parent marital status** |  |
| Parents married | 4,343 (81%) |
| Divorced | 481 (9.0%) |
| Parents were never married | 315 (5.9%) |
| One or both parents had died | 154 (2.9%) |
| (Missing) | 75 (1.4%) |
| **Subjective financial status of family growing up** |  |
| Lived comfortably | 2,552 (48%) |
| Got by | 1,933 (36%) |
| Found it difficult | 632 (12%) |
| Found it very difficult | 230 (4.3%) |
| (Missing) | 22 (0.4%) |
| **Experienced Abuse** |  |
| Yes | 864 (16%) |
| No | 4,455 (83%) |
| (Missing) | 49 (0.9%) |
| **Felt like an outsider in the family** |  |
| Yes | 1,017 (19%) |
| No | 4,308 (80%) |
| (Missing) | 43 (0.8%) |
| **Self-rated health growing up** |  |
| Excellent | 2,154 (40%) |
| Very good | 1,736 (32%) |
| Good | 995 (19%) |
| Fair | 332 (6.2%) |
| Poor | 130 (2.4%) |
| (Missing) | 20 (0.4%) |
| **Immigration status** |  |
| Born in this country | 4,659 (87%) |
| Born in another country | 682 (13%) |
| (Missing) | 27 (0.5%) |
| **Religious service attendance (age 12)** |  |
| At least 1/week | 1,732 (32%) |
| 1-3/month | 733 (14%) |
| <1/month | 903 (17%) |
| Never | 1,972 (37%) |
| (Missing) | 28 (0.5%) |
| **Year of birth** |  |
| 1998-2005; age 18-24 | 490 (9.1%) |
| 1993-1998; age 25-29 | 391 (7.3%) |
| 1983-1993; age 30-39 | 946 (18%) |
| 1973-1983; age 40-49 | 827 (15%) |
| 1963-1973; age 50-59 | 949 (18%) |
| 1953-1963; age 60-69 | 889 (17%) |
| 1943-1953; age 70-79 | 711 (13%) |
| 1943 or earlier; age 80+ | 163 (3.0%) |
| (Missing) | 1 (<0.1%) |
| **Gender** |  |
| Male | 2,557 (48%) |
| Female | 2,789 (52%) |
| Other | 14 (0.3%) |
| (Missing) | 9 (0.2%) |
| **Religious affiliation** |  |
| Christianity | 3,461 (64%) |
| Islam | 230 (4.3%) |
| Hinduism | 88 (1.6%) |
| Buddhism | 15 (0.3%) |
| Judaism | 59 (1.1%) |
| Sikhism | 30 (0.6%) |
| Baha'i | 5 (<0.1%) |
| Jainism | 0 (<0.1%) |
| Shinto | 0 (0%) |
| Taoism | 2 (<0.1%) |
| Confucianism | 3 (<0.1%) |
| Primal, Animist, or Folk religion | 22 (0.4%) |
| Spiritism | 0 (0%) |
| Umbanda, Candomble, and other African-derived religions | 0 (0%) |
| Chinese folk/traditional religion | 0 (0%) |
| Some other religion | 24 (0.5%) |
| No religion/Atheist/Agnostic | 1,409 (26%) |
| (Missing) | 21 (0.4%) |
| **Race/Ethnicity** |  |
| Asian | 426 (7.9%) |
| Black | 152 (2.8%) |
| Other | 96 (1.8%) |
| White | 4,647 (87%) |
| (Missing) | 47 (0.9%) |
| ^1^n (%) | |

Table S21b. Regression of Community Participation on Childhood Predictors for United Kingdom

|  | | Secular Community Participation | | | |  | Religious Service Attendance | | | |
| --- | --- | --- | --- | --- | --- | --- | --- | --- | --- | --- |
| Variable | Category | Risk-Ratio | RR 95% CI | log(RR) SE | Global p-value |  | Risk-Ratio | RR 95% CI | log(RR) SE | Global p-value |
| Relationship with mother | (Ref: Very bad/somewhat bad) |  |  |  | 0.065 |  |  |  |  | 0.147 |
|  | Very good/somewhat good | 1.28 | (0.98,1.66) | 0.13 |  |  | 1.27 | (0.92,1.74) | 0.16 |  |
| Relationship with father | (Ref: Very bad/somewhat bad) |  |  |  | 0.853 |  |  |  |  | 0.021 |
|  | Very good/somewhat good | 1.01 | (0.81,1.26) | 0.11 |  |  | 1.38 | (1.05,1.81) | 0.14 |  |
| Parent marital status | (Ref: Parents married) |  |  |  | 0.209 |  |  |  |  | 0.069 |
|  | Divorced | 0.80 | (0.60,1.06) | 0.14 |  |  | 0.62 | (0.39,0.98) | 0.24 |  |
|  | Parents were never married | 0.97 | (0.67,1.39) | 0.19 |  |  | 0.76 | (0.44,1.31) | 0.28 |  |
|  | One or both parents had died | 1.27 | (0.86,1.88) | 0.20 |  |  | 1.38 | (0.79,2.39) | 0.28 |  |
| Subjective financial status of family growing up | (Ref: Got by) |  |  |  | 0.041 |  |  |  |  | 0.824 |
|  | Lived comfortably | 1.20 | (1.03,1.40) | 0.08 |  |  | 0.93 | (0.78,1.11) | 0.09 |  |
|  | Found it difficult | 0.93 | (0.72,1.19) | 0.13 |  |  | 1.01 | (0.76,1.34) | 0.14 |  |
|  | Found it very difficult | 1.16 | (0.78,1.72) | 0.20 |  |  | 1.05 | (0.64,1.75) | 0.26 |  |
| Experienced Abuse | (Ref: No) |  |  |  | 0.588 |  |  |  |  | 0.319 |
|  | Yes | 1.05 | (0.88,1.24) | 0.09 |  |  | 1.11 | (0.90,1.38) | 0.11 |  |
| Felt like an outsider in the family | (Ref: No) |  |  |  | <.001 |  |  |  |  | <.001 |
|  | Yes | 1.30 | (1.11,1.51) | 0.08 |  |  | 1.43 | (1.18,1.74) | 0.10 |  |
| Self-rated health growing up | (Ref: Good) |  |  |  | 0.337 |  |  |  |  | 0.486 |
|  | Excellent | 1.04 | (0.85,1.28) | 0.10 |  |  | 1.29 | (0.98,1.69) | 0.14 |  |
|  | Very good | 1.07 | (0.87,1.30) | 0.10 |  |  | 1.20 | (0.92,1.58) | 0.14 |  |
|  | Fair | 0.73 | (0.50,1.07) | 0.19 |  |  | 1.16 | (0.74,1.83) | 0.23 |  |
|  | Poor | 0.92 | (0.53,1.57) | 0.28 |  |  | 1.16 | (0.60,2.24) | 0.34 |  |
| Immigration status | (Ref: Born in this country) |  |  |  | 0.564 |  |  |  |  | 0.412 |
|  | Born in another country | 0.94 | (0.76,1.16) | 0.11 |  |  | 0.91 | (0.72,1.14) | 0.12 |  |
| Religious service attendance (age 12) | (Ref: Never) |  |  |  | <.001 |  |  |  |  | <.001 |
|  | At least 1/week | 1.86 | (1.51,2.30) | 0.11 |  |  | 11.55 | (7.03,18.97) | 0.25 |  |
|  | 1-3/month | 1.83 | (1.45,2.33) | 0.12 |  |  | 8.45 | (5.03,14.21) | 0.26 |  |
|  | < 1/month | 1.53 | (1.21,1.94) | 0.12 |  |  | 2.31 | (1.28,4.15) | 0.30 |  |
| Year of birth | (Ref: 1998-2005; current age: 18-24) |  |  |  | 0.067 |  |  |  |  | <.001 |
|  | 1993-1998; age 25-29 | 0.95 | (0.67,1.34) | 0.18 |  |  | 1.68 | (1.11,2.54) | 0.21 |  |
|  | 1983-1993; age 30-39 | 0.95 | (0.70,1.29) | 0.16 |  |  | 1.63 | (1.11,2.40) | 0.20 |  |
|  | 1973-1983; age 40-49 | 0.83 | (0.60,1.15) | 0.17 |  |  | 1.16 | (0.77,1.74) | 0.21 |  |
|  | 1963-1973; age 50-59 | 0.78 | (0.56,1.09) | 0.17 |  |  | 1.04 | (0.67,1.61) | 0.22 |  |
|  | 1953-1963; age 60-69 | 0.66 | (0.47,0.92) | 0.17 |  |  | 0.92 | (0.58,1.44) | 0.23 |  |
|  | 1943-1953; age 70-79 | 0.89 | (0.64,1.25) | 0.17 |  |  | 0.90 | (0.57,1.42) | 0.23 |  |
|  | 1943 or earlier; age 80+ | 0.75 | (0.47,1.20) | 0.24 |  |  | 1.19 | (0.69,2.06) | 0.28 |  |
| Gender | (Ref: Male) |  |  |  | <.001 |  |  |  |  | <.001 |
|  | Female | 0.77 | (0.67,0.89) | 0.07 |  |  | 0.70 | (0.59,0.83) | 0.09 |  |
|  | Other | 0.22 | (0.03,1.77) | 1.05 |  |  | 0.50 | (0.06,3.88) | 1.05 |  |
| Religious affiliation | (Ref: No religion/Atheist/Agnostic) |  |  |  | 0.204 |  |  |  |  | <.001 |
|  | Islam | 1.10 | (0.72,1.68) | 0.22 |  |  | 2.83 | (1.79,4.46) | 0.23 |  |
|  | Christianity | 1.24 | (1.01,1.52) | 0.10 |  |  | 2.05 | (1.40,2.99) | 0.19 |  |
|  | Collapsed affiliations with prevalence<3% | 1.04 | (0.69,1.56) | 0.21 |  |  | 1.53 | (0.81,2.90) | 0.33 |  |
| Race/ethnicity | (Ref: Plurality group) |  |  |  | 0.408 |  |  |  |  | 0.006 |
|  | Non-plurality groups | 0.89 | (0.67,1.19) | 0.15 |  |  | 1.43 | (1.11,1.85) | 0.13 |  |

Table S21c. Sensitivity of the regression estimates to unmeasured confounding for United Kingdom

|  | | Secular Community Participation | |  | Religious Service Attendance | |
| --- | --- | --- | --- | --- | --- | --- |
| Variable | Category | E-value for Estimate | E-value for 95% CI |  | E-value for Estimate | E-value for 95% CI |
| Relationship with mother | (Ref: Very bad/somewhat bad) |  |  |  |  |  |
|  | Very good/somewhat good | 1.87 | 1.00 |  | 1.85 | 1.00 |
| Relationship with father | (Ref: Very bad/somewhat bad) |  |  |  |  |  |
|  | Very good/somewhat good | 1.11 | 1.00 |  | 2.10 | 1.27 |
| Parent marital status | (Ref: Parents married) |  |  |  |  |  |
|  | Divorced | 1.81 | 1.00 |  | 2.62 | 1.15 |
|  | Parents were never married | 1.23 | 1.00 |  | 1.97 | 1.00 |
|  | One or both parents had died | 1.86 | 1.00 |  | 2.10 | 1.00 |
| Subjective financial status of family growing up | (Ref: Got by) |  |  |  |  |  |
|  | Lived comfortably | 1.70 | 1.22 |  | 1.36 | 1.00 |
|  | Found it difficult | 1.37 | 1.00 |  | 1.11 | 1.00 |
|  | Found it very difficult | 1.59 | 1.00 |  | 1.29 | 1.00 |
| Experienced Abuse | (Ref: No) |  |  |  |  |  |
|  | Yes | 1.27 | 1.00 |  | 1.47 | 1.00 |
| Felt like an outsider in the family | (Ref: No) |  |  |  |  |  |
|  | Yes | 1.92 | 1.46 |  | 2.22 | 1.64 |
| Self-rated health growing up | (Ref: Good) |  |  |  |  |  |
|  | Excellent | 1.25 | 1.00 |  | 1.89 | 1.00 |
|  | Very good | 1.33 | 1.00 |  | 1.70 | 1.00 |
|  | Fair | 2.07 | 1.00 |  | 1.59 | 1.00 |
|  | Poor | 1.40 | 1.00 |  | 1.58 | 1.00 |
| Immigration status | (Ref: Born in this country) |  |  |  |  |  |
|  | Born in another country | 1.33 | 1.00 |  | 1.43 | 1.00 |
| Religious service attendance (age 12) | (Ref: Never) |  |  |  |  |  |
|  | At least 1/week | 3.13 | 2.39 |  | 22.58 | 13.53 |
|  | 1-3/month | 3.07 | 2.25 |  | 16.39 | 9.53 |
|  | < 1/month | 2.43 | 1.70 |  | 4.04 | 1.89 |
| Year of birth | (Ref: 1998-2005; current age: 18-24) |  |  |  |  |  |
|  | 1993-1998; age 25-29 | 1.29 | 1.00 |  | 2.74 | 1.45 |
|  | 1983-1993; age 30-39 | 1.29 | 1.00 |  | 2.65 | 1.46 |
|  | 1973-1983; age 40-49 | 1.70 | 1.00 |  | 1.58 | 1.00 |
|  | 1963-1973; age 50-59 | 1.89 | 1.00 |  | 1.24 | 1.00 |
|  | 1953-1963; age 60-69 | 2.39 | 1.38 |  | 1.41 | 1.00 |
|  | 1943-1953; age 70-79 | 1.48 | 1.00 |  | 1.46 | 1.00 |
|  | 1943 or earlier; age 80+ | 2.01 | 1.00 |  | 1.67 | 1.00 |
| Gender | (Ref: Male) |  |  |  |  |  |
|  | Female | 1.91 | 1.51 |  | 2.21 | 1.70 |
|  | Other | 8.37 | 1.00 |  | 3.42 | 1.00 |
| Religious affiliation | (Ref: No religion/Atheist/Agnostic) |  |  |  |  |  |
|  | Islam | 1.44 | 1.00 |  | 5.10 | 2.98 |
|  | Christianity | 1.78 | 1.09 |  | 3.51 | 2.15 |
|  | Collapsed affiliations with prevalence<3% | 1.23 | 1.00 |  | 2.43 | 1.00 |
| Race/ethnicity | (Ref: Plurality group) |  |  |  |  |  |
|  | Non-plurality groups | 1.49 | 1.00 |  | 2.21 | 1.45 |

Table S22a. Nationally representative descriptive statistics for United States

| **Characteristic** | **N = 38,312**^1^ |
| --- | --- |
| **Relationship with mother** |  |
| Very good | 20,590 (54%) |
| Somewhat good | 11,525 (30%) |
| Somewhat bad | 3,523 (9.2%) |
| Very bad | 1,874 (4.9%) |
| Does not apply | 694 (1.8%) |
| (Missing) | 106 (0.3%) |
| **Relationship with father** |  |
| Very good | 15,313 (40%) |
| Somewhat good | 12,665 (33%) |
| Somewhat bad | 4,879 (13%) |
| Very bad | 2,604 (6.8%) |
| Does not apply | 2,811 (7.3%) |
| (Missing) | 38 (0.1%) |
| **Parent marital status** |  |
| Parents married | 27,415 (72%) |
| Divorced | 6,325 (17%) |
| Parents were never married | 3,048 (8.0%) |
| One or both parents had died | 1,024 (2.7%) |
| (Missing) | 500 (1.3%) |
| **Subjective financial status of family growing up** |  |
| Lived comfortably | 15,116 (39%) |
| Got by | 15,682 (41%) |
| Found it difficult | 5,152 (13%) |
| Found it very difficult | 2,342 (6.1%) |
| (Missing) | 19 (<0.1%) |
| **Experienced Abuse** |  |
| Yes | 10,026 (26%) |
| No | 28,045 (73%) |
| (Missing) | 242 (0.6%) |
| **Felt like an outsider in the family** |  |
| Yes | 10,185 (27%) |
| No | 27,714 (72%) |
| (Missing) | 413 (1.1%) |
| **Self-rated health growing up** |  |
| Excellent | 16,866 (44%) |
| Very good | 12,108 (32%) |
| Good | 6,444 (17%) |
| Fair | 2,303 (6.0%) |
| Poor | 520 (1.4%) |
| (Missing) | 71 (0.2%) |
| **Immigration status** |  |
| Born in this country | 34,865 (91%) |
| Born in another country | 3,020 (7.9%) |
| (Missing) | 427 (1.1%) |
| **Religious service attendance (age 12)** |  |
| At least 1/week | 18,609 (49%) |
| 1-3/month | 6,644 (17%) |
| <1/month | 5,829 (15%) |
| Never | 7,085 (18%) |
| (Missing) | 145 (0.4%) |
| **Year of birth** |  |
| 1998-2005; age 18-24 | 2,682 (7.0%) |
| 1993-1998; age 25-29 | 3,540 (9.2%) |
| 1983-1993; age 30-39 | 7,284 (19%) |
| 1973-1983; age 40-49 | 5,649 (15%) |
| 1963-1973; age 50-59 | 6,745 (18%) |
| 1953-1963; age 60-69 | 6,832 (18%) |
| 1943-1953; age 70-79 | 4,054 (11%) |
| 1943 or earlier; age 80+ | 1,525 (4.0%) |
| (Missing) | 0 (0%) |
| **Gender** |  |
| Male | 18,222 (48%) |
| Female | 19,562 (51%) |
| Other | 392 (1.0%) |
| (Missing) | 136 (0.4%) |
| **Religious affiliation** |  |
| Christianity | 30,444 (79%) |
| Islam | 220 (0.6%) |
| Hinduism | 203 (0.5%) |
| Buddhism | 172 (0.4%) |
| Judaism | 787 (2.1%) |
| Sikhism | 47 (0.1%) |
| Baha'i | 4 (<0.1%) |
| Jainism | 18 (<0.1%) |
| Shinto | 6 (<0.1%) |
| Taoism | 17 (<0.1%) |
| Confucianism | 8 (<0.1%) |
| Primal, Animist, or Folk religion | 67 (0.2%) |
| Spiritism | 0 (0%) |
| Umbanda, Candomble, and other African-derived religions | 0 (0%) |
| Chinese folk/traditional religion | 0 (0%) |
| Some other religion | 359 (0.9%) |
| No religion/Atheist/Agnostic | 5,845 (15%) |
| (Missing) | 115 (0.3%) |
| **Race/Ethnicity** |  |
| Asian | 2,466 (6.4%) |
| Black | 4,501 (12%) |
| Hispanic | 6,724 (18%) |
| Other | 997 (2.6%) |
| White | 23,605 (62%) |
| (Missing) | 20 (<0.1%) |
| ^1^n (%) | |

Table S22b. Regression of Community Participation on Childhood Predictors for United States

|  | | Secular Community Participation | | | |  | Religious Service Attendance | | | |
| --- | --- | --- | --- | --- | --- | --- | --- | --- | --- | --- |
| Variable | Category | Risk-Ratio | RR 95% CI | log(RR) SE | Global p-value |  | Risk-Ratio | RR 95% CI | log(RR) SE | Global  p-value |
| Relationship with mother | (Ref: Very bad/somewhat bad) |  |  |  | 0.204 |  |  |  |  | 0.021 |
|  | Very good/somewhat good | 0.89 | (0.74,1.07) | 0.09 |  |  | 1.19 | (1.03,1.38) | 0.08 |  |
| Relationship with father | (Ref: Very bad/somewhat bad) |  |  |  | 0.199 |  |  |  |  | 0.003 |
|  | Very good/somewhat good | 1.11 | (0.94,1.30) | 0.08 |  |  | 1.18 | (1.06,1.33) | 0.06 |  |
| Parent marital status | (Ref: Parents married) |  |  |  | 0.898 |  |  |  |  | 0.108 |
|  | Divorced | 1.04 | (0.89,1.22) | 0.08 |  |  | 0.87 | (0.77,0.99) | 0.06 |  |
|  | Parents were never married | 0.95 | (0.65,1.38) | 0.19 |  |  | 0.87 | (0.67,1.13) | 0.14 |  |
|  | One or both parents had died | 0.99 | (0.72,1.35) | 0.16 |  |  | 0.84 | (0.63,1.12) | 0.14 |  |
| Subjective financial status of family growing up | (Ref: Got by) |  |  |  | 0.003 |  |  |  |  | 0.018 |
|  | Lived comfortably | 1.19 | (1.07,1.33) | 0.05 |  |  | 0.97 | (0.90,1.04) | 0.04 |  |
|  | Found it difficult | 0.99 | (0.84,1.18) | 0.09 |  |  | 1.15 | (1.01,1.31) | 0.07 |  |
|  | Found it very difficult | 1.24 | (0.97,1.59) | 0.13 |  |  | 1.21 | (1.01,1.45) | 0.09 |  |
| Experienced Abuse | (Ref: No) |  |  |  | 0.888 |  |  |  |  | 0.522 |
|  | Yes | 1.00 | (0.89,1.13) | 0.06 |  |  | 0.97 | (0.88,1.07) | 0.05 |  |
| Felt like an outsider in the family | (Ref: No) |  |  |  | 0.643 |  |  |  |  | <.001 |
|  | Yes | 0.97 | (0.83,1.13) | 0.08 |  |  | 0.68 | (0.61,0.77) | 0.06 |  |
| Self-rated health growing up | (Ref: Good) |  |  |  | 0.072 |  |  |  |  | 0.136 |
|  | Excellent | 1.19 | (1.00,1.41) | 0.09 |  |  | 1.12 | (0.99,1.26) | 0.06 |  |
|  | Very good | 1.06 | (0.89,1.26) | 0.09 |  |  | 1.11 | (0.98,1.26) | 0.06 |  |
|  | Fair | 0.98 | (0.71,1.35) | 0.16 |  |  | 0.85 | (0.65,1.11) | 0.14 |  |
|  | Poor | 1.42 | (0.88,2.27) | 0.24 |  |  | 1.04 | (0.68,1.60) | 0.22 |  |
| Immigration status | (Ref: Born in this country) |  |  |  | 0.326 |  |  |  |  | 0.838 |
|  | Born in another country | 0.89 | (0.71,1.12) | 0.11 |  |  | 0.98 | (0.83,1.17) | 0.09 |  |
| Religious service attendance (age 12) | (Ref: Never) |  |  |  | <.001 |  |  |  |  | <.001 |
|  | At least 1/week | 1.43 | (1.22,1.68) | 0.08 |  |  | 3.17 | (2.65,3.80) | 0.09 |  |
|  | 1-3/month | 1.35 | (1.11,1.63) | 0.10 |  |  | 1.62 | (1.31,2.01) | 0.11 |  |
|  | < 1/month | 1.12 | (0.92,1.36) | 0.10 |  |  | 1.33 | (1.08,1.64) | 0.11 |  |
| Year of birth | (Ref: 1998-2005; current age: 18-24) |  |  |  | <.001 |  |  |  |  | <.001 |
|  | 1993-1998; age 25-29 | 0.94 | (0.65,1.37) | 0.19 |  |  | 1.12 | (0.75,1.69) | 0.21 |  |
|  | 1983-1993; age 30-39 | 0.82 | (0.58,1.15) | 0.17 |  |  | 1.15 | (0.79,1.66) | 0.19 |  |
|  | 1973-1983; age 40-49 | 0.85 | (0.61,1.19) | 0.17 |  |  | 1.21 | (0.83,1.75) | 0.19 |  |
|  | 1963-1973; age 50-59 | 0.73 | (0.53,1.02) | 0.17 |  |  | 1.35 | (0.93,1.95) | 0.19 |  |
|  | 1953-1963; age 60-69 | 0.81 | (0.59,1.12) | 0.16 |  |  | 1.43 | (0.99,2.06) | 0.19 |  |
|  | 1943-1953; age 70-79 | 1.01 | (0.74,1.39) | 0.16 |  |  | 1.48 | (1.03,2.13) | 0.19 |  |
|  | 1943 or earlier; age 80+ | 1.19 | (0.83,1.71) | 0.18 |  |  | 1.72 | (1.18,2.51) | 0.19 |  |
| Gender | (Ref: Male) |  |  |  | <.001 |  |  |  |  | <.001 |
|  | Female | 0.84 | (0.76,0.92) | 0.05 |  |  | 1.16 | (1.08,1.24) | 0.04 |  |
|  | Other | 2.00 | (1.22,3.26) | 0.25 |  |  | 0.19 | (0.08,0.46) | 0.45 |  |
| Religious affiliation | (Ref: No religion/Atheist/Agnostic) |  |  |  | 0.059 |  |  |  |  | <.001 |
|  | Christianity | 0.85 | (0.71,1.01) | 0.09 |  |  | 1.79 | (1.45,2.21) | 0.11 |  |
|  | Collapsed affiliations with prevalence<3% | 1.03 | (0.79,1.34) | 0.13 |  |  | 1.20 | (0.87,1.66) | 0.16 |  |
| Race/ethnicity | (Ref: Plurality group) |  |  |  | <.001 |  |  |  |  | 0.029 |
|  | Non-plurality groups | 0.79 | (0.69,0.91) | 0.07 |  |  | 1.10 | (1.01,1.21) | 0.05 |  |

Table S22c. Sensitivity of the regression estimates to unmeasured confounding for United States

|  | | Secular Community Participation | |  | Religious Service Attendance | |
| --- | --- | --- | --- | --- | --- | --- |
| Variable | Category | E-value for Estimate | E-value for 95% CI |  | E-value for Estimate | E-value for 95% CI |
| Relationship with mother | (Ref: Very bad/somewhat bad) |  |  |  |  |  |
|  | Very good/somewhat good | 1.50 | 1.00 |  | 1.66 | 1.19 |
| Relationship with father | (Ref: Very bad/somewhat bad) |  |  |  |  |  |
|  | Very good/somewhat good | 1.45 | 1.00 |  | 1.65 | 1.31 |
| Parent marital status | (Ref: Parents married) |  |  |  |  |  |
|  | Divorced | 1.25 | 1.00 |  | 1.57 | 1.13 |
|  | Parents were never married | 1.30 | 1.00 |  | 1.57 | 1.00 |
|  | One or both parents had died | 1.12 | 1.00 |  | 1.66 | 1.00 |
| Subjective financial status of family growing up | (Ref: Got by) |  |  |  |  |  |
|  | Lived comfortably | 1.68 | 1.35 |  | 1.22 | 1.00 |
|  | Found it difficult | 1.08 | 1.00 |  | 1.56 | 1.10 |
|  | Found it very difficult | 1.79 | 1.00 |  | 1.71 | 1.09 |
| Experienced Abuse | (Ref: No) |  |  |  |  |  |
|  | Yes | 1.04 | 1.00 |  | 1.21 | 1.00 |
| Felt like an outsider in the family | (Ref: No) |  |  |  |  |  |
|  | Yes | 1.22 | 1.00 |  | 2.28 | 1.91 |
| Self-rated health growing up | (Ref: Good) |  |  |  |  |  |
|  | Excellent | 1.66 | 1.00 |  | 1.48 | 1.00 |
|  | Very good | 1.31 | 1.00 |  | 1.45 | 1.00 |
|  | Fair | 1.16 | 1.00 |  | 1.62 | 1.00 |
|  | Poor | 2.19 | 1.00 |  | 1.24 | 1.00 |
| Immigration status | (Ref: Born in this country) |  |  |  |  |  |
|  | Born in another country | 1.48 | 1.00 |  | 1.14 | 1.00 |
| Religious service attendance (age 12) | (Ref: Never) |  |  |  |  |  |
|  | At least 1/week | 2.22 | 1.74 |  | 5.80 | 4.73 |
|  | 1-3/month | 2.03 | 1.47 |  | 2.62 | 1.94 |
|  | < 1/month | 1.48 | 1.00 |  | 1.99 | 1.36 |
| Year of birth | (Ref: 1998-2005; current age: 18-24) |  |  |  |  |  |
|  | 1993-1998; age 25-29 | 1.31 | 1.00 |  | 1.50 | 1.00 |
|  | 1983-1993; age 30-39 | 1.74 | 1.00 |  | 1.56 | 1.00 |
|  | 1973-1983; age 40-49 | 1.64 | 1.00 |  | 1.70 | 1.00 |
|  | 1963-1973; age 50-59 | 2.06 | 1.00 |  | 2.03 | 1.00 |
|  | 1953-1963; age 60-69 | 1.76 | 1.00 |  | 2.22 | 1.00 |
|  | 1943-1953; age 70-79 | 1.14 | 1.00 |  | 2.32 | 1.19 |
|  | 1943 or earlier; age 80+ | 1.67 | 1.00 |  | 2.84 | 1.65 |
| Gender | (Ref: Male) |  |  |  |  |  |
|  | Female | 1.67 | 1.38 |  | 1.58 | 1.37 |
|  | Other | 3.41 | 1.75 |  | 9.85 | 3.76 |
| Religious affiliation | (Ref: No religion/Atheist/Agnostic) |  |  |  |  |  |
|  | Christianity | 1.64 | 1.00 |  | 2.98 | 2.25 |
|  | Collapsed affiliations with prevalence<3% | 1.21 | 1.00 |  | 1.69 | 1.00 |
| Race/ethnicity | (Ref: Plurality group) |  |  |  |  |  |
|  | Non-plurality groups | 1.83 | 1.44 |  | 1.44 | 1.11 |

Table S23. Population weighted meta-analysis of regression results.

|  |  | Secular Community Participation | |  | Religious Service Attendance | |
| --- | --- | --- | --- | --- | --- | --- |
| Variable | Category | RR | 95% CI |  | RR | 95% CI |
| Relationship with mother | (Ref: Very bad/somewhat bad) |  |  |  |  |  |
|  | Very good/somewhat good | 0.96 | (0.82,1.12) |  | 1.07 | (0.99,1.15) |
| Relationship with father | (Ref: Very bad/somewhat bad) |  |  |  |  |  |
|  | Very good/somewhat good | 1.14 | (0.96,1.34) |  | 1.13 | (1.06,1.21) |
| Parent marital status | (Ref: Parents married) |  |  |  |  |  |
|  | Divorced | 1.12 | (0.94,1.32) |  | 0.91 | (0.84,0.98) |
|  | Single, never married | 0.73 | (0.65,0.81) |  | 0.94 | (0.89,1.00) |
|  | One or both parents had died | 0.85 | (0.76,0.94) |  | 0.93 | (0.87,0.98) |
| Subjective financial status of family growing up | (Ref: Got by) |  |  |  |  |  |
|  | Lived comfortably | 1.12 | (1.05,1.18) |  | 1.02 | (0.99,1.06) |
|  | Found it difficult | 0.99 | (0.92,1.06) |  | 1.03 | (0.99,1.07) |
|  | Found it very difficult | 1.03 | (0.94,1.13) |  | 1.02 | (0.97,1.07) |
| Experienced Abuse | (Ref: No) |  |  |  |  |  |
|  | Yes | 1.25 | (1.17,1.33) |  | 1.03 | (0.99,1.06) |
| Felt like an outsider in the family | (Ref: No) |  |  |  |  |  |
|  | Yes | 1.03 | (0.96,1.10) |  | 1.01 | (0.98,1.05) |
| Self-rated health growing up | (Ref: Good) |  |  |  |  |  |
|  | Excellent | 1.09 | (1.01,1.17) |  | 1.05 | (1.01,1.08) |
|  | Very good | 1.06 | (0.99,1.12) |  | 1.03 | (1.00,1.07) |
|  | Fair | 0.94 | (0.87,1.02) |  | 0.98 | (0.93,1.03) |
|  | Poor | 1.09 | (0.93,1.27) |  | 1.04 | (0.96,1.13) |
| Immigration status | (Ref: Born in this country) |  |  |  |  |  |
|  | Born in another country | 1.08 | (0.84,1.40) |  | 1.02 | (0.92,1.13) |
| Religious service attendance (age 12) | (Ref: Never) |  |  |  |  |  |
|  | At least 1/week | 1.49 | (1.37,1.63) |  | 2.10 | (2.00,2.21) |
|  | 1-3/month | 1.42 | (1.29,1.57) |  | 1.61 | (1.52,1.69) |
|  | < 1/month | 1.21 | (1.09,1.33) |  | 1.15 | (1.08,1.21) |
| Year of birth | (Ref: 1998-2005; age 18-24) |  |  |  |  |  |
|  | 1993-1998; age 25-29 | 0.90 | (0.83,0.98) |  | 1.08 | (1.02,1.15) |
|  | 1983-1993; age 30-39 | 0.79 | (0.74,0.85) |  | 1.09 | (1.03,1.15) |
|  | 1973-1983; age 40-49 | 0.70 | (0.64,0.76) |  | 1.10 | (1.04,1.16) |
|  | 1963-1973; age 50-59 | 0.69 | (0.63,0.76) |  | 1.15 | (1.08,1.22) |
|  | 1953-1963; age 60-69 | 0.70 | (0.63,0.79) |  | 1.20 | (1.13,1.28) |
|  | 1943-1953; age 70-79 | 0.73 | (0.60,0.88) |  | 1.20 | (1.11,1.30) |
|  | 1943 or earlier; age 80+ | 0.23 | (0.17,0.32) |  | 1.34 | (1.18,1.52) |
| Gender | (Ref: Male) |  |  |  |  |  |
|  | Female | 0.71 | (0.67,0.74) |  | 0.97 | (0.95,0.99) |
|  | Other | 0.31 | (0.23,0.43) |  | 0.29 | (0.22,0.39) |

Table S24. Population weighted meta-analysis of E-values.

|  |  | Secular Community Participation | |  | Religious Service Attendance | |
| --- | --- | --- | --- | --- | --- | --- |
| Variable | Category | E-value for Estimate | E-value for 95% CI |  | E-value for Estimate | E-value for 95% CI |
| Relationship with mother | (Ref: Very bad/somewhat bad) |  |  |  |  |  |
|  | Very good/somewhat good | 1.26 | 1.00 |  | 1.33 | 1.00 |
| Relationship with father | (Ref: Very bad/somewhat bad) |  |  |  |  |  |
|  | Very good/somewhat good | 1.53 | 1.00 |  | 1.52 | 1.31 |
| Parent marital status | (Ref: Parents married) |  |  |  |  |  |
|  | Divorced | 1.48 | 1.00 |  | 1.44 | 1.16 |
|  | Single, never married | 2.10 | 1.78 |  | 1.32 | 1.03 |
|  | One or both parents had died | 1.64 | 1.32 |  | 1.37 | 1.14 |
| Subjective financial status of family growing up | (Ref: Got by) |  |  |  |  |  |
|  | Lived comfortably | 1.47 | 1.28 |  | 1.18 | 1.00 |
|  | Found it difficult | 1.14 | 1.00 |  | 1.19 | 1.00 |
|  | Found it very difficult | 1.21 | 1.00 |  | 1.15 | 1.00 |
| Experienced Abuse | (Ref: No) |  |  |  |  |  |
|  | Yes | 1.80 | 1.62 |  | 1.19 | 1.00 |
| Felt like an outsider in the family | (Ref: No) |  |  |  |  |  |
|  | Yes | 1.19 | 1.00 |  | 1.14 | 1.00 |
| Self-rated health growing up | (Ref: Good) |  |  |  |  |  |
|  | Excellent | 1.40 | 1.12 |  | 1.26 | 1.11 |
|  | Very good | 1.30 | 1.00 |  | 1.22 | 1.06 |
|  | Fair | 1.31 | 1.00 |  | 1.17 | 1.00 |
|  | Poor | 1.39 | 1.00 |  | 1.23 | 1.00 |
| Immigration status | (Ref: Born in this country) |  |  |  |  |  |
|  | Born in another country | 1.38 | 1.00 |  | 1.16 | 1.00 |
| Religious service attendance (age 12) | (Ref: Never) |  |  |  |  |  |
|  | At least 1/week | 2.35 | 2.07 |  | 3.62 | 3.42 |
|  | 1-3/month | 2.20 | 1.90 |  | 2.59 | 2.42 |
|  | < 1/month | 1.70 | 1.41 |  | 1.55 | 1.39 |
| Year of birth | (Ref: 1998-2005; age 18-24) |  |  |  |  |  |
|  | 1993-1998; age 25-29 | 1.46 | 1.18 |  | 1.37 | 1.14 |
|  | 1983-1993; age 30-39 | 1.84 | 1.64 |  | 1.40 | 1.21 |
|  | 1973-1983; age 40-49 | 2.22 | 1.97 |  | 1.42 | 1.23 |
|  | 1963-1973; age 50-59 | 2.23 | 1.95 |  | 1.57 | 1.39 |
|  | 1953-1963; age 60-69 | 2.20 | 1.86 |  | 1.70 | 1.51 |
|  | 1943-1953; age 70-79 | 2.09 | 1.53 |  | 1.69 | 1.46 |
|  | 1943 or earlier; age 80+ | 8.07 | 5.73 |  | 2.01 | 1.63 |
| Gender | (Ref: Male) |  |  |  |  |  |
|  | Female | 2.19 | 2.03 |  | 1.20 | 1.08 |
|  | Other | 5.81 | 4.12 |  | 6.30 | 4.56 |

| **Secular Community Participation** | **Religious Service Attendance** |
| --- | --- |
| 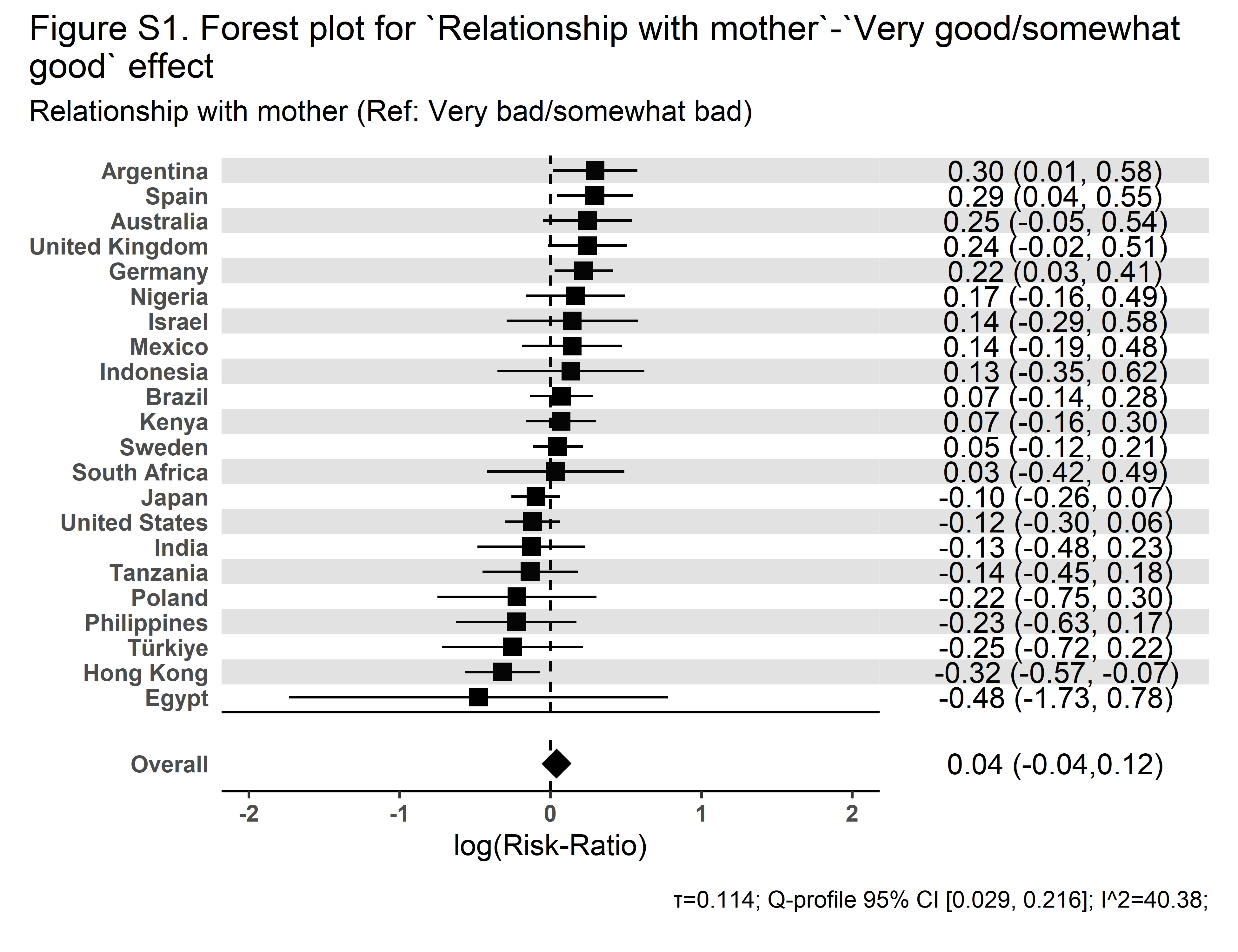 | 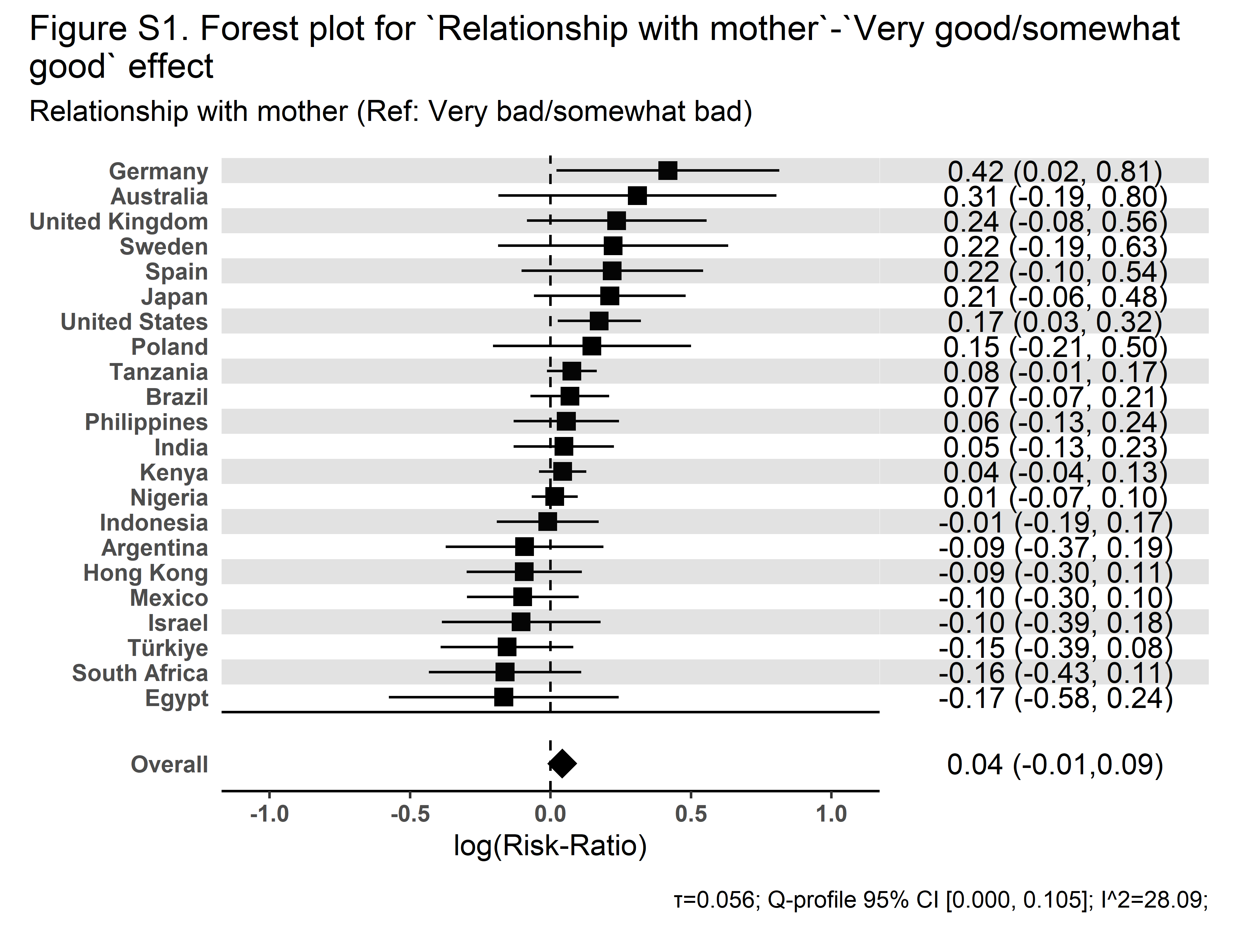 |

| **Secular Community Participation** | **Religious Service Attendance** |
| --- | --- |
| 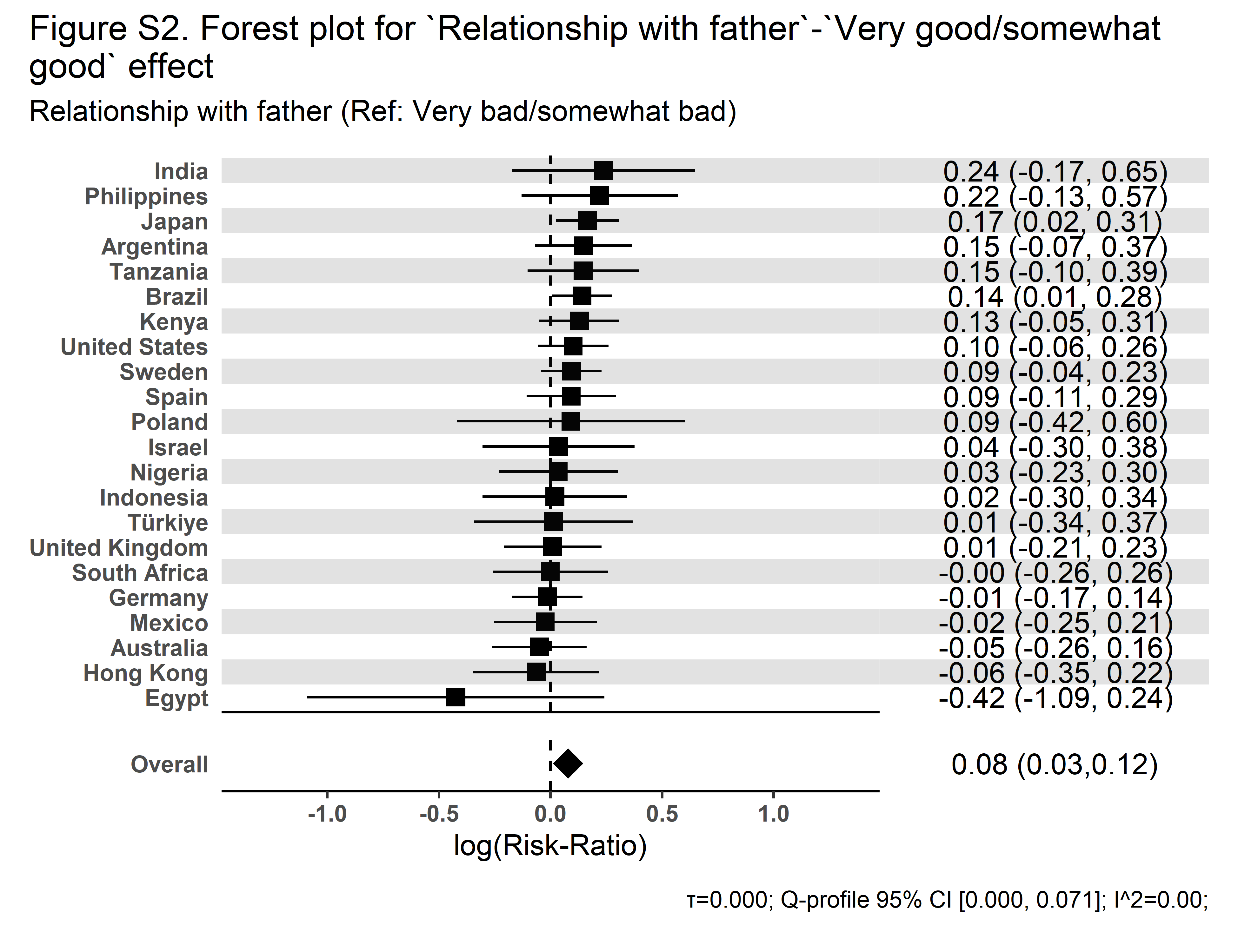 | 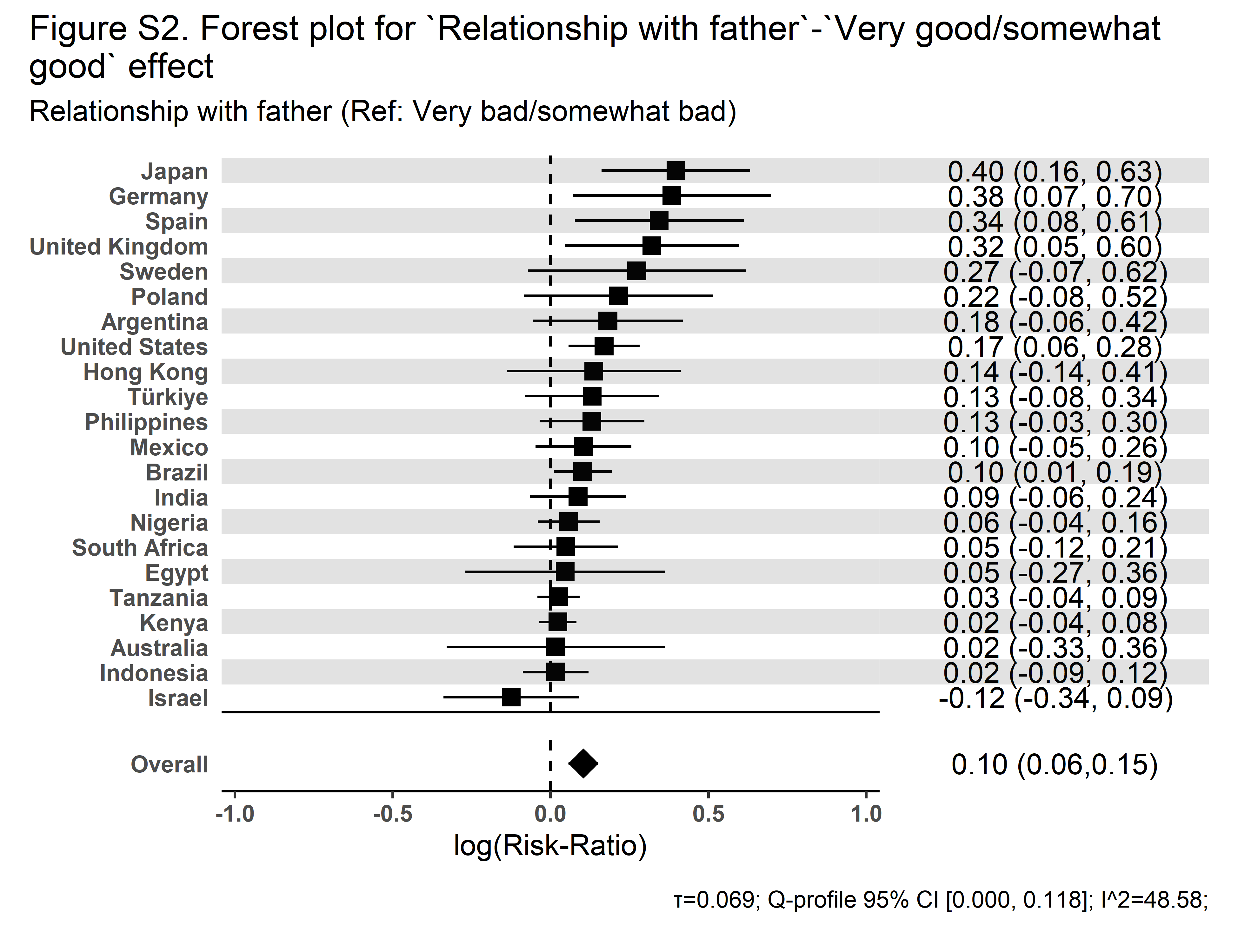 |

| **Secular Community Participation** | **Religious Service Attendance** |
| --- | --- |
| 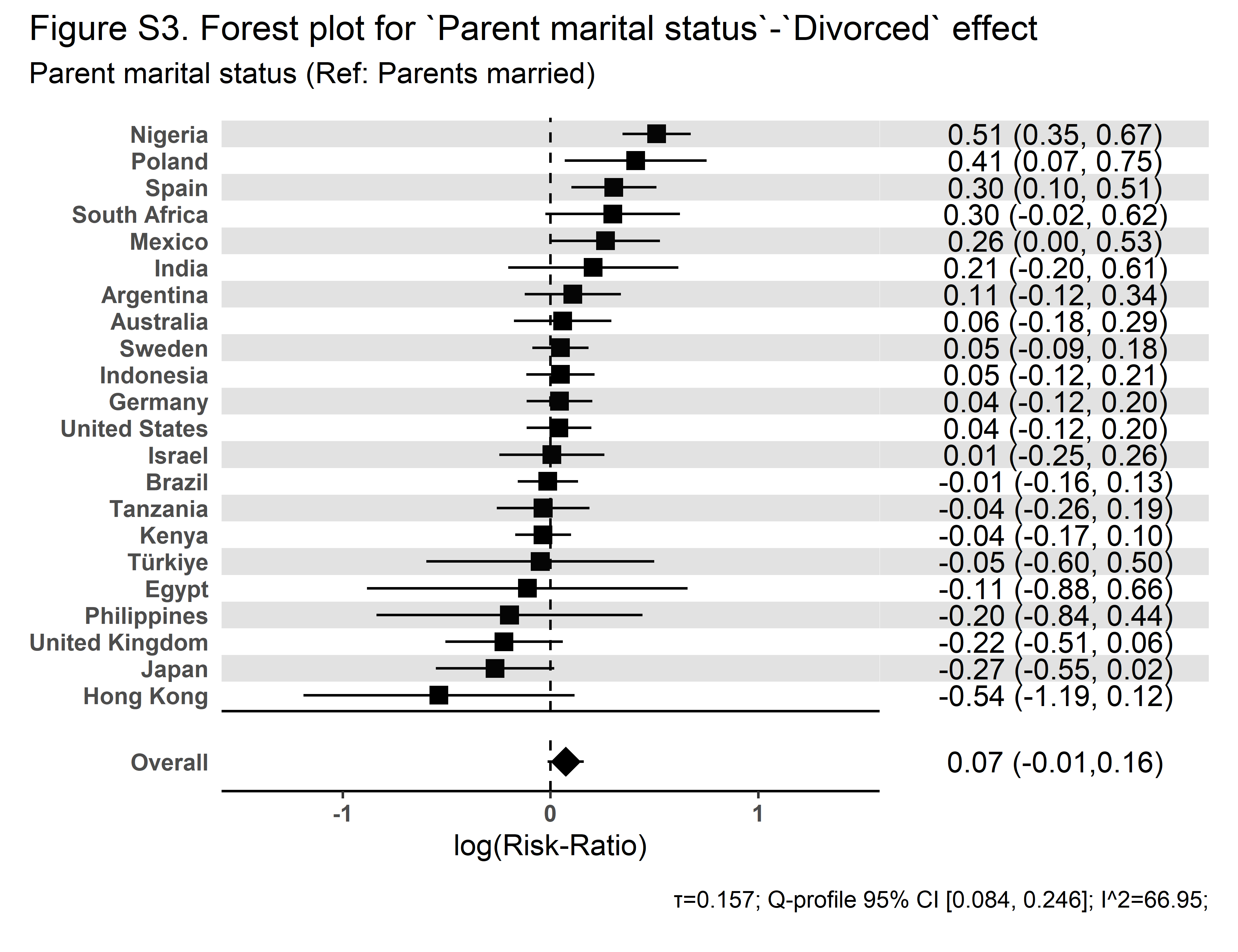 | 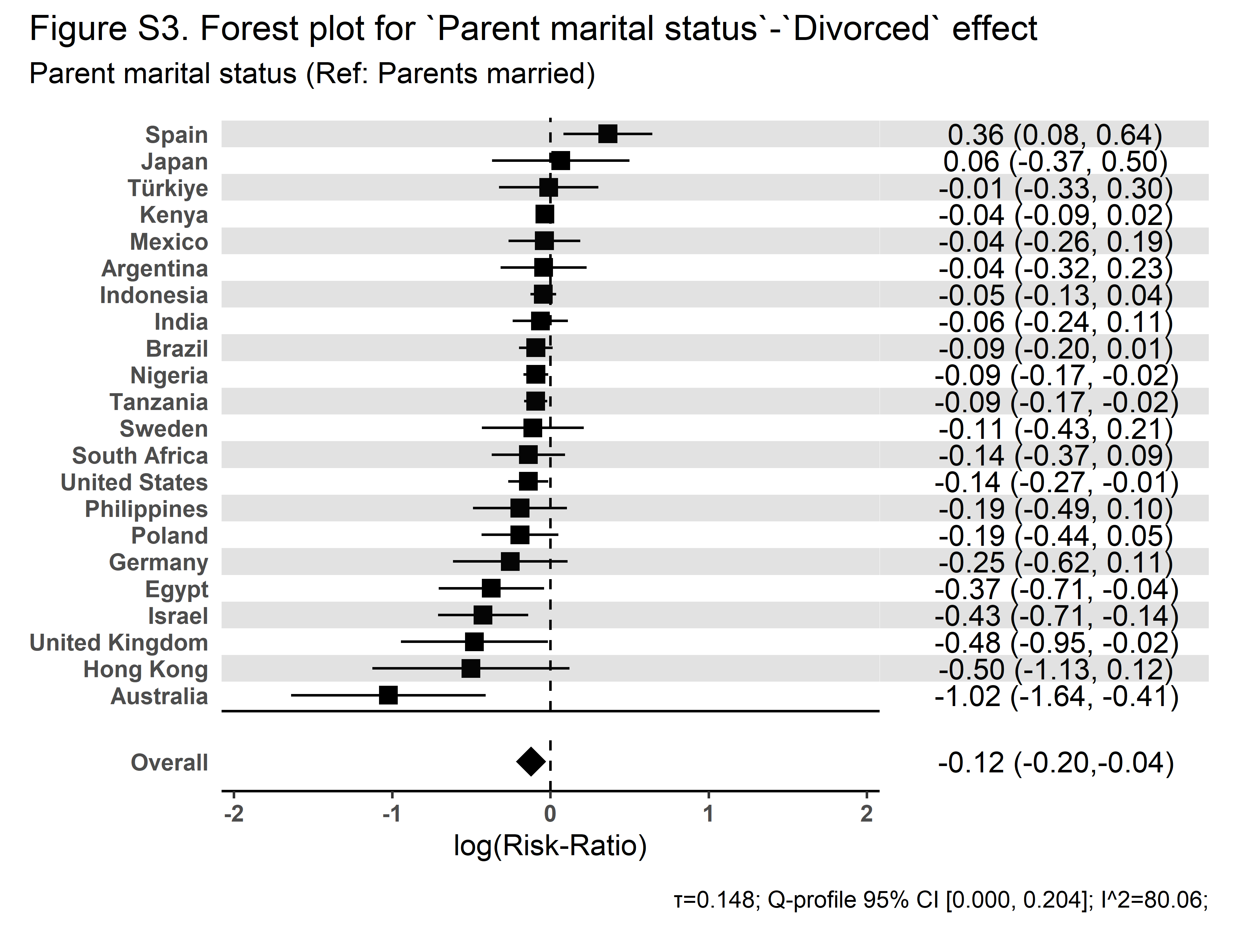 |

| **Secular Community Participation** | **Religious Service Attendance** |
| --- | --- |
| 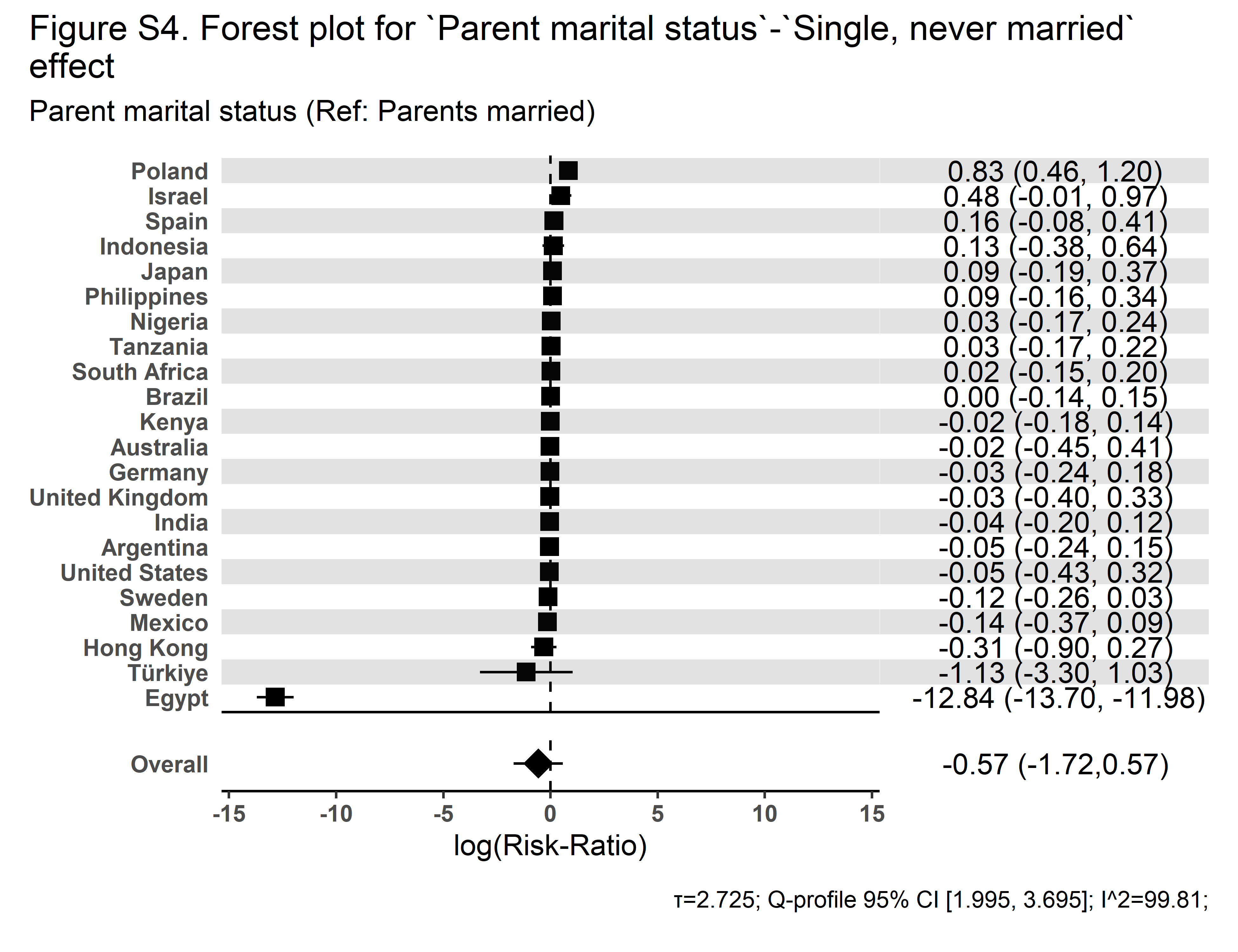 | 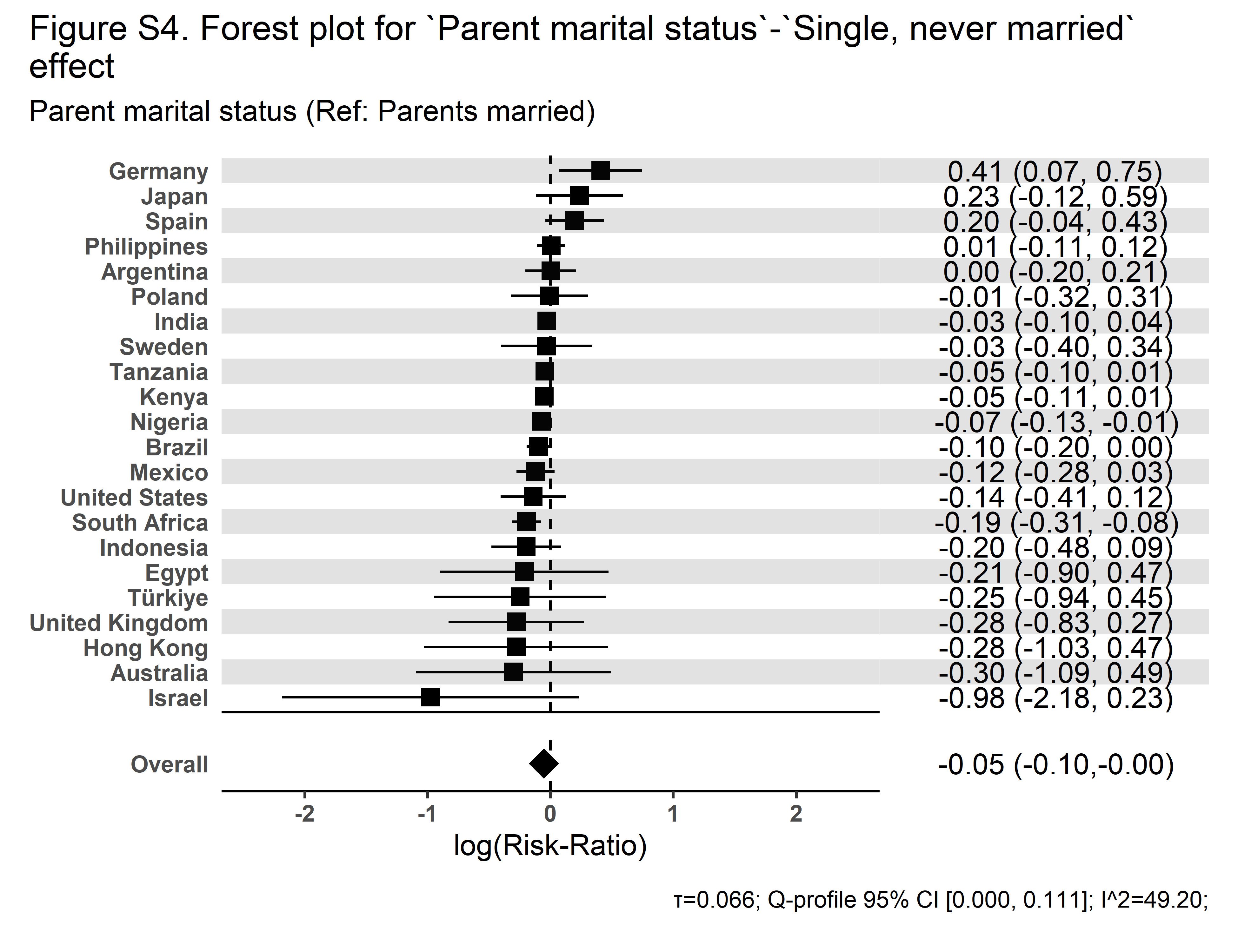 |

| **Secular Community Participation** | **Religious Service Attendance** |
| --- | --- |
| 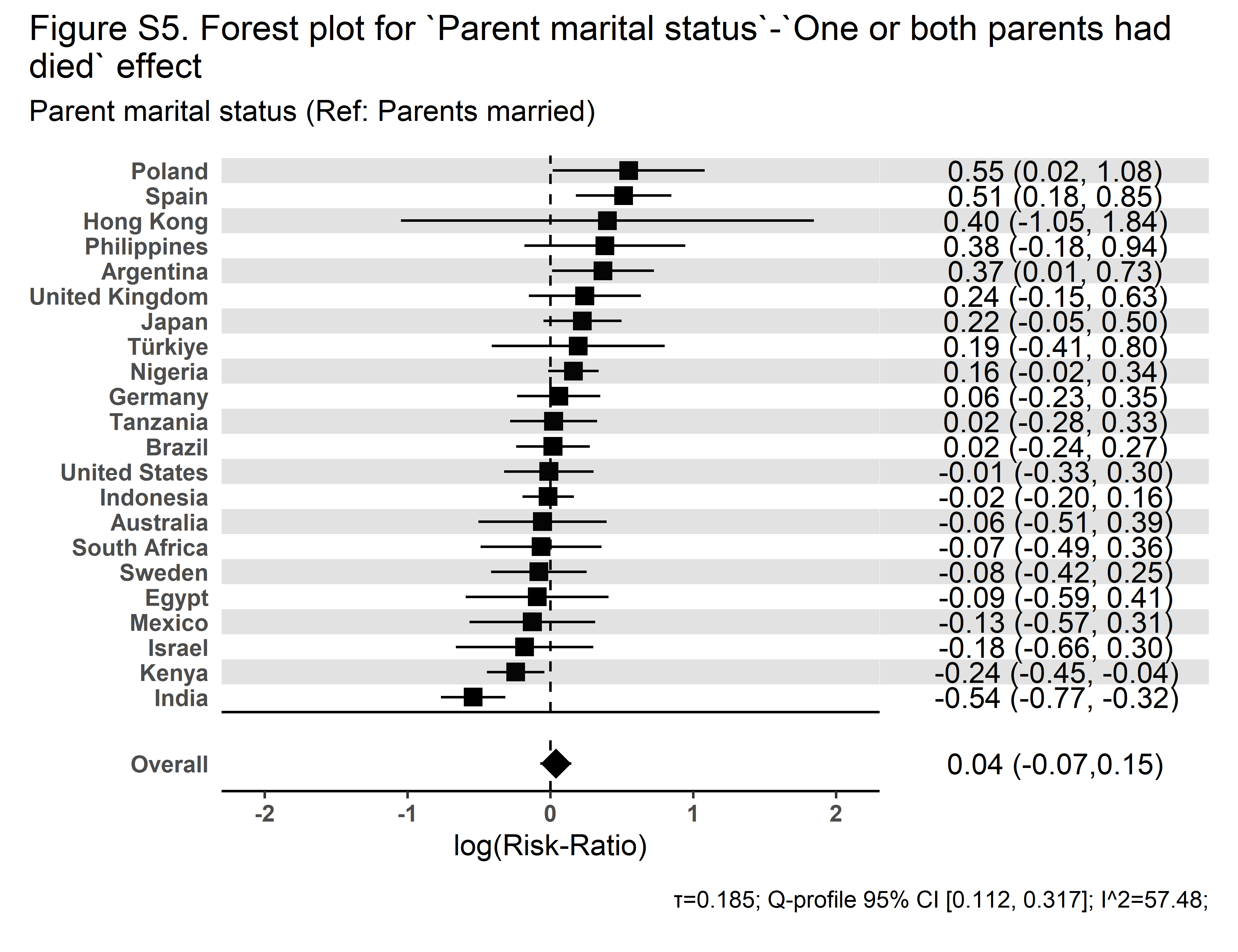 | 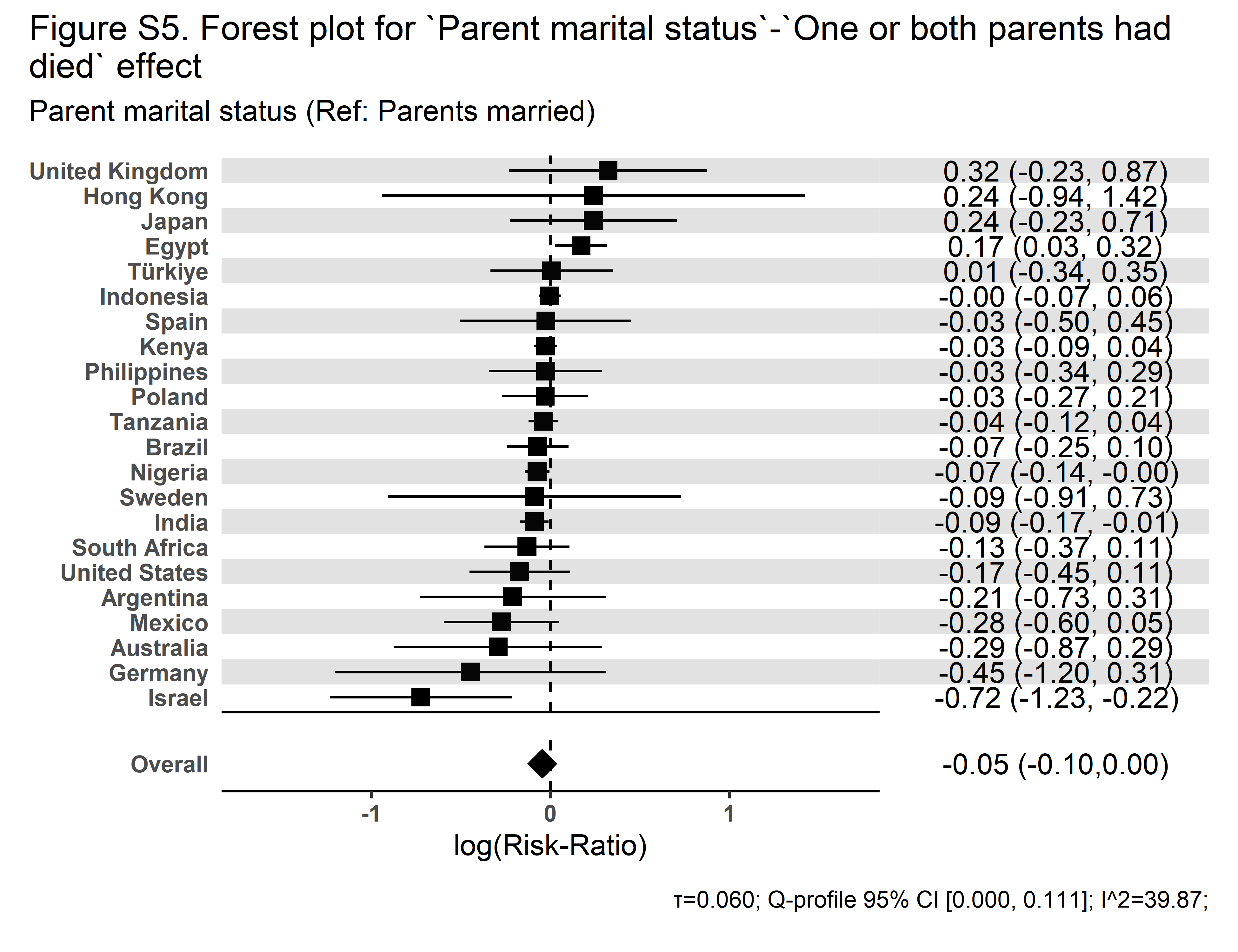 |

| **Secular Community Participation** | **Religious Service Attendance** |
| --- | --- |
| 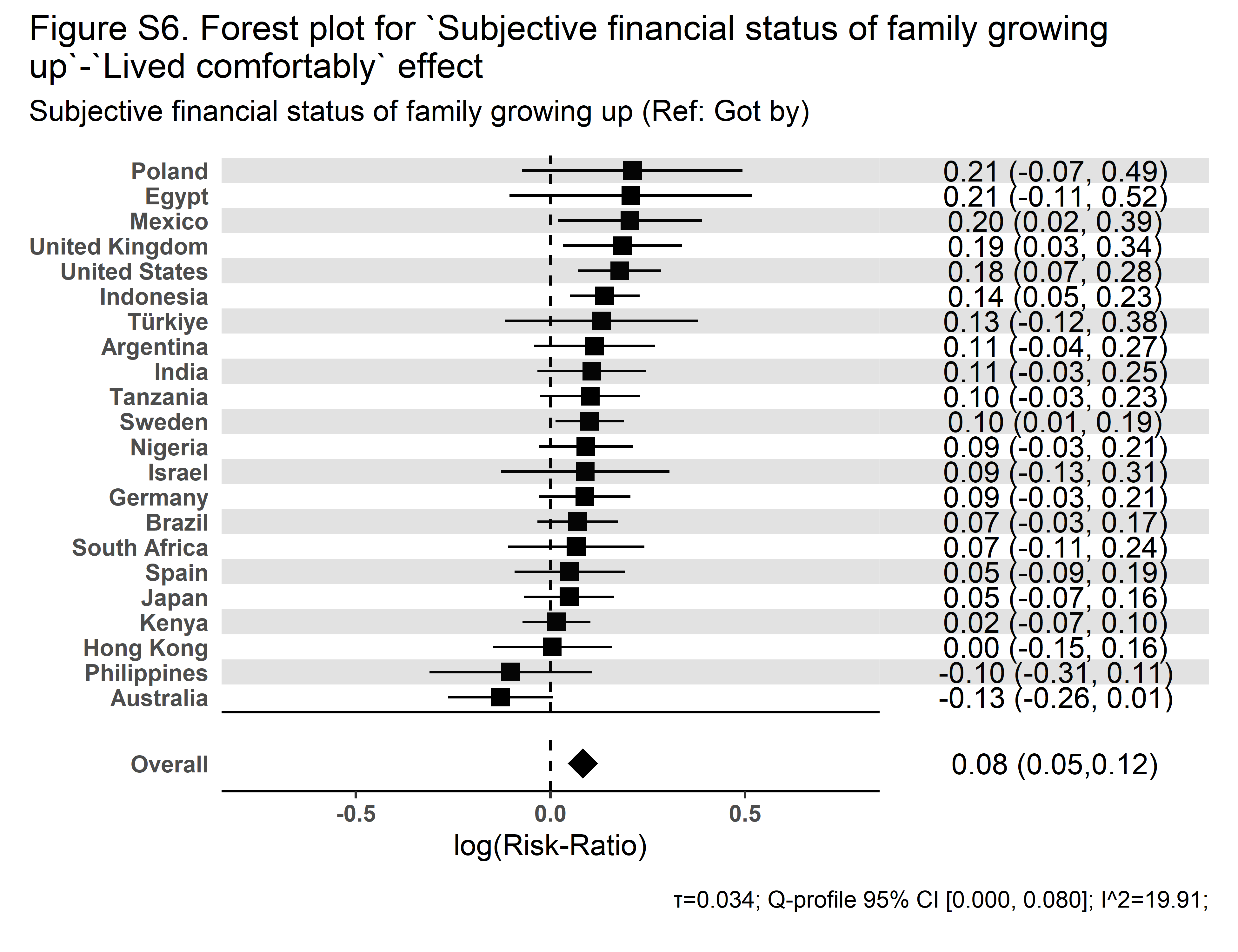 | 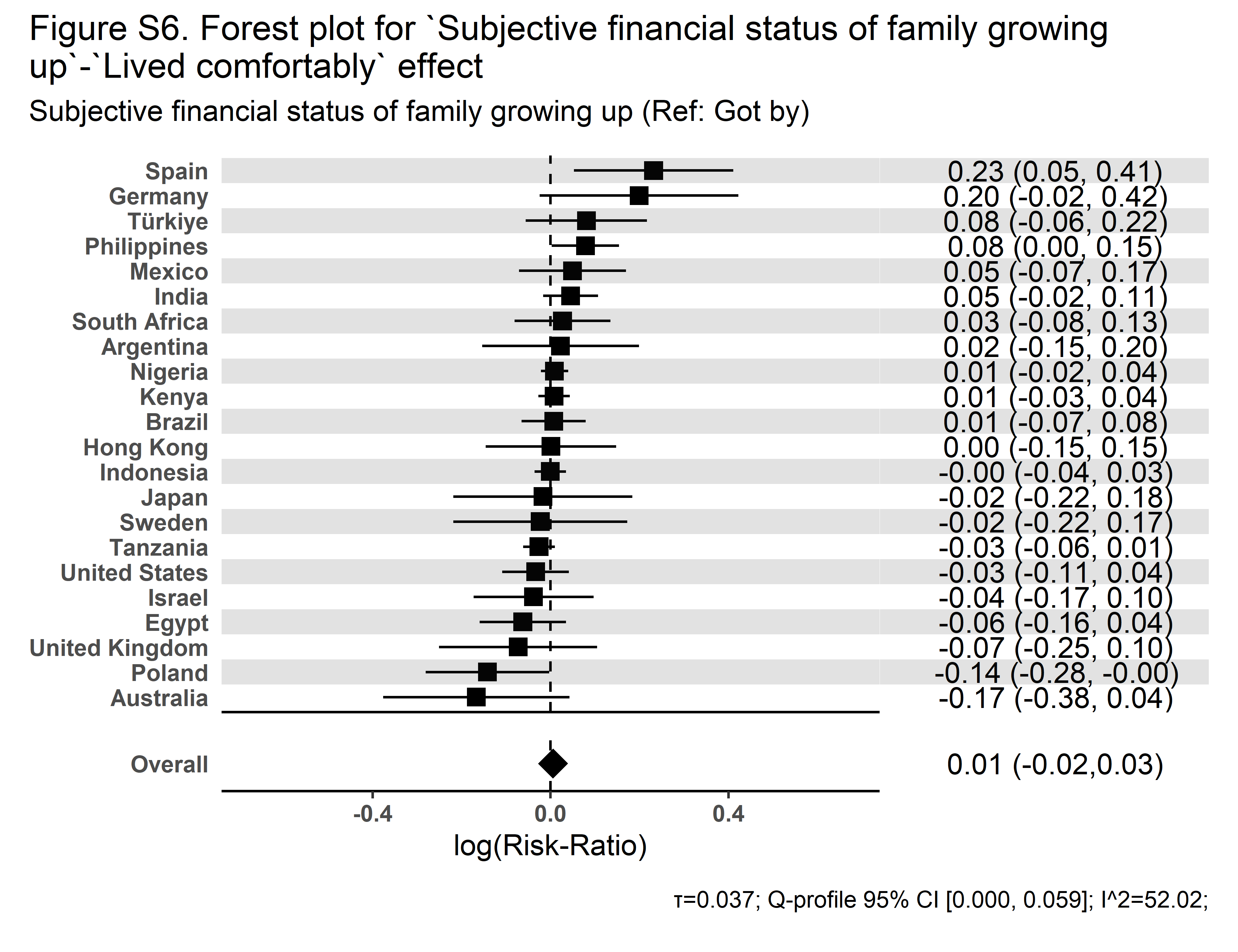 |

| **Secular Community Participation** | **Religious Service Attendance** |
| --- | --- |
| 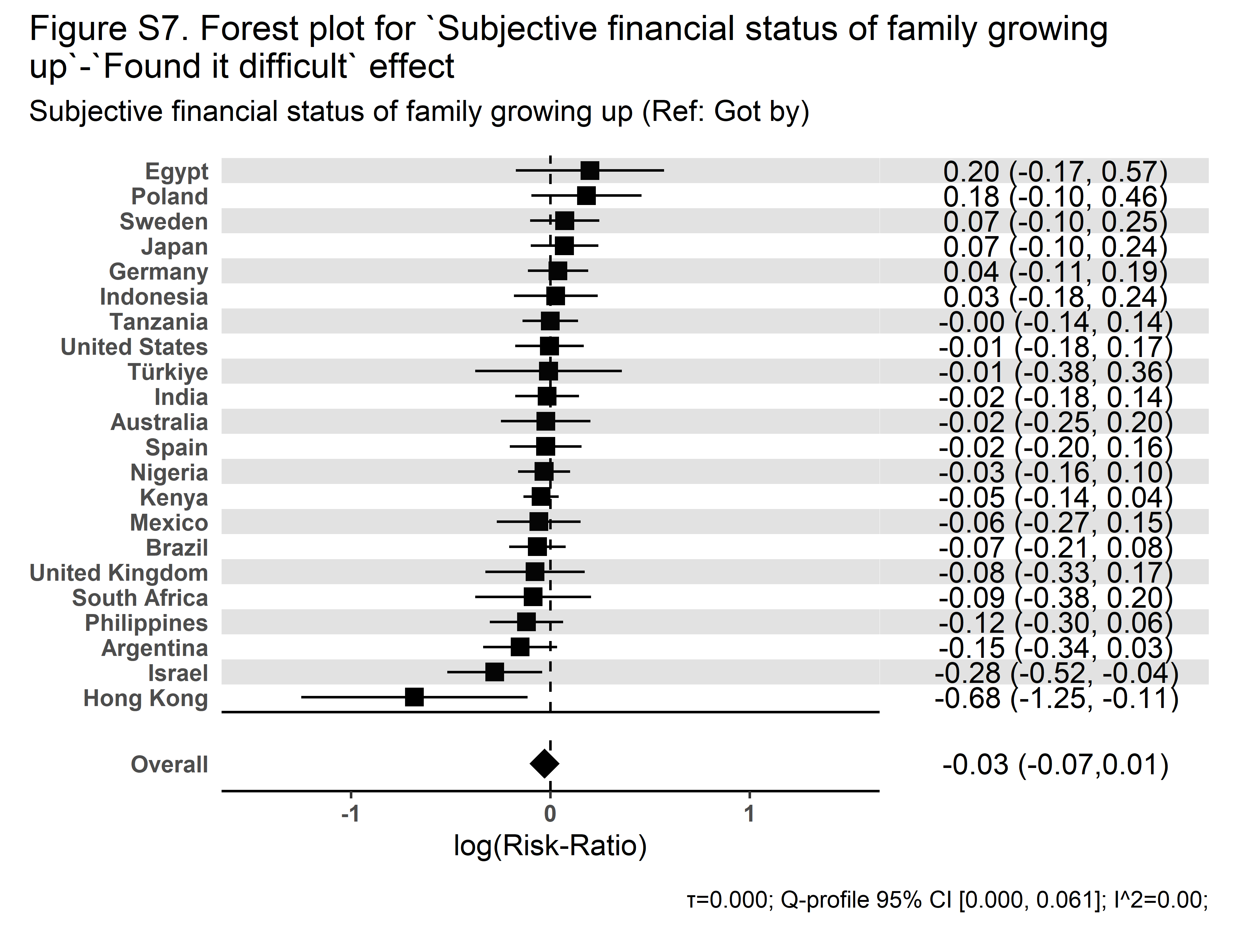 | 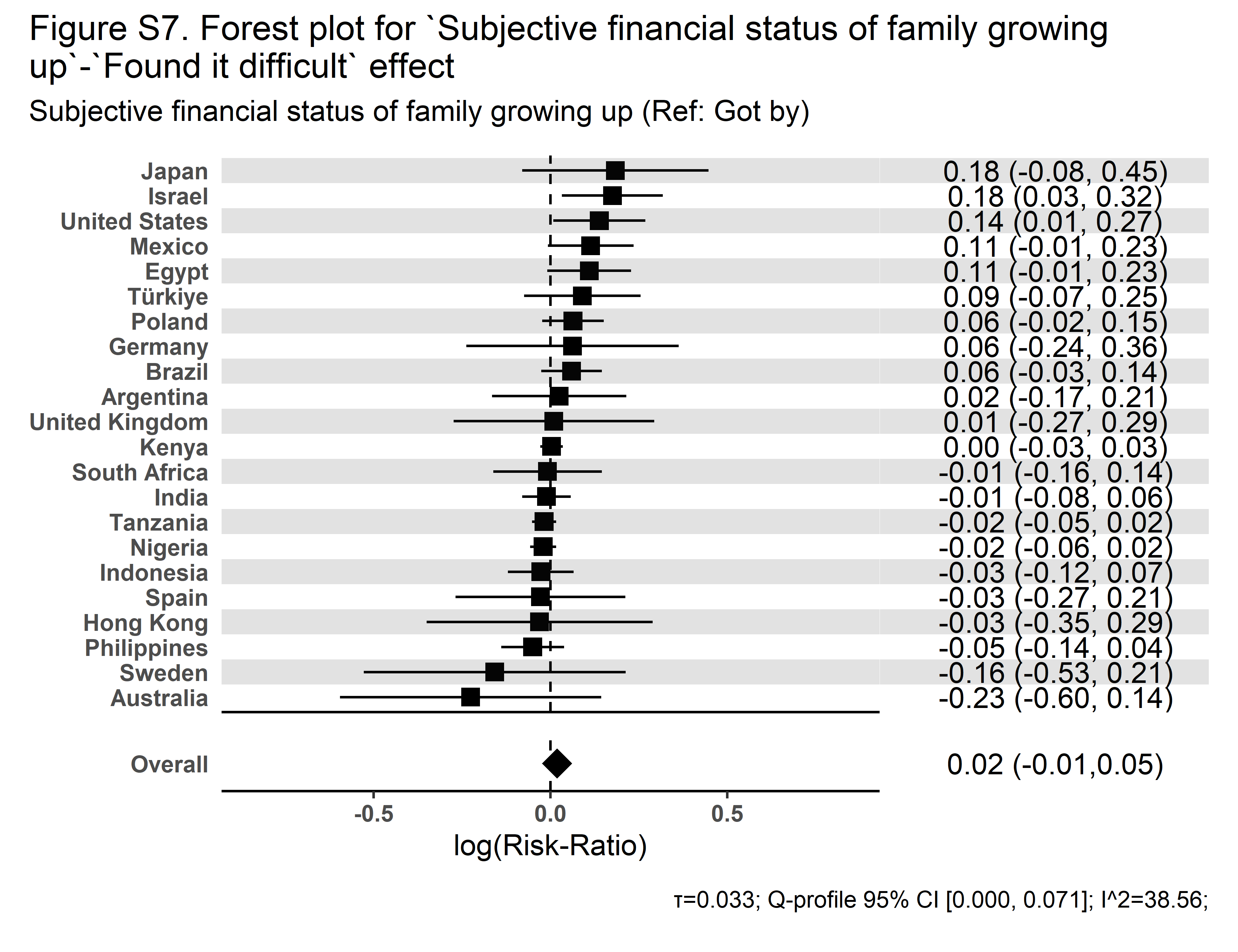 |

| **Secular Community Participation** | **Religious Service Attendance** |
| --- | --- |
| 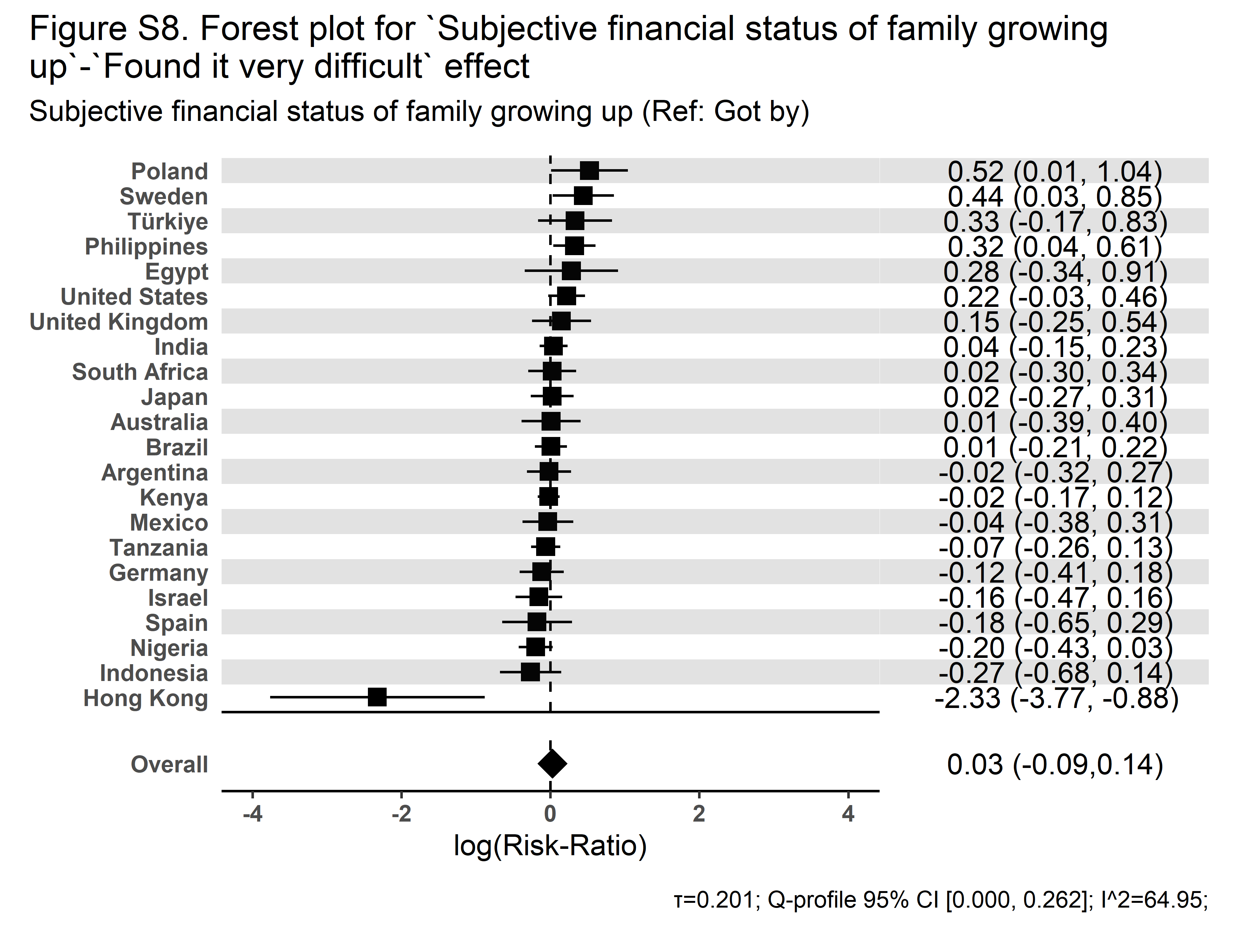 | 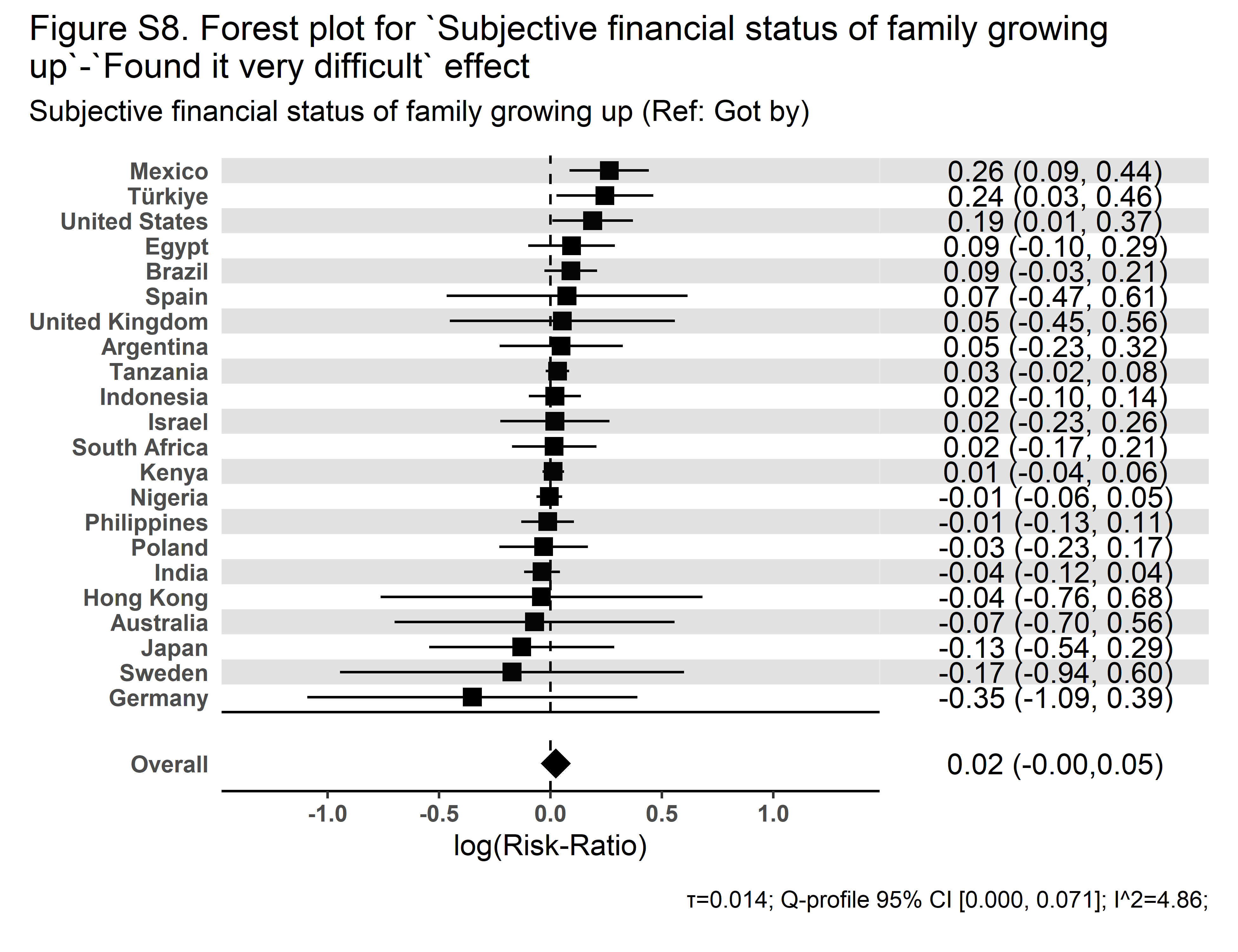 |

| **Secular Community Participation** | **Religious Service Attendance** |
| --- | --- |
| 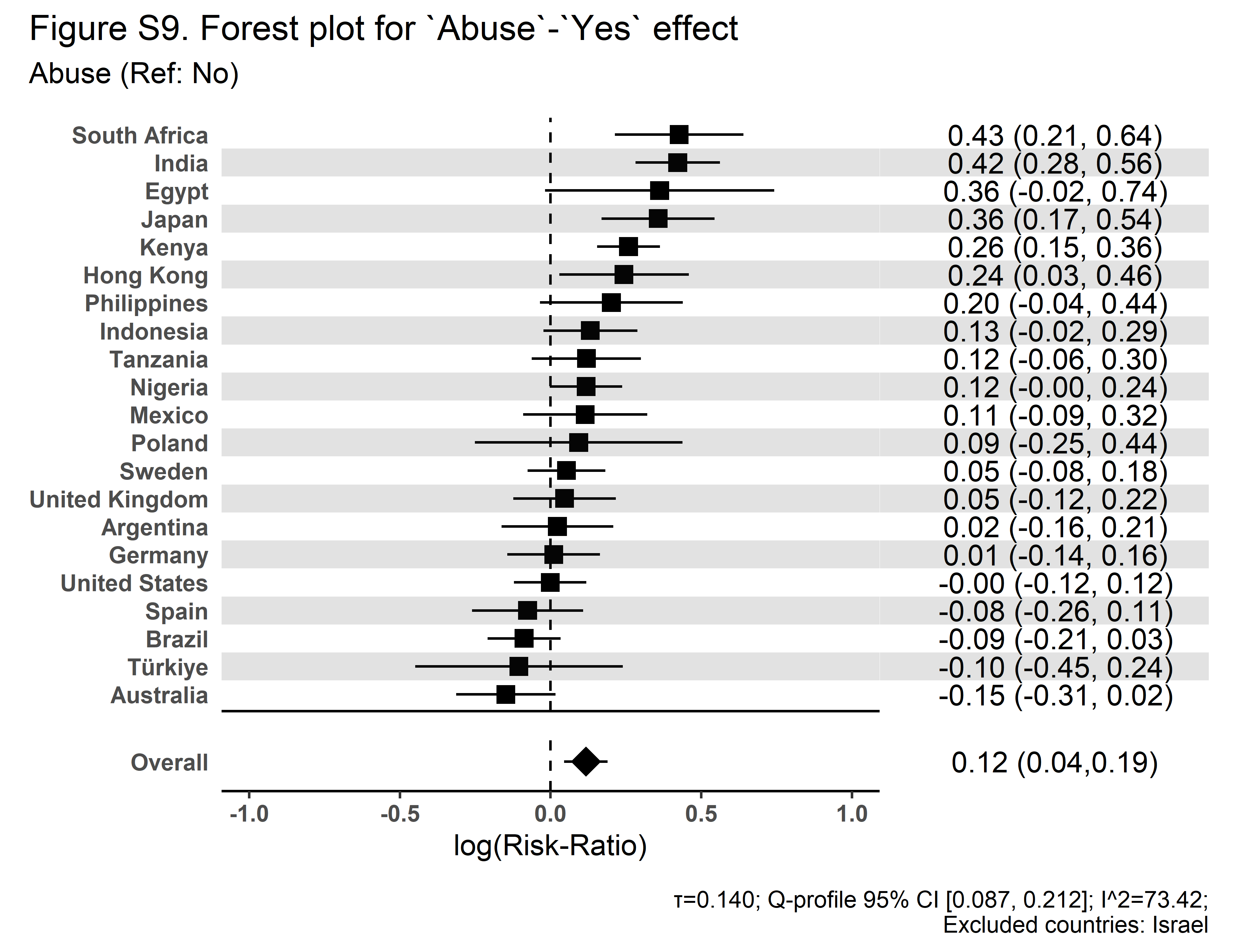 | 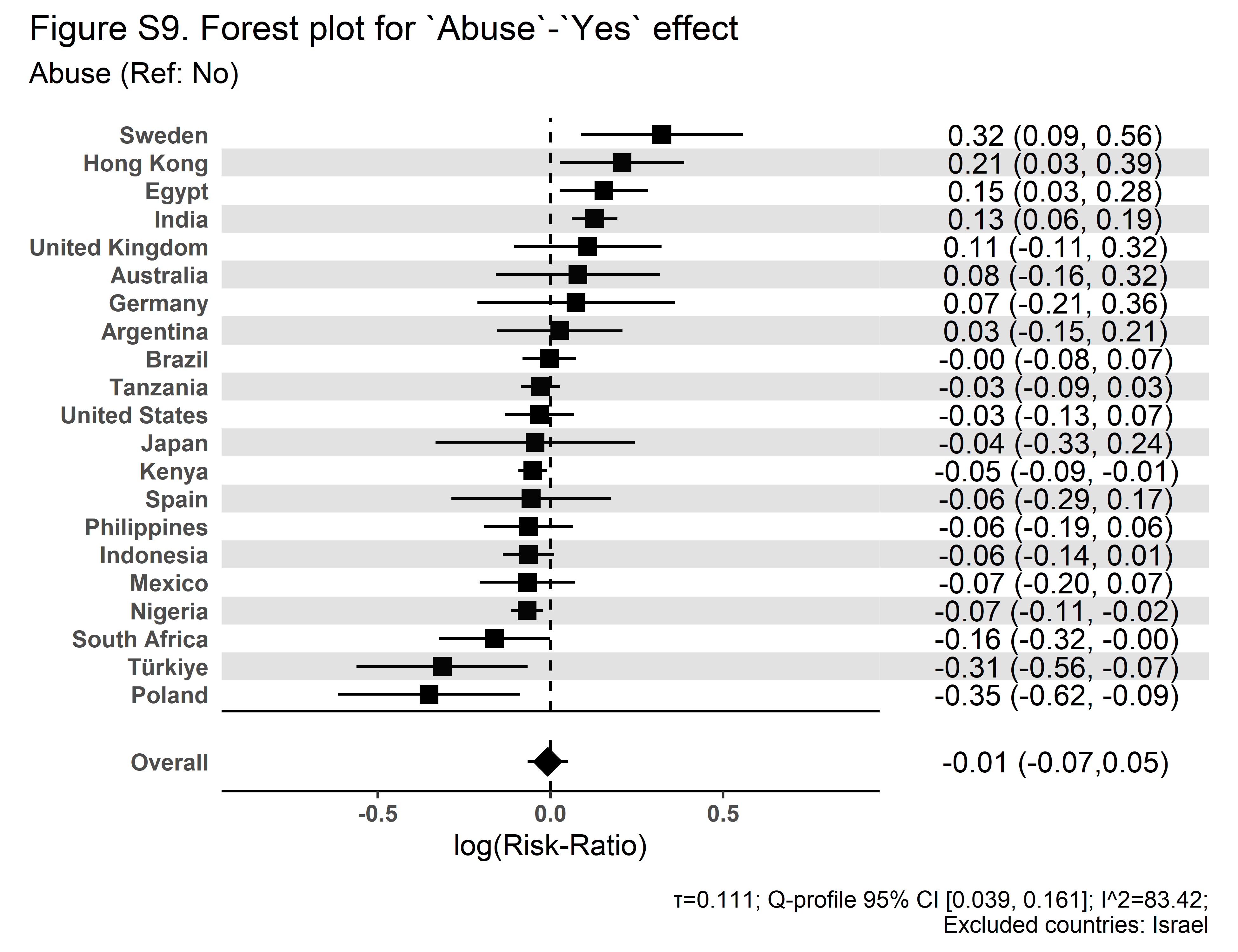 |

| **Secular Community Participation** | **Religious Service Attendance** |
| --- | --- |
| 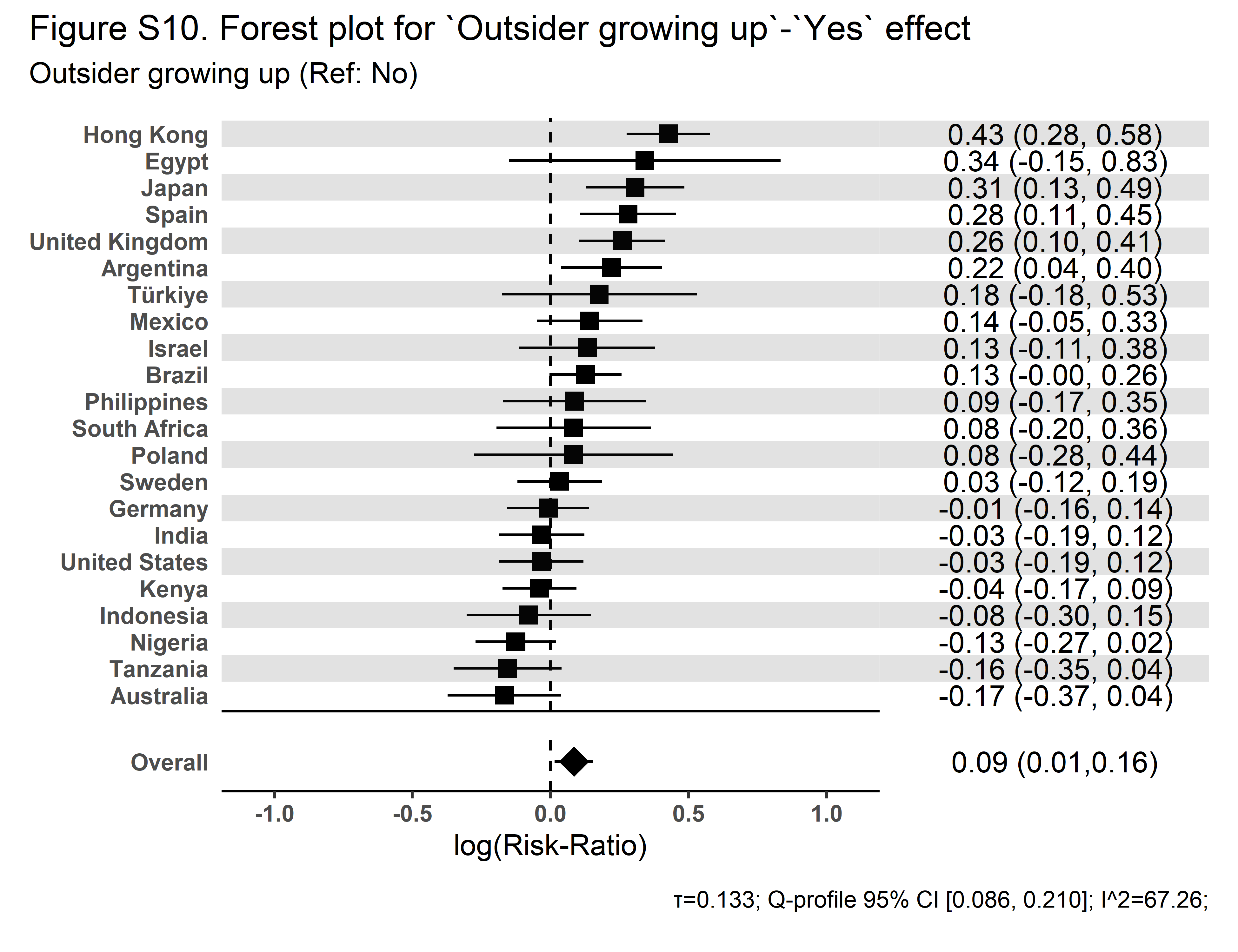 | 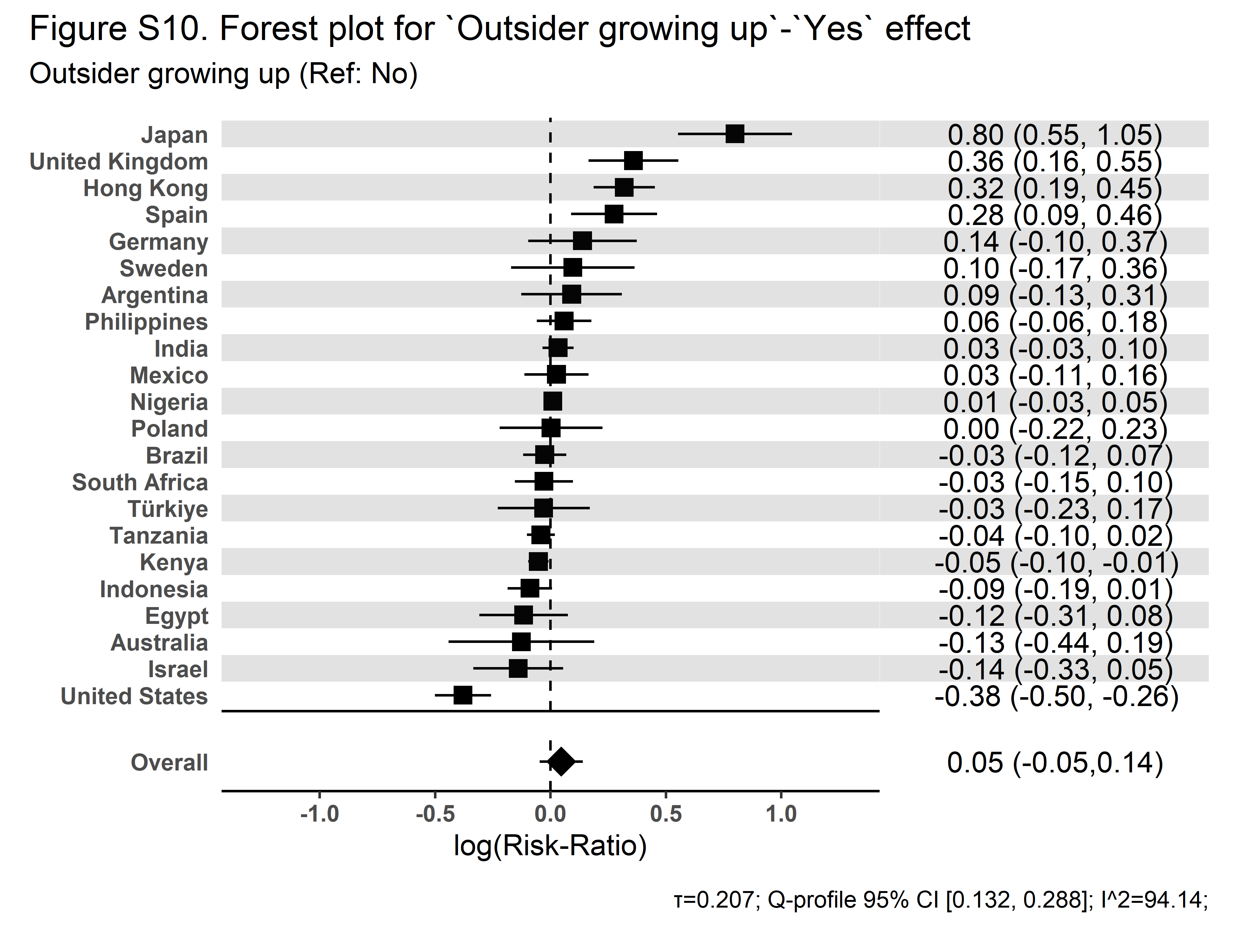 |

| **Secular Community Participation** | **Religious Service Attendance** |
| --- | --- |
| 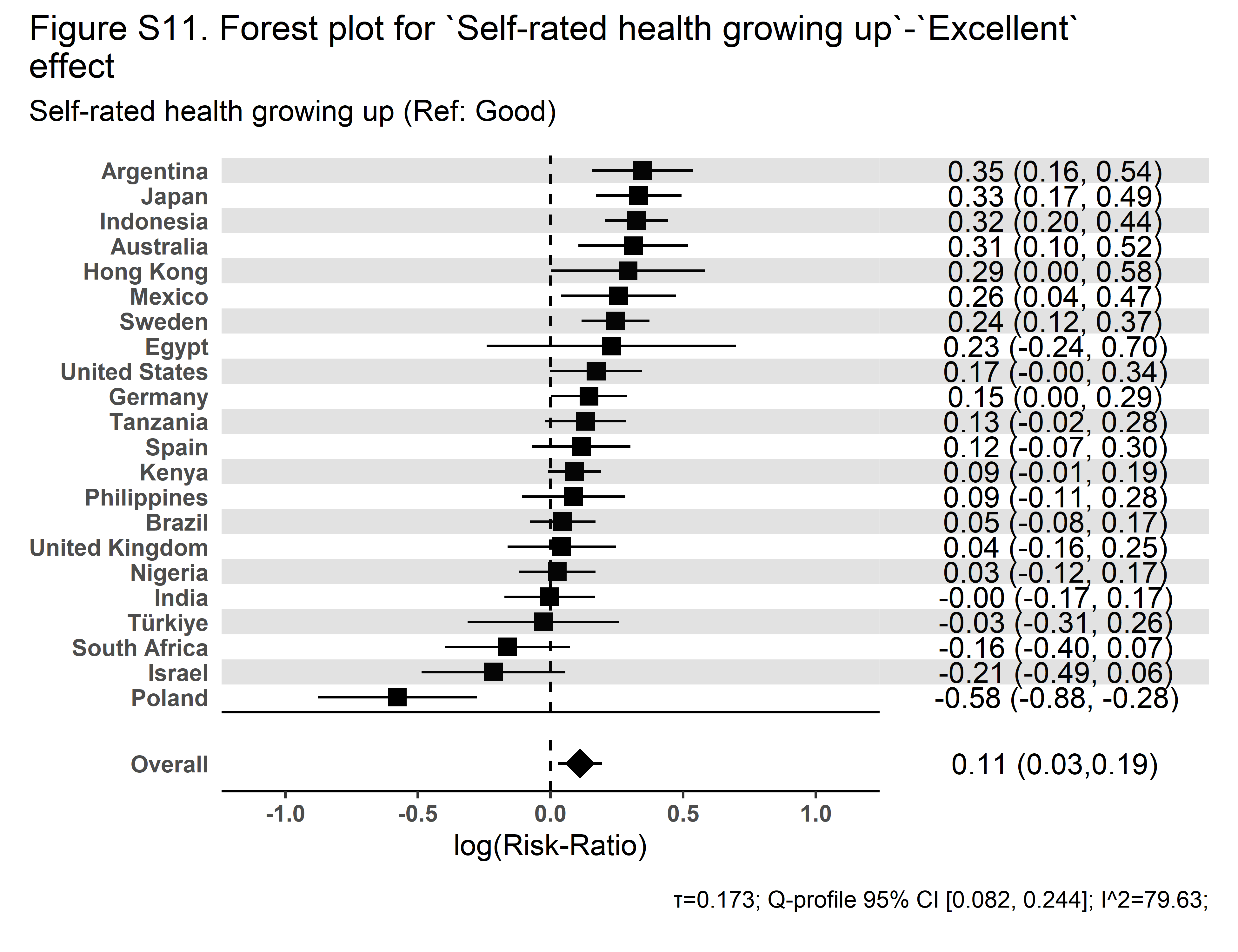 | 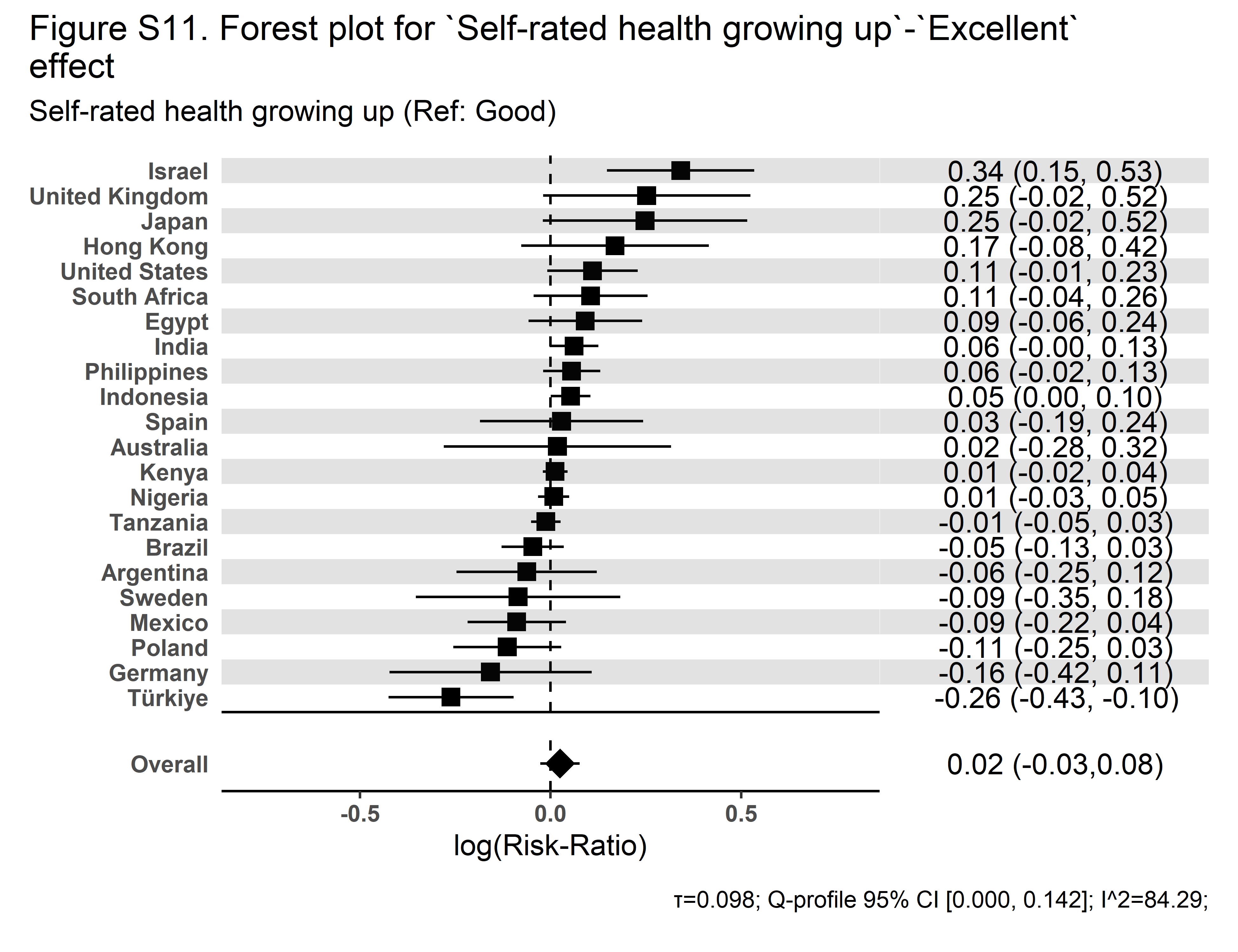 |

| **Secular Community Participation** | **Religious Service Attendance** |
| --- | --- |
| 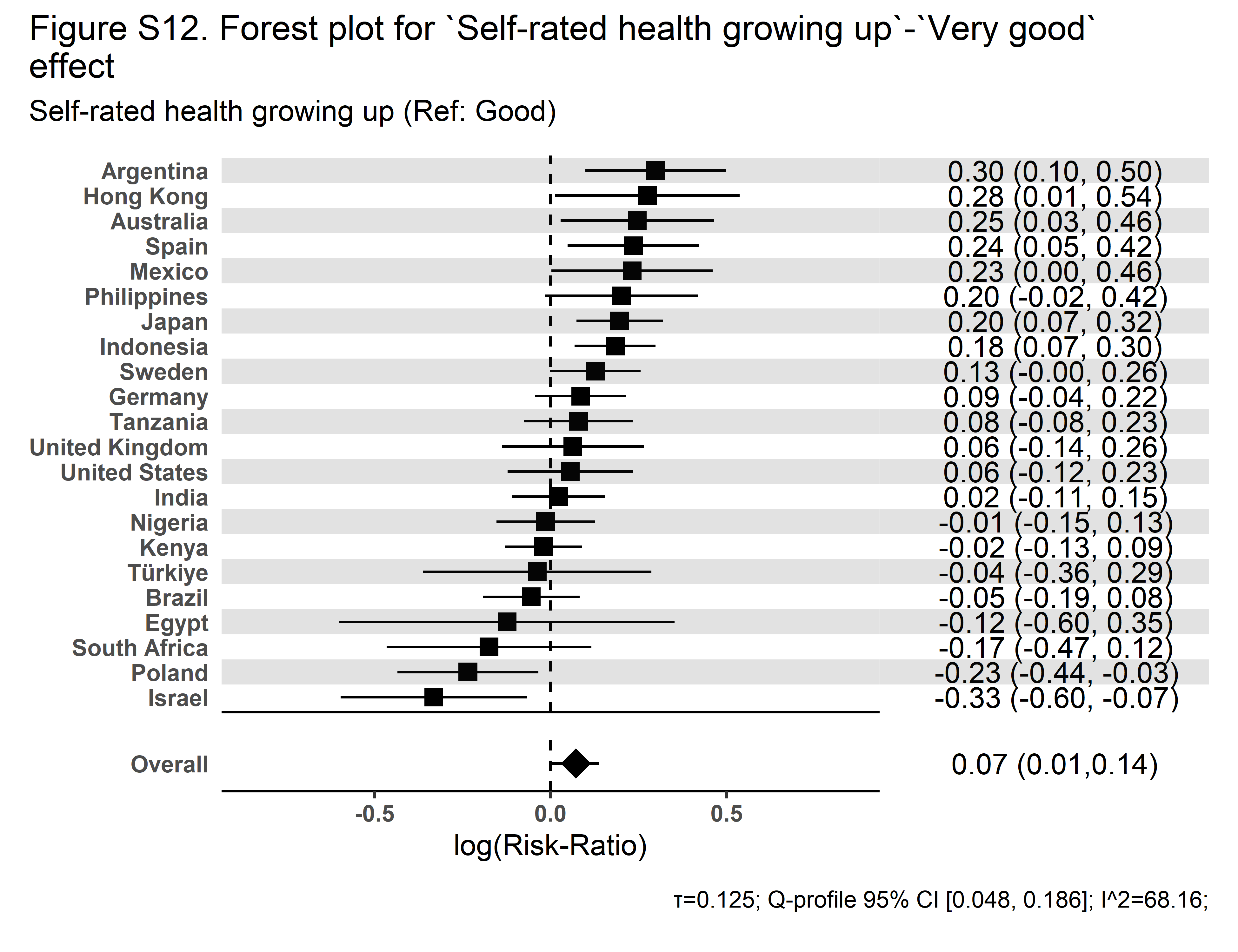 | 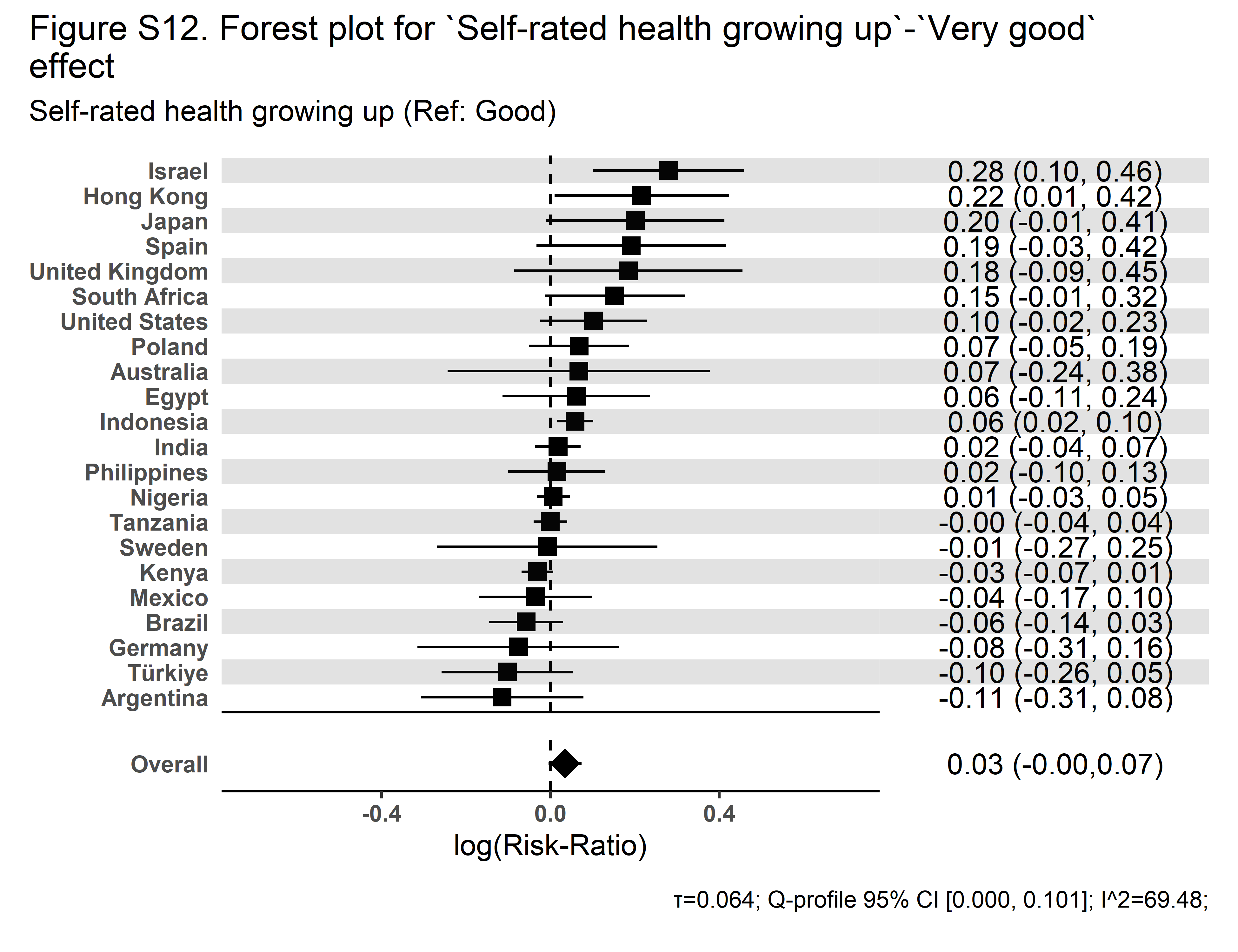 |

| **Secular Community Participation** | **Religious Service Attendance** |
| --- | --- |
| 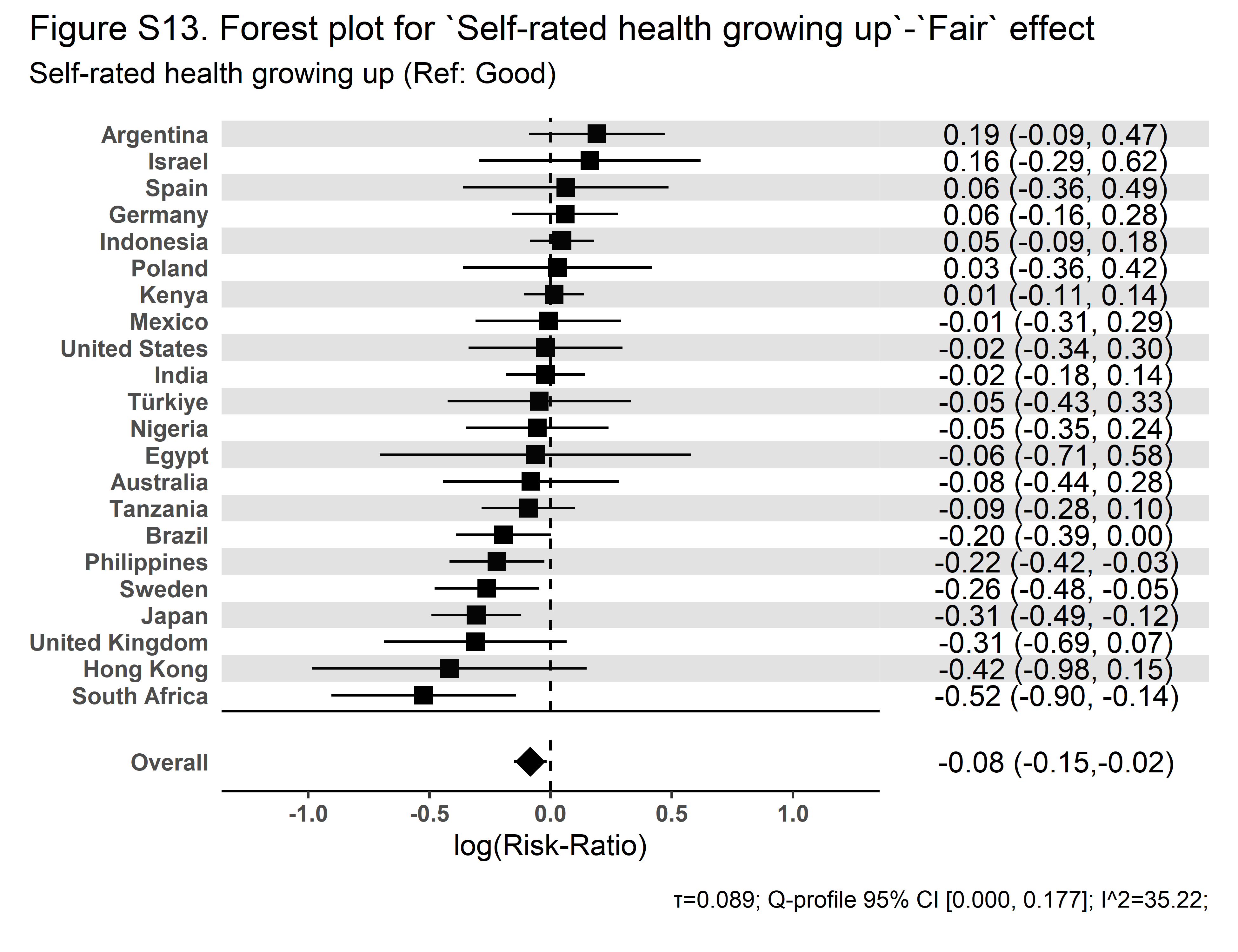 | 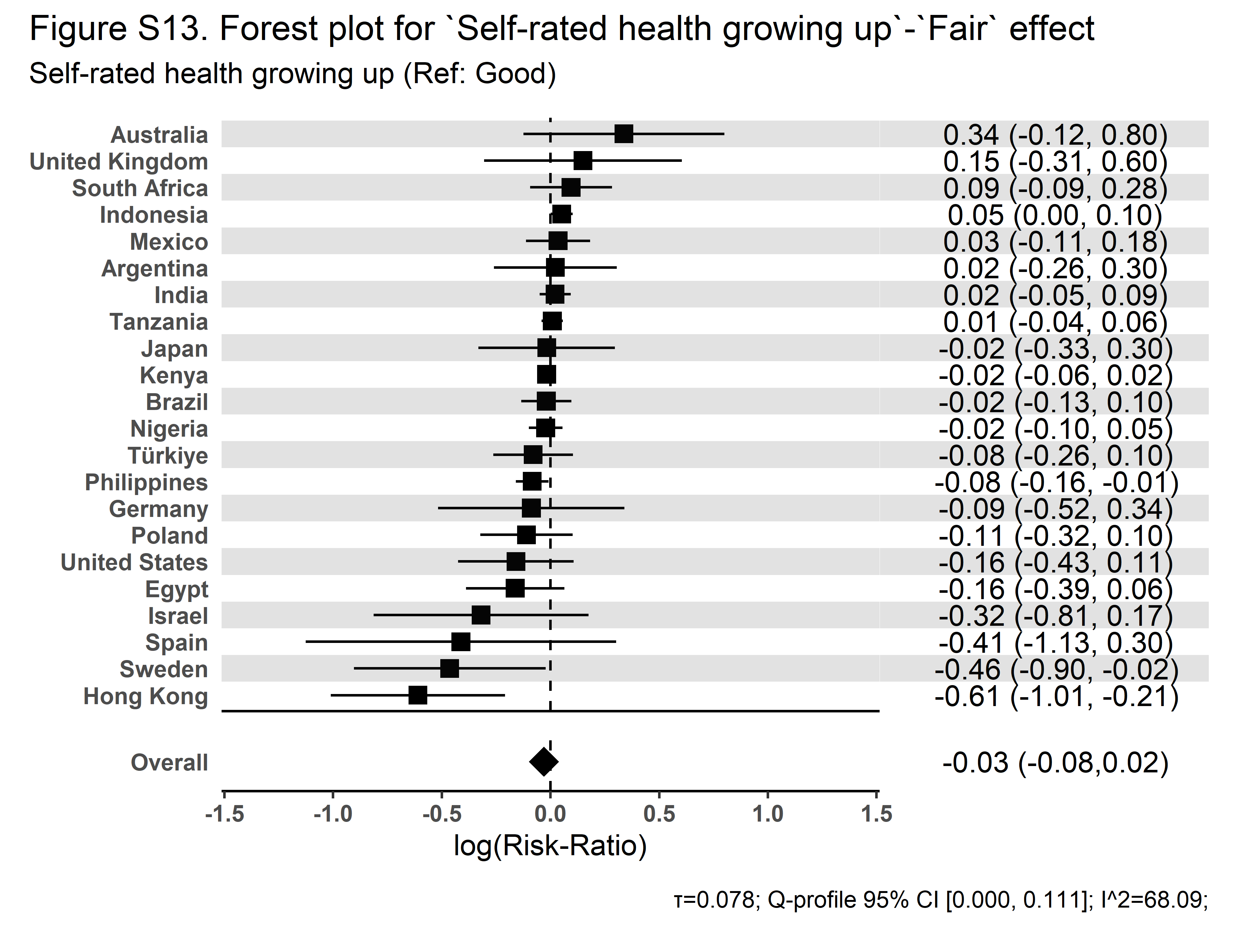 |

| **Secular Community Participation** | **Religious Service Attendance** |
| --- | --- |
| 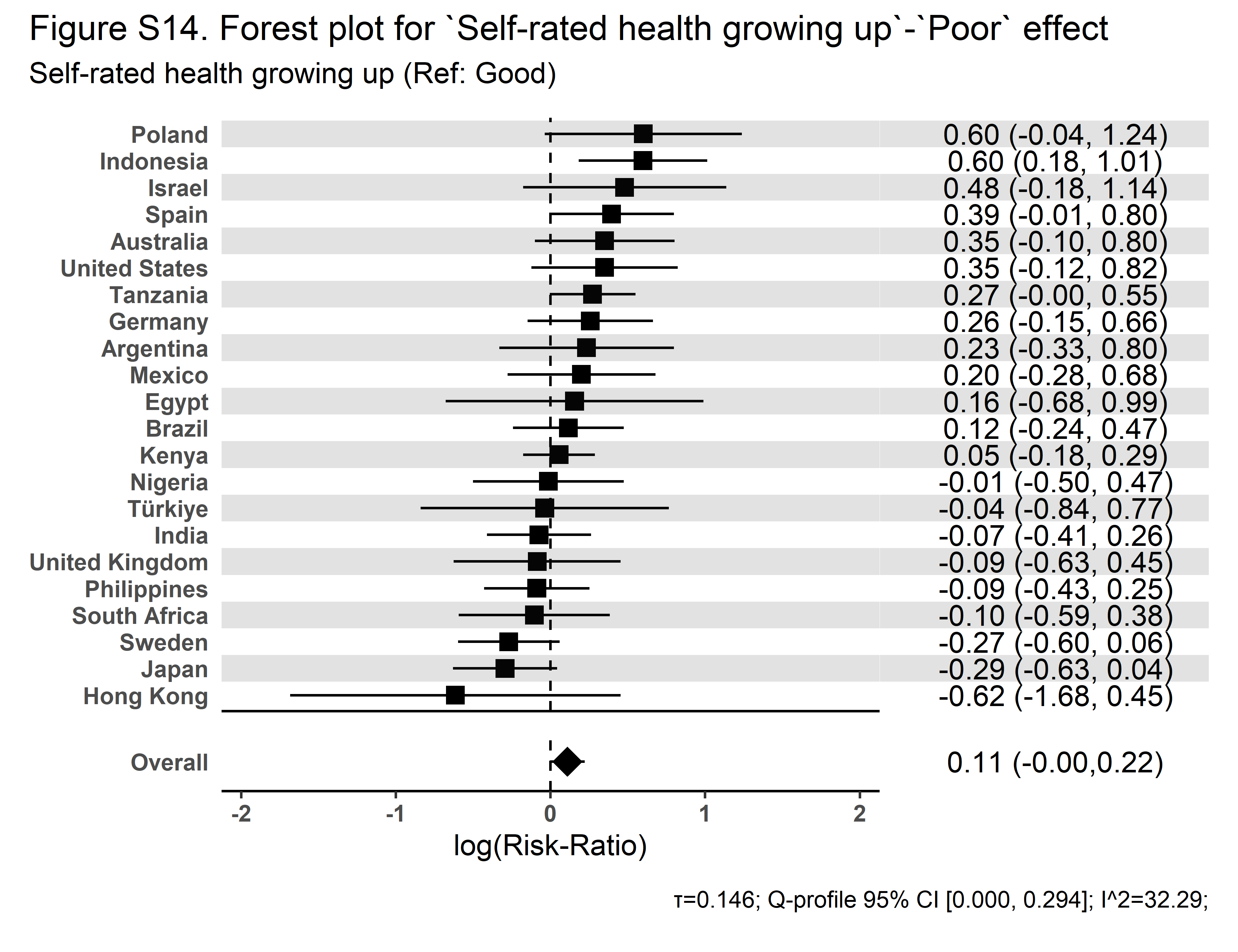 | 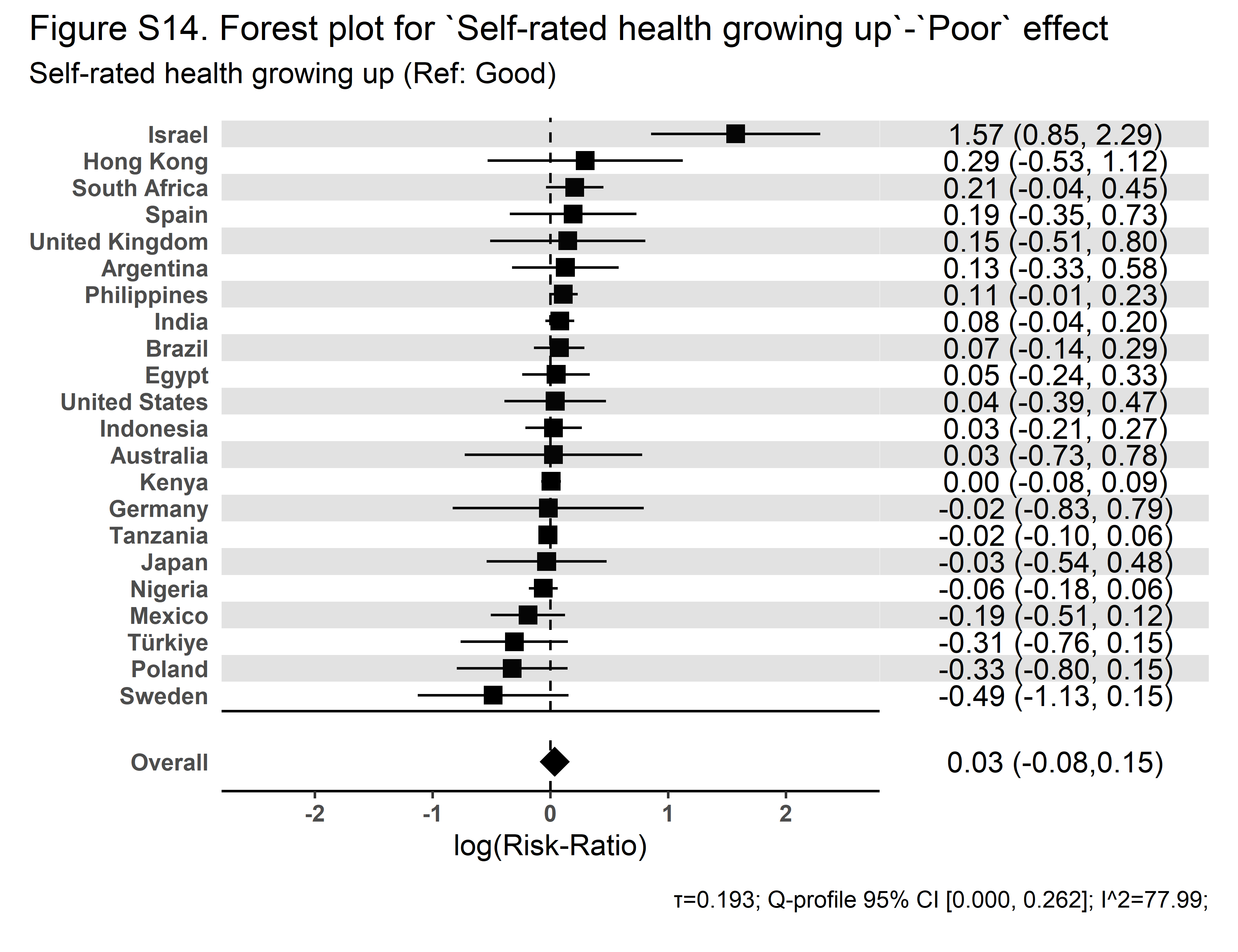 |

| **Secular Community Participation** | **Religious Service Attendance** |
| --- | --- |
| 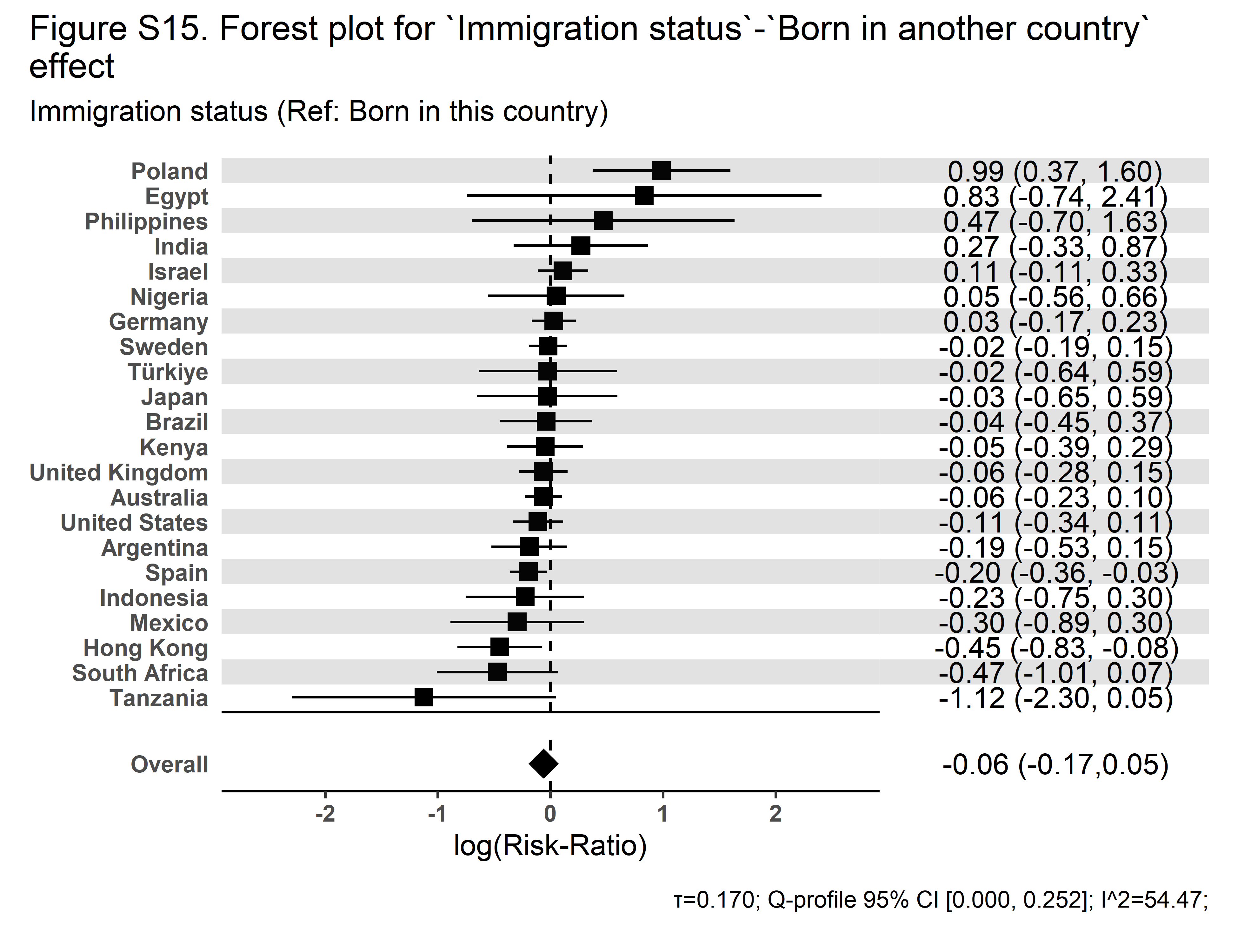 | 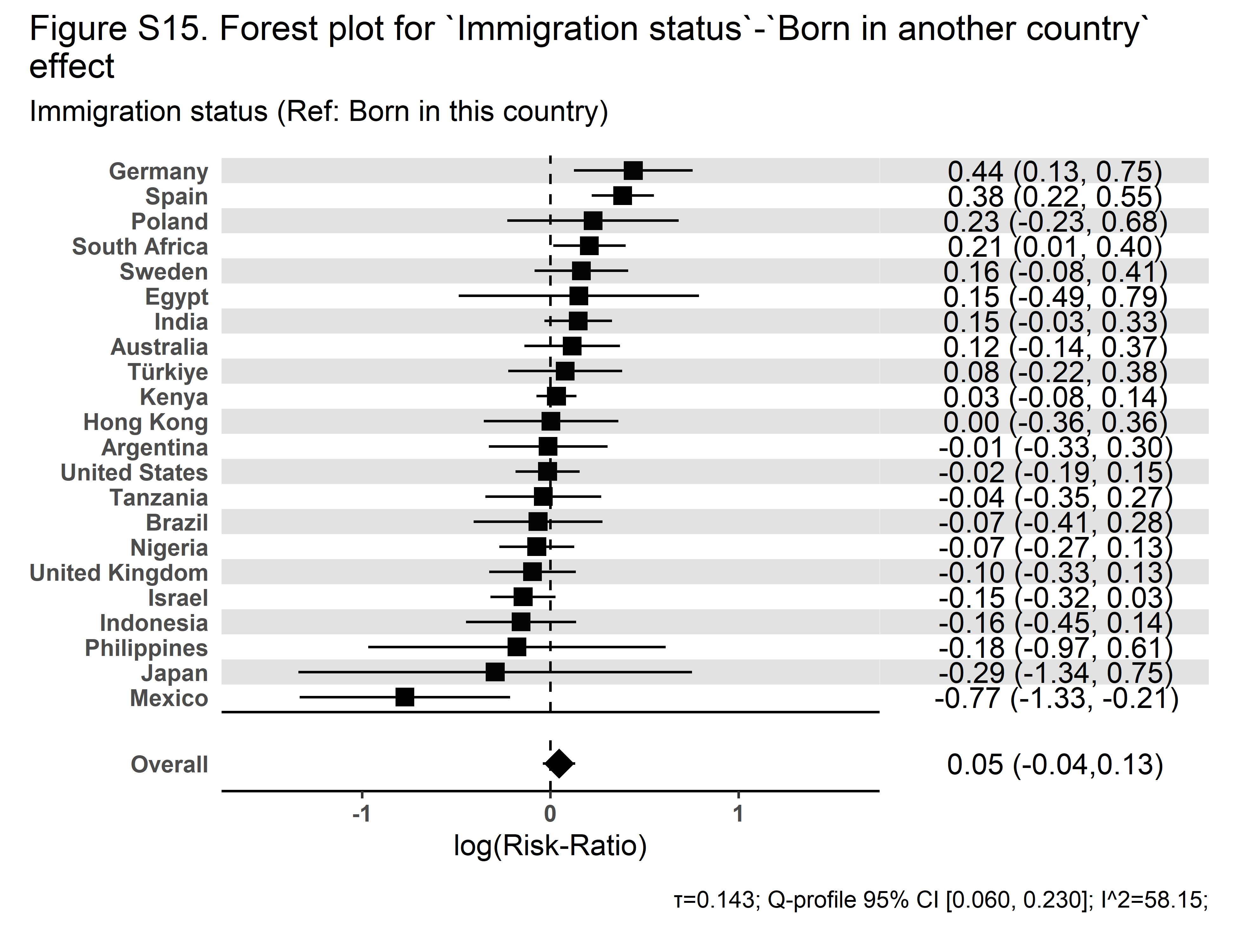 |

| **Secular Community Participation** | **Religious Service Attendance** |
| --- | --- |
| 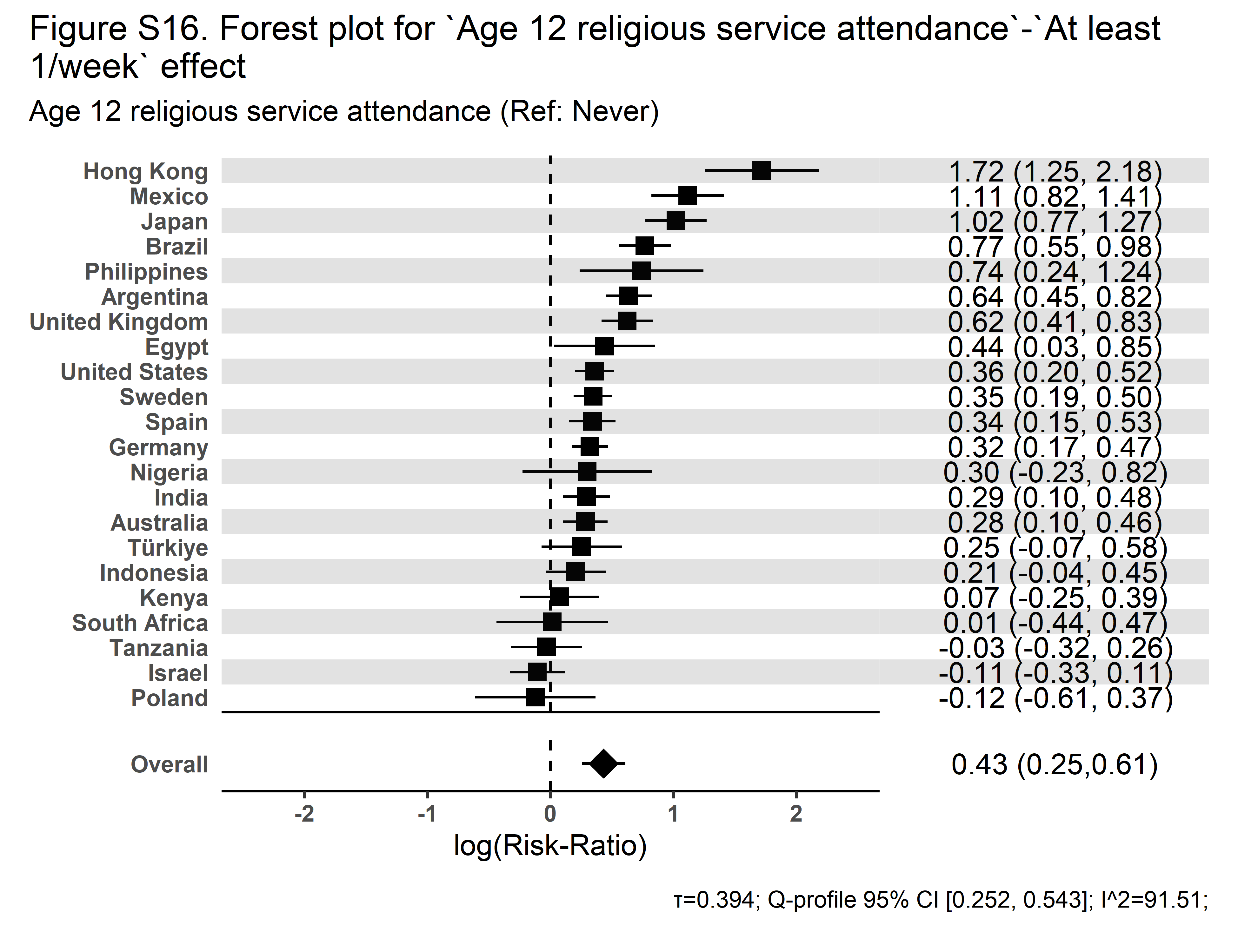 | 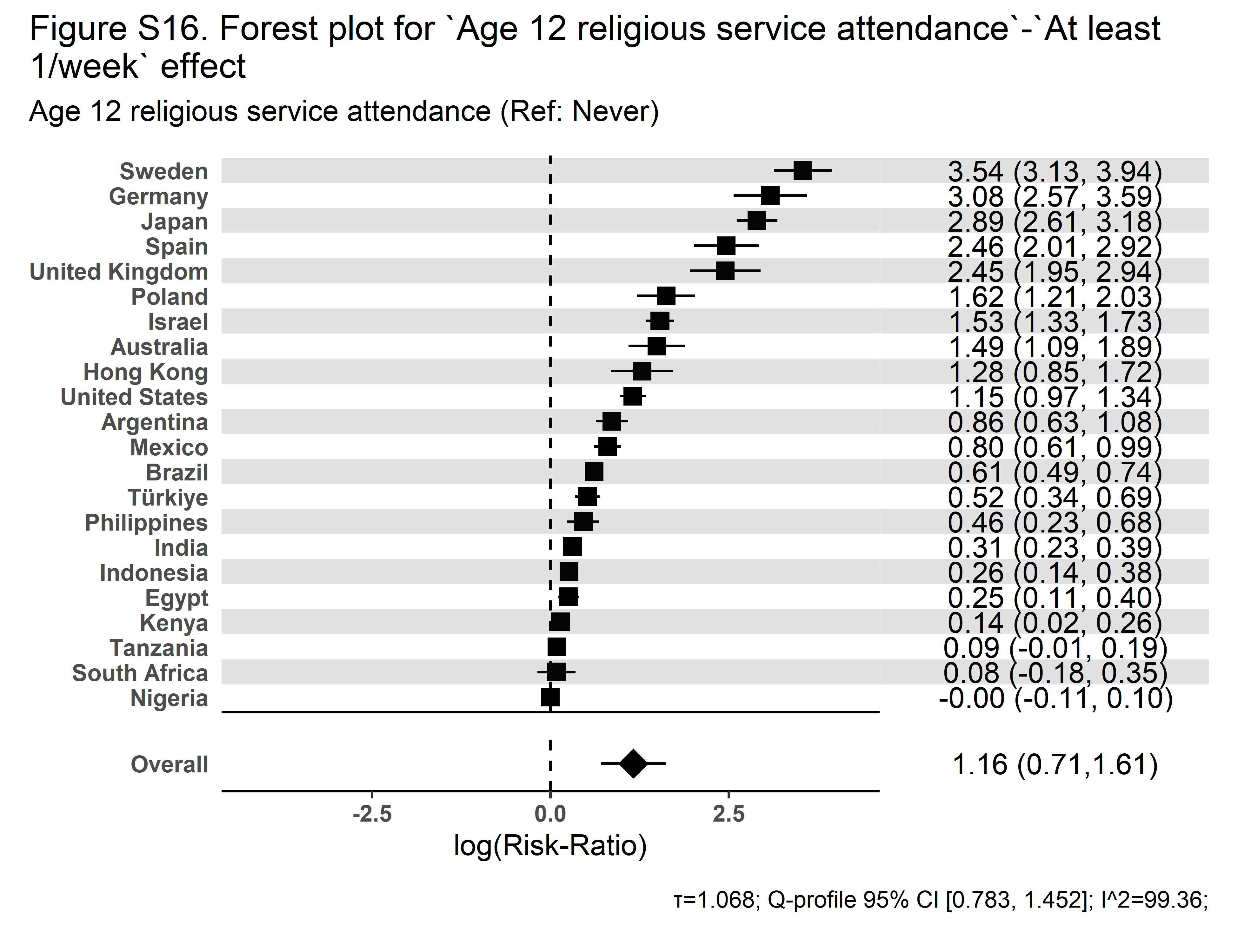 |

| **Secular Community Participation** | **Religious Service Attendance** |
| --- | --- |
| 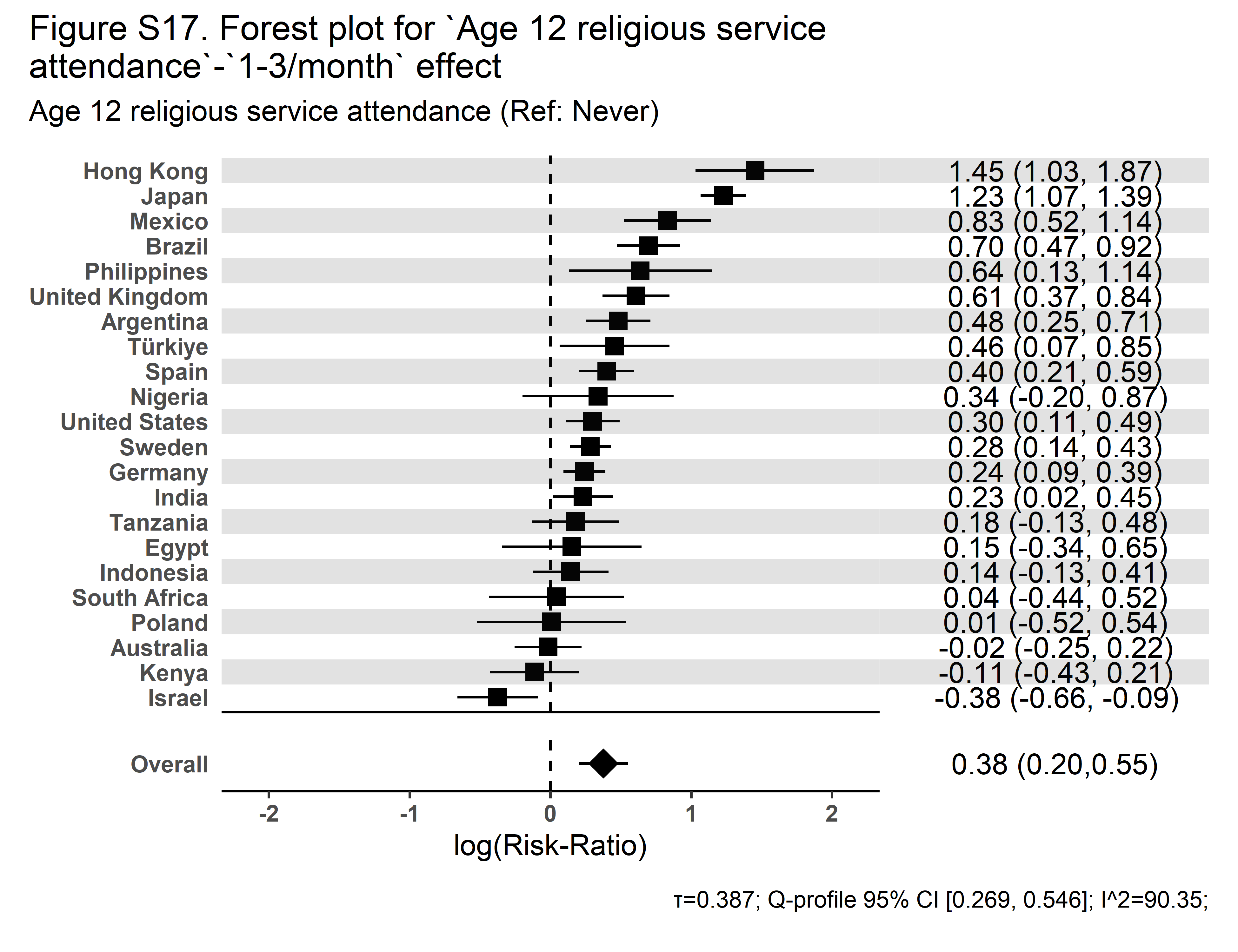 | 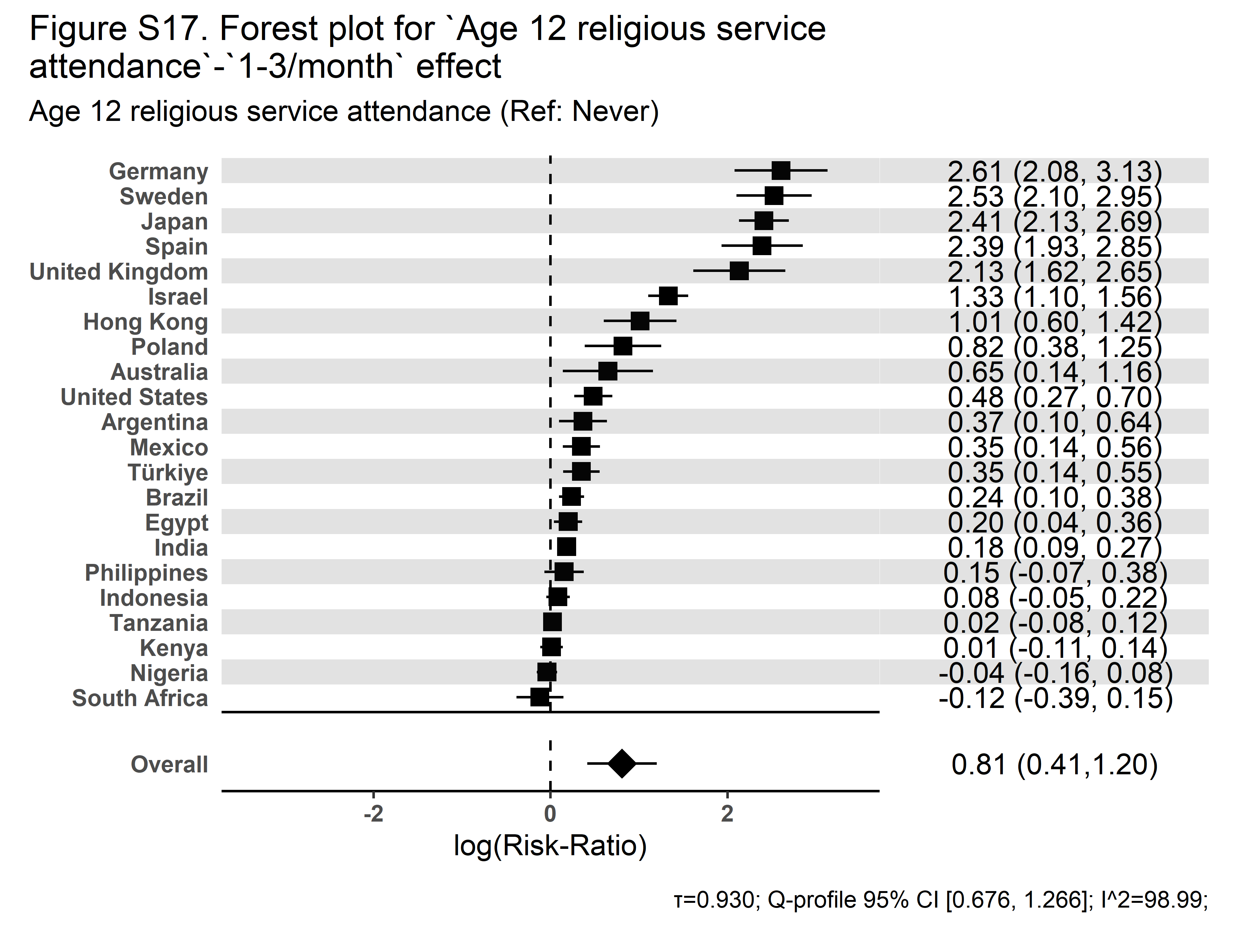 |

| **Secular Community Participation** | **Religious Service Attendance** |
| --- | --- |
| 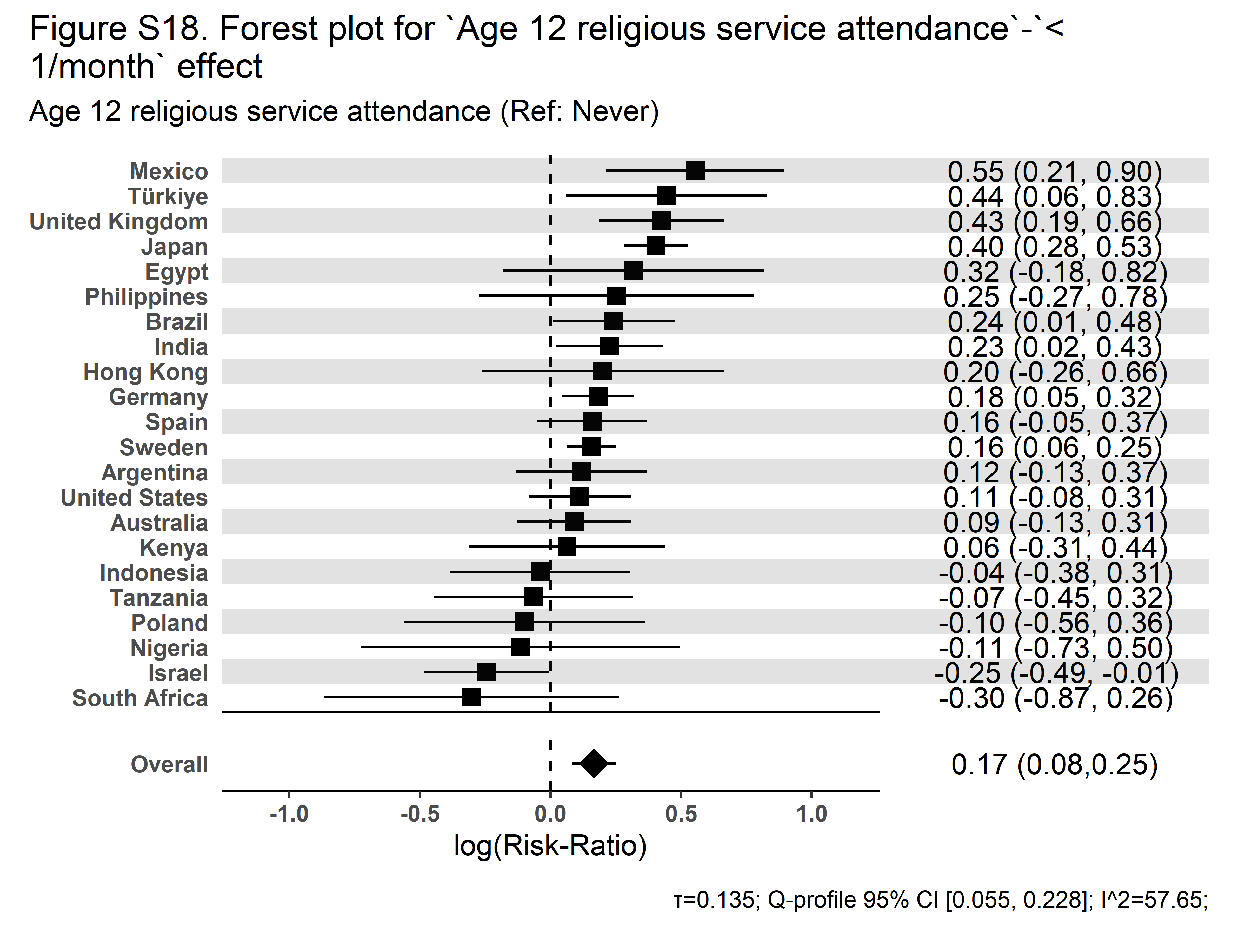 | 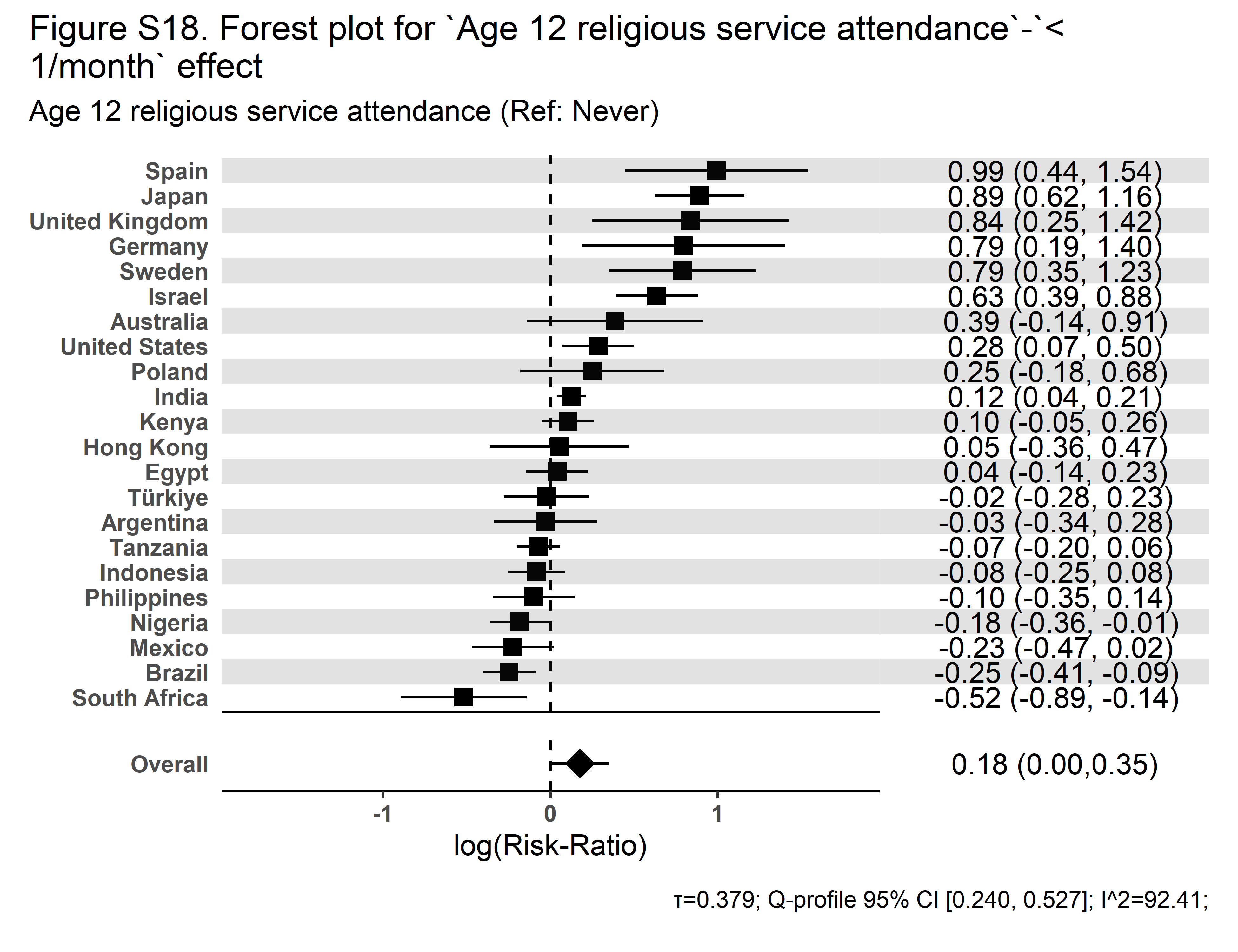 |

| **Secular Community Participation** | **Religious Service Attendance** |
| --- | --- |
| 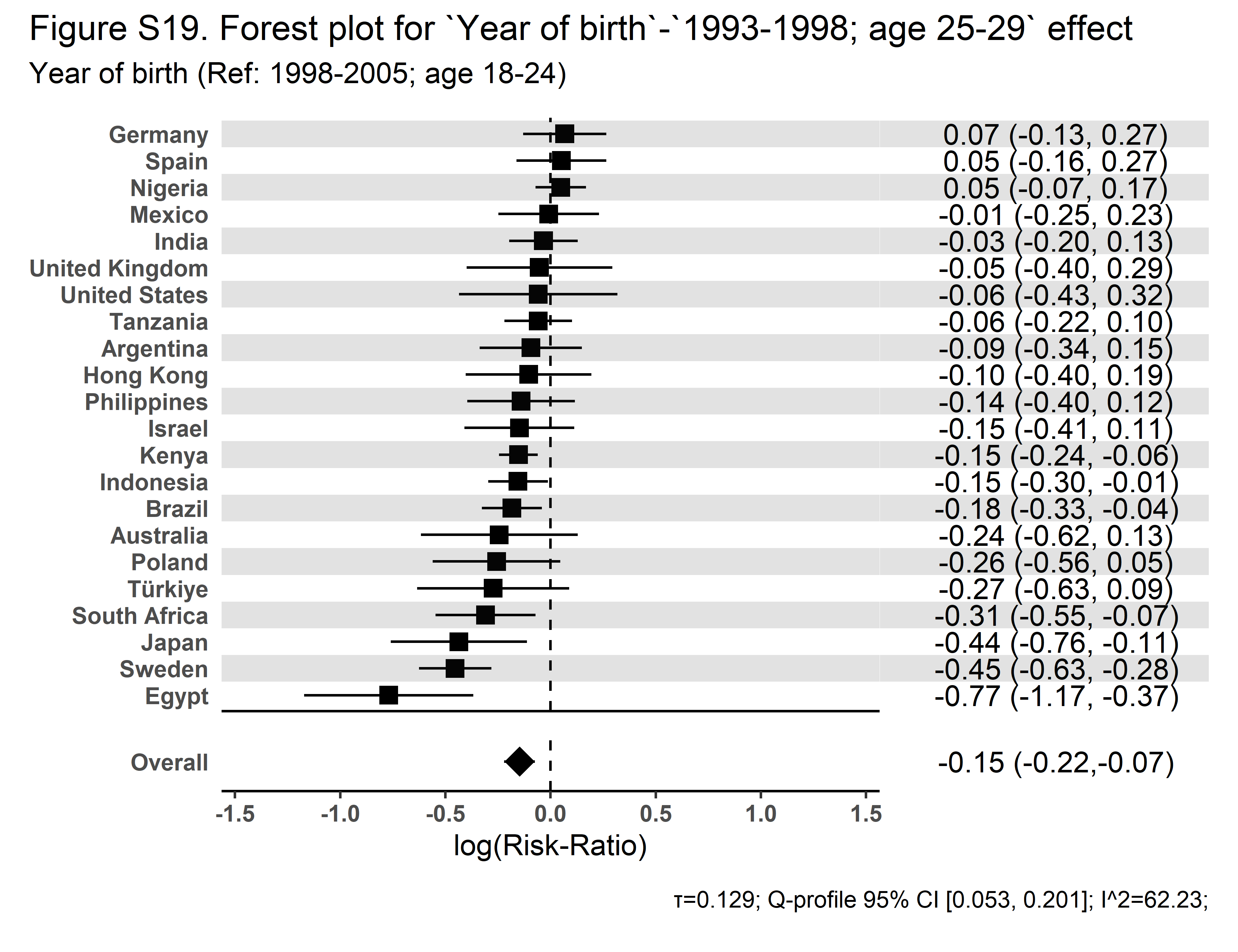 | 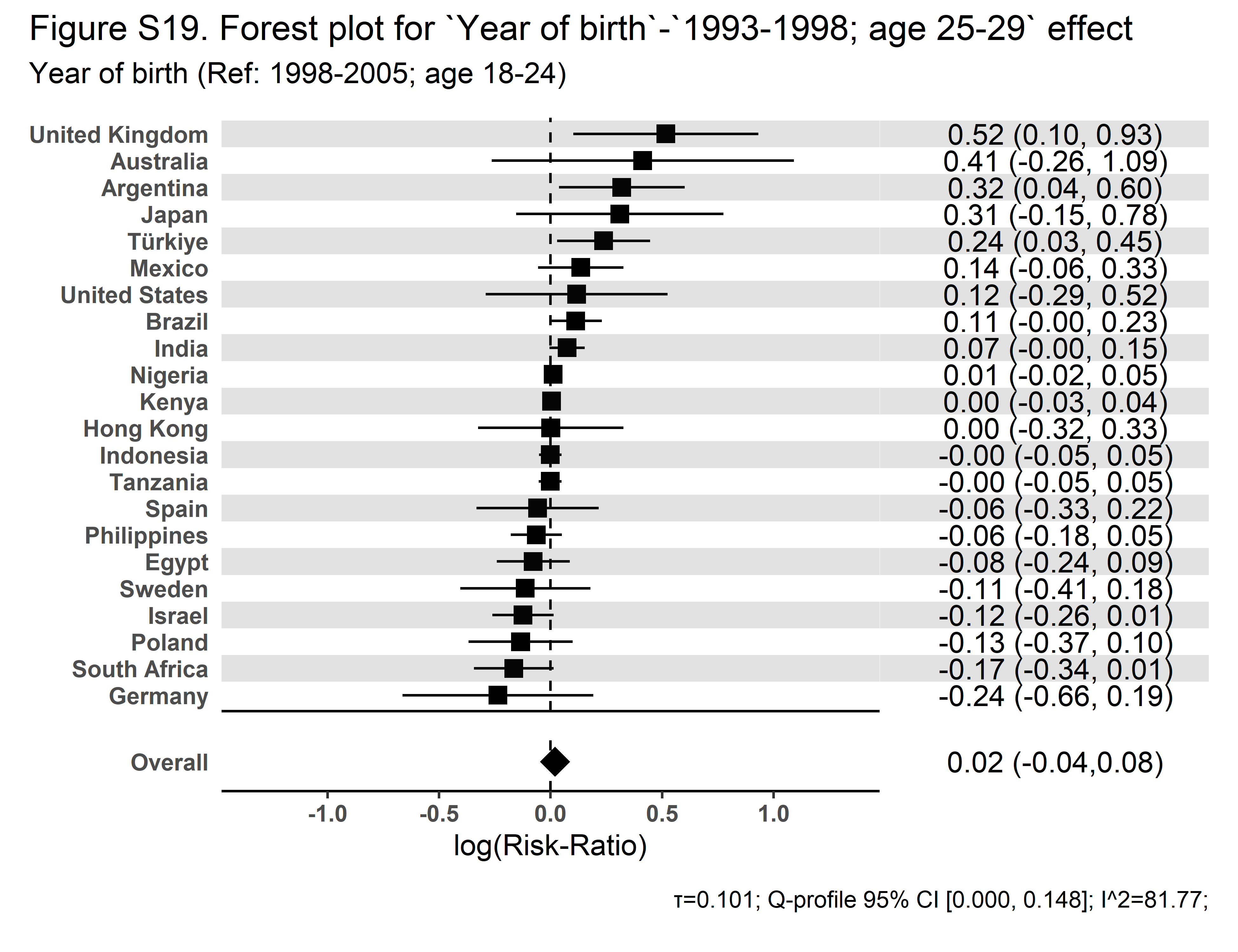 |

| **Secular Community Participation** | **Religious Service Attendance** |
| --- | --- |
| 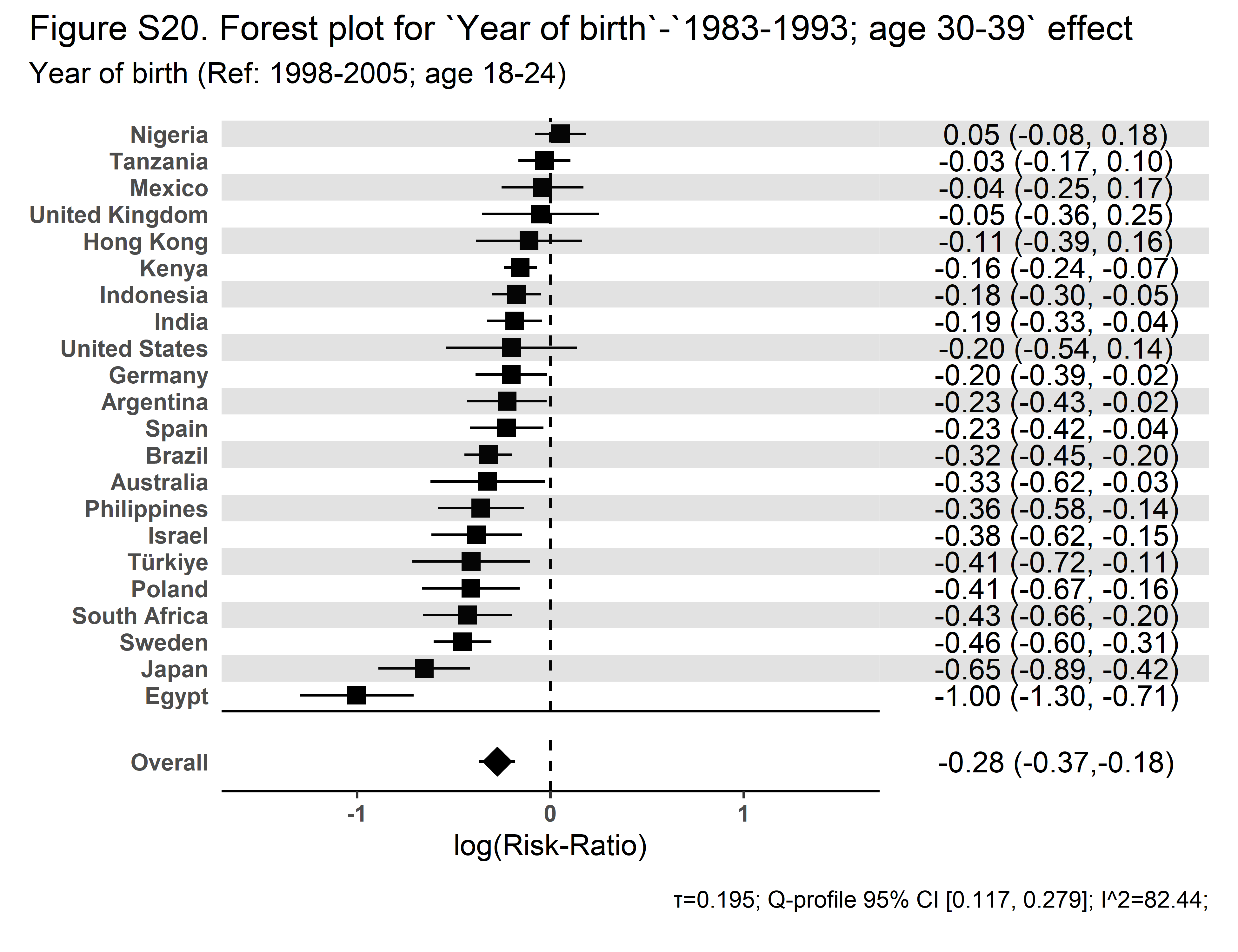 | 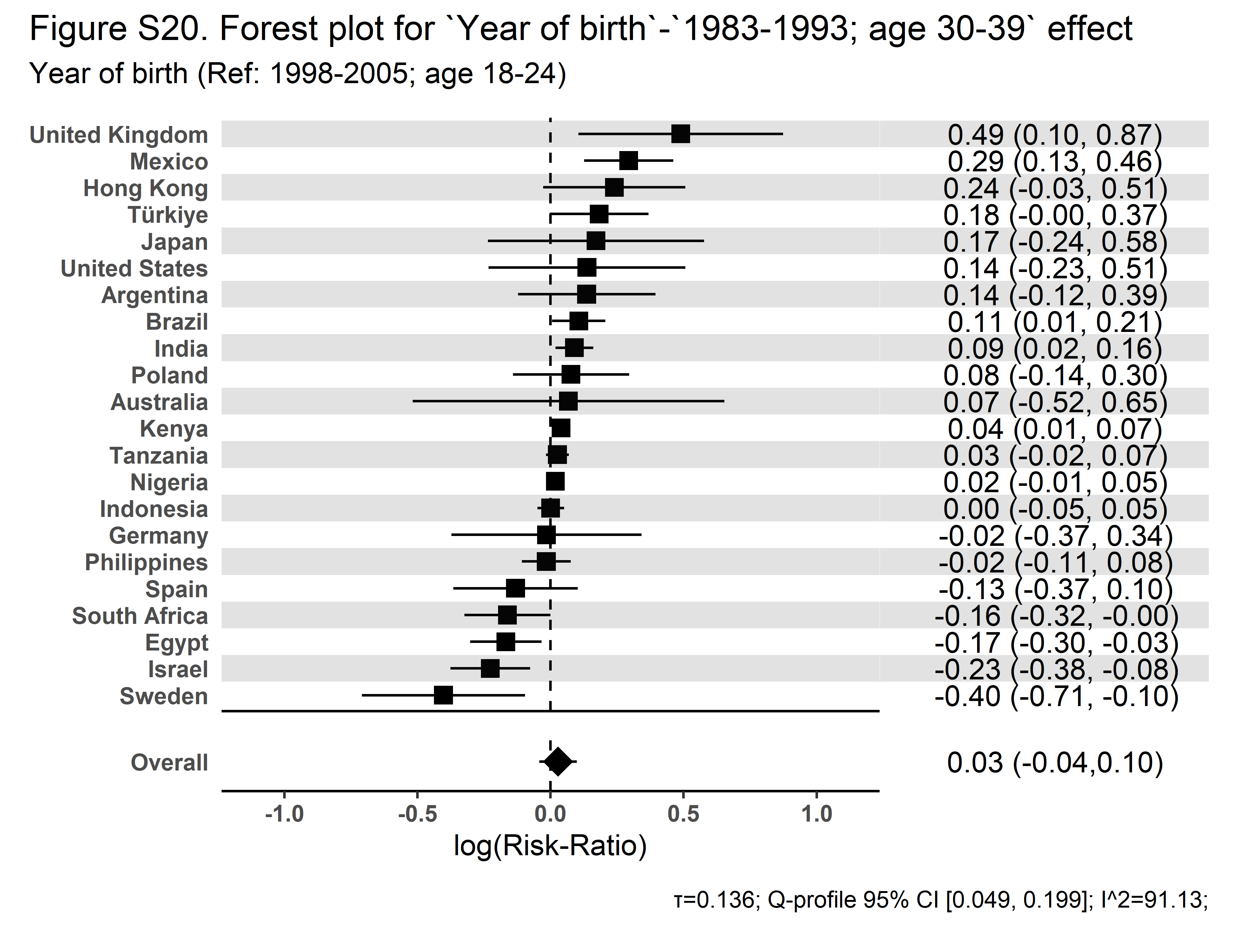 |

| **Secular Community Participation** | **Religious Service Attendance** |
| --- | --- |
| 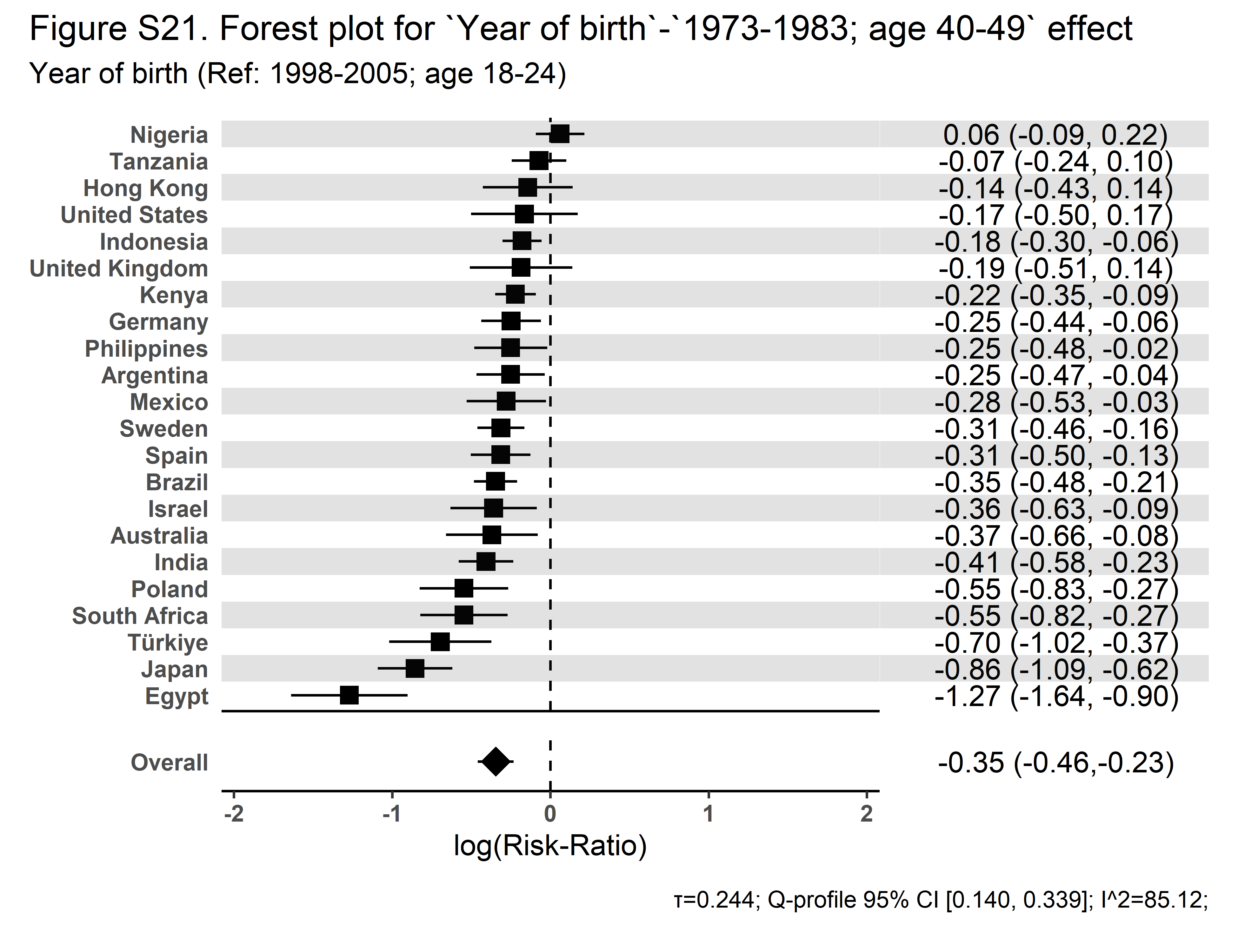 | 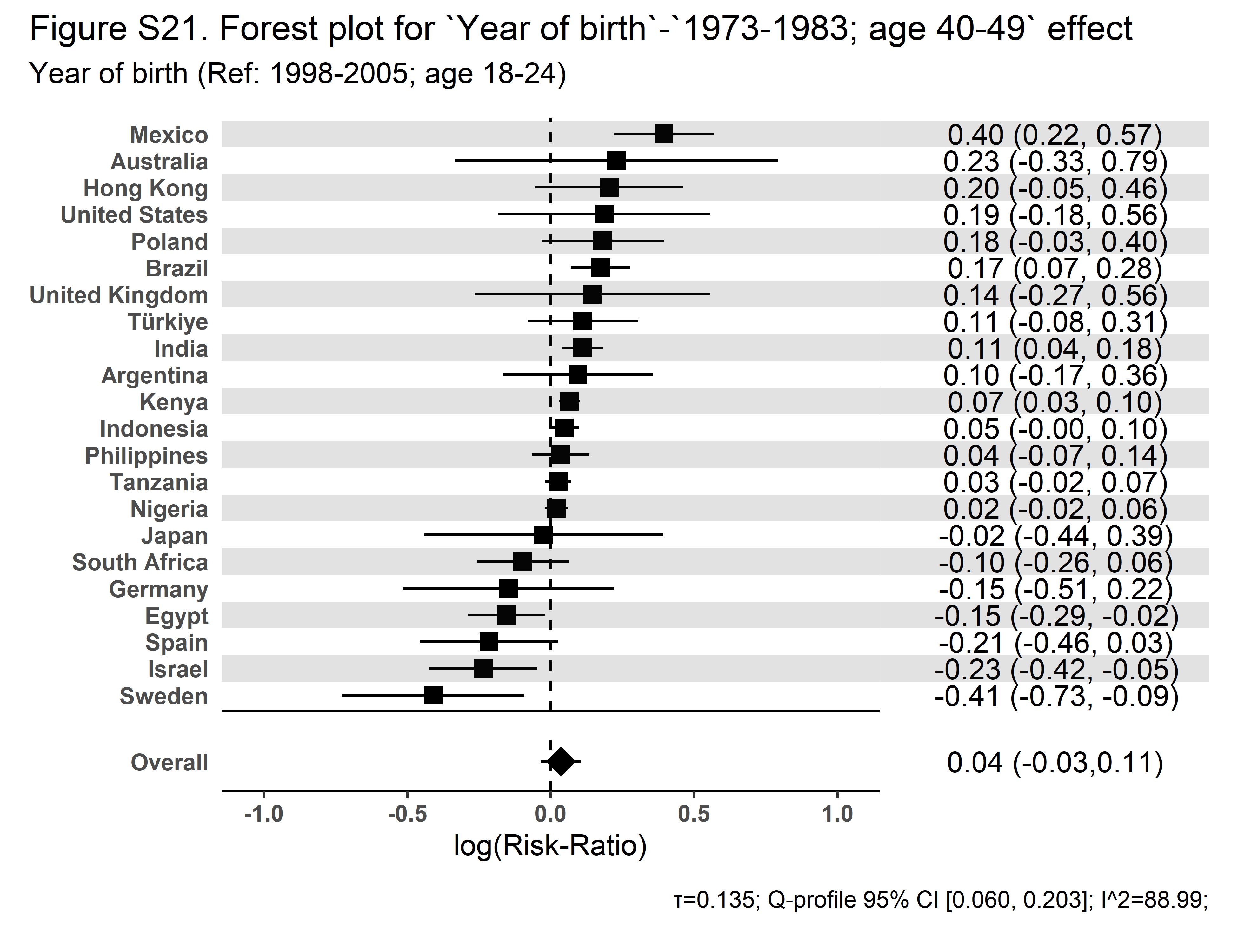 |

| **Secular Community Participation** | **Religious Service Attendance** |
| --- | --- |
| 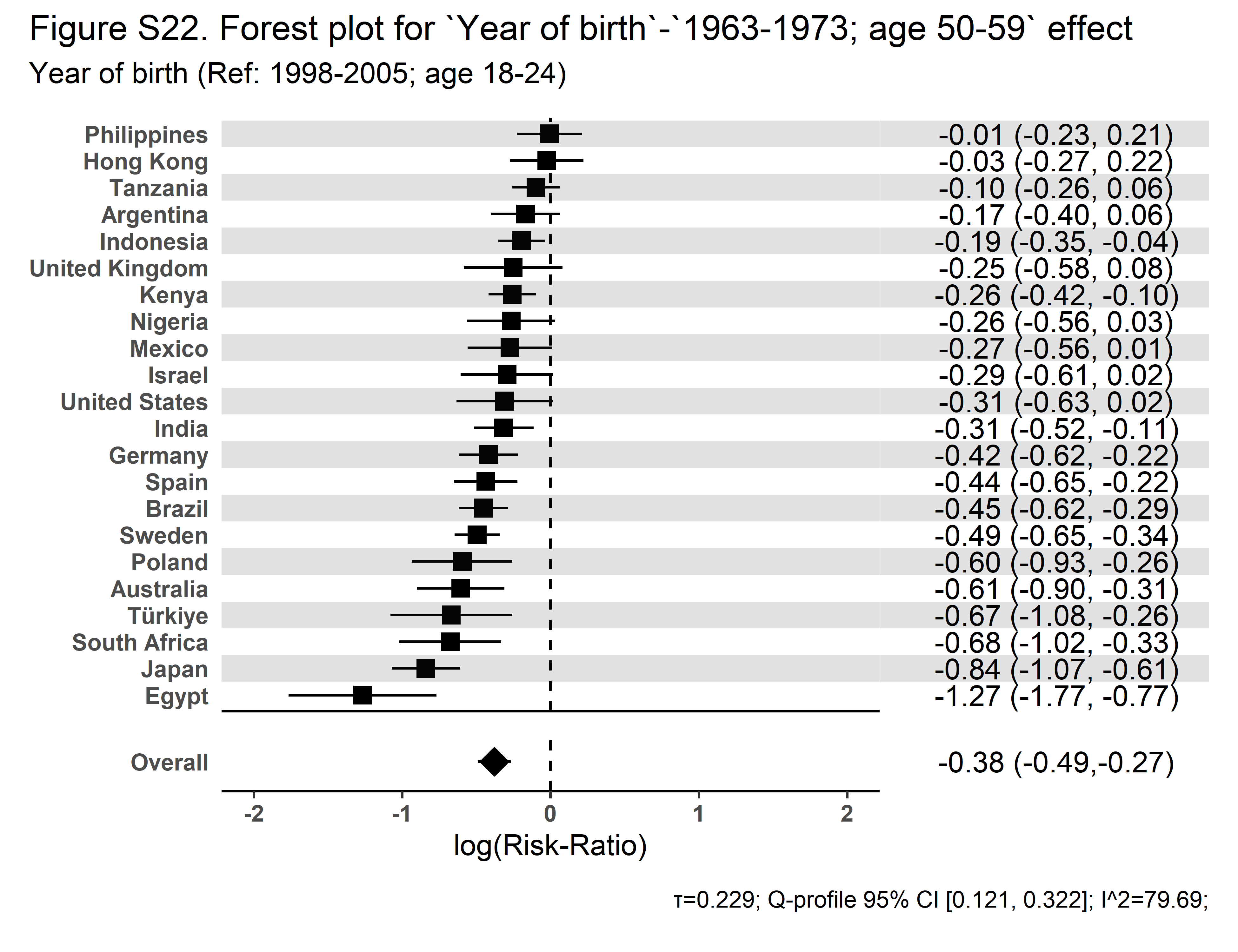 | 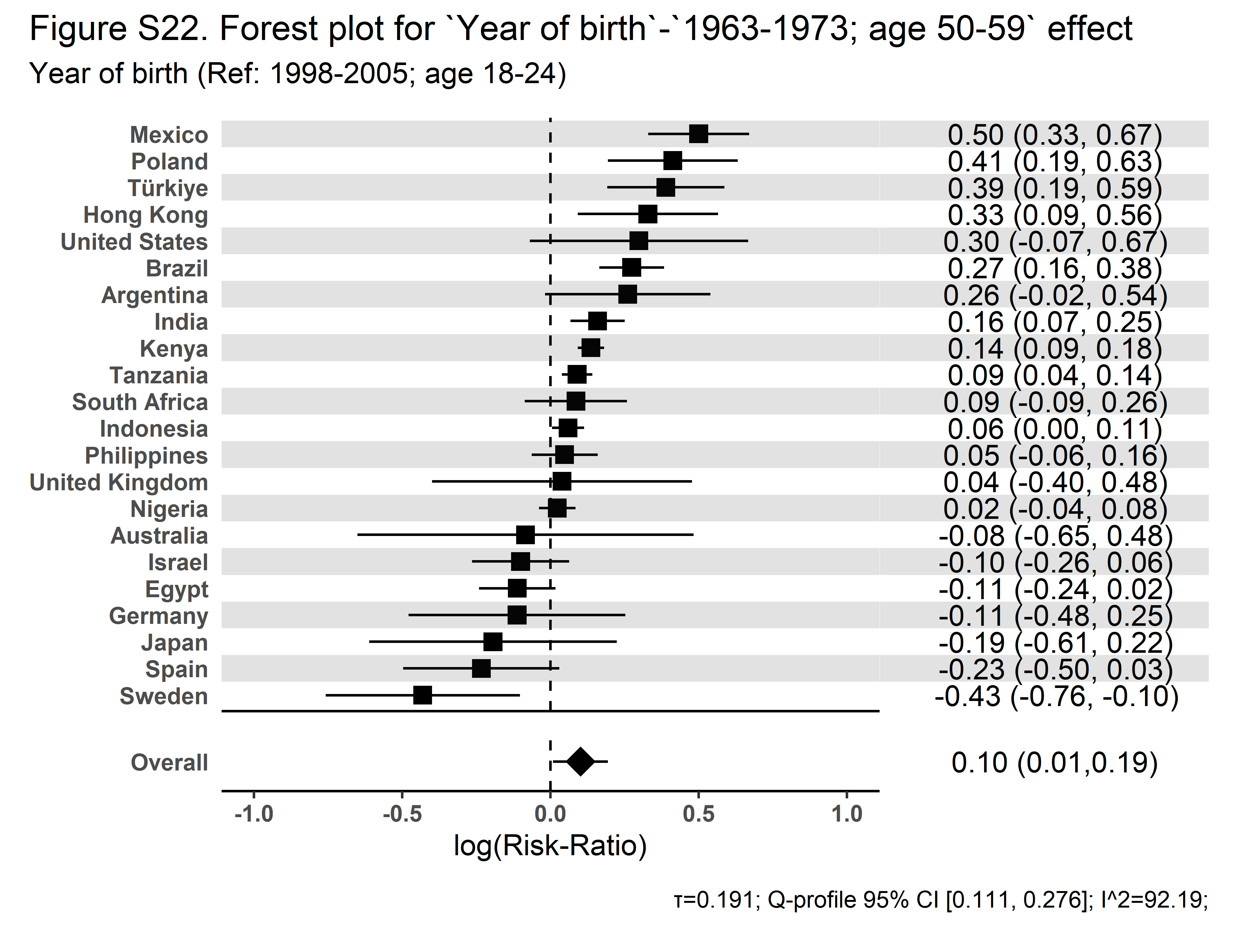 |

| **Secular Community Participation** | **Religious Service Attendance** |
| --- | --- |
| 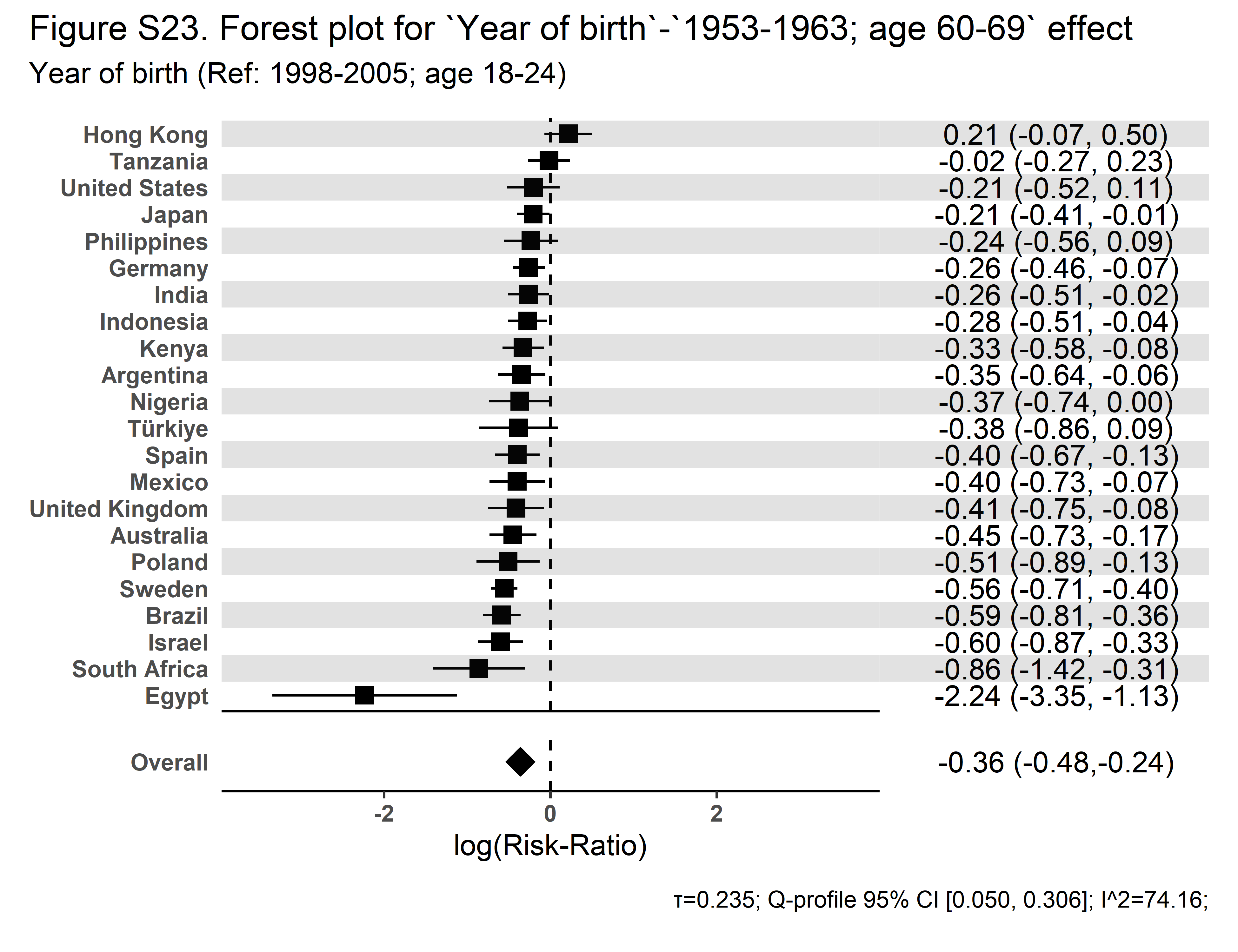 | 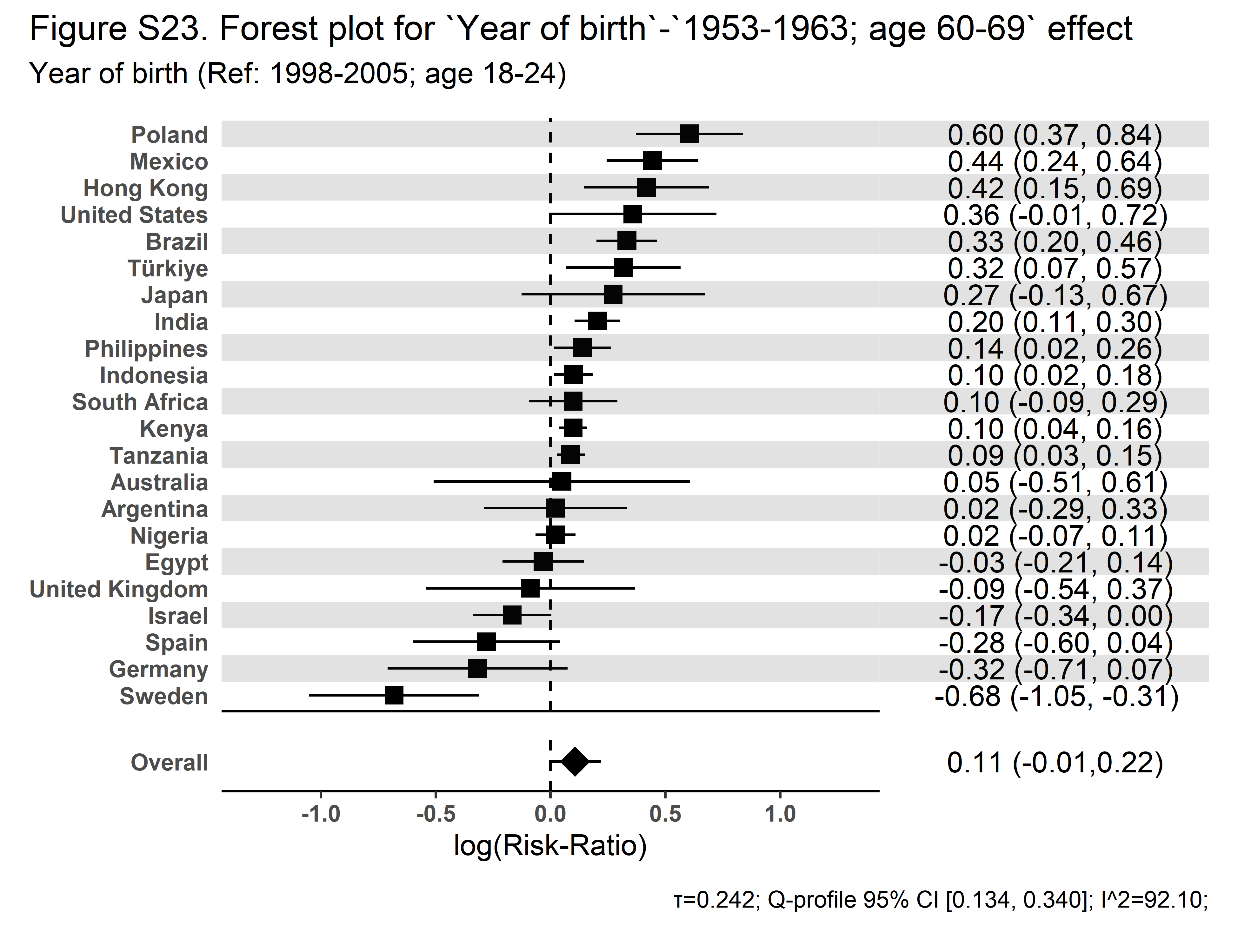 |

| **Secular Community Participation** | **Religious Service Attendance** |
| --- | --- |
| 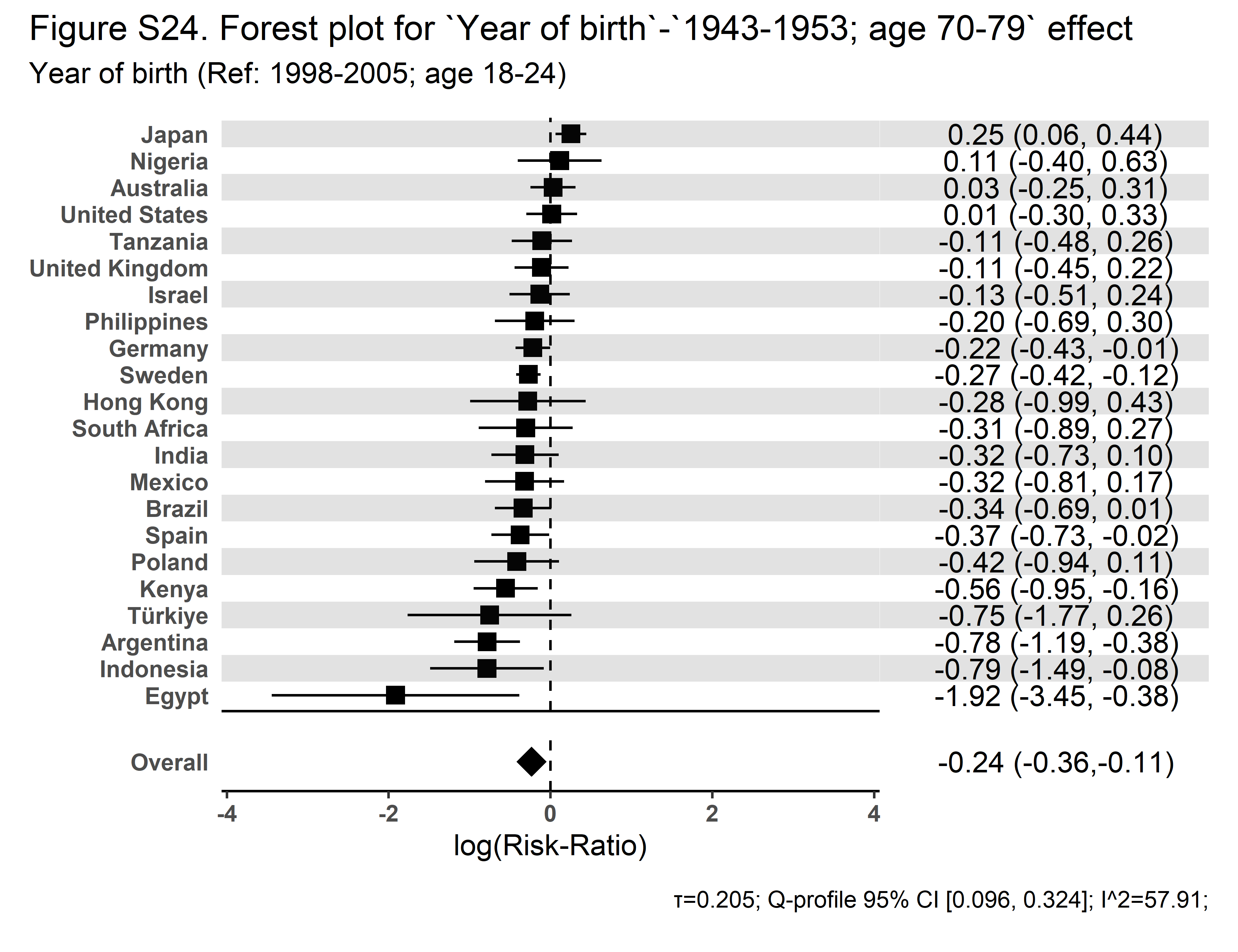 | 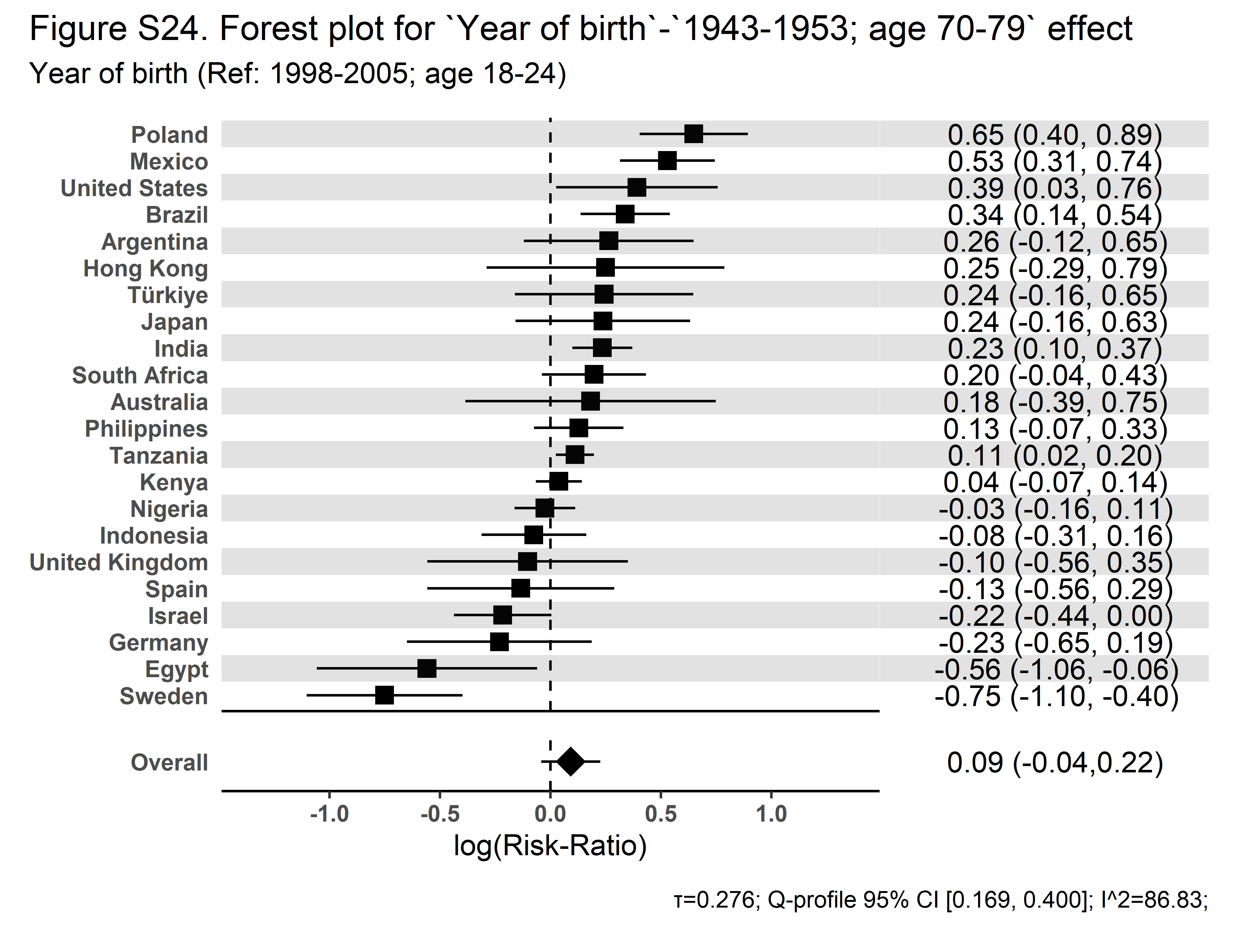 |

| **Secular Community Participation** | **Religious Service Attendance** |
| --- | --- |
| 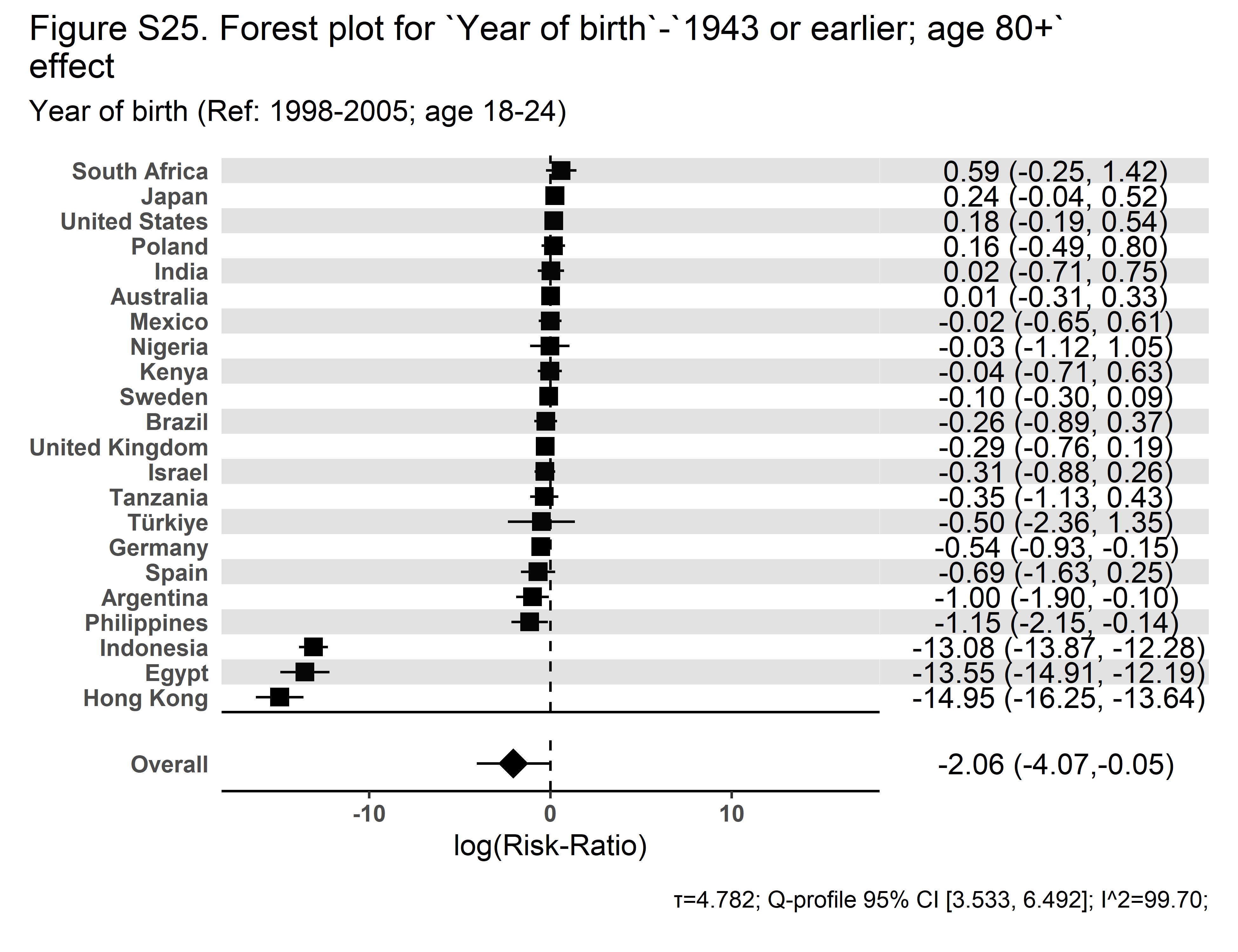 | 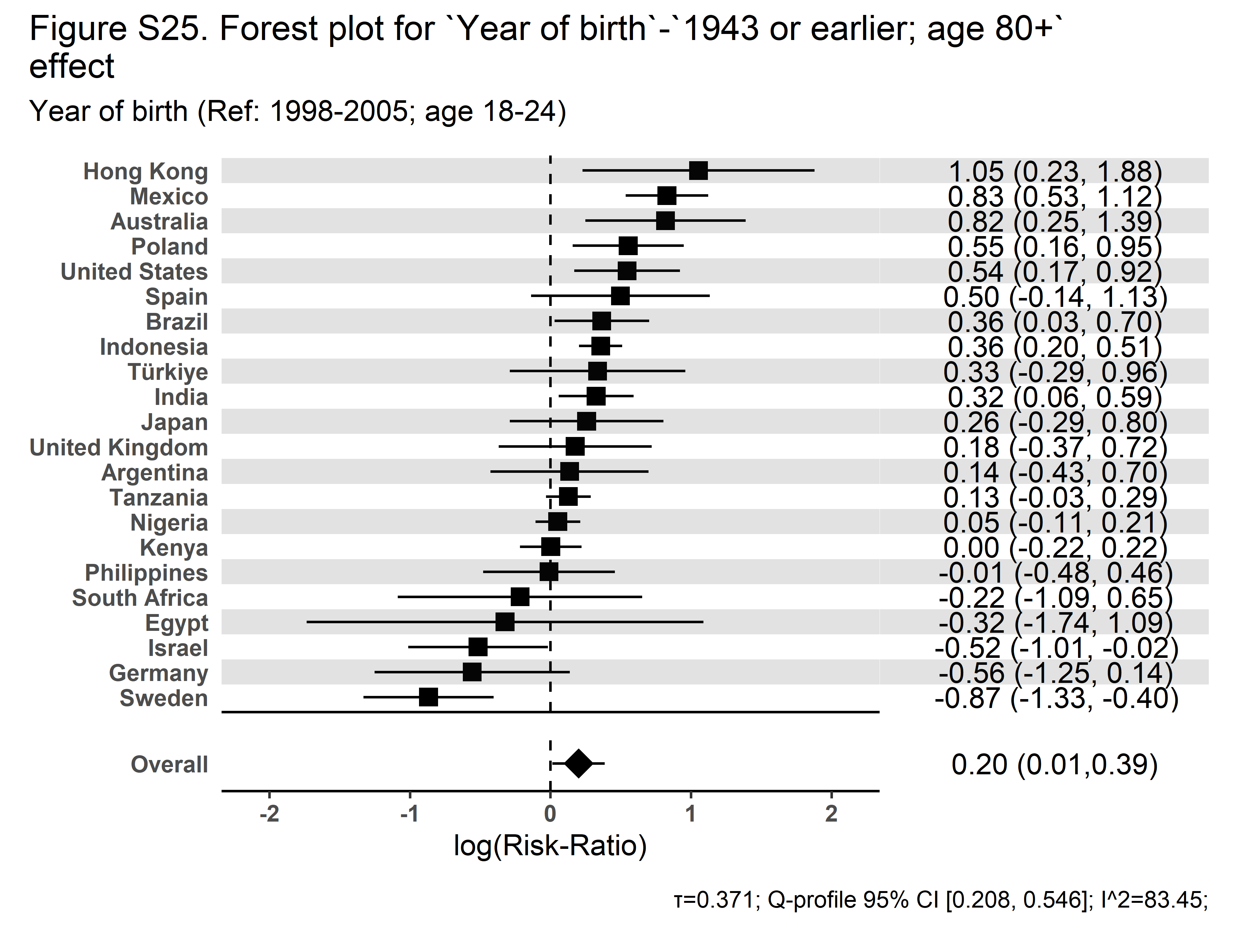 |

| **Secular Community Participation** | **Religious Service Attendance** |
| --- | --- |
| 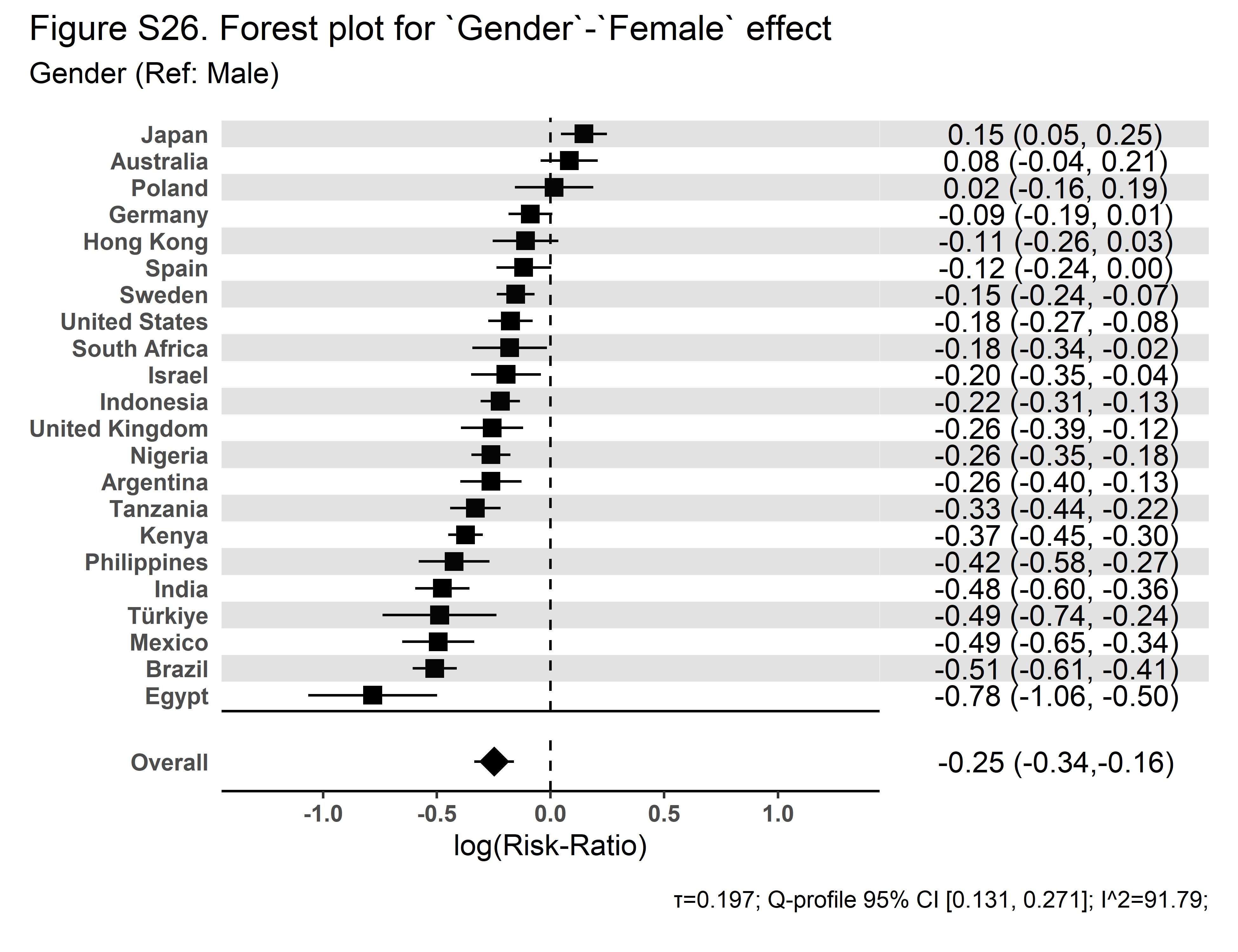 | 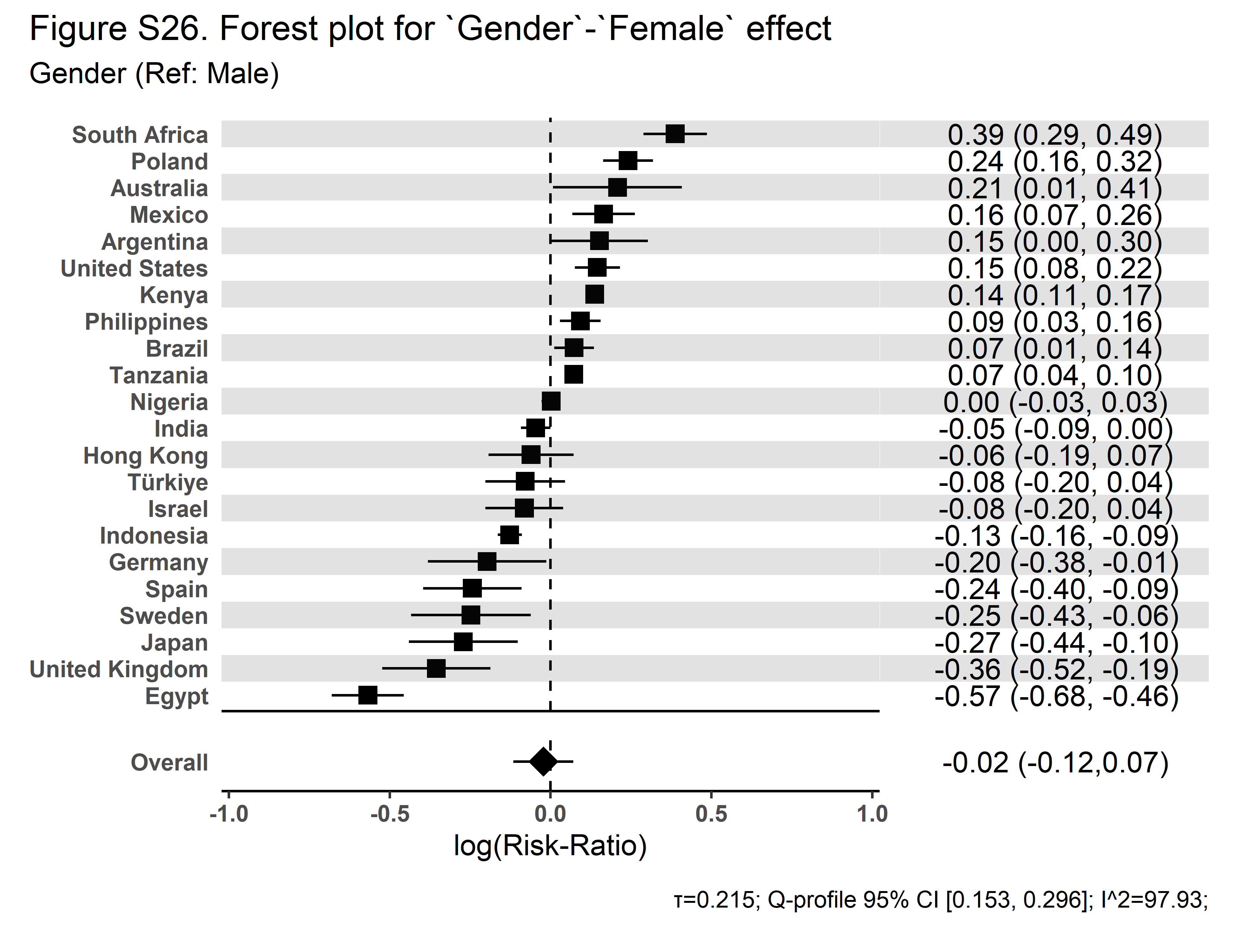 |

| **Secular Community Participation** | **Religious Service Attendance** |
| --- | --- |
| 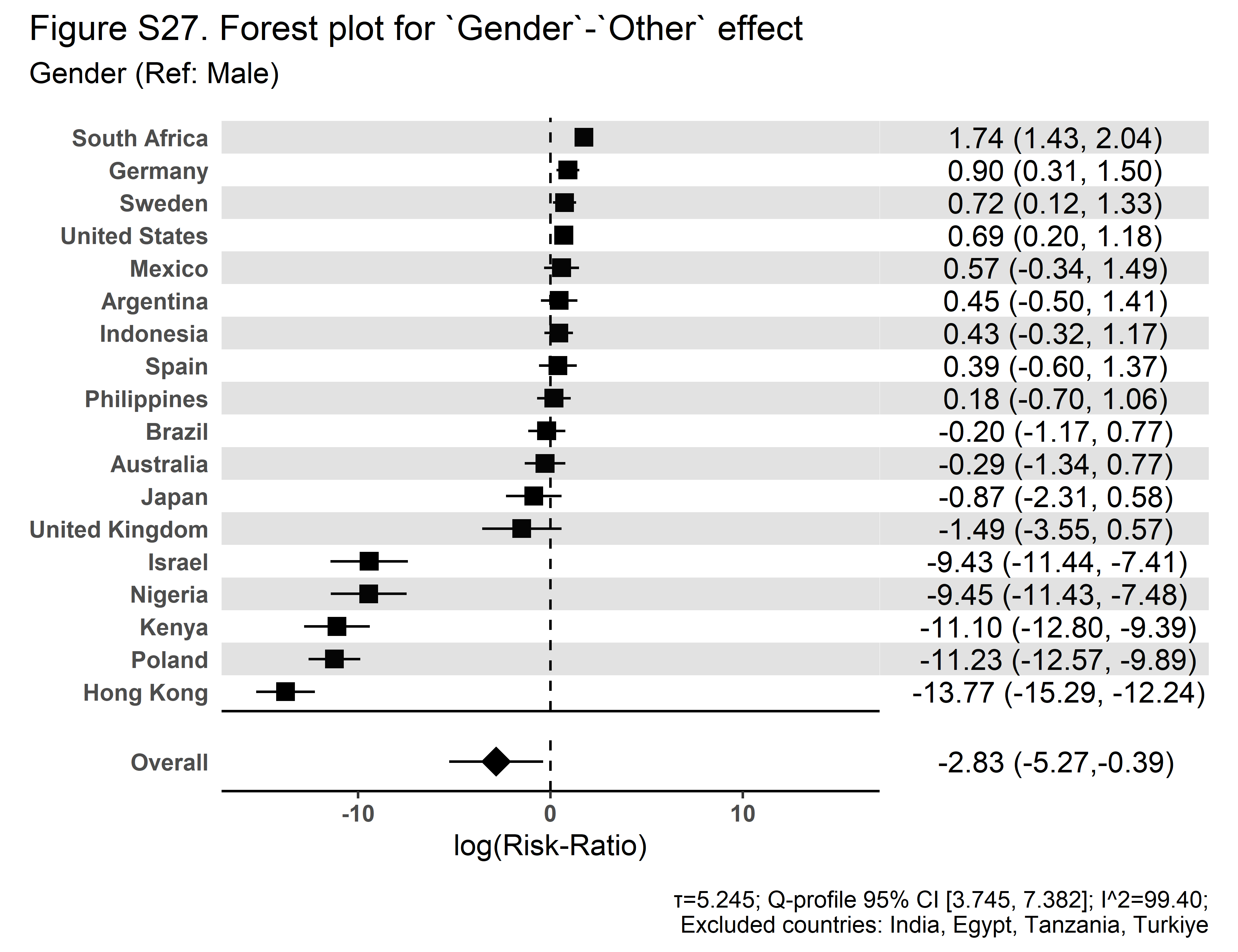 | 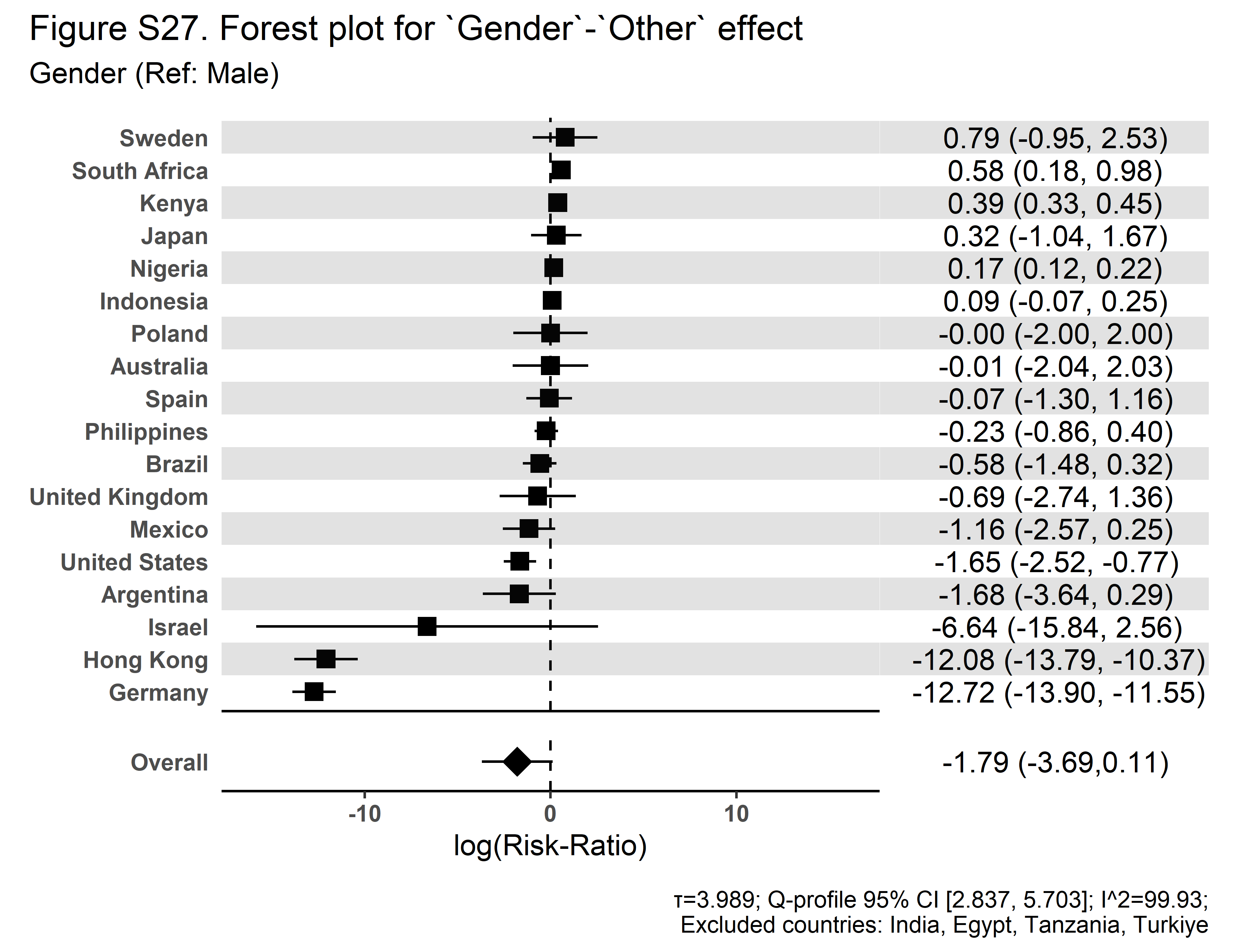 |
